# A prebiotic diet changes neural correlates of food decision-making in overweight adults: a randomized controlled within-subject cross-over trial

**DOI:** 10.1101/2023.05.30.23290707

**Authors:** Evelyn Medawar, Frauke Beyer, Ronja Thieleking, Sven-Bastiaan Haange, Ulrike Rolle-Kampczyk, Madlen Reinicke, Rima Chakaroun, Martin von Bergen, Michael Stumvoll, Arno Villringer, A. Veronica Witte

## Abstract

**Objective:** Animal studies suggest that prebiotic, plant-derived nutrients could improve homeostatic and hedonic brain functions through improvements in microbiome-gut-brain communication. However, little is known if these results are applicable to humans. Therefore, we tested the effects of high-dosed prebiotic fiber on reward-related food decision-making in a randomized controlled within-subject cross-over study and assayed potential microbial and metabolic markers.

**Design:** 59 overweight young adults (19 females, 18-42 years, body mass index 25-30 kg/m^2^) underwent functional task MRI before and after 14 days of supplementary intake of 30 g/d of inulin (prebiotics) and equicaloric placebo, respectively. Short chain fatty acids (SCFA), gastrointestinal hormones, glucose/lipid and inflammatory markers were assayed in fasting blood. Gut microbiota and SCFA were measured in stool.

**Results:** Compared to placebo, participants showed decreased brain activation towards high-caloric wanted food stimuli in the ventral tegmental area and right orbitofrontal cortex after prebiotics (pre-registered, pFWE < 0.05). While fasting blood levels remained largely unchanged, 16S-rRNA sequencing showed significant shifts in the microbiome towards increased occurrence of, among others, SCFA-producing *Bifidobacteriacea*, and changes in >90 predicted functional signaling pathways after prebiotic intake. Changes in brain activation correlated with changes in *Actinobacteria* microbial abundance and associated activity linked with SCFA production, such as ABC transporter metabolism.

**Conclusions:** In this proof-of-concept study, a prebiotic intervention attenuated reward-related brain activation during food-decision making, paralleled by shifts in gut microbiota indicative of higher SCFA production.

## Introduction

Plant-based diets, recognized as a major effector of planetary health ^[1]^, are more beneficial for cardiovascular and brain health compared to conventional Western diets ^[2, 3]^. Plant-based food and related prebiotic nutrients are less dense in calories and have been suggested to modulate brain function ^[4]^ including feeding ^[5]^ and psychological functioning ^[6]^ via the microbiota-gut-brain axis.

Microbiota-derived metabolites of plant-based dietary fiber such as short-chain fatty acids (SCFA), can cross the blood-brain barrier ^[7]^ to modulate hypothalamic signaling ^[8]^. First experimental studies showed that oral intake of the SCFA butyrate or of the butyrate-producing bacteria *Akkermansia spp*. lowered body weight (in humans ^[9]^) and restored obesity-induced functional brain changes (in mice ^[10]^). Moreover, one week of colonic SCFA delivery modulated hypothalamic-pituitary-adrenal axis-dependent stress-induced cortisol response in a study including 66 healthy men ^[11]^, and intake of autologous feces-derived microbiota from a dietary weight-loss period enhanced weight loss maintenance in humans^[12]^.

Earlier trials in humans showed that supplementary intake of prebiotic fiber such as inulin-type fructans reduced subjective hunger and improved gut hormonal-driven appetite regulation through changes in postprandial glucagon-like peptide (GLP)-1, neuropeptide y (PYY) ^[13]^ (n = 10), and ghrelin ^[14, 15]^ (both n < 50). In another randomized clinical trial (RCT) in >100 obese patients, inulin compared to placebo induced greater weight loss ^[16]^ and exploratory results indicated mood improvements in a microbiota-based subgroup with elevated relative *Coprococcus* abundance at baseline ^[17]^. Own results from two cross-sectional analyses indicated that habitual overall dietary fiber intake links to specific microbiota genera including *Parabacteriodes*, which in turn explained variance in eating behavior in adults with overweight, and treatment success after bariatric surgery ^[18]^.

However, neuroimaging evidence of how prebiotic diets and diet-related microbial changes affect the brain with regard to eating behavior remains to be shown. At the brain level, food decision-making relies on a complex interplay of homeostatic and hedonic signaling, orchestrated by a variety of subcortical and cortical networks involving the brainstem and hypothalamus, striatum and prefrontal cortex areas ^[19]^. The neurobiological underpinnings of (unhealthy) eating behavior, however, have not been fully understood. Functional magnetic resonance imaging (fMRI) studies indicated that presentation of highly palatable food cues leads to a stronger brain response in reward areas than equicaloric, non-palatable food cues ^[20]^. In parallel, disinhibition and unhealthy food craving, sometimes controversially described as food addiction ^[21]^, have been linked with subtle structural differences in the reward network ^[22, 23]^ and with differential brain activation in the ventromedial prefrontal cortex in response to high-caloric food stimuli ^[24]^. Whether these effects can be mitigated by prebiotic dietary targeting the gut-brain axis ^[25]^ is yet far from understood.

We here aimed to test the hypothesis that a high-dosed prebiotic fiber intervention can alter the gut microbiome and thereby neural activation patterns of food reward in a population at risk for weight gain and insulin resistance. To this end, we conducted a RCT in overall healthy adults in a randomized within-subject cross-over design and assessed food wanting using functional MRI before and after 14 days of daily 30 g supplementary intake of inulin (prebiotic fiber) and equicaloric maltodextrin (placebo), respectively. Suggested microbial and metabolic mediators of potential effects were measured using feces and serum proxies collected at all four timepoints. The study and analyses were preregistered at ClinicalTrials.gov/NCT03829189 and <u>osf.io/ynkxw</u>.

## Methods

### Study design

In this within-subject cross-over design, participants underwent screening and, if eligible, received both verum and placebo in a randomized order (2 arms) for 14 days each, separated by a wash-out period of at least two weeks (**Fig. 1**). Verum (prebiotic fiber) consisted of 30 g inulin (63 kcal, 26.7 g fiber, Orafti® Beneo Synergy1, BENEO GmbH, Mannheim, Germany) per day compared to calorie-matched placebo consisting of 16 g maltodextrin (63 kcal, 0 g fiber), each provided as two sachets per day.

**Figure 1:**
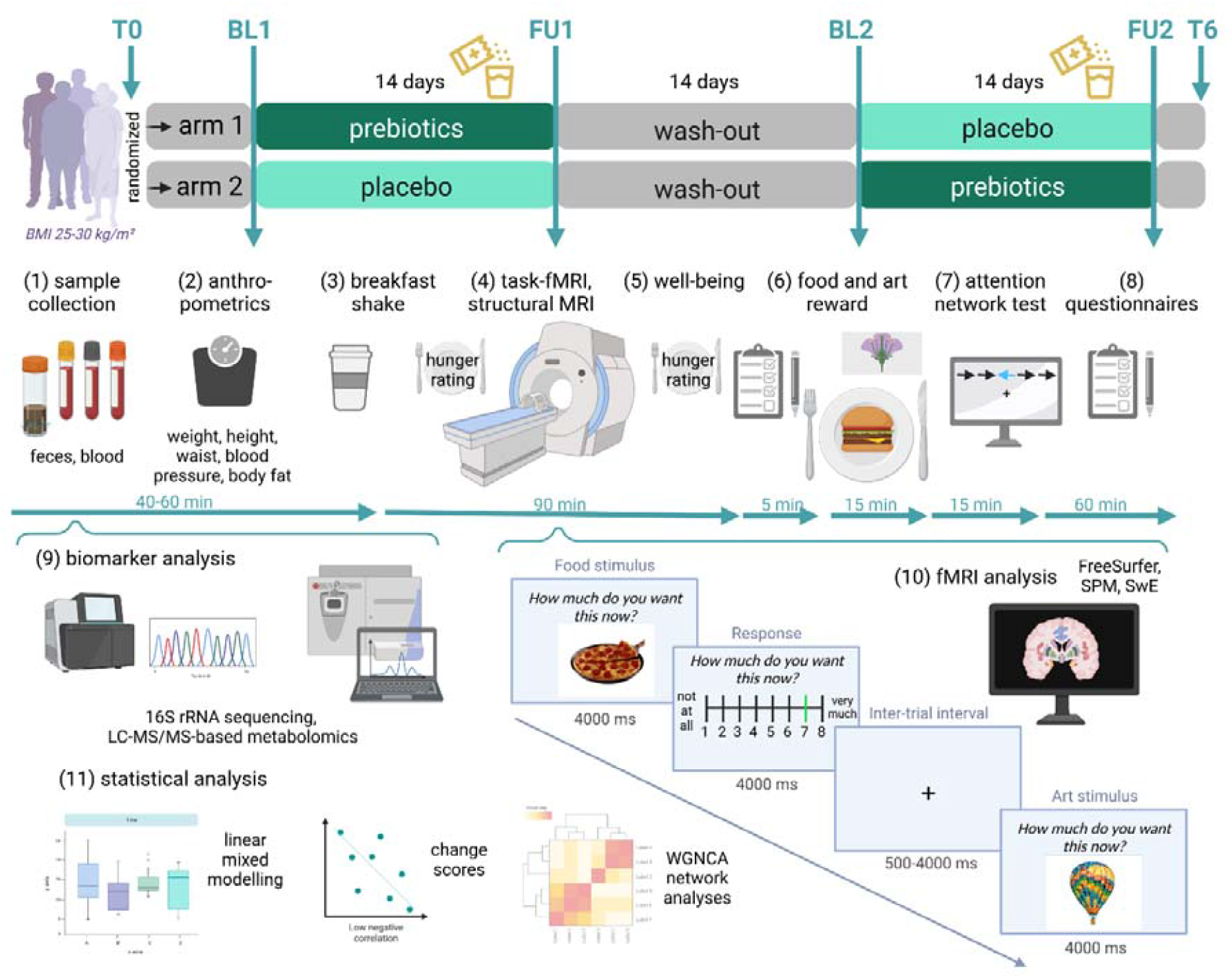
Study design. Within-subject cross-over dietary intervention design with two study arms and up to six measurement timepoints (upper panel, T0: screening; BL1/2: baseline 1/2, FU1/2: follow-up 1/2, T6: additional follow-up). Participants were randomly assigned to receive first prebiotics and second placebo (arm 1), or vice versa (arm 2), for 14 days each, separated by a 14 days washout period. Following the same timeline, at BL1, FU1, BL2, and FU2, participants provided stool samples and underwent fasting blood draw (1), anthropometric measurements (2), received a standard breakfast shake (3) and MRI assessments (4), followed by brief surveys (5), food remuneration (6) and further tests and questionnaires (7-8). Steps (9-11) indicate data processing and statistical analysis. Screens give fMRI wanting task paradigm scheme and timing. Abbreviations: BL, baseline; FU, follow-up; fMRI, functional magnetic resonance imaging; LC-MS/MS, liquid chromatography–mass spectrometry; SPM, statistical parametric mapping, SwE, sandwich estimator, WGNCA, weighted graph network correlational analysis. Created with BioRender.com.

All participants were invited to baseline and follow-up visits for each condition, resulting in four study visits with feces and fasting blood sample collection, fMRI and questionnaires. Briefly, after fasting blood draw and anthropometrics (∼45min), participants received a neutral drink covering 10% of their individual daily energy requirement. Right after, the MRI assessment followed (∼2h), which was then followed by further computer-based assessments (∼1.5h), see **SI_general** for further details.

### Participants

Volunteers of all gender were recruited via online and local advertisements and the institute’s local database. Inclusion criteria were a body-mass-index of 25-30 kg/m^2^, no MRI contraindications, aged 18-45 years, women: intake of oral contraceptives. Exclusion criteria were: neurological or psychiatric disease; intake of medication acting on the central nervous system; diabetes mellitus type 2; severe untreated internal disease including the gastrointestinal tract, lung, heart, vasculature, liver and kidneys; eating disorder or unconventional eating habits; women: pregnancy, breastfeeding as well as daily consumption of > 50 g alcohol, > 10 cigarettes, or > 6 cups of coffee. Out of 106 initially recruited volunteers with screening assessment, 59 participants (19 women, 40 men) took part in the study, with 45 completing all 4 measurement visits (**Fig. 2**).

**Figure 2:**
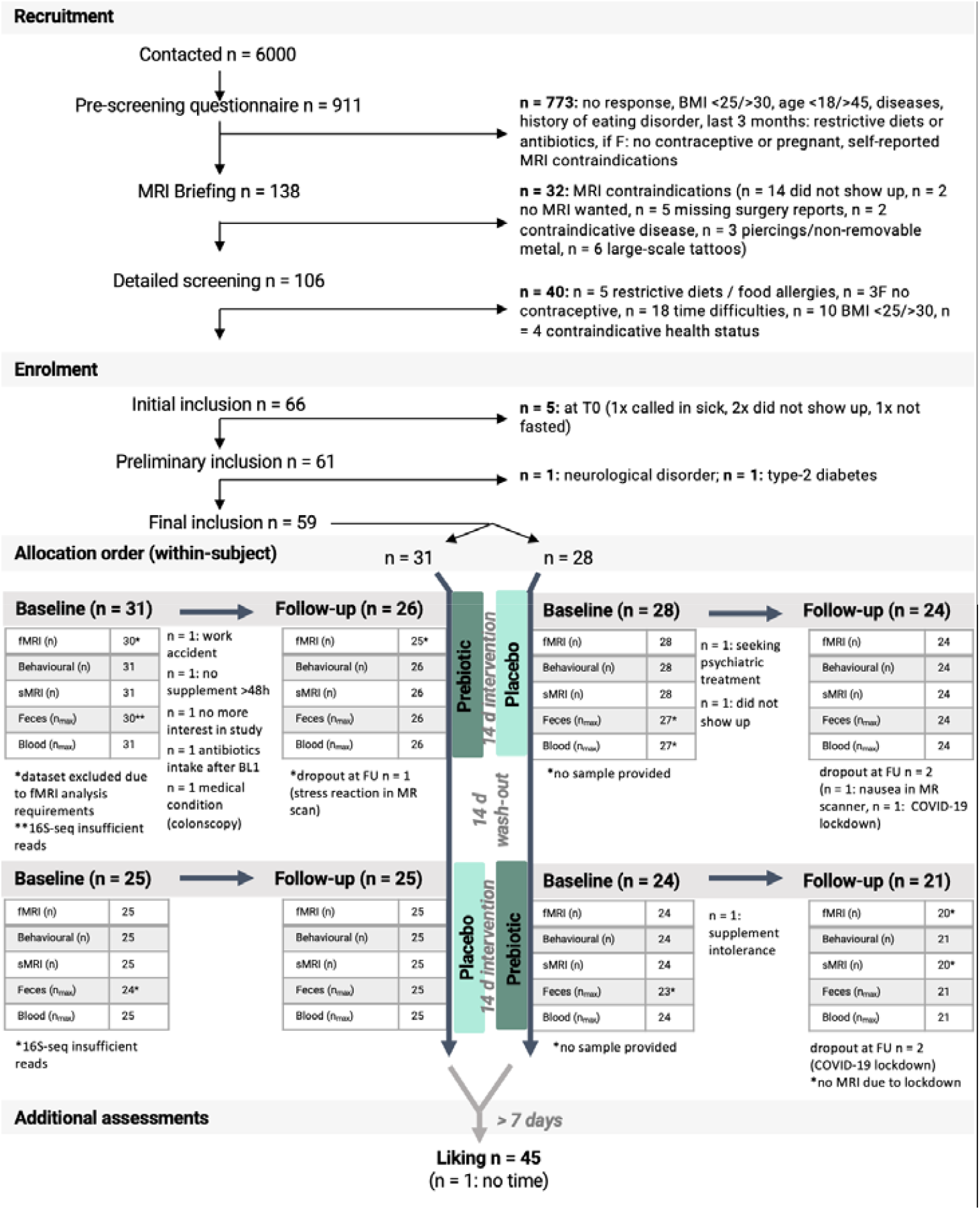
CONSORT flow diagram. Participants underwent a randomized controlled dietary intervention trial in a within-subject cross-over design.

### Ethics, registration and blinding

The ethics board of the Medical Faculty of the University of Leipzig, Germany, raised no concerns regarding the study protocol (228/18-ek) and all participants provided written informed consent. Participants received a small reimbursement of 9-10€/h for testing days and additionally 30€ for study completion. The study was registered at https://clinicaltrials.gov/ct2/show/NCT03829189 and https://osf.io/f6qz5 (14-01-2019) prior to recruitment and data acquisition. Additionally, details on fMRI (pre)processing were uploaded before start of data analysis https://osf.io/ynkxw (11-05-2021). Participants and staff members were blinded regarding the study intervention/placebo allocation.

### Patient and public involvement

The authors acknowledge a missed opportunity of not following a tailored approach to involve patients or the public in the design of the study. We invited and collected comments and assessments from all participants throughout the study to inform the design of upcoming research studies.

### MRI

MRI was performed on a 3T Siemens Prismafit scanner with a 32-channel head coil. FMRI was done in an event-related design assessing wanting of food and art, respectively. Participants were presented with 4 sets of images across 4 sessions (randomized order). Each stimulus was shown for 4000 ms with the question “How much do you want this now?”, followed by a 4000 ms response period, followed by 500-4000 ms inter-stimulus interval with a 500 ms jitter until the next stimulus was presented (**Fig. 1**). Wanting ratings were done on a 8-point Likert scale with 1 labelled as “not at all” and 8 as “absolutely”. Participants were informed about receiving a reward right after the scanning session outside the scanner, for food and art, respectively, based on their highest ratings in that session. The reward was given as a dish to eat right away and as a carton-based art print to take home with.

Preprocessing was done using fMRIPprep 1.2.5 ^[26]^. As preregistered, first-level contrasts of interest were global difference between food and art viewing, food compared to art wanting slope, and wanting modulation (*Design A*), food wanting by caloric or fiber density (*Design B*) and considering liking ratings as modulator (*Design C*). See **SI_fMRI** for further details.

### Additional behavioral assessments

Dietary habits, lifestyle factors including gastrointestinal quality of life, sleep, physical activity, mental well-being and mood was assessed at each timepoint. Additionally, we assessed potential traits associated with food decision-making at baseline, i.e. on personality, eating behavior, anxiety and well-being, as well as on art knowledge (see **SI_behav** for details).

### Blood and feces markers

To assess serum SCFA, gut hormones (ghrelin, GLP-1, PYY), markers of glucose/lipid metabolism (glucose, insulin, HbA1c, high- and low density lipoprotein, HDL/ LDL, triglycerides), inflammatory markers (high sensitive C-reactive protein, CRP, interleukin-6, TNFalpha), and other markers (trimethylamine-n-oxid, TMAO, and amino acids), blood was obtained in fasting state (12.5 ± 2.2 h fasted) at the same time per participant for each session. Stool samples were taken 1-2 days before the testing day to assess fecal SCFA and microbial markers.

### Microbial analysis

For 16S-rRNA gene profiling, DNA was extracted and V3-V4 variable regions of the 16S-rRNA genes were amplified by PCR and a library was constructed, followed by paired-end 2x250bp Illumina sequencing. Raw sequencing data analysis was done on the inhouse Galaxy server using a pipeline implemented with the DADA2 R-package processed data in fastq format ^[27]^. For further details, see **SI_microbiome**.

### Statistical analysis

On a behavioral level, we hypothesized that participant’s wanting ratings scored higher for food compared to art (*H_behav_1*), and that wanting would change after prebiotic intervention *(H_behav_2)*, dependent on caloric density of the food item *(H_behav_3)*. Linear mixed models were performed in R (version >3.6) using lmer(), for a model-of-interest and a null model for each effect of interest. Model residuals were tested for normal distribution using the R package performance() with the command check_normality(x, effects = “random”), see **SI_behav** for details.

On a neural level, we hypothesized that food evaluation elicits different regional brain activation compared to art evaluation (*H_neural_1)*, and that this differential brain response changes after prebiotic intervention *(H_neural_2)*. Inference tests were performed using a homeostatic and reward-related region-of-interest brain mask on first-level contrasts (*Designs A-C*) and second-level factors time (baseline, follow-up), group (prebiotics/placebo), and time*group interactions, using the Sandwich Estimator (SwE v2.2.2, https://fsl.fmrib.ox.ac.uk/fsl/fslwiki/Swe, implemented in SPM 12.7486 run in MATLAB version > 9.0) and R (version >3.6). Significant results were reported according to threshold-free cluster enhancement (TFCE) methods with alpha < 0.05 and family wise error (FWE) correction for multiple comparisons. For details see fMRI preregistration and **SI_fMRI**.

Further effects of prebiotic intervention on microbial and blood and feces markers, as well as potential relations to changes in neural and behavioral markers, were explored using mixed effects inference with one sample t-test on summary statistics as well as using correlation matrices and network analyses. Significance threshold was set at p < 0.05.

### Data and code availability

Data and code are available at https://gitlab.gwdg.de/omega-lab/prebiotic-intervention-on-fmri-food-wanting. All unthresholded MRI TFCE maps are available at https://identifiers.org/neurovault.collection:14111.

## Results

A total of 59 well-characterized overweight/obese adults were included in main analyses (19 women, 40 men, mean age 28 years ± 6.2 SD, BMI 27.3 kg/m^2^ ± 1.4 SD, socioeconomic status, SES, 14.2 ± 3.2; **Table 1, SI_general-Table1**).

**Table 1:**
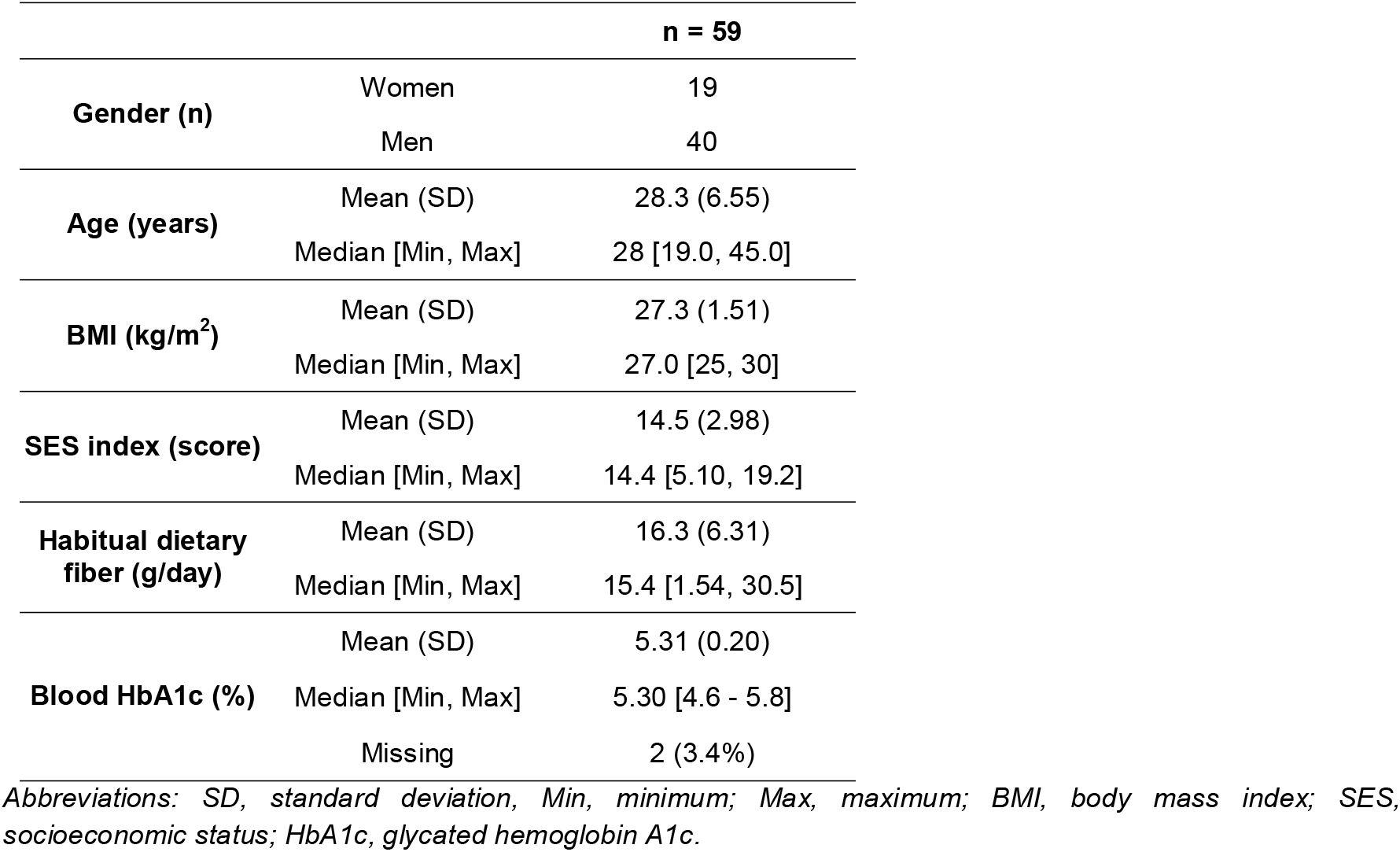
Baseline characteristics.

### Neurobehavioral correlates of reward-related decision-making

Overall, wanting and liking ratings in the fMRI preference task were higher for food than for art stimuli (*H_behav_1;* n_obs_ = 32,111, n_subj_ = 59, b = 1.0, t = 54.8, p < .001, **Fig. 3A-B)**. Food evaluation activated large parts of the reward network, including ventral tegmental area (VTA), hypothalamus, nucleus accumbens (NAc), basal ganglia and ventromedial thalamus, as well as anterior insula, amygdala, cingulate, ventromedial prefrontal cortex (vmPFC) and distinct parts of the orbitofrontal cortex (OFC) (*H_neural_1*, n = 57, *Design A*, p_FWE_ < 0.05; **Fig. 3C**). Similarly, higher wanting ratings for food compared to art elicited higher brain activation ubiquitously across these brain areas, yet particularly in the vmPFC and OFC (*Design A*, p_FWE_ < 0.05; **Fig. 3D**).

**Figure 3:**
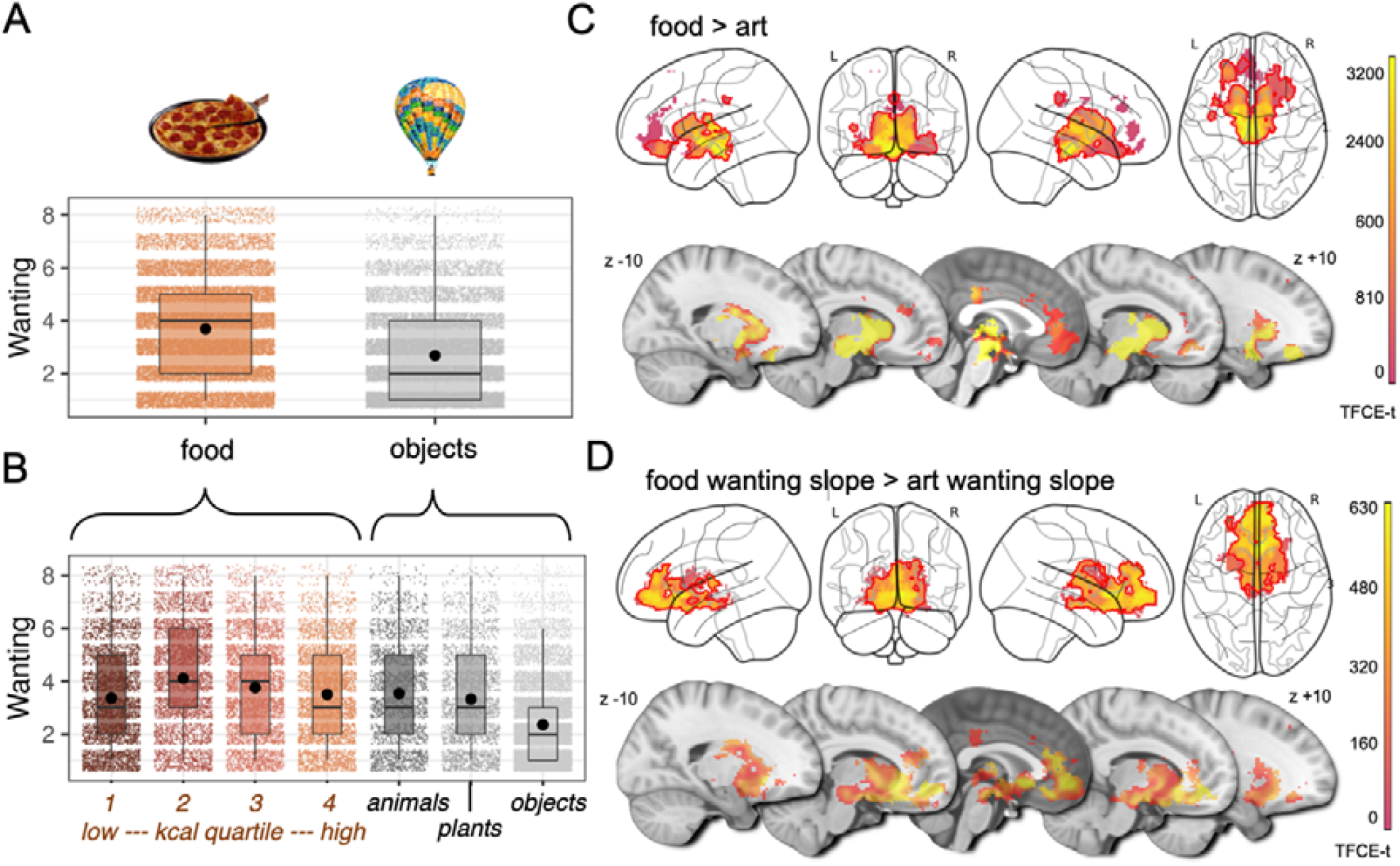
Behavioral (A-B) and neural response (C-D) to food and art stimuli in overweight adults during decision-making. Participants responded to food with higher wanting scores compared to art (n_obs_ = 32,111, n_subj_ = 59) (**A**), showing highest mean values for moderately high caloric stimuli, and lowest mean values for art objects (n_obs_ = 32,111, n_subj_ = 59) (**B**). Food compared to art valuation elicited stronger brain activation particularly in subcortical areas of the reward network (n_subj_ = 57) (**C**), while additional parametric modulation with wanting scores indicated a stronger brain activation in ventromedial prefrontal cortex and orbitofrontal cortex when comparing food vs. art (n_subj_ = 57) (**D**). Statistics were done with linear mixed effect modelling, up to 4 timepoints per participant x 120 stimuli on wanting scores (**A, B**) and on voxel-wise blood-oxygen-level-dependent signal using the sandwich estimator toolbox with threshold-free cluster enhancement (TFCE) family wise-error correction (FWE) of multiple comparisons (**C, D**). Color bars depict parametric TFCE statistic (TFCE-t > 50 for visualization purposes) with wild-boot strapped p-_FWE_ < 0.05 marked in red outline.

### Effect of prebiotics on food decision-making

At the behavioral level, individuals’ overall wanting scores were not different after the two-week prebiotic intervention regarding food vs. art and when accounting for calories or fiber (*H_behav_2+3* ; n_obs_ = 16,071, n_subj_ = 59, b_all_ < |0.00005|, t_all_ < |1.0|; p_all_ > 0.34). Exploratory analysis showed however that prebiotics compared to placebo led to significantly higher wanting scores for *moderately low* and *moderately high* caloric content (n_obs_ = 32,111, n_subj_ = 59, b > 0.03, t > 0.19, p > 0.002, **Fig. 4A**).

**Figure 4:**
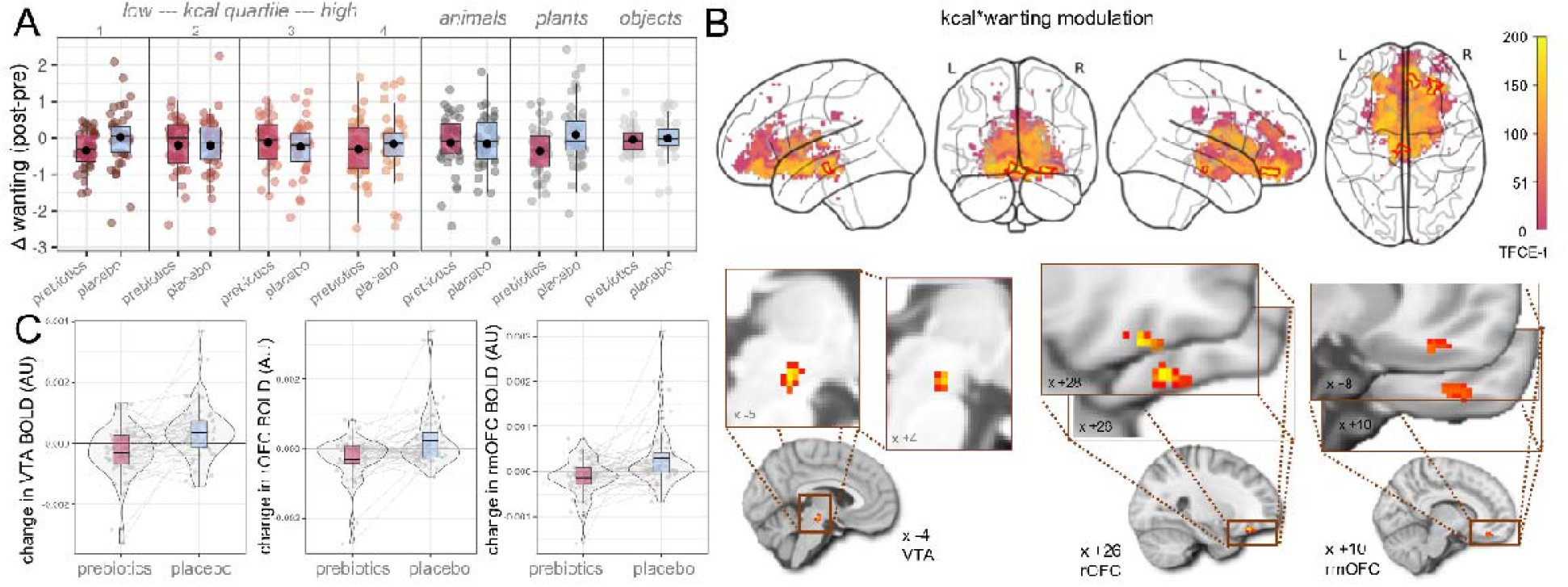
Effects of prebiotic intervention on food decision-making. After the intervention, participants decreased wanting scores for food from caloric quartiles 1 and 4 as well as animals (n_subj_ = 59, **A**). At the neural level, brain activation decreased in the VTA and two clusters in OFC towards high-caloric, wanted food stimuli (n_subj_ = 57, **B, C**). Statistics according to linear mixed effects modelling, up to 4 timepoints per participant x 120 stimuli on wanting scores and on voxel-wise blood-oxygen-level-dependent signal using the sandwich estimator toolbox with threshold-free cluster enhancement (TFCE) family wise-error correction (FWE) of multiple comparisons. Color bars depict parametric TFCE statistic with wild-boot strapped p-_FWE_ < 0.05 marked in red outline (upper righthand panel) and as enlargement (lower righthand panel).

According to fMRI (*H_neural_2), we* did not observe changes in regional brain response after prebiotics in food compared to art viewing, food compared to art wanting slope, or wanting modulation (*Design A*). However, brain activation towards wanted, high-caloric food (*Design B*) decreased after prebiotics compared to placebo in three clusters, in the VTA (p_FWE-corr_ = 0.042), in the right OFC (rOFC, p_FWE-corr_ < 0.05), and in the right medial OFC (rmOFC, p_FWE-corr_ < 0.05) (n = 57, **Fig. 4B-C, Table 2**). In addition, art liking compared to food liking increased in a small cluster in the right NAc after prebiotics compared to placebo (*Design C*, **Table 2**). See **SI_fMRI-Results** for further results.

**Table 2:**
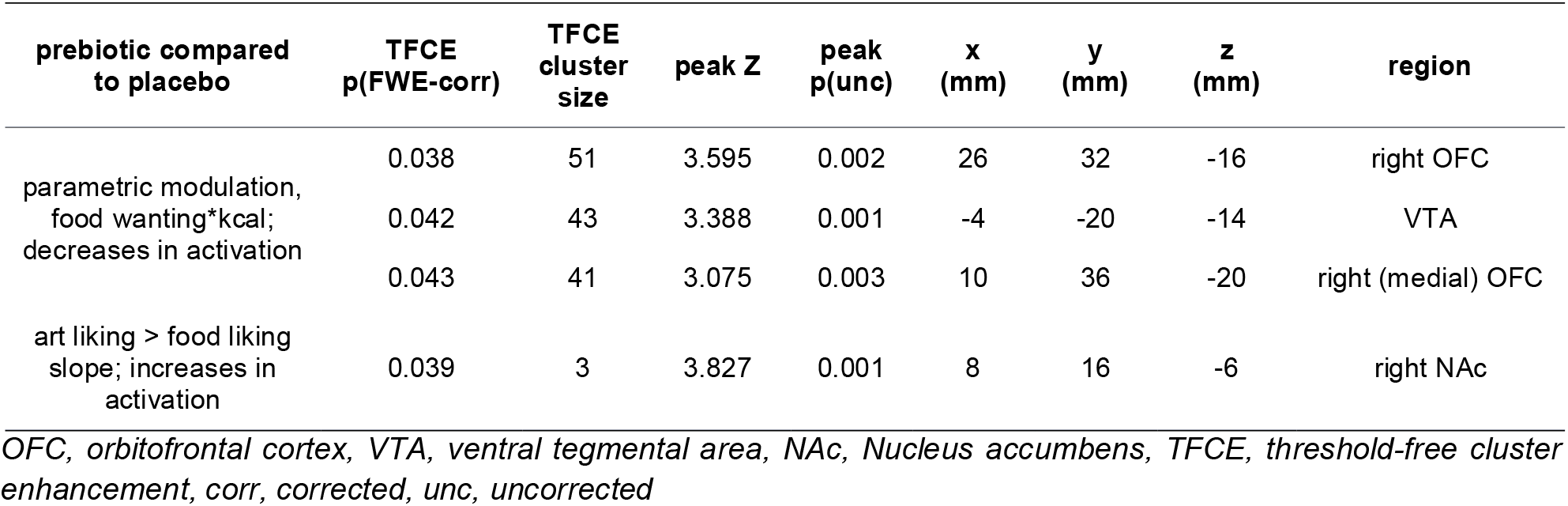
Localization of significant changes in brain activation to visual food and art stimuli during functional MRI, after prebiotic compared to placebo intervention.

| prebiotic compared to placebo | TFCE $p(\text{FWE-corr})$ | TFCE cluster size | peak Z | peak $p(\text{unc})$ | x (mm) | y (mm) | z (mm) | region |
| --- | --- | --- | --- | --- | --- | --- | --- | --- |
| parametric modulation, food wanting*kcal; decreases in activation | 0.038 | 51 | 3.595 | 0.002 | 26 | 32 | -16 | right OFC |
|  | 0.042 | 43 | 3.388 | 0.001 | -4 | -20 | -14 | VTA |
|  | 0.043 | 41 | 3.075 | 0.003 | 10 | 36 | -20 | right (medial) OFC |
| art liking > food liking slope; increases in activation | 0.039 | 3 | 3.827 | 0.001 | 8 | 16 | -6 | right NAc |

OFC, orbitofrontal cortex, VTA, ventral tegmental area, NAc, Nucleus accumbens, TFCE, threshold-free cluster enhancement, corr, corrected, unc, uncorrected

### Anthropometric and blood markers after prebiotics

While both intervention and placebo supplements contained the same amounts of calories and participants reported equally high compliance in taking the daily supplements, we observed decreases in body fat after placebo (timepoint*intervention, b = 0.16, p = 0.01; **Fig. 5A**). In addition, lipid markers were significantly lower after placebo intake compared to prebiotics (b_all_ > 0.25, t_all_ = 2.6, p_all_ < 0.02; **Fig. 5B-C**). BMI, waist-to-hip ratio, blood pressure and subjective hunger ratings during fMRI did not change significantly, which was also true for fasting ghrelin, GLP-1 and PYY, glucose, insulin, amino acids, as well as inflammatory markers (see **SI_behav** and **SI_general 2-5**).

**Figure 5:**
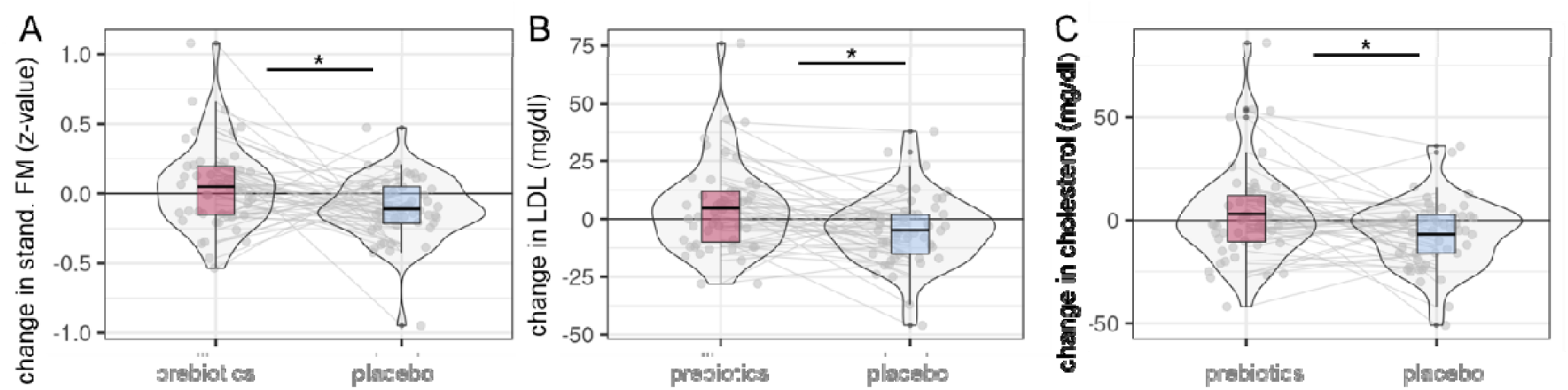
Changes in secondary eating behavior-related outcomes after prebiotic (red) compared to placebo condition (light blue). According to linear mixed effects modelling, gender-standardized body fat mass (FM, **A**), serum lipid markers low-density lipoprotein (LDL, **B**), and cholesterol (CHOL, **C**) significantly decreased after placebo.

In exploratory bivariate correlation analysis on change scores after the prebiotic intervention, mean BOLD activation in the three outlined VTA and OFC clusters decreased in correlation with decreases in fasting PYY (Spearman’s r_all_ > 0.32, p_all_ < 0.05).

### Changes in gut microbiota and parameters

The prebiotic intervention led to increases in stool frequency (b = 1.3, t = 2.0, p = 0.04, **Fig. 6A**). Through 16S-rRNA analysis, we detected significantly decreased richness, evenness and alpha diversity after prebiotics compared to placebo (n_obs_ = 205, n_subj_ = 57, all p < 0.01 **Fig. 6B-D, SI_microbiome_Table1**). Beta diversity on Amplicon Sequencing Variant (ASV) was significantly different after fiber intervention (**Fig. 6E**), and there were abundance changes in families of *Actinobacteria* and *Firmicutes* (all p < 0.02, **Fig. 6F**). Zooming at the genera level, prebiotics induced significant shifts in various abundances, including increases in *Bifidobacteria* (**Table 3**).

**Figure 6:**
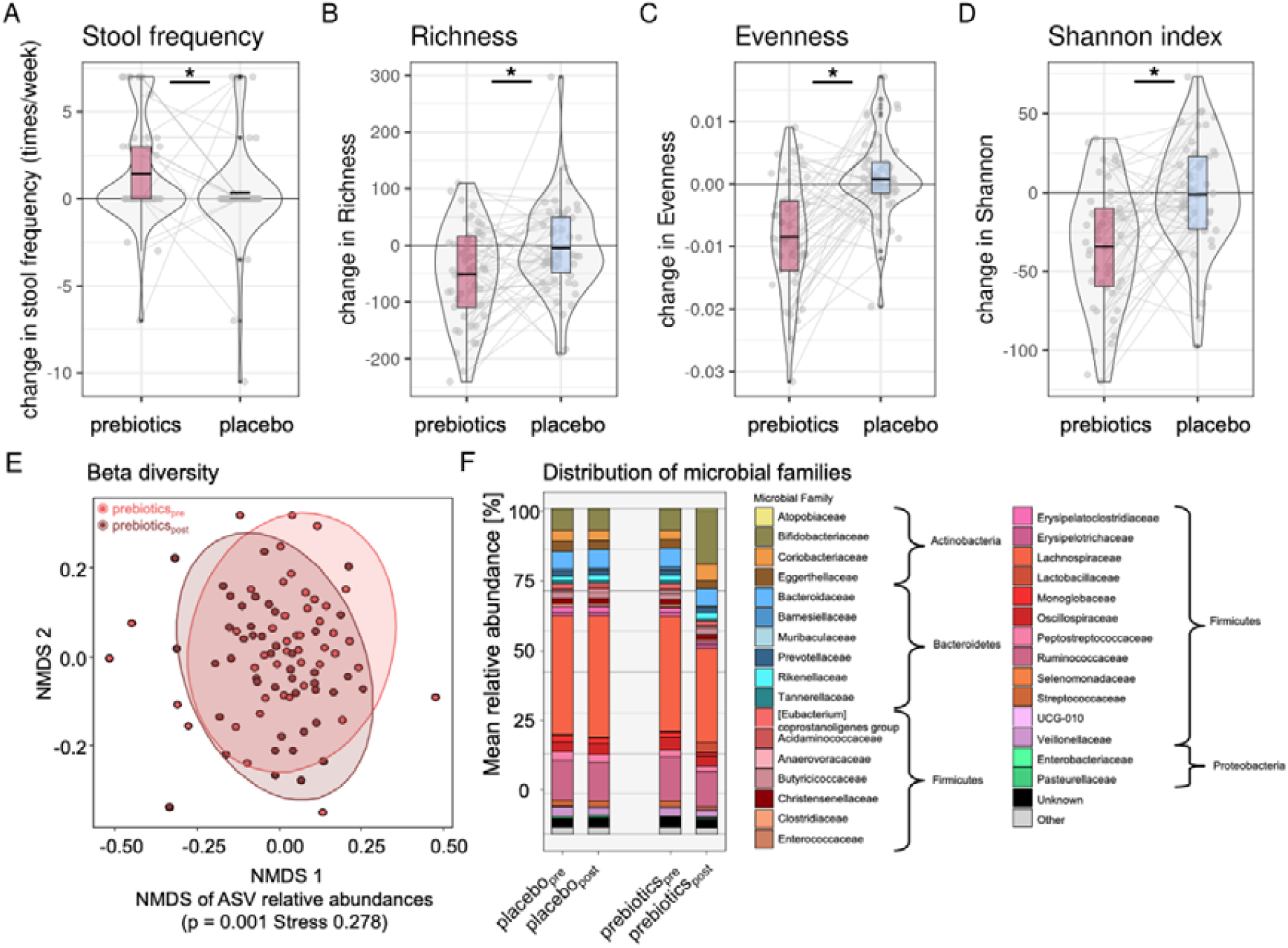
Microbiota-related shifts after two-week prebiotic intervention. Increases in stool frequency (**A**) and decreases in (**B**) microbiota richness, (**C**) evenness, (**D**) Shannon index, (**E**) beta diversity changes compared by dissimilarity gradients and (**F**) shifts in microbial family distribution according to group and timepoint after prebiotics (pink) compared to placebo (blue). Asterisks in (**A)-(D)** indicating significant ANOVA results for null-full model comparisons (p < 0.05). Abbreviations: NMDS: Nonmetric multidimensional scaling

**Table 3:**
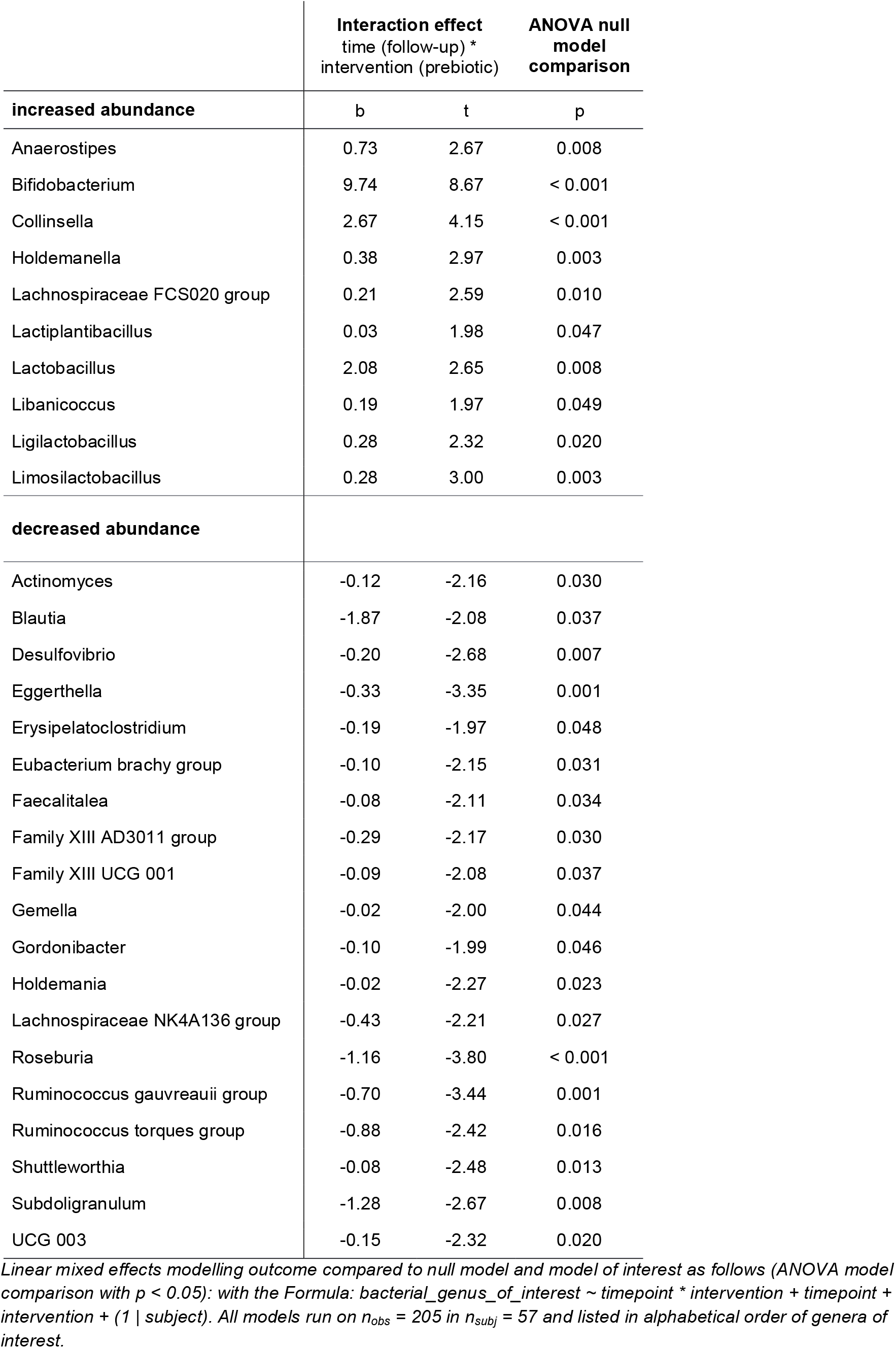
**Significant shifts in microbiota relative abundances on the genera level after prebiotic intervention**, according to 16S-rRNA sequencing and linear mixed effects modelling.

### Changes in microbiota genera link to changes in neurobehavioral outcomes

We further explored whether the observed changes in microbial genera predicted intervention-induced changes in neurobehavior. Accordingly, a less severe decrease in *Subdoligranulum* correlated with intervention-induced decreases in VTA brain activation towards wanted, high caloric food stimuli after prebiotic intervention (r = -0.42, p < 0.01 **Fig. 7A**). Note that bacterial abundance was measured in percentage, thus a relative decrease in *Subdoligranulum* does not necessarily display absolute decrease after prebiotics.

**Figure 7:**
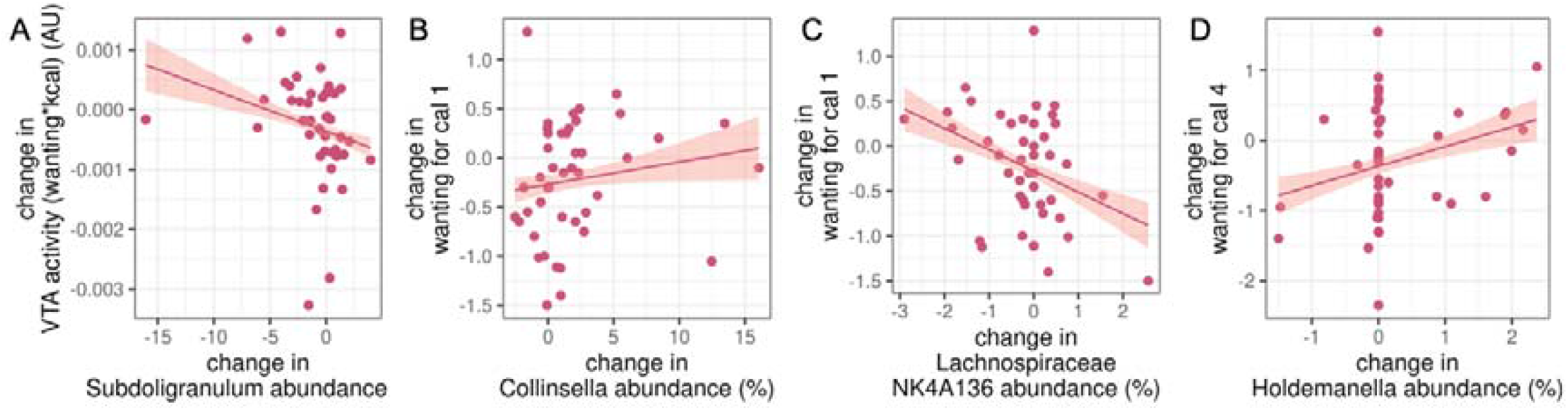
Correlations between significant shifts in genera abundance and reward-related brain activation. Spearman correlation plots of significantly changes in microbial measures: (**A**) *Subdoligranulum* abundance and VTA activity, (**B**) *Collinsella* abundance and wanting for very low caloric food items, (**C**) *Lachnospiracea* abundance and wanting for very low caloric food items, (**D**) *Holdemanella* abundance and wanting for very high caloric food items.VTA, ventral tegmental area, cal, caloric quartile

Additionally, increases in *Lactiplantibacillus* (lactic acid producing bacteria) were significantly related to increases in rmOFC activation (r = 0.30, p > 0.05), however abundance of this bacterium did not change in most participants. Changes in wanting ratings showed significant correlations between wanting for lowest caloric stimuli positively for *Collinsella* abundance and negatively for *Lachnospiraceaca NK4A136 group* abundance and wanting for highest caloric items positively with *Holdemanella* abundance (r_all_ > | 0.32 |, p_all_ < 0.05, **Fig. 7B-D**). Complementary weighted network analyses in a subgroup of available participant data from all four timepoints (n = 35) did not provide compelling evidence that clusters of microbial taxa related to neurobehavioral outcomes (**SI_microbiome)**.

### SCFA and microbial functional capacity prediction

We could not detect changes in SCFA acetate, butyrate and propionate after intervention, neither in fasting serum nor in fecal concentrations (n_obs_ ≥ 122, n_subj_ ≥ 40, p_all_ > 0.39, **Table 3**).

**Table 3:** Concentrations of feces and serum SCFA levels.

|  | total (μmol/g) |  |  |  |  |  | butyrate (μmol/g) |  |  |  |  |  | acetate (μmol/g) |  |  |  |  |  | propionate (μmol/g) |  |  |  |  |  |
| --- | --- | --- | --- | --- | --- | --- | --- | --- | --- | --- | --- | --- | --- | --- | --- | --- | --- | --- | --- | --- | --- | --- | --- | --- |
|  | n | pre, mean ± SD | n | post, mean ± SD | change (%) | p | n | pre, mean ± SD | n | post, mean ± SD | change (%) | p | n | pre, mean ± SD | n | post, mean ± SD | change (%) | p | n | pre, mean ± SD | n | post, mean ± SD | change (%) | p |
| **feces** |  |  |  |  |  |  |  |  |  |  |  |  |  |  |  |  |  |  |  |  |  |  |  |  |
| prebiotics | 42 | 26.4 ± 10.9 | 40 | 24.6 ± 8.5 | -6.8 | 0.39 | 42 | 18.3 ± 8.7 | 41 | 16.8 ± 8.0 | -8.2 | 0.73 | 42 | 7.9 ± 2.7 | 41 | 8.1 ± 2.4 | 2.5 | 0.76 | 42 | 7.1 ± 2.4 | 41 | 6.2 ± 3.2 | -12.7 | 0.65 |
| placebo | 42 | 23.9 ± 8.8 | 42 | 23.2 ± 12.2 | -2.9 |  | 42 | 16.2 ± 6.9 | 42 | 15.5 ± 9.4 | -4.3 |  | 42 | 7.4 ± 2.4 | 42 | 7.4 ± 3.2 | 0.0 |  | 42 | 7.1 ± 3.1 | 42 | 6.6 ± 3.4 | -7.0 |  |
| **serum** |  |  |  |  |  |  |  |  |  |  |  |  |  |  |  |  |  |  |  |  |  |  |  |  |
| prebiotics | 27 | 5.8 ± 1.3 | 27 | 5.5 ± 1.5 | -5.2 | 0.65 | 36 | 0.5 ± 0.3 | 34 | 0.5 ± 0.4 | 0.0 | 0.66 | 30 | 3.1 ± 3.2 | 32 | 3.8 ± 3.0 | 22.6 | 0.91 | 28 | 0.5 ± 0.5 | 24 | 0.6 ± 0.6 | 20.0 | 0.84 |
| placebo | 29 | 5.5 ± 1.4 | 25 | 5.3 ± 1.5 | -3.6 |  | 34 | 0.5 ± 0.4 | 37 | 0.4 ± 0.3 | -20.0 |  | 32 | 2.4 ± 2.3 | 28 | 3.1 ± 3.2 | 29.2 |  | 27 | 0.4 ± 0.4 | 28 | 0.4 ± 0.4 | 0.0 |  |

P, according to ANOVA null-full model p-value, total = sum of butyrate, acetate and propionate.

Next, we explored changes induced by microbial shifts on the metagenomic level according to Kyoto Encyclopedia of Genes and Genomes (KEGG) ^[28]^ analysis. Changes in KEGG orthologue relative abundance were significantly different after prebiotics (**Fig. 8A**, NMDS: prebiotics p = 0.001, placebo p = 0.99). The KEGG orthologues were annotated to 158 pathways out of which about 50%, i.e. 79, were significantly altered in relative abundance after prebiotic intervention compared to placebo, including pathways related to carbohydrate, protein and fat metabolism, plant degradation or cell repair (p_all_ < .05, **SI_microbiome_Table 2**).

**Figure 8:**
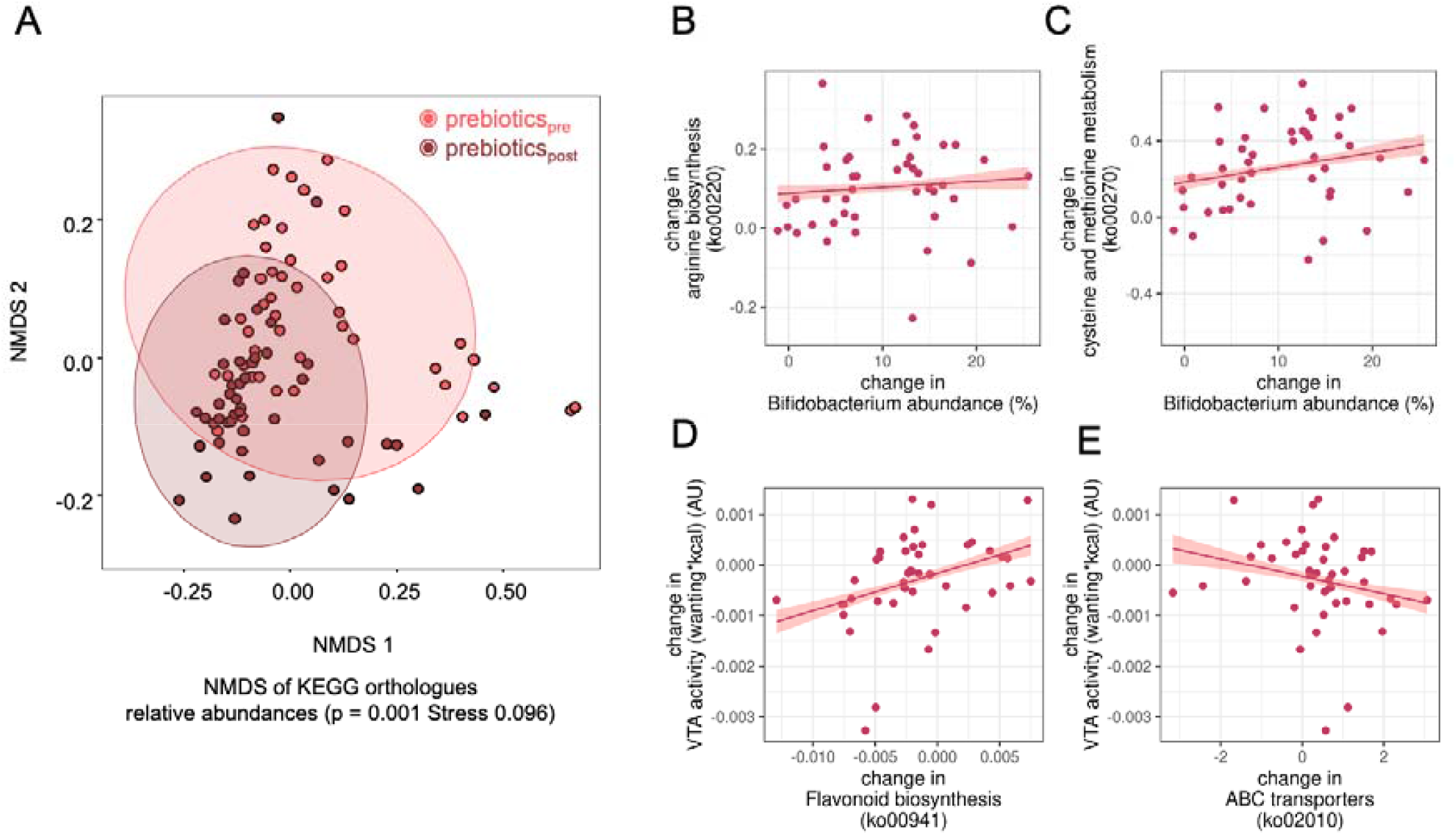
Predicted functional shifts and their correlations with changes in microbiota genera and in reward-related brain activation after prebiotic intervention. (**A**) Dissimilarity of functional composition of microbiome pre- to versus post-prebiotic intervention based on principal component analysis of relative abundance of predicted KEGG orthologues statistics, calculated by PERMANOVA (p = 0.001). (**B**) Changes scores of bifidobacterium abundance and arginine biosynthesis (ko00220), (**C**) Bifidobacterium abundance and cysteine and methionine metbolism (ko00270), (**D**) flavonoid biosynthesis (ko00941) and changes in reward-elated brain response, (**E**) stilbenoid, gingerol biosynthesis (ko00945) and reward-related brain response. B-E, all r > 0.32, all p < 0.05 according to Spearman’s correlation, line gives regression fit with 95% confidence interval.

More specifically, increases in relative abundance of *Bifidobacteria* correlated significantly with increases in metabolic pathways related to taurine, seleno-compounds, nicotinate, and amino acids, and with decreases related to porphyrin metabolism, steroid degradation, (unsaturated) fatty acid biosynthesis, and DNA repair functions (exemplary **Fig. 8B-C**; Spearman’s r_all_ > 0.32, p_all_ < 0.05). In addition, increases in *Lactobacillus* and decreases in *Gordonibacter* correlated with increases in pyruvate metabolism pathway, a precursor of SCFA (note that not all participants changed in *Lactobacillus* and *Gordonibacter* abundance, though). Further exploratory analyses indicated that decreases in VTA brain activation after prebiotic intervention correlated with intervention-induced significant decreases in pathways involved in flavonoid and stilbenoid biosynthesis, two-component signal transduction, biofilm formation, amino sugar and nucleotide sugar metabolism, citrate cycle (r_all_ > 0.37, p_all_ < 0.05), and with significant increases in ATP-binding cassette transporters (ABC, r = -0.39, p < 0.05, exemplary **Fig. 8D-E**). Decreases in rOFC activation after prebiotics correlated with significant decreases in caprolactam degradation (r = 0.33, p < 0.05). Decreases in rmOFC activation after prebiotics correlated with significant increases in oxidative phosphorylation (r = -0.31, p < 0.05). For details see **Fig. 9, SI_microbiome_Table3**.

**Figure 9:**
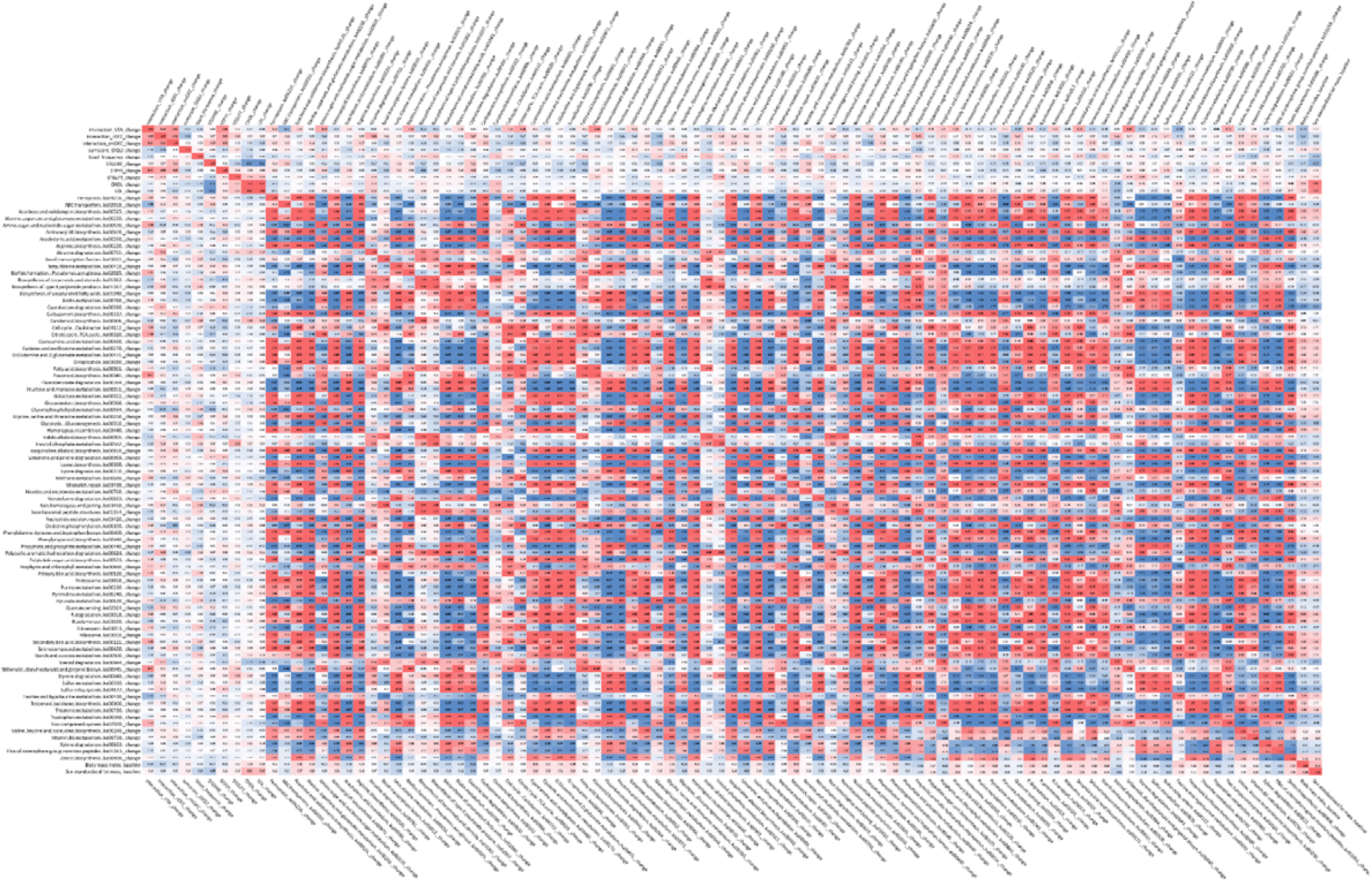
Heatmaps of bivariate correlations between changes in reward-related brain activation and changes in blood and microbial markers. Left-hand triangle: brain activation, blood markers and predicted microbial functional pathways. Right-hand triangle: brain activation, blood markers and microbiota genera. Color according to Spearman’s r, red, positive correlations, blue, negative correlations, bold: p < 0.05, italic, p < 0.01. VTA, ventral tegmental are, OFC, orbitofrontal cortex, r, right, m, middle.

## Discussion

In this proof-of-concept study, we tested the effects of a prebiotic intervention on food decision-making in a randomized within-subject cross-over design including 59 well-characterized, overweight adults. In pre-registered analyses, we found that 14 days of high-dose dietary prebiotics, compared to placebo, led to decreases in BOLD-related brain activation towards high caloric, wanted food in the VTA and right OFC measured using 3T fMRI. In parallel, prebiotics led to significant shifts in relative abundance of the gut microbiota, including increases in SCFA-producers such as *Bifidobacteria, Lactobacillus*, and *Collinsella*. Intervention-induced changes in relative abundance and predicted metabolic pathways correlated with changes in VTA brain activation. While fasting gut hormones, inflammatory markers and SCFA in blood and feces remained unchanged, we observed that prebiotics-induced decreases in brain activation in reward areas related to decreases in fasting PYY.

### Changes in functional brain activation

Only few studies with moderate sample size have addressed whether manipulating the microbiome can alter brain functions. A parallel trial in 34 females indicated that four weeks of fermented milk consumption (including *Bifidobacteria*) induced resting-state functional connectivity changes in the midbrain ^[29]^. Another randomized trial reported that four weeks of probiotic supplementary powder containing *Bifidobacteria* and *Lactobacillae* resulted in changes in microbial genera abundance that correlated with improvements of emotional attention and memory, paralleled by differences in related brain activation ^[30]^. Our findings now present prebiotics-induced changes in brain function with implications for food craving and decision-making: Neural responses within VTA and OFC have been suggested to underly dopamine-related reward anticipation and subjective value attribution of food, respectively, linking stronger BOLD-related activation to higher reward values and decision-making ^[31]^. Indeed, midbrain and medial OFC activation during fMRI in response to milkshake taste predicted the amount of milkshake intake after the scan ^[32]^.

While the neuronal processes underlying human eating behavior are far from fully understood, neuroimaging studies indicate that drivers of reward considering food (such as caloric content) modulate subjective value particularly in the OFC ^[33]^, and the right OFC has been specifically implicated in food-related motivation ^[34]^. Notably, decreases in brain activation towards high caloric food cues such as ice-cream in the OFC has for example been shown using fMRI when participants were instructed to consider health aspects or long-term consequences of consumption, compared to ‘naive’ viewing ^[35]^. The intervention-induced decreases in VTA and rOFC in the current study might thus indicate a diminished anticipation of reward, and a smaller subjective value attribution to high-caloric wanted foods after prebiotic treatment, potentially translating in a subtle reduction of the desire for high-caloric food. While we could not confirm a general reduction in food wanting at the behavioral level, exploratory analysis of less wanting of very high caloric food supports this hypothesis.

### Microbiota-related mechanisms

For example, the gut microbiome has only recently been shown to be relevant for host nutritional foraging in rats, e.g., through changing circulating amino acids and bacterial tryptophan ^[36]^. Another fecal transplantation rat study indicated that microbiota from obese donors resulted in changes in food preference and expression of dopaminergic markers in the striatum ^[37]^. In humans, a single-group study in 26 females suggested that increased consumption of vegetables rich in inulin-type fructans over two weeks increased *Bifidobacteria* and decreased the desire to eat sweet, salty, and fatty food ^[38]^. In the current study, we similarly observed changes in multiple bacterial genera abundances after prebiotics compared to placebo, mainly increases in Actinobacteria phylum (*e*.*g*., *Bifidobacteria*) and Firmicutes phylum (e.g. *Lactobacillus*). This suggests a marked increase in fiber-degrading, SCFA producing bacteria that are present in the gut, which is in line with previous human trials ^[16, 39, 40, 41, 42]^.

Functional capacity prediction analyses further yielded a multitude of different pathways that were selectively changed after prebiotic intervention, among them pathways involved in SCFA production capable to modify systemic SCFA signaling. For example, one of the most strongly upregulated pathways related to the ABC transporters (ko02010). It has been shown that *Lactobacillus* use dietary fiber (e.g. inulin) via ABC transporters to produce acetate ^[43]^, which can be further degraded to butyrate ^[44]^. Multiple of the upregulated microbiota genera after prebiotics have been classified to produce SCFA, e.g. *Anaerostipes, Bifidobacterium*, and *Holdemanella* ^[45]^. Moreover, pointing to a dose-effect relationship, a less severe decrease in relative *Subdoligranulum* abundance (also SCFA producer), as well as the increases in prebiotics-induced upregulation of ABC transporters, correlated with significant decreases in prebiotics-induced VTA brain activation, suggesting a potential mechanistic route of higher SCFA production leading to lessened reward anticipation.

In contrast to our *a priori* hypothesis, we did not observe changes in fecal or fasting blood levels of acetate, butyrate or propionate. Similarly, previous small scale studies could not show increases in fecal SCFA after inulin, e.g. in healthy young adults (n = 49) ^[46]^. Others observed SCFA increases, e.g. in type-2 diabetes mellitus (n = 25, ^[47]^), or even decreases in fecal SCFA (n = 30) [40]. These conflicting results might be explained by unknown complexity of local and systemic microbial effects, and/or by pre-existing differences such as microbiota patterns at baseline (note higher relative *Firmicutes* in our overweight group compared to obesity studies), or differences in stool frequency, weight and fluidity (note significant changes in Bristol stool scale after prebiotics in the current study).

Body fat and lipid markers slightly improved after placebo condition and worsened after prebiotics in the current study. While we did not observe changes in lifestyle habits according to questionnaires, beneficial effects of e.g. increased energy expenditure in the placebo phase cannot be ruled out. Also, inulin might challenge liver cholesterol metabolism, as postulated in mice under certain conditions ^[48]^. For serum SCFA, others did find short-term increases in SCFA after inulin ^[49]^, and the postprandial increase in SCFA correlated with decreases in serum ghrelin ^[50]^. Prebiotics and SCFA also stimulate the expression of PYY and GLP-1 in the gut ^[51]^, that may contribute to changes in central reward-related food responses. In humans, PYY injections induced changes in BOLD-related fMRI signaling in the hypothalamus, VTA and OFC ^[52, 53]^. We found that decreases in fasting PYY correlated with decreases in brain activation in the VTA and OFC clusters after intervention, pointing to a similar mechanisms. However, a (postprandial) increase in serum SCFA or gut hormones due to prebiotics in our sample might have been masked after overnight fasting.

### Limitations

Our study should be discussed in light of several limitations. First, 14 days of intervention can be considered too short to induce long-lasting effects on neuronal processes involved in eating behavior. Still, advanced neuroimaging techniques and in-depth pre-registration of statistical thresholding ensured confidence in the robustness of the observed effects. Second, KEGG analyses need to be considered indirect only and microbiome samples were not time-locked to MRI sessions. Due to the within-subject crossover design, however, interindividual differences at baseline determining microbiota responses could be kept to a minimum. Also, participants belonged to a Western, Educated, Industrialized, Rich and Democratic (WEIRD) society and we did not recruit representative shares of female and diverse gender, limiting generalizability of results difficult.

### Conclusions

According to pre-registered RCT analysis of advanced 3T-fMRI, this proof-of-concept study suggests that a high-dosed microbiome-changing prebiotic intervention decreases neurobehavioral food craving within two weeks in overweight adults. Based on 16S-rRNA combined with functional pathway prediction and metabolomics, our findings lend support for a mechanistic link between prebiotic dietary intake, related increases in SCFA production and potentially PYY as underlying mechanisms to lessen reward-related brain activation during food-decision making. Neural response in reward-related areas during fMRI are predictive for behavior change ^[54]^, underlining implications for the treatment of unhealthy eating behaviors or overnutrition using microbiome-changing interventions. Future studies are needed to confirm these results in patients living with obesity and to assess clinical utility using longer intervention periods in combination with tailored intervention strategies.

## Supporting information

SI_general

SI_fMRI

SI_microbiome

SI_microbiome_Table3

SI_behav

## Data Availability

Data and code are available at https://gitlab.gwdg.de/omega-lab/prebiotic-intervention-on-fmri-food-wanting. All unthresholded MRI TFCE maps are available at https://identifiers.org/neurovault.collection:14111. Additional supplementary data of the present study are available upon request to the authors.

https://identifiers.org/neurovault.collection:14111

## Acknowledgments

We thank all the individuals who took part in the study. This work was supported by grants of the German Research Foundation (DFG), contract grant number 209933838 CRC1052-03 A1 to A.V.W and M.S., and by the Berlin School of Mind and Brain (stipend for E.M.) and the German Foundation for Environment (stipend for E.M.). The inulin supplement was sponsored by the manufacturer BENEO GmbH, Mannheim, Germany. For participant support we thank Maria Dreyer, Ramona Menger, Bettina Johst and Susan Prejawa and for technical support at the MRI we thank all MTAs and specifically Nicole Pampus, Sylvie Neubert, Mandy Jochemko, Anke Kummer and Domenica Klank, and Torsten Schlumm. For fMRI analysis support we thank Hannah Sophie Heinrichs. For lab support we thank Laura Hesse, Charlotte Wiegank, Lina Eisenberg, Emmy Töws and Anna-Luise Wehle. For all other data collection we highly appreciate the support of all our interns and student assistants Leonie Disch, Lukas Recker, Emira Shehabi, Niklas Hlubek, Lynn Mosesku, Larissa de Biasi, Hannah Stock, Lennard Schneidewind, Christian Schneider and Anne-Kathrin Brecht. We thank Lorenz Lemcke and Anna Bujanow for medical assistance. For SCFA analysis we thank Beatrice Engelmann and for blood analysis we thank Madlen Reinicke. For help with formatting and project coordination we thank Silke Friedrich.

## Author information

*Contributions*. Study conceptualization and design: EM, MvB, MS, AV, AVW. Code for tasks: RT. Data collection: EM, RT. Behavioral and fMRI analysis: EM, FB, AVW. Serum analysis: MR. Microbiome analysis: SBH. Metabolomics analysis: URK. Network analysis: SBH, EM, AV, RC. Manuscript draft: EM, AVW. All authors agreed on the content of the material.

