## Supplementary material for "A prebiotic diet changes neural correlates of food decision-making in overweight adults: a randomized controlled within-subject cross-over trial": SI_general

**Study details**

Data acquisition took place between 2019 and 2022 with some breaks due to lockdown regulations during the Sars-Cov2-pandemic). All participants reported omnivorous, non-restrictive eating habits.

At all four test measurements, time-of-day for each participant was standardized to 07:15AM, 08:00AM, 09:15AM, 10:30AM or 11:15AM, respectively. Participants were instructed to come in overnight fasted.

Participants underwent task-based fMRI in a semi-satiated state after receiving a standardized protein shake (mean 159 kcal ± 10 SD for women, mean 206 kcal ± 16 SD for men) subsequent to overnight fasting (12.5 h ± 2.2 SD).

The protein shake comprised a plant-based drink with 10% of energy need (Harris and Benedict 1918) based on protein powder (Vegan Protein Neutral, Foodspring, Berlin, Germany) and oat drink (EDEKA BIO Hafer Drink Classic Vegan, Germany). The following formulas were used to provide the drink:

Energy basal metabolic rate(men) = 66.473+13.752xbodyweight[kg]+5.3xbodyheight[cm]-6.755xage[years]

Energy basal metabolic rate (women) = 655.96+9.563xbodyweight[kg]+1.85xbodyheight[cm]-4.676xage[years]

**Intervention details**

Participants were instructed to take one sachet in the morning and one at lunchtime in any preferred form. Compliance scores were not different for each of the supplements over two weeks or 48h before the follow-up appointment (full-null model comparison, both b < -0.5, p > .09).

**Blood parameters**

Blood drawing was done using safety-multifly needles (21G, 200 mm) and BD Vacutainer Multiple Sample Luer Adapter and different monovettes (2x S-Monovette 9 ml Z-Gel, S-Monovette 2.7 ml FE for glucose, S-Monovette 2.7 ml K3E for whole blood, Greiner VACUETTE® TUBE 2.5 ml CAT Serum Separator Clot Activator for gut hormones). Gut hormones were collected in a 2.5 ml tube with instantly added inhibitors (25 μl DPP-IV inhibitor, Merck, Germany; 25 μl of dissolved Pefabloc ® SC (AEBSF), Roche, Germany), 30 min waiting time and then centrifuged with the other tubes. Blood samples were centrifuged at 3500 rpm at 7° C for 6 min and serum was aliquoted within 1 h of obtainment. Processed aliquots were stored at -80° C within 1 h of collection and further analyzed in one batch per marker. Analyses were conducted at Synevo Studien Service Labor GmbH c/o IMD Institut für Medizinische Diagnostik Berlin-Potsdam GbR, Berlin, Germany and the Institute for Laboratory Medicine, Clinical Chemistry and Molecular Diagnostics (ILM) Leipzig University, Leipzig, Germany. Measurements beyond the lower detection threshold were set to half of the value of the lower bound (e.g. for hCRP if lower bound is <0.30, then value set to 0.15). Biologically implausible values were excluded from the analysis (in total 3 values: TMAO > 1000 ng/ml, ghrelin > 1250 pg/ml, CRP > 85 mg/l).

**Baseline characteristics**

While participants were considered healthy without clinical diagnosis of metabolic disorder at screening, a moderate proportion showed signs of impaired glucose tolerance (n = 10, insulin > 25mlU/l or glucose > 5.6mmol/l or glycated hemoglobin A1c, HbA1c, > 5.7) or hyperlipidemia (n = 17, total cholesterol > 250 mg/dl or low-density lipoprotein, LDL, > 130 mg/dl or triglycerides > 150 mg/dl), based on fasted blood levels at first baseline sessions (**SI Table 1** and **SI Fig. 1**). Few participants were on regular medication (anti-hypertensives: n = 1, L-thyroxine: n = 1, asthma medication: n = 1), and more than half on certain testing days only (n = 35 including painkillers (NSAIDs) or vitamins, in one case a single-dose antibiotic leading to drop-out), all females were on hormonal contraception (pill: n = 11, IUD: n = 1, vaginal ring: n = 1, NA: n = 6).

SI-Table 1: Questionnaire's baseline characteristics at study timepoint T0.

|  | n = 59 | Mean (SD) | Median [Min, Max] |
| --- | --- | --- | --- |
| Barrat Impulsiveness Scale (BIS) | total | 31.0 (5.72) | 31.0 [19.0, 45.0] |
|  | attentional | 8.76 (2.17) | 9.00 [5.00, 14.0] |
|  | motor | 10.8 (2.67) | 10.0 [6.00, 17.0] |
|  | non-planning | 11.4 (2.60) | 12.0 [6.00, 17.0] |
| Eating Disorder Examination Questionnaire (EDEQ) | Eating Concern | 0.18 (0.27) | 0.20 [0, 3.60] |
|  | Restraint | 0.52 (0.69) | 0.20 [0, 3.60] |
|  | Shape Concern | 1.08 (0.98) | 0.81 [0, 3.50] |
|  | Weight Concern | 0.87 (0.89) | 0.60 [0, 3.60] |
| Three Factor Eating Questionnaire (TFEQ) | hunger | 4.29 (3.23) | 4.00 [0, 13.0] |
|  | cognitive restraint | 5.47 (3.68) | 5.00 [0, 13.0] |
|  | disinhibition | 5.41 (2.44) | 5.00 [1.00, 11.0] |
| Big Five Personality Questionnaire (NEO-FFI) | neuroticism | 1.38 (0.60) | 1.38 [0.17, 2.92] |
|  | extraversion | 2.43 (0.59) | 2.46 [1.00, 3.58] |
|  | openness | 2.67 (0.49) | 2.71 [1.42, 3.67] |
|  | agreeableness | 2.63 (0.53) | 2.58 [1.25, 3.83] |
|  | conscientiousness | 2.67 (0.49) | 2.71 [1.42, 3.75] |
| State-Trait Anxiety and Depression Inventory (STADI-T) | trait-dystymia | 7.43 (2.42) | 7.00 [5.00, 15.0] |
|  | trait-emotionality | 8.83 (2.04) | 9.00 [5.00, 13.0] |
|  | trait-euthymia | 15.3 (3.28) | 15.0 [6.00, 20.0] |
|  | trait-worry | 9.17 (3.08) | 9.00 [5.00, 19.0] |
| Vienna Art Interest and Art Knowledge (VAIAK) | total | 34.4 (14.5) | 31.0 [13.0, 67.0] |
| World Health Organisation (WHO)-5 Well being | total | 15.2 (4.85) | 15.0 [3.00, 24.0] |
| Eurohis well-being | total | 31.9 (5.02) | 31.5 [16.0, 40.0] |
| Beckett Depression Inventory (BDI) | total | 4.05 (4.24) | 3.0 [0, 21.0] |
| Smoking status | Non-Smoker  Smoker  Missing | 51 (86.4%)  7 (11.9%)  1 (1.7%) |  |
| Mode of feeding as a child | Bottle-fed  Brest-fed  Unknown  Missing | 5 (8.5%)  46 (78%)  7 (11.9%)  1 (1.7%) |  |
| Mode of birth | Cesarian  Vaginal  Unknown  Missing | 8 (13.6%)  47 (79.7%)  3 (5.1%)  1 (1.7%) |  |

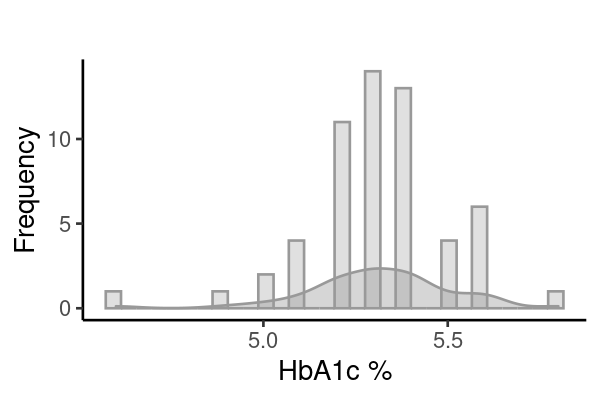

**SI-Fig. 1: Distribution of serum glycated hemoglobin A1c (HbA1c) levels (%)** at baseline**.**

**Results – Descriptives for change in anthropometrics and biomarkers.**

SI-Table 2: Anthropometric markers for all timepoints by intervention condition.

|  | **fiber** | | **placebo** | |
| --- | --- | --- | --- | --- |
|  | **BL** | **FU** | **BL** | **FU** |
|  | **(N=55)** | **(N=47)** | **(N=53)** | **(N=49)** |
| **BMI [kg/m^2^]** |  |  |  |  |
| Mean (SD) | 27.2 (1.50) | 27.3 (1.62) | 27.4 (1.61) | 27.3 (1.67) |
| Median [Min, Max] | 27.1 [24.5, 30.2] | 27.2 [24.2, 30.6] | 27.3 [25.0, 31.2] | 27.1 [24.9, 31.7] |
| **Fat mass [%]** |  |  |  |  |
| Mean (SD) | 26.2 (6.49) | 26.2 (6.24) | 27.0 (6.66) | 26.0 (6.46) |
| Median [Min, Max] | 24.8 [7.59, 38.5] | 24.9 [10.6, 39.0] | 26.7 [9.53, 41.6] | 25.2 [7.76, 38.9] |
| Missing | 0 (0%) | 0 (0%) | 1 (1.9%) | 0 (0%) |
| **Fat mass gender-standardized [%]** |  |  |  |  |
| Mean (SD) | -0.0671 (0.975) | -0.00729 (0.942) | 0.109 (1.05) | -0.0727 (1.03) |
| Median [Min, Max] | -0.0108 [-3.48, 2.41] | -0.154 [-2.82, 2.51] | 0.0217 [-3.05, 2.54] | -0.156 [-3.44, 2.40] |
| Missing | 0 (0%) | 0 (0%) | 1 (1.9%) | 0 (0%) |
| **Fat-free mass gender-standardized [kg]** |  |  |  |  |
| Mean (SD) | -0.0269 (1.00) | -0.00723 (0.953) | -0.0212 (0.973) | 0.0546 (1.02) |
| Median [Min, Max] | -0.220 [-1.94, 4.47] | -0.0759 [-2.11, 3.18] | -0.198 [-2.13, 3.80] | -0.238 [-2.18, 4.02] |
| Missing | 0 (0%) | 0 (0%) | 1 (1.9%) | 0 (0%) |
| **Waist-to-hip ratio** |  |  |  |  |
| Mean (SD) | 0.820 (0.0540) | 0.816 (0.0618) | 0.821 (0.0563) | 0.814 (0.0550) |
| Median [Min, Max] | 0.811 [0.700, 0.942] | 0.816 [0.694, 0.970] | 0.824 [0.686, 0.980] | 0.809 [0.712, 0.981] |
| **10% of daily energy requirement [kcal]** |  |  |  |  |
| Mean (SD) | 191 (26.2) | 194 (26.8) | 193 (26.0) | 192 (26.6) |
| Median [Min, Max] | 194 [142, 249] | 196 [141, 245] | 194 [141, 246] | 194 [141, 247] |
| Missing | 0 (0%) | 1 (2.1%) | 0 (0%) | 0 (0%) |

SI-Table 3: Serum markers for all timepoints by intervention condition.

|  | **fiber** | | **placebo** | |
| --- | --- | --- | --- | --- |
|  | **BL** | **FU** | **BL** | **FU** |
|  | **(N=55)** | **(N=47)** | **(N=53)** | **(N=49)** |
| **Time fasted [h]** |  |  |  |  |
| Mean (SD) | 12.5 (2.25) | 12.3 (1.70) | 12.5 (2.25) | 12.3 (1.70) |
| Median [Min, Max] | 12.3 [6.00, 18.0] | 12.0 [6.50, 15.0] | 12.3 [6.00, 18.0] | 12.0 [6.50, 15.0] |
| Missing | 1 (1.8%) | 0 (0%) | 1 (1.8%) | 0 (0%) |
| **Triglycerides [mg/dl]** |  |  |  |  |
| Mean (SD) | 103 (47.3) | 98.0 (47.5) | 103 (47.3) | 98.0 (47.5) |
| Median [Min, Max] | 92.0 [40.0, 285] | 89.0 [26.0, 229] | 92.0 [40.0, 285] | 89.0 [26.0, 229] |
| Missing | 0 (0%) | 0 (0%) | 0 (0%) | 0 (0%) |
| **Cholesterol [mg/dl]** |  |  |  |  |
| Mean (SD) | 170 (27.9) | 173 (41.8) | 170 (27.9) | 173 (41.8) |
| Median [Min, Max] | 166 [120, 259] | 164 [112, 345] | 166 [120, 259] | 164 [112, 345] |
| Missing | 0 (0%) | 0 (0%) | 0 (0%) | 0 (0%) |
| **LDL [mg/dl]** |  |  |  |  |
| Mean (SD) | 97.7 (24.5) | 102 (33.9) | 97.7 (24.5) | 102 (33.9) |
| Median [Min, Max] | 94.0 [35.0, 160] | 97.0 [44.0, 236] | 94.0 [35.0, 160] | 97.0 [44.0, 236] |
| Missing | 0 (0%) | 0 (0%) | 0 (0%) | 0 (0%) |
| **HDL [mg/dl]** |  |  |  |  |
| Mean (SD) | 50.3 (11.0) | 49.9 (12.9) | 50.3 (11.0) | 49.9 (12.9) |
| Median [Min, Max] | 51.0 [25.0, 77.0] | 48.0 [27.0, 79.0] | 51.0 [25.0, 77.0] | 48.0 [27.0, 79.0] |
| Missing | 0 (0%) | 0 (0%) | 0 (0%) | 0 (0%) |
| **Leptin [ng/ml]** |  |  |  |  |
| Mean (SD) | 11.6 (11.7) | 11.8 (11.3) | 11.6 (11.7) | 11.8 (11.3) |
| Median [Min, Max] | 6.10 [0.100, 51.9] | 7.40 [0.100, 40.7] | 6.10 [0.100, 51.9] | 7.40 [0.100, 40.7] |
| Missing | 0 (0%) | 0 (0%) | 0 (0%) | 0 (0%) |
| **Insulin [uU/ml]** |  |  |  |  |
| Mean (SD) | 10.5 (6.64) | 10.5 (7.94) | 10.5 (6.64) | 10.5 (7.94) |
| Median [Min, Max] | 8.80 [3.20, 34.2] | 8.50 [3.20, 54.4] | 8.80 [3.20, 34.2] | 8.50 [3.20, 54.4] |
| Missing | 0 (0%) | 0 (0%) | 0 (0%) | 0 (0%) |
| **Glucose [mmol/ml]** |  |  |  |  |
| Mean (SD) | 4.97 (0.457) | 5.02 (0.444) | 4.97 (0.457) | 5.02 (0.444) |
| Median [Min, Max] | 4.96 [4.05, 6.54] | 4.97 [4.03, 6.67] | 4.96 [4.05, 6.54] | 4.97 [4.03, 6.67] |
| Missing | 0 (0%) | 2 (4.3%) | 0 (0%) | 2 (4.3%) |
| **Ghrelin [pg/ml]** |  |  |  |  |
| Mean (SD) | 147 (107) | 148 (102) | 147 (107) | 148 (102) |
| Median [Min, Max] | 132 [4.15, 505] | 144 [4.83, 393] | 132 [4.15, 505] | 144 [4.83, 393] |
| Missing | 2 (3.6%) | 2 (4.3%) | 2 (3.6%) | 2 (4.3%) |
| **GLP-1 [pg/ml]** |  |  |  |  |
| Mean (SD) | 117 (51.5) | 116 (49.1) | 117 (51.5) | 116 (49.1) |
| Median [Min, Max] | 110 [1.30, 234] | 108 [39.8, 245] | 110 [1.30, 234] | 108 [39.8, 245] |
| Missing | 2 (3.6%) | 2 (4.3%) | 2 (3.6%) | 2 (4.3%) |
| **PYY [pg/ml]** |  |  |  |  |
| Mean (SD) | 56.3 (59.9) | 58.9 (58.3) | 56.3 (59.9) | 58.9 (58.3) |
| Median [Min, Max] | 45.5 [6.80, 235] | 46.5 [6.80, 299] | 45.5 [6.80, 235] | 46.5 [6.80, 299] |
| Missing | 2 (3.6%) | 2 (4.3%) | 2 (3.6%) | 2 (4.3%) |
| **IL-6 [pg/ml]** |  |  |  |  |
| Mean (SD) | 1.56 (2.08) | 1.11 (0.360) | 1.56 (2.08) | 1.11 (0.360) |
| Median [Min, Max] | 1.00 [1.00, 12.0] | 1.00 [1.00, 2.30] | 1.00 [1.00, 12.0] | 1.00 [1.00, 2.30] |
| Missing | 0 (0%) | 0 (0%) | 0 (0%) | 0 (0%) |
| **TNF-alpha [pg/ml]** |  |  |  |  |
| Mean (SD) | 5.94 (1.79) | 6.12 (1.84) | 5.94 (1.79) | 6.12 (1.84) |
| Median [Min, Max] | 6.00 [2.00, 10.2] | 6.10 [2.00, 9.90] | 6.00 [2.00, 10.2] | 6.10 [2.00, 9.90] |
| Missing | 0 (0%) | 0 (0%) | 0 (0%) | 0 (0%) |
| **HCRP [mg/l]** |  |  |  |  |
| Mean (SD) | 3.13 (4.82) | 2.62 (3.92) | 3.13 (4.82) | 2.62 (3.92) |
| Median [Min, Max] | 1.39 [0.150, 27.6] | 1.51 [0.150, 24.3] | 1.39 [0.150, 27.6] | 1.51 [0.150, 24.3] |
| Missing | 1 (1.8%) | 0 (0%) | 1 (1.8%) | 0 (0%) |
| **TMAO [ng/ml]** |  |  |  |  |
| Mean (SD) | 281 (210) | 234 (231) | 281 (210) | 234 (231) |
| Median [Min, Max] | 216 [54.0, 944] | 191 [14.0, 1130] | 216 [54.0, 944] | 191 [14.0, 1130] |
| Missing | 0 (0%) | 1 (2.1%) | 0 (0%) | 1 (2.1%) |
| **Tryptophan [umol/l]** |  |  |  |  |
| Mean (SD) | 33.1 (7.66) | 33.9 (8.48) | 33.1 (7.66) | 33.9 (8.48) |
| Median [Min, Max] | 31.7 [20.2, 58.4] | 32.5 [20.9, 58.7] | 31.7 [20.2, 58.4] | 32.5 [20.9, 58.7] |
| Missing | 2 (3.6%) | 3 (6.4%) | 2 (3.6%) | 3 (6.4%) |
| **Tyrosine [umol/l]** |  |  |  |  |
| Mean (SD) | 52.6 (11.8) | 54.6 (10.4) | 52.6 (11.8) | 54.6 (10.4) |
| Median [Min, Max] | 53.1 [25.5, 79.1] | 55.2 [27.7, 73.3] | 53.1 [25.5, 79.1] | 55.2 [27.7, 73.3] |
| Missing | 0 (0%) | 3 (6.4%) | 0 (0%) | 3 (6.4%) |
| **ALAT [ukat/l]** |  |  |  |  |
| Mean (SD) | 0.381 (0.136) | 0.473 (0.312) | 0.381 (0.136) | 0.473 (0.312) |
| Median [Min, Max] | 0.370 [0.190, 0.790] | 0.395 [0.200, 1.96] | 0.370 [0.190, 0.790] | 0.395 [0.200, 1.96] |
| Missing | 0 (0%) | 1 (2.1%) | 0 (0%) | 1 (2.1%) |
| **ASAT [ukat/l]** |  |  |  |  |
| Mean (SD) | 0.395 (0.0769) | 0.425 (0.132) | 0.395 (0.0769) | 0.425 (0.132) |
| Median [Min, Max] | 0.390 [0.260, 0.640] | 0.400 [0.220, 0.970] | 0.390 [0.260, 0.640] | 0.400 [0.220, 0.970] |
| Missing | 0 (0%) | 1 (2.1%) | 0 (0%) | 1 (2.1%) |
| **TSH [mU/l]** |  |  |  |  |
| Mean (SD) | 62.5 (223) | 131 (290) | 62.5 (223) | 131 (290) |
| Median [Min, Max] | 1.94 [1.01, 959] | 2.13 [1.09, 956] | 1.94 [1.01, 959] | 2.13 [1.09, 956] |
| Missing | 0 (0%) | 1 (2.1%) | 0 (0%) | 1 (2.1%) |
| **Creatinine [umol/l]** |  |  |  |  |
| Mean (SD) | 81.4 (12.3) | 82.2 (13.5) | 81.4 (12.3) | 82.2 (13.5) |
| Median [Min, Max] | 82.0 [56.0, 104] | 82.5 [54.0, 109] | 82.0 [56.0, 104] | 82.5 [54.0, 109] |
| Missing | 0 (0%) | 1 (2.1%) | 0 (0%) | 1 (2.1%) |

Anthropometric markers did not significantly change across measurement timepoints, i.e. BMI, gender-standardized waist-to-hip ratio, fat-free mass and blood pressure (interaction timepoint*intervention, p_all_ > 0.05), except for gender-standardized fat mass (%), which increased significantly after prebiotic intake (interaction timepoint*intervention, b = 0.16, p = 0.01). All models were adjusted for age, gender, and person as random factors. Both intervention and placebo supplements contained the same amounts of calories and participants reported equally high compliance in taking the daily supplements. Blood marker analyses were adjusted for age, gender, individual as random factors, time of day at blood withdrawal and time fasted.

**Results – Linear mixed model results for changes in anthropometric biomarkers**.

**SI-Table 4: Mixed effects linear model results on anthropometric markers for post-prebiotic intervention.**

|  | **n_obs_** | **n_subj_** | **fixed effects** | **estimate** | **SE** | **t-value** | **full-null model comparison p** |
| --- | --- | --- | --- | --- | --- | --- | --- |
| BMI | 204 | 59 | (intercept) | 28.04 | 0.91 | 31.00 |  |
|  |  |  | time (follow-up) | -0.10 | 0.07 | -1.42 |  |
|  |  |  | intervention (prebiotic) | -0.13 | 0.07 | -1.81 |  |
|  |  |  | age | -0.04 | 0.03 | -1.17 |  |
|  |  |  | gender (male) | 0.48 | 0.30 | 1.61 |  |
|  |  |  | time (follow-up) * intervention (prebiotic) | 0.18 | 0.10 | 1.87 | 0.06 |
| Waist-to-hip ratio | 204 | 59 | (intercept) | 0.71 | 0.02 | 28.86 |  |
|  |  |  | time (follow-up) | -0.01 | 0.01 | -1.53 |  |
|  |  |  | intervention (prebiotic) | 0.00 | 0.01 | -0.38 |  |
|  |  |  | age | 0.01 | 0.00 | 3.20 |  |
|  |  |  | gender (male) | 0.06 | 0.01 | 5.15 |  |
|  |  |  | time (follow-up) * intervention (prebiotic) | 0.00 | 0.01 | 0.27 | 0.78 |
| % Fat mass gender-standardized | 203 | 59 | (intercept) | -0.01 | 0.58 | -0.02 |  |
|  |  |  | time (follow-up) | -0.11 | 0.04 | -2.59 |  |
|  |  |  | intervention (prebiotic) | -0.11 | 0.04 | -2.43 |  |
|  |  |  | age | 0.00 | 0.02 | 0.07 |  |
|  |  |  | gender (male) | 0.10 | 0.19 | 0.52 |  |
|  |  |  | **time (follow-up) * intervention (prebiotic)** | **0.16** | **0.06** | **2.57** | **0.01** |
| Fat mass gender-standardized [kg] | 203 | 59 | (intercept) | 0.03 | 0.57 | 0.06 |  |
|  |  |  | time (follow-up) | -0.09 | 0.04 | -2.55 |  |
|  |  |  | intervention (prebiotic) | -0.10 | 0.04 | -2.86 |  |
|  |  |  | age | 0.00 | 0.02 | -0.10 |  |
|  |  |  | gender (male) | 0.13 | 0.17 | 0.78 |  |
|  |  |  | **time (follow-up) * intervention (prebiotic)** | **0.14** | **0.05** | **2.71** | **0.0067** |
| Fat-free mass gender-standardized [kg] | 203 | 59 | (intercept) | 0.39 | 0.56 | 0.71 |  |
|  |  |  | time (follow-up) | 0.04 | 0.04 | 0.99 |  |
|  |  |  | intervention (prebiotic) | 0.02 | 0.04 | 0.50 |  |
|  |  |  | age | -0.02 | 0.02 | -0.96 |  |
|  |  |  | gender (male) | 0.08 | 0.16 | 0.48 |  |
|  |  |  | time (follow-up) * intervention (prebiotic) | -0.04 | 0.05 | -0.79 | 0.43 |
| 10% of daily energy requirement | 203 | 59 | (intercept) | 181.55 | 10.22 | 17.76 |  |
|  |  |  | time (follow-up) | -1.89 | 0.95 | -1.98 |  |
|  |  |  | intervention (prebiotic) | -1.47 | 0.95 | -1.55 |  |
|  |  |  | age | -0.30 | 0.36 | -0.85 |  |
|  |  |  | gender (male) | 26.92 | 3.74 | 7.19 |  |
|  |  |  | time (follow-up) * intervention (prebiotic) | 2.19 | 1.36 | 1.61 | 0.10 |

*Formula: variable_of_interest ~ intervention * timepoint + (1 | subject) + age + gender. REML criterion at convergence > 158. Significance, *** p < 0.001.*

**SI-Table 5: Mixed effects linear model results on serum markers for post-prebiotic intervention.**

|  | n_obs_ | n_subj_ | **fixed effects** | **estimate** | **SE** | **t-value** | **full-null model comparison p** |
| --- | --- | --- | --- | --- | --- | --- | --- |
| hsCRP | 195 | 58 | (intercept) | 4.74 | 2.41 | 1.97 |  |
|  |  |  | time (follow-up) | 0.20 | 0.67 | 0.30 |  |
|  |  |  | intervention (prebiotic) | 0.46 | 0.66 | 0.69 |  |
|  |  |  | age | -0.08 | 0.06 | -1.21 |  |
|  |  |  | gender (male) | -3.62 | 0.87 | -4.17 |  |
|  |  |  | time of day (8:00 AM) | -1.72 | 1.66 | -1.04 |  |
|  |  |  | time of day (9:15 AM) | -0.22 | 0.94 | -0.23 |  |
|  |  |  | time of day (10:30 AM) | -1.95 | 1.79 | -1.09 |  |
|  |  |  | time of day (11:15 AM) | 0.37 | 1.19 | 0.31 |  |
|  |  |  | time fasted (hours) | 0.24 | 0.14 | 1.69 |  |
|  |  |  | time (follow-up) * intervention (prebiotic) | -0.58 | 0.94 | -0.62 | 0.56 |
| IL-6 | 196 | 58 | (intercept) | 1.16 | 0.86 | 1.33 |  |
|  |  |  | time (follow-up) | 0.19 | 0.31 | 0.60 |  |
|  |  |  | intervention (prebiotic) | 0.26 | 0.30 | 0.85 |  |
|  |  |  | age | 0.001 | 0.02 | 0.04 |  |
|  |  |  | gender (male) | 0.22 | 0.28 | 0.78 |  |
|  |  |  | time of day (8:00 AM) | -0.86 | 0.53 | -1.62 |  |
|  |  |  | time of day (9:15 AM) | -0.53 | 0.30 | -1.78 |  |
|  |  |  | time of day (10:30 AM) | -0.50 | 0.59 | -0.85 |  |
|  |  |  | time of day (11:15 AM) | -0.75 | 0.39 | -1.94 |  |
|  |  |  | time fasted (hours) | 0.03 | 0.06 | 0.58 |  |
|  |  |  | time (follow-up) * intervention (prebiotic) | -0.68 | 0.43 | -1.58 | 0.11 |
| TNF | 196 | 58 | (intercept) | 3.57 | 1.12 | 3.19 |  |
|  |  |  | time (follow-up) | -0.007 | 0.25 | -0.003 |  |
|  |  |  | intervention (prebiotic) | 0.02 | 0.24 | 0.08 |  |
|  |  |  | age | 0.04 | 0.03 | 1.29 |  |
|  |  |  | gender (male) | 0.58 | 0.43 | 1.33 |  |
|  |  |  | time of day (8:00 AM) | 0.86 | 0.85 | 1.01 |  |
|  |  |  | time of day (9:15 AM) | -0.05 | 0.48 | -0.11 |  |
|  |  |  | time of day (10:30 AM) | 0.04 | 0.89 | 0.04 |  |
|  |  |  | time of day (11:15 AM) | 0.20 | 0.60 | 0.33 |  |
|  |  |  | time fasted (hours) | 0.05 | 0.06 | 0.96 |  |
|  |  |  | time (follow-up) * intervention (prebiotic) | 0.18 | 0.35 | 0.52 | 0.60 |
| HDL | 197 | 58 | (intercept) | 63.65 | 6.41 | 9.93 |  |
|  |  |  | time (follow-up) | -0.18 | 0.93 | -0.19 |  |
|  |  |  | intervention (prebiotic) | 0.90 | 0.92 | 0.98 |  |
|  |  |  | age | -0.30 | 0.21 | -1.45 |  |
|  |  |  | gender (male) | -10.05 | 2.59 | -3.88 |  |
|  |  |  | time of day (8:00 AM) | 1.92 | 5.50 | 0.35 |  |
|  |  |  | time of day (9:15 AM) | 1.41 | 3.12 | 0.45 |  |
|  |  |  | time of day (10:30 AM) | -1.89 | 5.62 | -0.34 |  |
|  |  |  | time of day (11:15 AM) | 2.53 | 3.63 | 0.70 |  |
|  |  |  | time fasted (hours) | 0.02 | 0.23 | 0.08 |  |
|  |  |  | time (follow-up) * intervention (prebiotic) | -0.34 | 1.30 | -0.26 | 0.79 |
| LDL | 197 | 58 | (intercept) | 89.03 | 18.13 | 4.91 |  |
|  |  |  | time (follow-up) | -5.02 | 2.46 | -2.04 |  |
|  |  |  | intervention (prebiotic) | -1.83 | 2.43 | -0.75 |  |
|  |  |  | age | 0.22 | 0.60 | 0.36 |  |
|  |  |  | gender (male) | 0.69 | 7.27 | 0.10 |  |
|  |  |  | time of day (8:00 AM) | 0.11 | 15.72 | 0.01 |  |
|  |  |  | time of day (9:15 AM) | 9.55 | 8.90 | 1.07 |  |
|  |  |  | time of day (10:30 AM) | 11.86 | 16.02 | 0.74 |  |
|  |  |  | time of day (11:15 AM) | 13.25 | 10.28 | 1.29 |  |
|  |  |  | time fasted (hours) | -0.22 | 0.60 | -0.38 |  |
|  |  |  | **time (follow-up) * intervention (prebiotic)** | **10.41**** | **3.44** | **3.03** | **0.003** |
| LDL/  HDL | 197 | 58 | (intercept) | 1.09 | 0.50 | 2.20 |  |
|  |  |  | time (follow-up) | -0.09 | 0.05 | -1.65 |  |
|  |  |  | intervention (prebiotic) | -0.08 | 0.05 | -1.46 |  |
|  |  |  | age | 0.03 | 0.02 | 2.04 |  |
|  |  |  | gender (male) | 0.27 | 0.19 | 1.41 |  |
|  |  |  | time of day (8:00 AM) | -0.27 | 0.44 | -0.61 |  |
|  |  |  | time of day (9:15 AM) | -0.002 | 0.25 | -0.01 |  |
|  |  |  | time of day (10:30 AM) | 0.17 | 0.45 | 0.37 |  |
|  |  |  | time of day (11:15 AM) | 0.08 | 0.28 | 0.29 |  |
|  |  |  | time fasted (hours) | -0.01 | 0.01 | -0.67 |  |
|  |  |  | **time (follow-up) * intervention (prebiotic)** | **0.25***** | **0.07** | **3.40** | **0.0007** |
| Triglycerides | 197 | 58 | (intercept) | 69.54 | 32.95 | 2.11 |  |
|  |  |  | time (follow-up) | -9.34 | 7.07 | -1.32 |  |
|  |  |  | intervention (prebiotic) | -4.04 | 6.94 | -0.58 |  |
|  |  |  | age | 1.50 | 0.97 | 1.55 |  |
|  |  |  | gender (male) | -8.67 | 12.92 | -0.67 |  |
|  |  |  | time of day (8:00 AM) | -39.10 | 25.29 | -1.55 |  |
|  |  |  | time of day (9:15 AM) | -26.27 | 14.47 | -1.82 |  |
|  |  |  | time of day (10:30 AM) | -19.08 | 26.57 | -0.72 |  |
|  |  |  | time of day (11:15 AM) | -37.28 | 17.75 | -2.10 |  |
|  |  |  | time fasted (hours) | 1.86 | 1.64 | 1.14 |  |
|  |  |  | time (follow-up) * intervention (prebiotic) | 4.12 | 9.90 | 0.42 | 0.67 |
| Cholesterol | 197 | 58 | (intercept) | 163.46 | 21.96 | 7.45 |  |
|  |  |  | time (follow-up) | -6.80 | 2.98 | -2.28 |  |
|  |  |  | intervention (prebiotic) | -1.00 | 2.95 | -0.34 |  |
|  |  |  | age | 0.34 | 0.72 | 0.47 |  |
|  |  |  | gender (male) | -10.15 | 8.80 | -1.15 |  |
|  |  |  | time of day (8:00 AM) | -4.21 | 19.03 | -0.22 |  |
|  |  |  | time of day (9:15 AM) | 6.61 | 10.77 | 0.61 |  |
|  |  |  | time of day (10:30 AM) | 7.46 | 19.40 | 0.39 |  |
|  |  |  | time of day (11:15 AM) | 6.64 | 12.44 | 0.53 |  |
|  |  |  | time fasted (hours) | 0.08 | 0.73 | 0.11 |  |
|  |  |  | **time (follow-up) * intervention (prebiotic)** | **10.90**** | **4.18** | **2.61** | **0.0086** |
| Cholesterol  (without sub-47) | 196 | 58 | (intercept) | 142.22 | 19.41 | 7.33 |  |
|  |  |  | time (follow-up) | -6.80 | 2.79 | -2.44 |  |
|  |  |  | intervention (prebiotic) | -0.34 | 2.76 | -0.12 |  |
|  |  |  | age | 0.84 | 0.63 | 1.33 |  |
|  |  |  | gender (male) | -4.32 | 7.80 | -0.55 |  |
|  |  |  | time of day (8:00 AM) | -7.71 | 16.30 | -0.47 |  |
|  |  |  | time of day (9:15 AM) | -0.93 | 9.37 | -0.10 |  |
|  |  |  | time of day (10:30 AM) | 5.27 | 16.66 | 0.32 |  |
|  |  |  | time of day (11:15 AM) | 1.55 | 10.82 | 0.14 |  |
|  |  |  | time fasted (hours) | 0.50 | 0.68 | 0.73 |  |
|  |  |  | **time (follow-up) * intervention (prebiotic)** | **9.07*** | **3.93** | **2.31** | **0.020** |
| Insulin | 197 | 58 | (intercept) | 14.64 | 3.96 | 3.70 |  |
|  |  |  | time (follow-up) | -0.14 | 0.81 | -0.17 |  |
|  |  |  | intervention (prebiotic) | 0.86 | 0.80 | 1.08 |  |
|  |  |  | age | -0.18 | 0.12 | -1.53 |  |
|  |  |  | gender (male) | 0.09 | 1.57 | 0.06 |  |
|  |  |  | time of day (8:00 AM) | -1.62 | 3.10 | -0.52 |  |
|  |  |  | time of day (9:15 AM) | -0.68 | 1.77 | -0.38 |  |
|  |  |  | time of day (10:30 AM) | -0.42 | 3.24 | -0.13 |  |
|  |  |  | time of day (11:15 AM) | 0.55 | 2.16 | 0.25 |  |
|  |  |  | time fasted (hours) | 0.04 | 0.19 | 0.23 |  |
|  |  |  | time (follow-up) * intervention (prebiotic) | -0.04 | 1.13 | -0.04 | 0.98 |
| Ghrelin | 193 | 58 | (intercept) | 221.45 | 60.23 | 3.68 |  |
|  |  |  | time (follow-up) | 16.27 | 14.31 | 1.14 |  |
|  |  |  | intervention (prebiotic) | -6.90 | 14.18 | -0.49 |  |
|  |  |  | age | 0.37 | 1.71 | 0.21 |  |
|  |  |  | gender (male) | -72.75 | 22.89 | -3.18 |  |
|  |  |  | time of day (8:00 AM) | 95.04 | 44.19 | 2.15 |  |
|  |  |  | time of day (9:15 AM) | 32.14 | 25.34 | 1.27 |  |
|  |  |  | time of day (10:30 AM) | -5.45 | 47.08 | -0.12 |  |
|  |  |  | time of day (11:15 AM) | 22.52 | 31.45 | 0.72 |  |
|  |  |  | time fasted (hours) | -4.19 | 3.23 | -1.30 |  |
|  |  |  | time (follow-up) * intervention (prebiotic) | -11.25 | 20.30 | -0.55 | 0.58 |
| GLP-1 | 194 | 58 | (intercept) | 144.47 | 30.09 | 4.80 |  |
|  |  |  | time (follow-up) | -1.06 | 6.44 | -0.16 |  |
|  |  |  | intervention (prebiotic) | 1.95 | 6.40 | 0.31 |  |
|  |  |  | age | -0.81 | 0.89 | -0.91 |  |
|  |  |  | gender (male) | 24.00 | 11.73 | 2.05 |  |
|  |  |  | time of day (8:00 AM) | -61.57 | 23.06 | -2.67 |  |
|  |  |  | time of day (9:15 AM) | -33.10 | 13.20 | -2.51 |  |
|  |  |  | time of day (10:30 AM) | -31.28 | 24.26 | -1.29 |  |
|  |  |  | time of day (11:15 AM) | -22.89 | 16.18 | -1.42 |  |
|  |  |  | time fasted (hours) | 0.10 | 1.50 | 0.07 |  |
|  |  |  | time (follow-up) * intervention (prebiotic) | -2.39 | 9.18 | -0.26 | 0.79 |
| PYY | 194 | 58 | (intercept) | 37.37 | 33.27 | 1.12 |  |
|  |  |  | time (follow-up) | -3.20 | 7.91 | -0.40 |  |
|  |  |  | intervention (prebiotic) | 2.46 | 7.83 | 0.32 |  |
|  |  |  | age | 0.85 | 0.94 | 0.90 |  |
|  |  |  | gender (male) | 2.39 | 12.62 | 0.19 |  |
|  |  |  | time of day (8:00 AM) | -34.81 | 24.37 | -1.43 |  |
|  |  |  | time of day (9:15 AM) | -11.05 | 13.96 | -0.79 |  |
|  |  |  | time of day (10:30 AM) | -1.79 | 25.98 | -0.07 |  |
|  |  |  | time of day (11:15 AM) | -24.23 | 17.35 | -1.40 |  |
|  |  |  | time fasted (hours) | -0.08 | 1.709 | 0.05 |  |
|  |  |  | time (follow-up) * intervention (prebiotic) | 6.21 | 11.26 | 0.55 | 0.58 |
| Glucose | 195 | 58 | (intercept) | 4.55 | 0.25 | 18.01 |  |
|  |  |  | time (follow-up) | -0.04 | 0.05 | -0.74 |  |
|  |  |  | intervention (prebiotic) | 0.04 | 0.05 | 0.76 |  |
|  |  |  | age | 0.01 | 0.01 | 1.18 |  |
|  |  |  | gender (male) | 0.22 | 0.11 | 2.05 |  |
|  |  |  | time of day (8:00 AM) | 0.01 | 0.20 | 0.03 |  |
|  |  |  | time of day (9:15 AM) | -0.14 | 0.11 | -1.26 |  |
|  |  |  | time of day (10:30 AM) | -0.01 | 0.21 | -0.03 |  |
|  |  |  | time of day (11:15 AM) | -0.13 | 0.14 | -0.93 |  |
|  |  |  | time fasted (hours) | 0.00 | 0.01 | 0.38 |  |
|  |  |  | time (follow-up) * intervention (prebiotic) | 0.05 | 0.07 | 0.72 | 0.46 |
| Leptin | 197 | 58 | (intercept) | 35.91 | 4.94 | 7.27 |  |
|  |  |  | time (follow-up) | -0.16 | 0.80 | -0.20 |  |
|  |  |  | intervention (prebiotic) | -0.48 | 0.79 | -0.61 |  |
|  |  |  | age | -0.18 | 0.16 | -1.15 |  |
|  |  |  | gender (male) | -18.74 | 2.19 | -8.55 |  |
|  |  |  | time of day (8:00 AM) | -5.16 | 4.18 | -1.24 |  |
|  |  |  | time of day (9:15 AM) | -0.42 | 2.38 | -0.18 |  |
|  |  |  | time of day (10:30 AM) | -7.51 | 4.29 | -1.75 |  |
|  |  |  | time of day (11:15 AM) | -1.60 | 2.81 | -0.57 |  |
|  |  |  | time fasted (hours) | -0.34 | 0.19 | -1.78 |  |
|  |  |  | time (follow-up) * intervention (prebiotic) | 0.96 | 1.12 | 0.86 | 0.75 |
| Betain | 195 | 58 | (intercept) | 1.69 | 0.68 | 2.47 |  |
|  |  |  | time (follow-up) | -0.03 | 0.12 | -0.20 |  |
|  |  |  | intervention (prebiotic) | 0.17 | 0.12 | 1.36 |  |
|  |  |  | age | 0.03 | 0.02 | 1.57 |  |
|  |  |  | gender (male) | 1.76 | 0.30 | 5.95 |  |
|  |  |  | time of day (8:00 AM) | 0.27 | 0.56 | 0.47 |  |
|  |  |  | time of day (9:15 AM) | -0.14 | 0.32 | -0.44 |  |
|  |  |  | time of day (10:30 AM) | -1.07 | 0.58 | -1.84 |  |
|  |  |  | time of day (11:15 AM) | -0.03 | 0.39 | -0.08 |  |
|  |  |  | time fasted (hours) | -0.03 | 0.03 | -1.16 |  |
|  |  |  | time (follow-up) * intervention (prebiotic) | -0.05 | 0.17 | -0.31 | 0.75 |
| Carnitin | 195 | 58 | (intercept) | 3.68 | 0.86 | 4.26 |  |
|  |  |  | time (follow-up) | 0.14 | 0.15 | 0.94 |  |
|  |  |  | intervention (prebiotic) | 0.05 | 0.15 | 0.31 |  |
|  |  |  | age | 0.06 | 0.03 | 2.02 |  |
|  |  |  | gender (male) | 2.30 | 0.38 | 6.09 |  |
|  |  |  | time of day (8:00 AM) | -0.71 | 0.72 | -0.99 |  |
|  |  |  | time of day (9:15 AM) | -0.22 | 0.41 | -0.53 |  |
|  |  |  | time of day (10:30 AM) | -0.67 | 0.74 | -0.90 |  |
|  |  |  | time of day (11:15 AM) | -0.82 | 0.49 | -1.68 |  |
|  |  |  | time fasted (hours) | -0.01 | 0.04 | -0.42 |  |
|  |  |  | time (follow-up) * intervention (prebiotic) | 0.01 | 0.21 | 0.07 | 0.95 |
| Cholin | 195 | 58 | (intercept) | 0.77 | 0.12 | 6.40 |  |
|  |  |  | time (follow-up) | -0.02 | 0.03 | -0.63 |  |
|  |  |  | intervention (prebiotic) | 0.02 | 0.03 | 0.83 |  |
|  |  |  | age | 0.005 | 0.003 | 1.42 |  |
|  |  |  | gender (male) | 0.09 | 0.05 | 1.82 |  |
|  |  |  | time of day (8:00 AM) | -0.04 | 0.09 | -0.47 |  |
|  |  |  | time of day (9:15 AM) | -0.05 | 0.05 | -1.07 |  |
|  |  |  | time of day (10:30 AM) | -0.17 | 0.09 | -1.80 |  |
|  |  |  | time of day (11:15 AM) | 0.01 | 0.06 | 0.18 |  |
|  |  |  | time fasted (hours) | -0.00 | 0.01 | -0.36 |  |
|  |  |  | time (follow-up) * intervention (prebiotic) | 0.02 | 0.04 | 0.45 | 0.65 |
| TMAO | 194 | 58 | (intercept) | 38.47 | 106.83 | 0.36 |  |
|  |  |  | time (follow-up) | 19.19 | 34.26 | 0.56 |  |
|  |  |  | intervention (prebiotic) | 84.00 | 33.14 | 2.54 |  |
|  |  |  | age | 0.58 | 2.71 | 0.21 |  |
|  |  |  | gender (male) | 3.64 | 36.98 | 0.10 |  |
|  |  |  | time of day (8:00 AM) | 57.71 | 68.17 | 0.85 |  |
|  |  |  | time of day (9:15 AM) | -16.56 | 39.29 | -0.42 |  |
|  |  |  | time of day (10:30 AM) | -184.62 | 76.05 | -2.43 |  |
|  |  |  | time of day (11:15 AM) | 17.42 | 50.63 | 0.34 |  |
|  |  |  | time fasted (hours) | 12.17 | 6.83 | 1.78 |  |
|  |  |  | time (follow-up) * intervention (prebiotic) | -68.42 | 47.81 | -1.43 | 0.16 |
| Tryptophan | 188 | 57 | (intercept) | 30.99 | 4.62 | 6.70 |  |
|  |  |  | time (follow-up) | -0.67 | 1.08 | -0.62 |  |
|  |  |  | intervention (prebiotic) | -1.06 | 1.07 | -0.99 |  |
|  |  |  | age | 0.10 | 0.14 | 0.73 |  |
|  |  |  | gender (male) | 2.84 | 1.86 | 1.53 |  |
|  |  |  | time of day (8:00 AM) | 8.25 | 3.47 | 2.38 |  |
|  |  |  | time of day (9:15 AM) | 0.17 | 1.99 | 0.09 |  |
|  |  |  | time of day (10:30 AM) | 9.46 | 3.65 | 2.59 |  |
|  |  |  | time of day (11:15 AM) | -0.56 | 2.52 | -0.22 |  |
|  |  |  | time fasted (hours) | -0.23 | 0.25 | -0.90 |  |
|  |  |  | time (follow-up) * intervention (prebiotic) | 1.51 | 1.52 | 1.00 | 0.30 |
| Tryptophan/LNAA | 187 | 57 | (intercept) | 0.1140 | 0.0179 | 6.347 |  |
|  |  |  | time (follow-up) | 0.0036 | 0.0034 | 1.058 |  |
|  |  |  | intervention (prebiotic) | 0.0027 | 0.0034 | 0.812 |  |
|  |  |  | age | 0.0002 | 0.0006 | 0.391 |  |
|  |  |  | gender (male) | -0.0123 | 0.0077 | -1.597 |  |
|  |  |  | time of day (8:00 AM) | 0.0415 | 0.0145 | 2.867 |  |
|  |  |  | time of day (9:15 AM) | 0.0078 | 0.0083 | 0.937 |  |
|  |  |  | time of day (10:30 AM) | 0.0598 | 0.0150 | 3.984 |  |
|  |  |  | time of day (11:15 AM) | 0.0106 | 0.0102 | 1.042 |  |
|  |  |  | time fasted (hours) | -0.0001 | 0.0008 | -0.093 |  |
|  |  |  | time (follow-up) * intervention (prebiotic) | -0.0048 | 0.0048 | -1.011 | 0.31 |
| Tyrosine | 193 | 58 | (intercept) | 59.76 | 6.10 | 9.80 |  |
|  |  |  | time (follow-up) | -0.56 | 1.59 | -0.35 |  |
|  |  |  | intervention (prebiotic) | -1.91 | 1.56 | -1.23 |  |
|  |  |  | age | 0.28 | 0.17 | 1.64 |  |
|  |  |  | gender (male) | 10.30 | 2.31 | 4.45 |  |
|  |  |  | time of day (8:00 AM) | -0.21 | 4.36 | -0.05 |  |
|  |  |  | time of day (9:15 AM) | -3.47 | 2.49 | -1.39 |  |
|  |  |  | time of day (10:30 AM) | -7.07 | 4.65 | -1.52 |  |
|  |  |  | time of day (11:15 AM) | -5.71 | 3.12 | -1.83 |  |
|  |  |  | time fasted (hours) | -1.37 | 0.35 | -3.94 |  |
|  |  |  | time (follow-up) * intervention (prebiotic) | 1.73 | 2.24 | 0.77 | 0.43 |
| Tyrosine/LNAA | 187 | 57 | (intercept) | 0.235 | 0.023 | 10.395 |  |
|  |  |  | time (follow-up) | 0.003 | 0.005 | 0.734 |  |
|  |  |  | intervention (prebiotic) | -0.001 | 0.005 | -0.291 |  |
|  |  |  | age | 0.001 | 0.001 | 1.954 |  |
|  |  |  | gender (male) | -0.002 | 0.009 | -0.177 |  |
|  |  |  | time of day (8:00 AM) | 0.022 | 0.018 | 1.261 |  |
|  |  |  | time of day (9:15 AM) | -0.002 | 0.010 | -0.170 |  |
|  |  |  | time of day (10:30 AM) | -0.011 | 0.018 | -0.575 |  |
|  |  |  | time of day (11:15 AM) | -0.009 | 0.013 | -0.686 |  |
|  |  |  | time fasted (hours) | -0.004 | 0.001 | -4.037 |  |
|  |  |  | time (follow-up) * intervention (prebiotic) | -0.004 | 0.007 | -0.662 | 0.51 |
| ASAT | 196 | 58 | (intercept) | 0.45 | 0.09 | 5.04 |  |
|  |  |  | time (follow-up) | 0.06 | 0.03 | 1.87 |  |
|  |  |  | intervention (prebiotic) | -0.00 | 0.03 | -0.07 |  |
|  |  |  | age | -0.00 | 0.00 | -1.13 |  |
|  |  |  | gender (male) | 0.10 | 0.03 | 3.25 |  |
|  |  |  | time of day (8:00 AM) | 0.02 | 0.06 | 0.31 |  |
|  |  |  | time of day (9:15 AM) | -0.03 | 0.03 | -1.01 |  |
|  |  |  | time of day (10:30 AM) | 0.05 | 0.06 | 0.74 |  |
|  |  |  | time of day (11:15 AM) | -0.01 | 0.04 | -0.22 |  |
|  |  |  | time fasted (hours) | -0.00 | 0.01 | -0.52 |  |
|  |  |  | time (follow-up) * intervention (prebiotic) | -0.03 | 0.04 | -0.79 | 0.42 |
| ALAT | 196 | 58 | (intercept) | 0.27 | 0.12 | 2.29 |  |
|  |  |  | time (follow-up) | -0.0005 | 0.03 | -0.02 |  |
|  |  |  | intervention (prebiotic) | -0.02 | 0.03 | -0.73 |  |
|  |  |  | age | 0.0001 | 0.003 | 0.03 |  |
|  |  |  | gender (male) | 0.23 | 0.04 | 5.20 |  |
|  |  |  | time of day (8:00 AM) | -0.04 | 0.08 | -0.52 |  |
|  |  |  | time of day (9:15 AM) | -0.08 | 0.05 | -1.63 |  |
|  |  |  | time of day (10:30 AM) | 0.05 | 0.09 | 0.57 |  |
|  |  |  | time of day (11:15 AM) | -0.08 | 0.06 | -1.41 |  |
|  |  |  | time fasted (hours) | 0.002 | 0.01 | 0.23 |  |
|  |  |  | time (follow-up) * intervention (prebiotic) | 0.08 | 0.04 | 1.94 | 0.05 |

*Formula: marker_of_interest ~ intervention * timepoint + (1 | subject) + age + gender + time_of_day + time_fasted. REML criterion at convergence > 700. Significance, *** p < 0.001.*
