## Supplementary material for "A prebiotic diet changes neural correlates of food decision-making in overweight adults: a randomized controlled within-subject cross-over trial": SI_fMRI

**MRI assessments**

**MRI acquisition.** Anatomical MRI was acquired using a T1-weighted MPRAGE sequence using the ADNI protocol with the following parameters: TR = 2300ms; TE = 2.98ms; flip angle = 9°; FOV: (256 mm)²; voxel size: (1.0mm)³; 176 slices. Task-based fMRI was acquired using T2*-weighted images: EPI BOLD: repetition time TR = 2000ms, echo time TE = 23.60ms, flip angle FA = 80°C, field-of-view FOV = (204 mm)^2^; voxel size 2 x 2 x 2 mm^3^; 60 slices; gap 0.26mm; orientation T>C -15°; multi-band = 3, interleaved. Field maps and ap/pa were acquired to be used for correcting scanner inhomogeneities in the preprocessing pipeline.

**Stimuli and sessions**. Stimuli for the fMRI task (wanting) and subsequent behavioral tasks (liking) were taken from validated databases, including food and art databases [1, 2, 3]. In total, for the wanting task 640 stimuli (320 food, 320 art) were chosen and split to 4 sets, in a randomized order over four sessions within each participant, matched by calorie content quartiles. Also, the four parallel versions of image sets matched in number of animals, plants, object stimuli and in low-level image characteristics (red, blue, green, object size, contrast: pall > 0.05). All stimuli were also included in the liking task. Stimuli size was normed to 600x450 pixels and presented on white background in Presentation (R) version 16.2 0.13.17 (Windows XP) using a mirror mounted display. Stimuli order was limited to maximally three stimuli of one group (food or art) in a row, and order was randomized per subject across all sessions.

Stimuli characteristics were derived from the databases including low-level characteristics, nutrient content (kcal / 100g for food only) and high-level characteristics based on normative ratings (i.e. population craving). We extended nutritional information and added (grams of fiber / 100g) based on the mean of four independent raters (inter-rater reliability ICC 0.76) and item type (1=Dairy & Eggs, 2=Fruits, 3=Vegetables, 4=Confectionery & Sweets, 5=Bakery Wares & Cereals, 6=Meat, 7=Fish, 8=Beverages, 9=Ready-to-eat Savories, 10=Prepared) (for details see [4]).

One session consisted of one block (with two 30s breaks, about 31 min), each with 160 stimuli, i.e. 80 food and 80 art images. The original German rating questions were: “Wie sehr möchten Sie dies jetzt haben?”. Ratings were acquired using a two-button box with the index and middle finger to move a cursor with a trial-by-trial random position. Orientation of the rating scale was randomized across subjects to left-to-right (1-8) or right-to-left (8-to-1). The original German label for 1 was “überhaupt nicht” and for 8 “unbedingt”. The reward item was randomly chosen for all equally high rated stimuli across one domain for maximal ratings by the study staff.

**Task instructions** were given verbally outside the scanner, followed by a test task inside the scanner (only on the first testing day), where participants were able to ask comprehension questions, followed by written instructions right before the task. [Detailed instructions in German: "Gleich startet die Bewertungsaufgabe. Zur Erinnerung: Eines der am höchsten bewerteten Lebensmittel bekommen Sie zum DIREKTEN Verzehr. Einen der am höchsten bewerteten Kunstdrucke bekommen Sie als Ausdruck zum Mitnehmen. Beachten Sie: 1. Ihre Antwort wird nur gezählt, wenn Sie den grünen Balken MINDESTENS EINMAL bewegen. 2. Merken Sie sich die Bilder so gut wie möglich. Im Anschluss fragen wir ab, ob Sie sich an diese erinnern können."]

**MRI Quality Controls**

Visual examination of the fMRIprep generated reports was done, checking for the correct extraction of the brainmask, correct surface reconstruction, for unusual artefacts and correct coregistration between fieldmap to EPI and EPI to T1 images. Smoothing with FWHM 6mm was done in SPM12. Slice Timing was not used. MRIQC was used for the individual and group level for both EPI BOLD and T1w sequences [5].

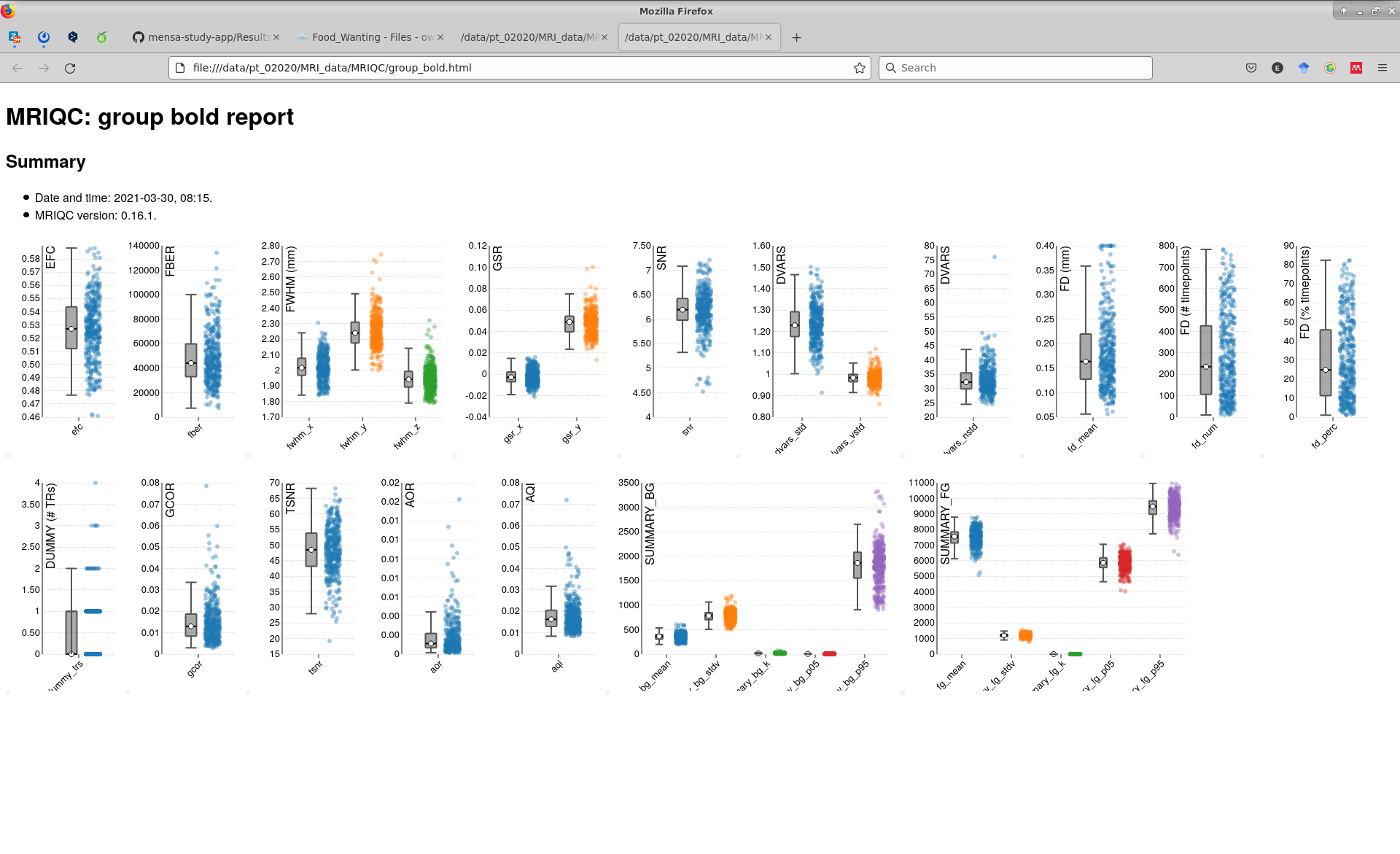

SI Figure 1: MRIQC report for BOLD sequences on the group level.

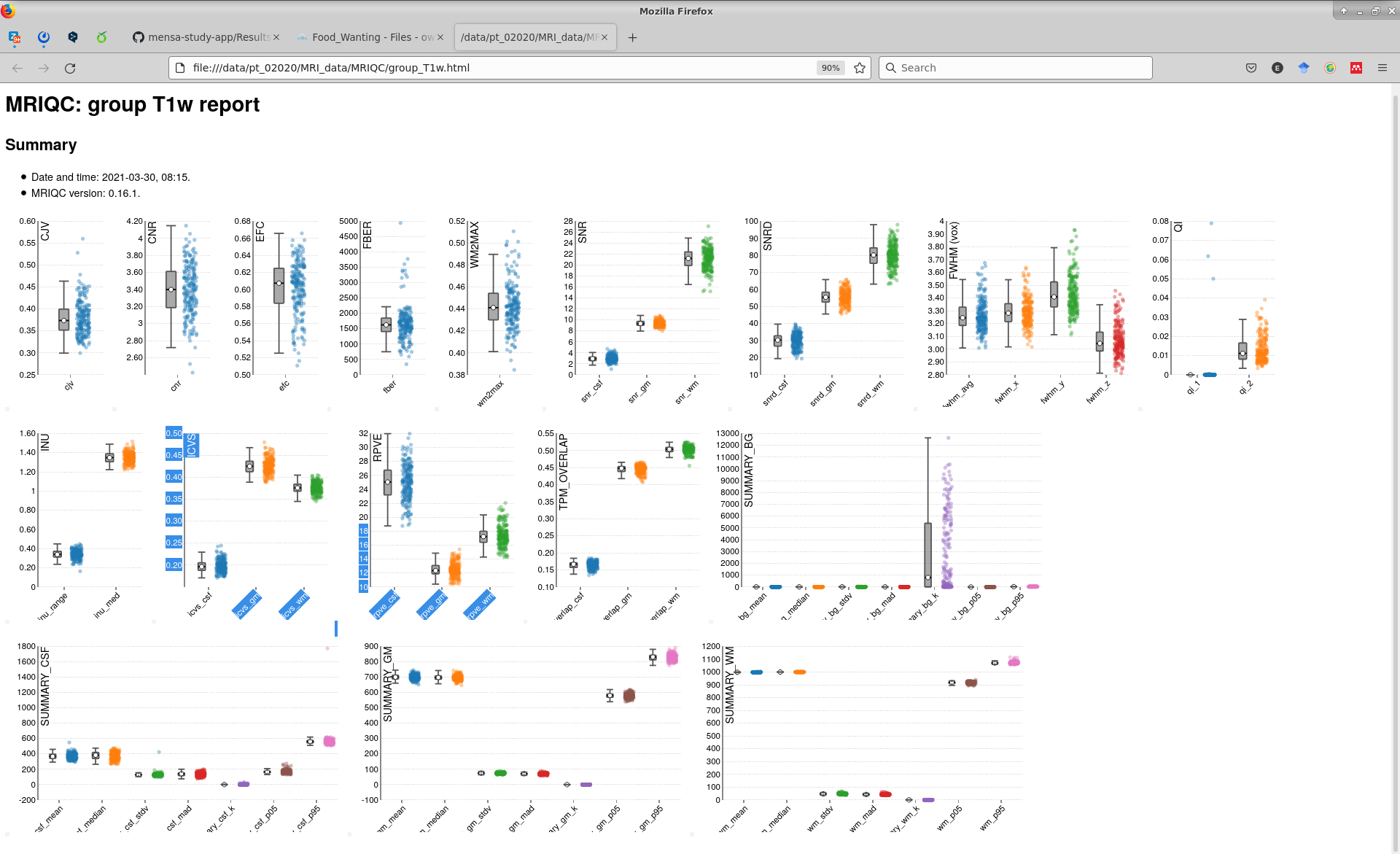

SI Figure 2: MRIQC report for T1w on the group level.

**First-level fMRI analysis specifications**

**Parameter settings.** SPM default parameters were used, and implicit masking was set to 0.4 to include all within-skull voxels. From picture onset, picture duration was modelled to 4000ms.

**Dataset exclusion.** Pre-defined exclusion criteria were employed: a) manual exclusion in case of severe brain pathology (based on T1w/FLAIR image) leading to errors in coregistration by visual inspection of study doctors (0 cases), b) fMRI data was excluded if there was an erroneous co-registration of EPI to T1w (quality assessment done on fmriprep outputs) (0 cases). In addition, timepoints in which participants showed no interest in the stimuli at all (if all stimuli are ranked 1 out 8 for either food or art) had to be excluded from the SPM analysis as the parametric modulator is collinear to the onset regressor (1 participant at 2 sessions).

**fMRI trial exclusion.** Missed trials during wanting fMRI (i.e. missed response) were handled in two ways according to the randomness of those. Trials missing at random vs. non-random trials: if missed trials occurred in blocks, those were considered non-random and likely due to inattentiveness or sleep of the participants and therefore excluded from the analysis (occurred in 10 subject-session cases), in contrast to missed trials scattered in time, which might have been randomly missed and therefore, worthwhile to impute the concurrent wanting rating to allow inferences about BOLD activity whilst picture evaluation (occurred in 24 subject-session cases). Visual inspection of logfiles were performed by one rater and if the decision is unclear, then group consensus decisions was made (not necessary). Imputation of single-item ratings was done only if missed items were up to 10% of all 160 stimuli of one session (up to 16 missed stimuli) and replaced with the individual average of the respective stimuli type, food or art, respectively, of that session. Missed trials during the behavioural liking task were treated likewise: if >=10% items (72 out 720 stimuli) were missed or not available due to a missing follow-up appointment, then the respective session of that participant was excluded from analysis for model C1/C2.

**Motion scrubbing. I**n all contrasts, the six rigid motion parameters derived by fmriprep, as well as a binary regressor for TRs exceeding a motion threshold of 0.9mm were included (derived by fsl_motion_outliers), as recommended by [6]

**First-level contrasts** were done in SPM12 with four predictors (placebo_BL, placebo_FU, verum_BL, verum_FU) and three design matrices. Design matrix A: food > art [1 0 -1 0], art > food [-1 0 1 0], food > art wanting slope [0 1 0 -1], art > food wanting slope [0 -1 0 1] and wanting modulation [0 0.5 0 0.5]; Design Matrix B1: kcal*wanting [0 0 1 0 0]; Design Matrix B2: fiber*wanting [0 0 1 0 0]; Design Matrix C1 for liking as parametric modulator of no interest: food > art [1 0 -1 0 0], art > food [-1 0 1 0 0], food > art wanting slope [0 1 0 -1 0], art > food wanting slope [0 -1 0 1 0] and wanting modulation [0 0.5 0 0.5 0]; Design Matrix C2 for liking as parametric modulator of interest: food > art liking slope [0 1 0 -1], art > food liking slope [0 -1 0 1] and liking modulation [0 0.5 0 0.5 0]. Orthogonalization in SPM was set to 1 is case of one parametric modulator per condition (model A and C2) and in case of more than one parametric modulator, orthogonalization was set to 0 (models B1, B2 and C1).

**Brain mask:** A region-of-interest (ROI) mask were created using 3D-volumes provided by pprox.th.org, from a meta-analysis based on 922 studies for “reward” as search term in combination with a meta-analysis based on 98 studies for “hypothalamus” (accessed on 19 April 2021). The reward-mask was thresholded at zmin= 1.96 (corresponding to alpha<0.05, uncorr.) and combined with the hypothalamus-mask thresholded at z=10 (chosen to get a constrained mask of the hypothalamus), to create a bilateral ROI mask for voxel-wise primary analysis. Primary mask was used as described above, a secondary mask, was used based on the peak voxel clusters and 4mm spheres around those (done in fslmaths).

**Power analysis.** A power analysis has been conducted based on a study by Tiedemann et al. 2017 Nature Communications, testing the effect of a metabolic intervention on the preference value for food. We estimated an effect size of n2 = 0,12 or f = 0,37 to reach significance for rejecting the null hypothesis based on repeated-measures ANOVA. Since power analysis resulted in a sample size of 50, we aimed for 60 participants including a 20% dropout-rate.

### **Brain mask: Neurosynth-based rawNeurosynth-based peak clusters**

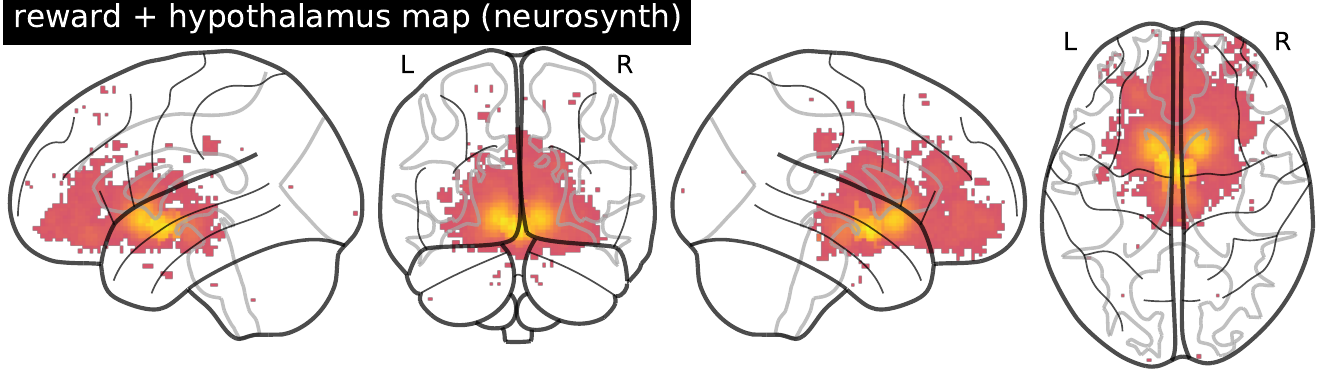

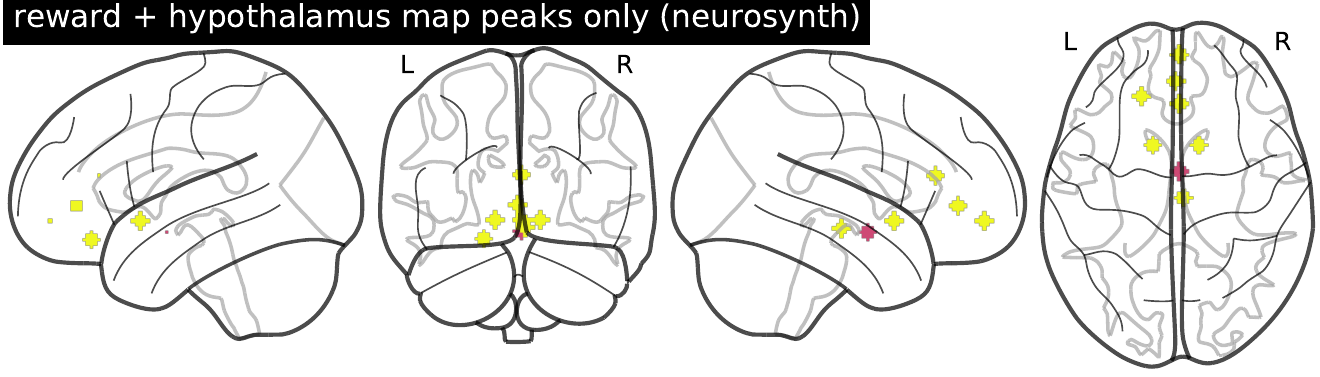

**fMRI 2nd level masks.** Clusters used for brain activity extraction and in network analysis. Interaction effects in VTA, rOFC and rmOFC (left), peak voxel spheres reward and hypothalamus network (right), main effect of food wanting (model A, con 01, 03, 05) (not shown).

**Whole-brain analysis.** Whole-brain results are also reported using MNI152 T1 2mm brain mask as explicit mask.

**fMRI 2nd level specifications**

**Hypothesis testing.** The modified SwE approach was selected, with all subjects modeled as one group assuming that all subjects share a common covariance matrix. Visits (1,2,3,4) and subjects (1 to 60) were entered according to SwE structure, and four binary covariates for intervention*timepoint were modeled (placebo_BL, placebo_FU, verum_BL, verum_FU) matching the input order of the images. Type 3 small sample bias adjustment and for estimating the degrees of freedom the “pprox. II” option to account for missing data without assuming a missing-data-bias were used.

**Contrast matrices**. Main effects were modelled as [0.25 0.25 0.25 0.25] and interaction effects as [-1 1 1 -1] with the predictors [placebo_BL, placebo_FU, verum_BL, verum_FU].

**Thresholding.** Repeated measures accounted for by within-subject correlation were estimated at TFCE-p-FEW<0.05 for masked brain areas using SwE default parameters (E = 0.5, H = 2). Additionally, results with TFCE-p<0.001 uncorrected are reported. Before non-parametric wild bootstrap resampling (999 permutations), residuals were small sample adjusted with “type C2” (referred to as type 3). TFCE is used to avoid a priori definition of a threshold, however, coming at the cost of a spatial bias [7].

**Results – fMRI 2^nd^ level main analyses.**

### Out of 59 included participants, fMRI data from 57 participants, from at least one and up to four timepoints passed quality control, summing up to in total 200 neuroimaging measurements for inclusion into main analyses.

### Results are reported for primary analysis using the thresholded Neurosynth mask (left) only.

### model A con_0001 food > art

### model A con_0002 art > food

### model A con_0003 food wanting slope > art wanting slope

### model A con_0004 art wanting slope > food wanting slope

### model A con_0005 wanting modulation

### model B1 con_0001 wanting*kcal

### con_0002 kcal

### model B2 con_0001 wanting*fiber

### con_0002 fiber

### model C1 con_0001 food > art

### con_0002 art > food

### con_0003 food wanting slope > art wanting slope

### con_0004 art wanting slope > food wanting slope

### con_0005 wanting modulation

### model C2 con_0001 food liking slope > art liking slope

### con_0002 art liking slope > food liking slope

### con_0003 liking modulation

### **Primary analysis (pre-registered):**

| **Models considering parametric wanting ratings**  **models A, B1, B2** | **Models considering parametric wanting and liking ratings**  **model C1** | **Models considering parametric liking ratings**  **model C2** |
| --- | --- | --- |
| food > art (TFCE-p_FWE_ < 0.05) 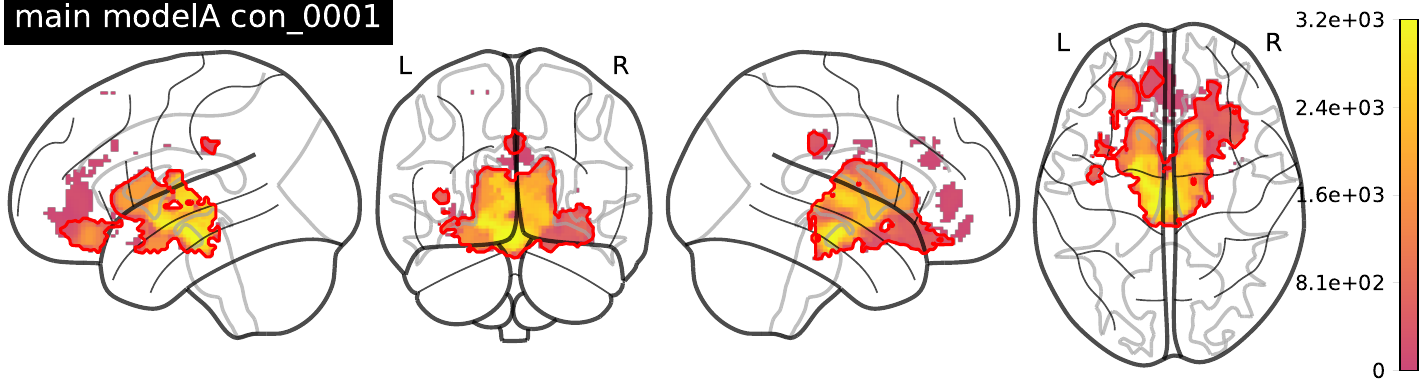 | food > art (corrected for liking as parametric modulator of no interest) (TFCE-p_FWE_ < 0.05) 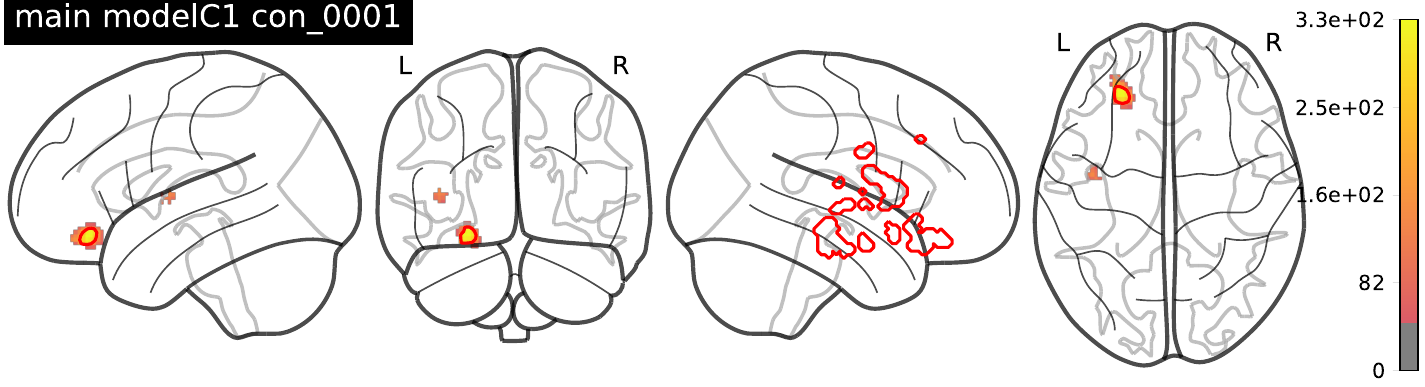 |  |
| art > food (TFCE-p_FWE_ < 0.05) 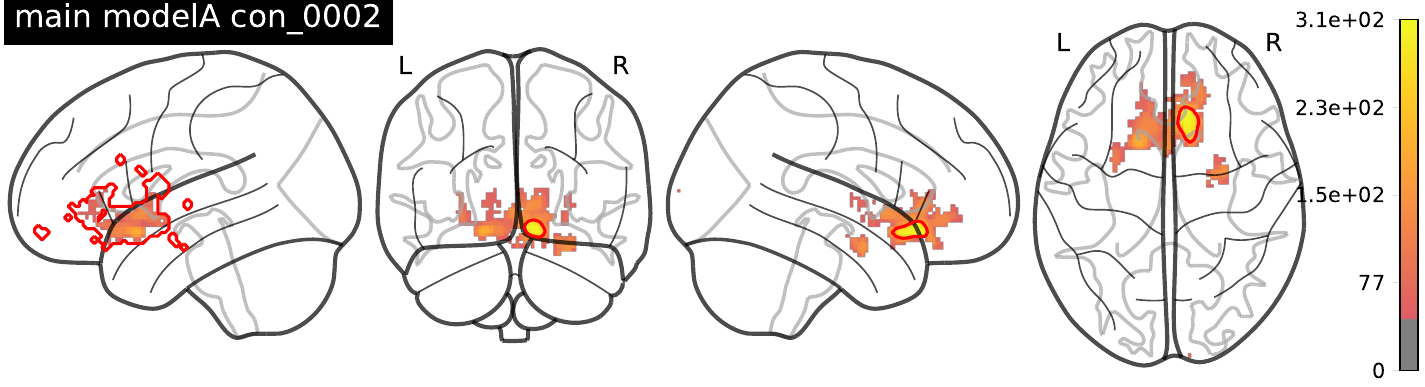 | art > food (corrected for liking as parametric modulator of no interest) (TFCE-p_FWE_ < 0.05) 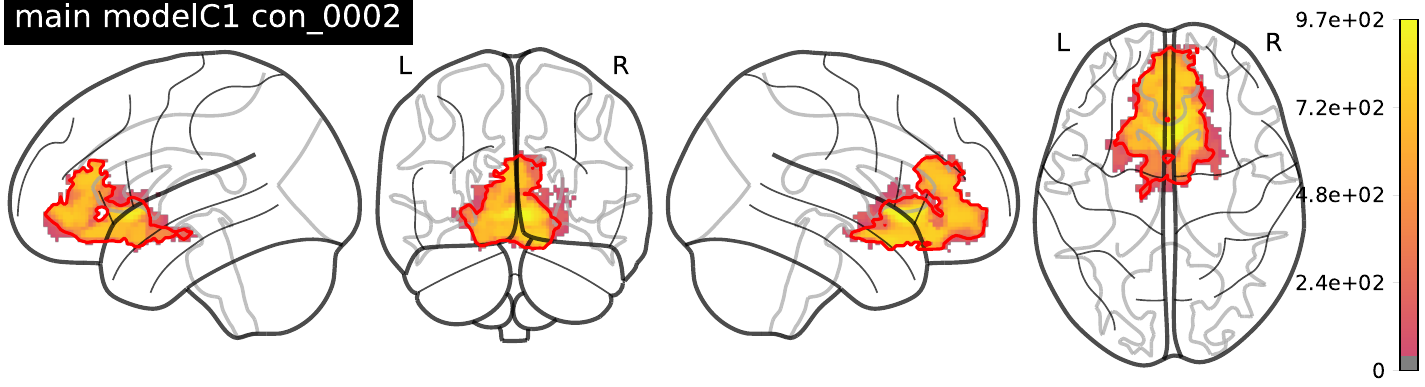 |  |
| food wanting slope > art wanting slope (TFCE-p_FWE_ < 0.05) 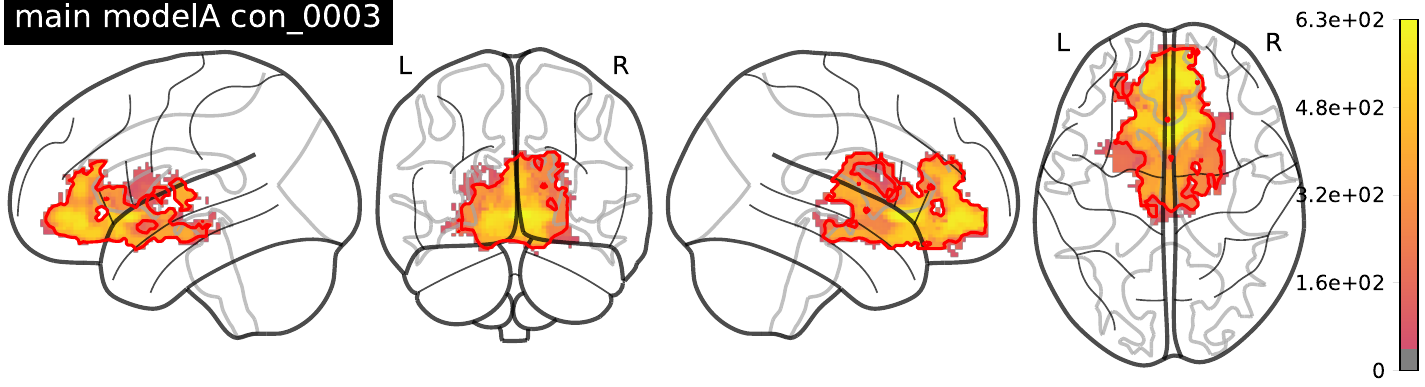 | food wanting slope > art wanting slope (corrected for liking as parametric modulator of no interest) (TFCE-p_FWE_ < 0.05) 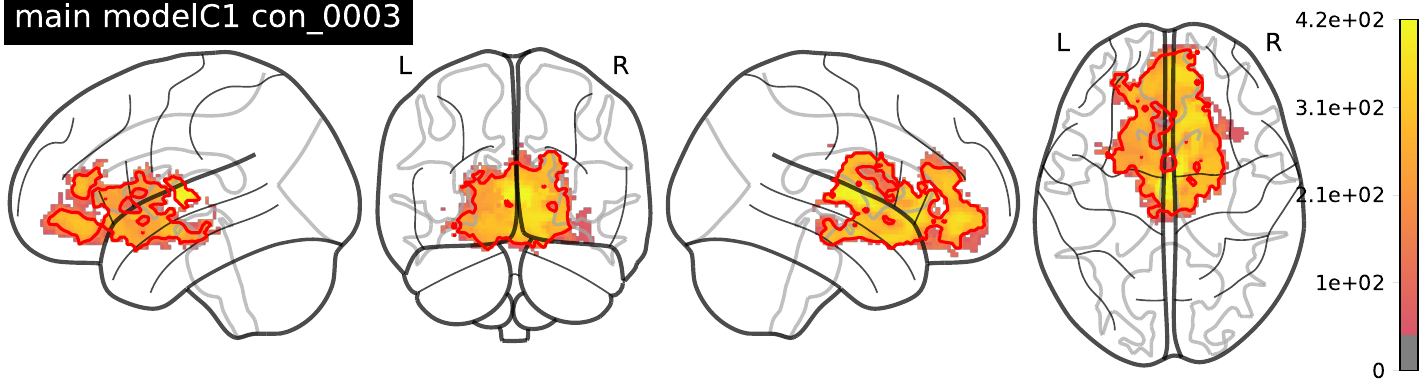 | food liking slope > art liking slope (TFCE-p_FWE_ < 0.05) 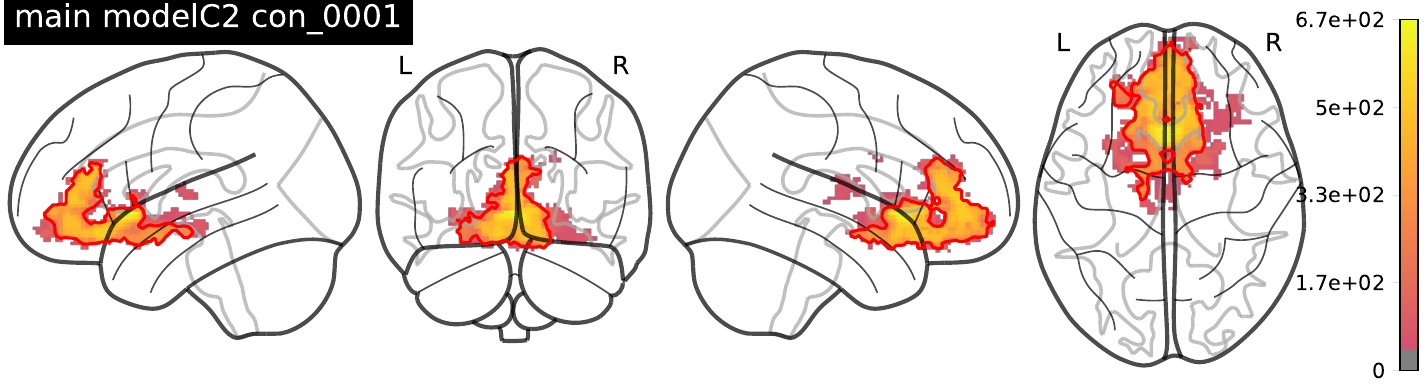 |
| art wanting slope > food wanting slope (n.s.) 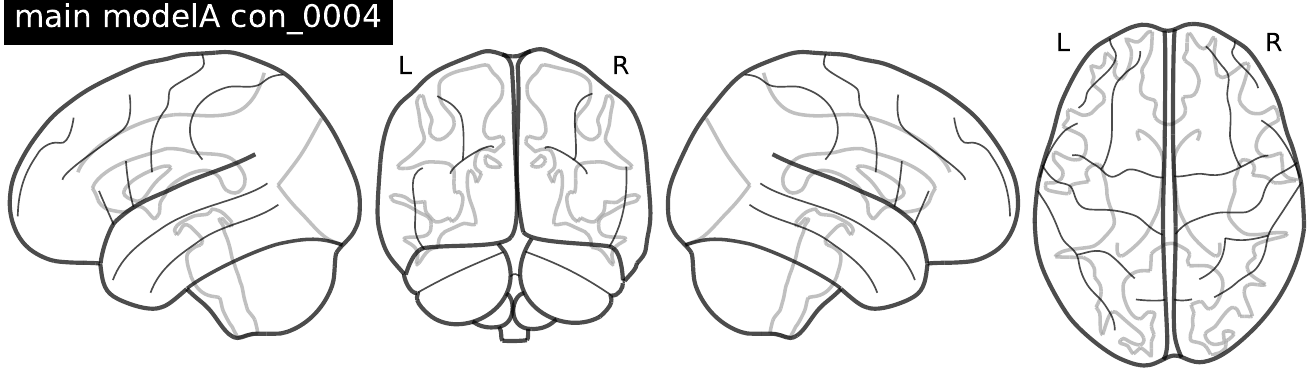 | art wanting slope > food wanting slope (corrected for liking as parametric modulator of no interest) 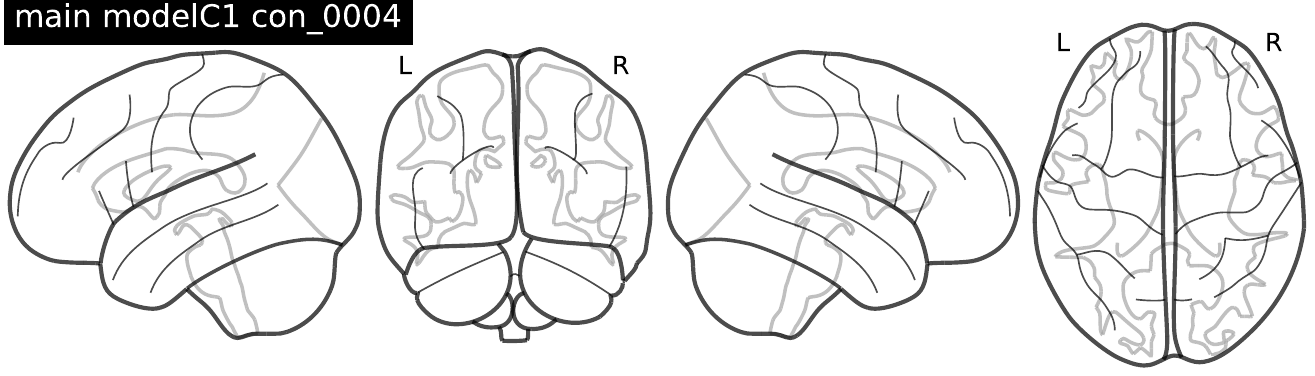 | art liking slope > food liking slope (n.s.) 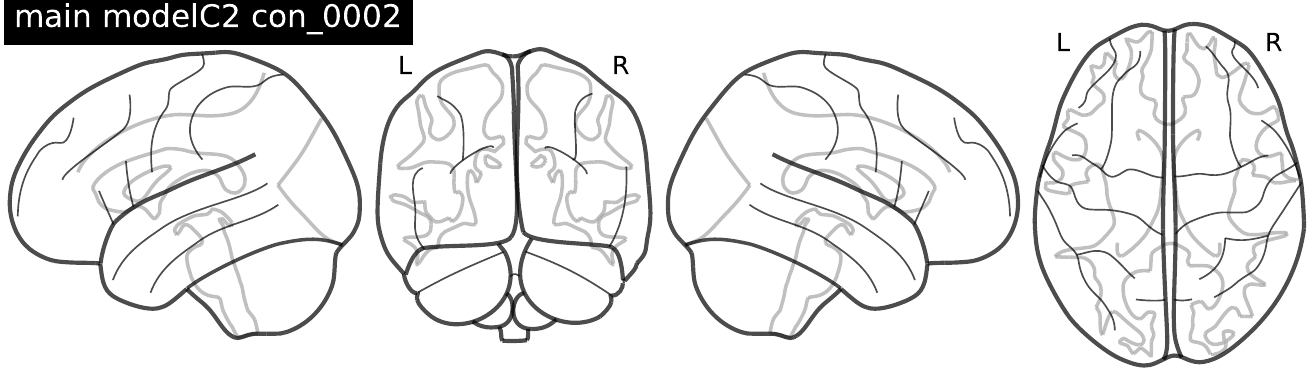 |
| wanting modulation (food & art) (TFCE-p_FWE_ < 0.05) 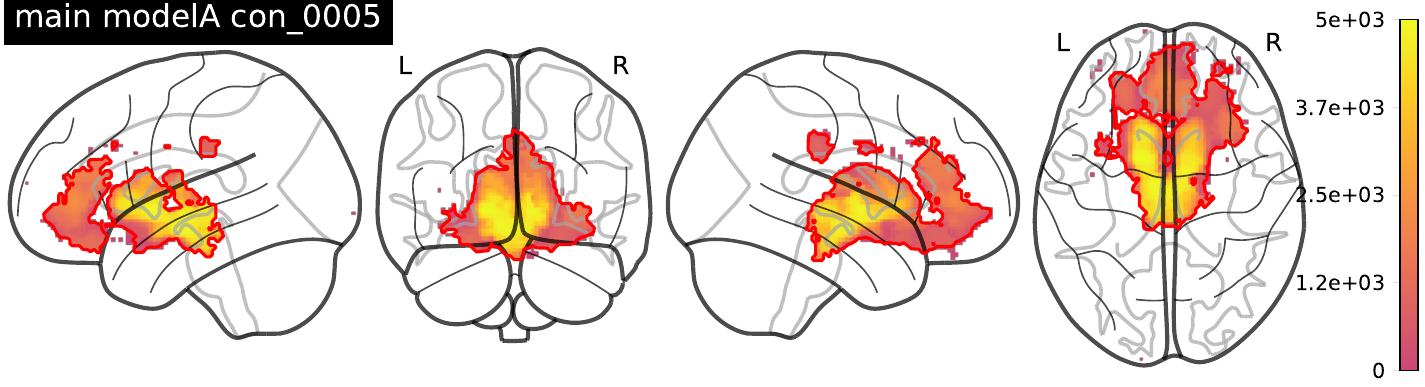 | wanting modulation (food & art) (corrected for liking as parametric modulator of no interest) (TFCE-p_FWE_ < 0.05) 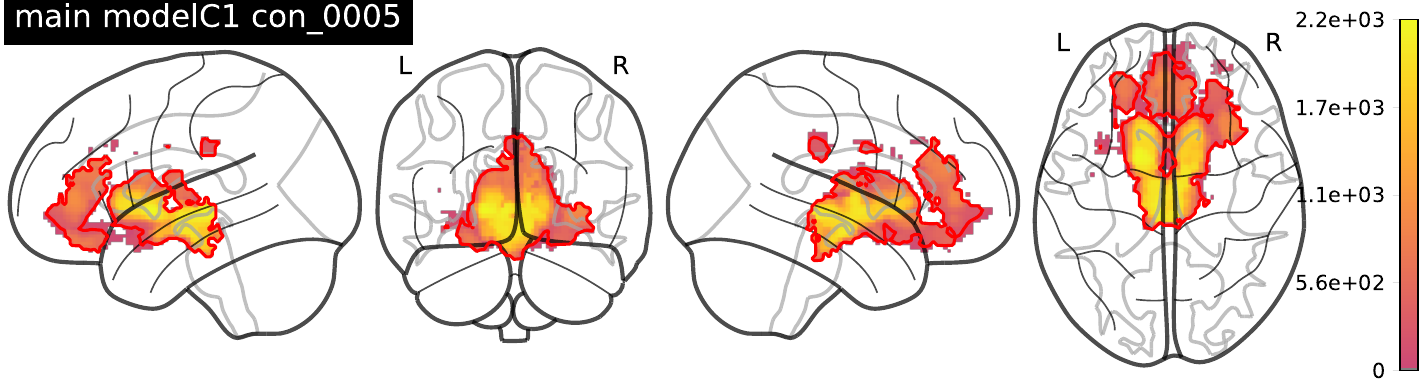 | liking modulation (food & art) (TFCE-p_FWE_ < 0.05) 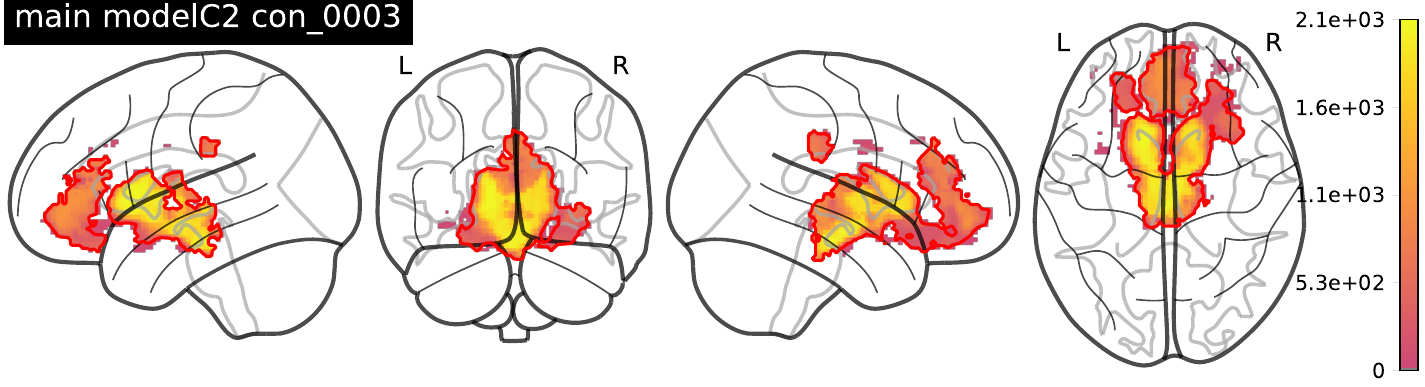 |
| wanting*kcal (n.s.) 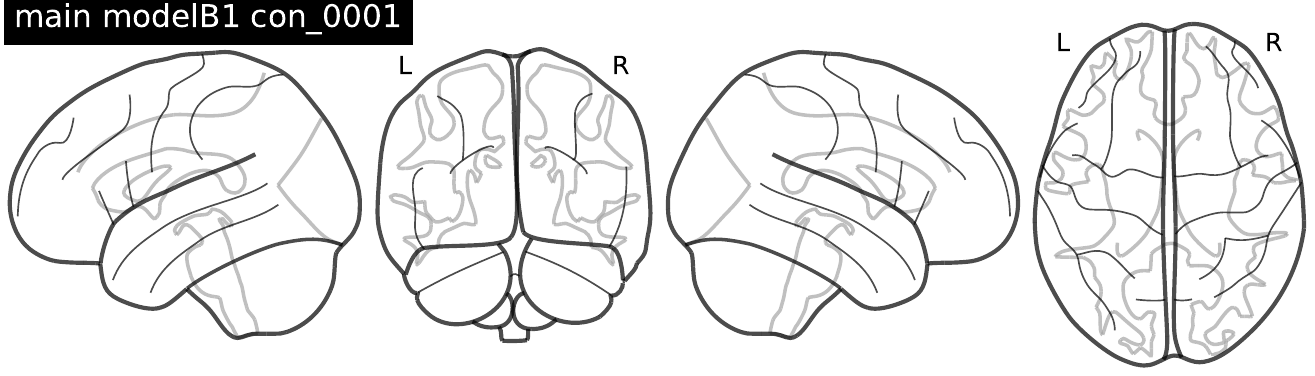 |  |  |
| kcal (TFCE-p_FWE_ > 0.05) 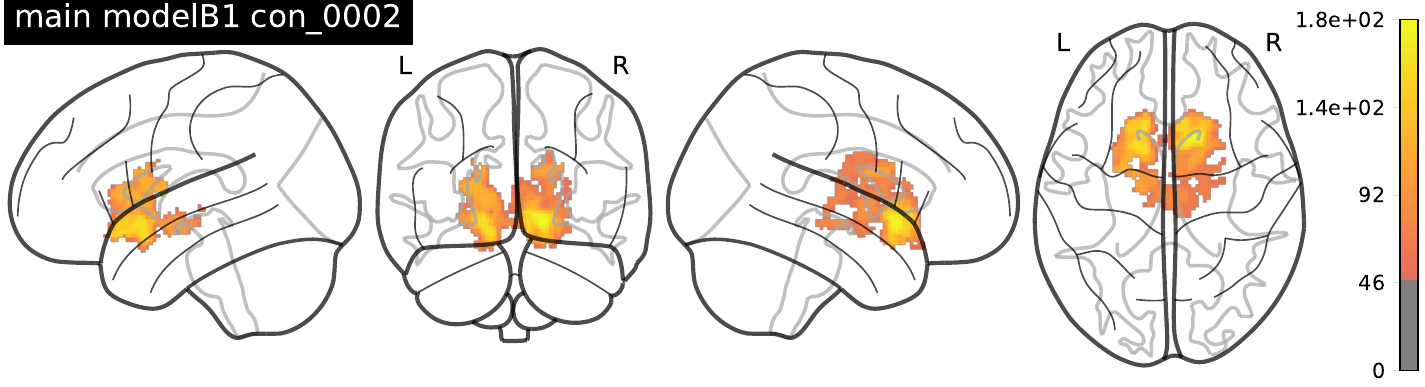 |  |  |
| wanting*fiber (n.s.) 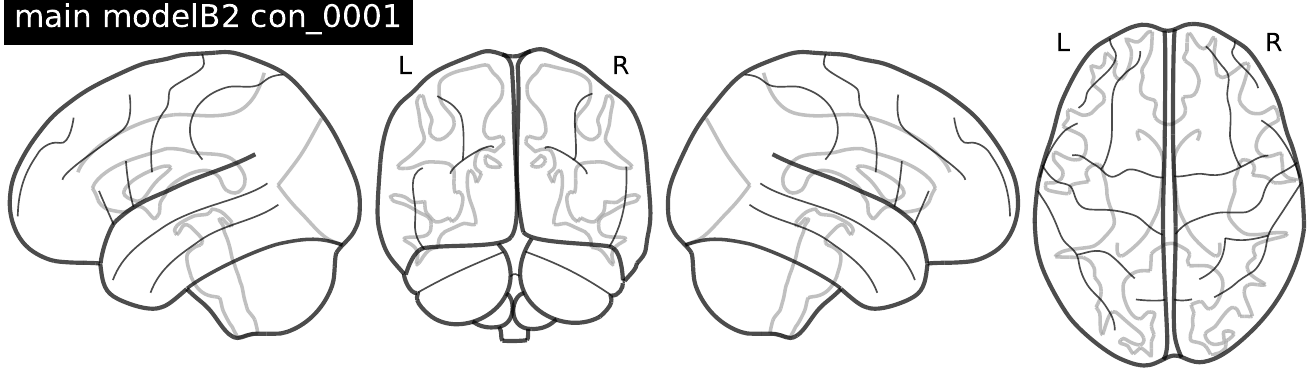 |  |  |
| fiber (n.s.) 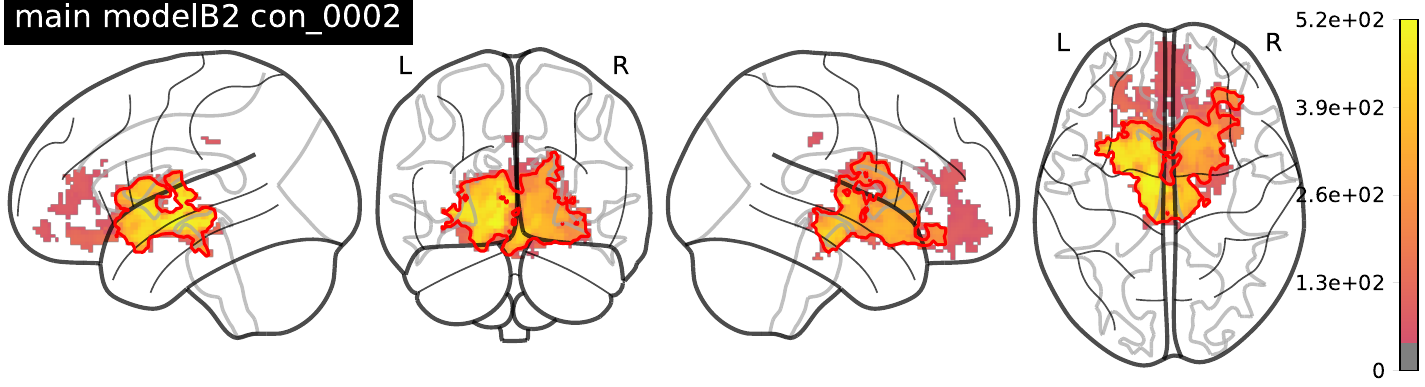 |  |  |

***SI Figure 3: Main effects for food wanting-related activity****. Statistically significant BOLD activity depicting parametric TFCE t-statistic (min = 50 for visualizing purposes) and wild-boot strapped p-_FWE_ < 0.05 (p_FWE_-corrected permutation test results delineated as red contour). Column 1 depicts all statistical models related to wanting ratings only, column 2 depicts models as in column 1 but additionally with liking ratings added as a confounder of no interest, and column 3 reflects certain models of column 1 only including parametric liking ratings only as a parametric modulator of interest. All models were run on the 2^nd^ level using Neurosynth mask raw as shown before.* *Input images computed with SwE toolbox and plotted with nilearn.*

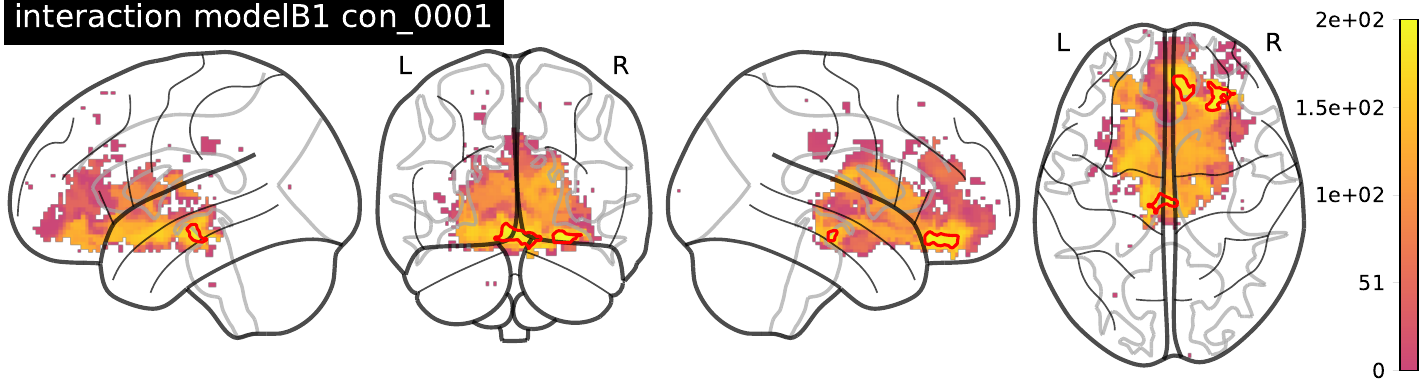

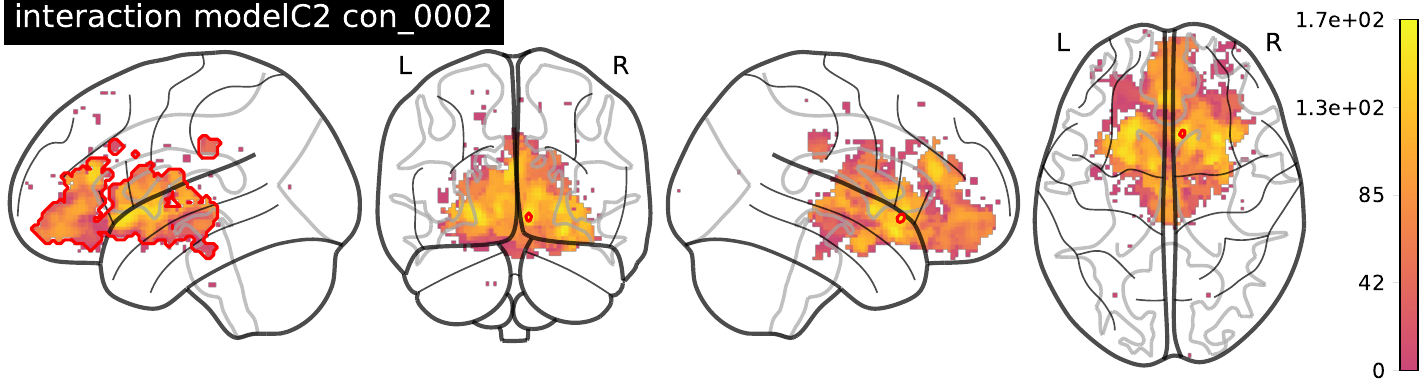

### ***SI Figure 4: Prebiotic diet induced changes in food wanting-related activity contrasted to post-placebo intervention****. Statistically significant BOLD activity depicting parametric TFCE t-statistic (min = 0) and wild-boot strapped p-_FWE_ < 0.05 (p_FWE_-corrected permutation test results delineated as red contour) for post-prebiotic intervention compared to post-placebo intervention for the activation contrast for wanting*kcal for all available MR datapoints (n=200 in 57 individuals). Neurosynth raw mask used as 2^nd^ level brain mask. Input images computed with SwE toolbox and plotted with nilearn.*

### **Secondary sensitivity analysis (pre-registered):**

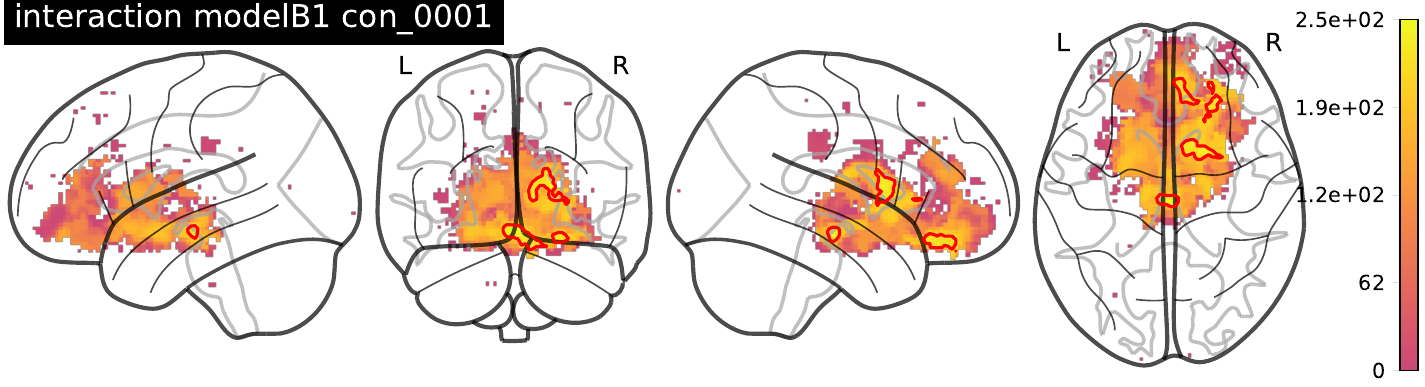

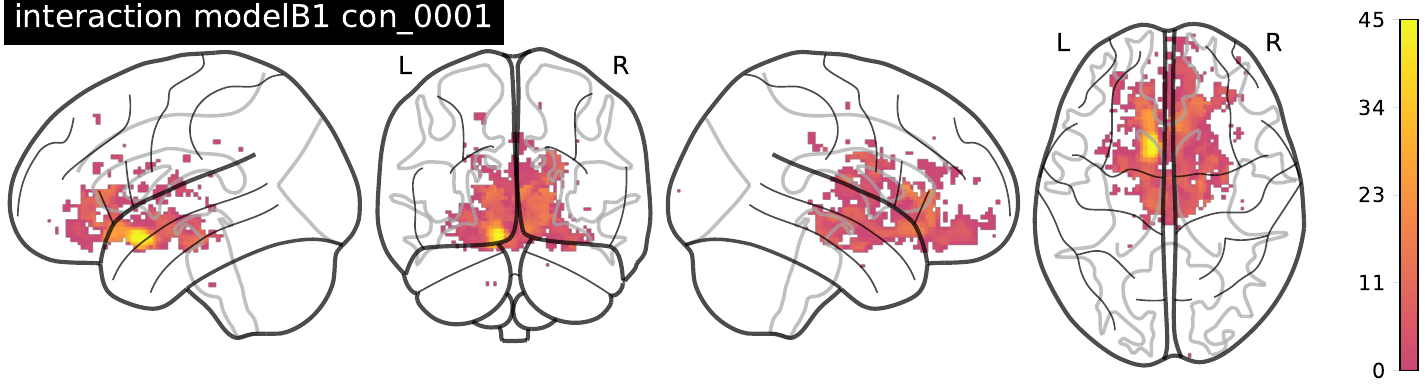

### ***SI Figure 5: Sex-stratified prebiotic diet induced changes in food wanting-related activity contrasted to post-placebo intervention****. Top: Male only (n = 38), Bottom: Female only (n = 19).*

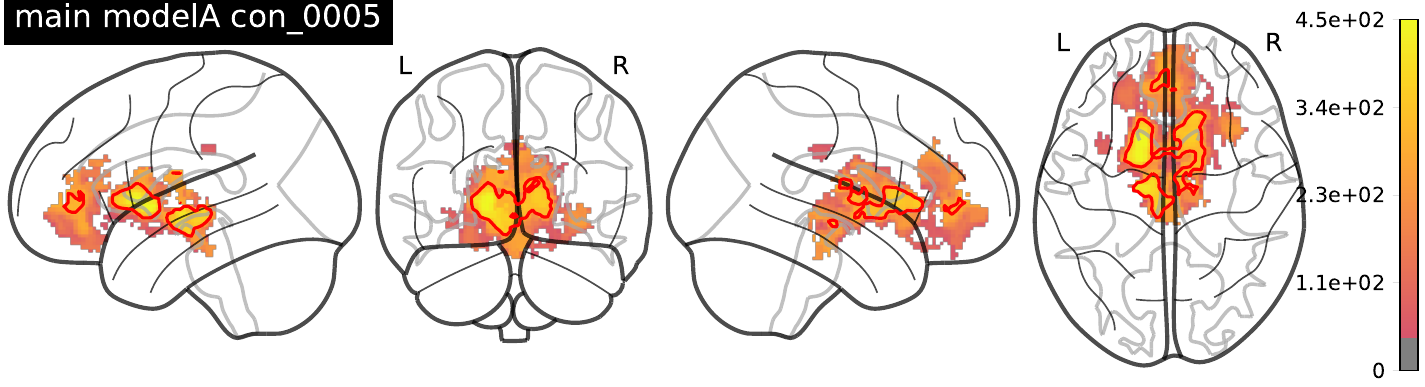

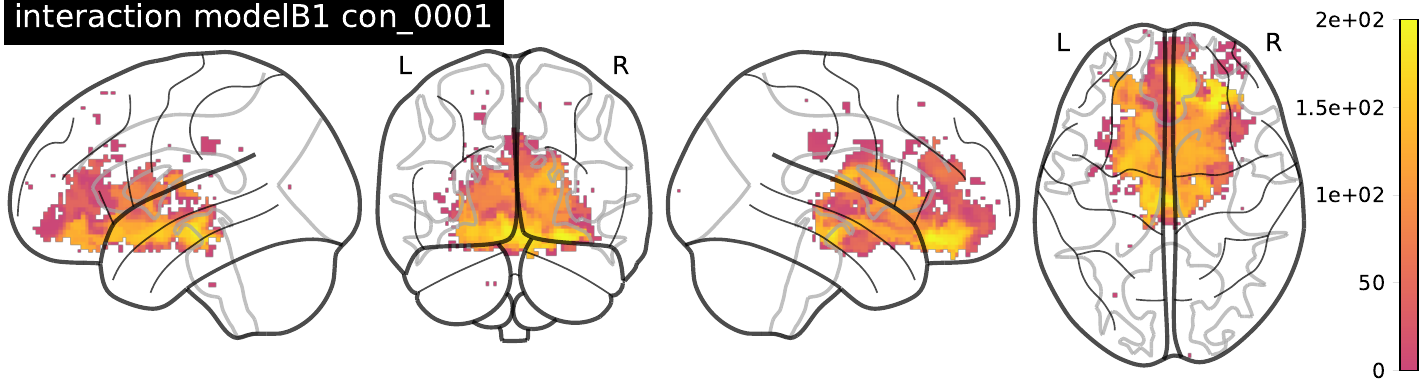

***SI Figure 6: Main results corrected for confounding factors age, sex and SES index.*** *Top: Main effect of wanting modulation (model A), Bottom: Interaction effect of timepoint*intervention showing prebiotic diet induced changes in food wanting-related activity contrasted to post-placebo intervention (model B1).*

### ***Sensitivity analyses for model B1 – kcal*wanting interaction effects***

| ***Additionally corrected for fiber intake*** 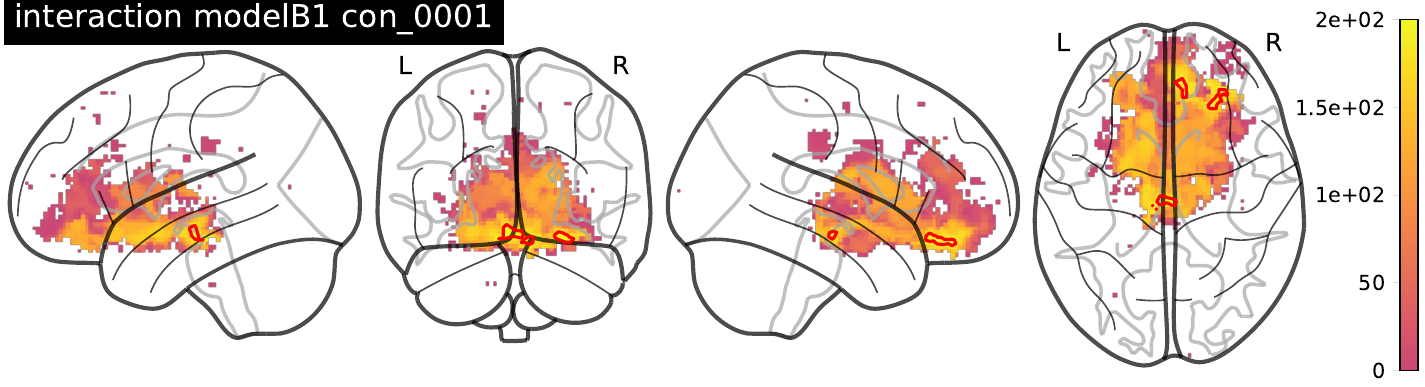 | ***Additionally corrected for post-hunger*** 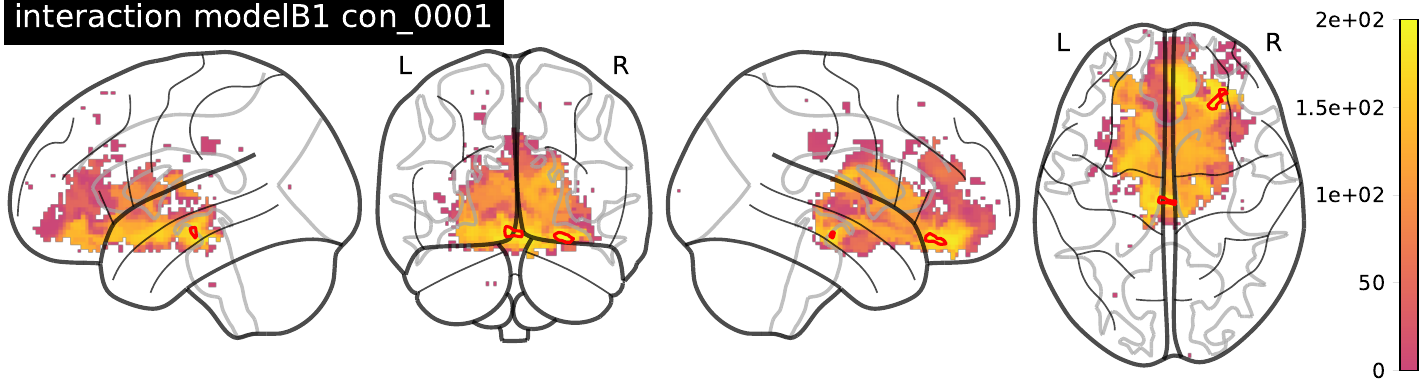 |
| --- | --- |
| ***Additionally corrected for body fat mass*** 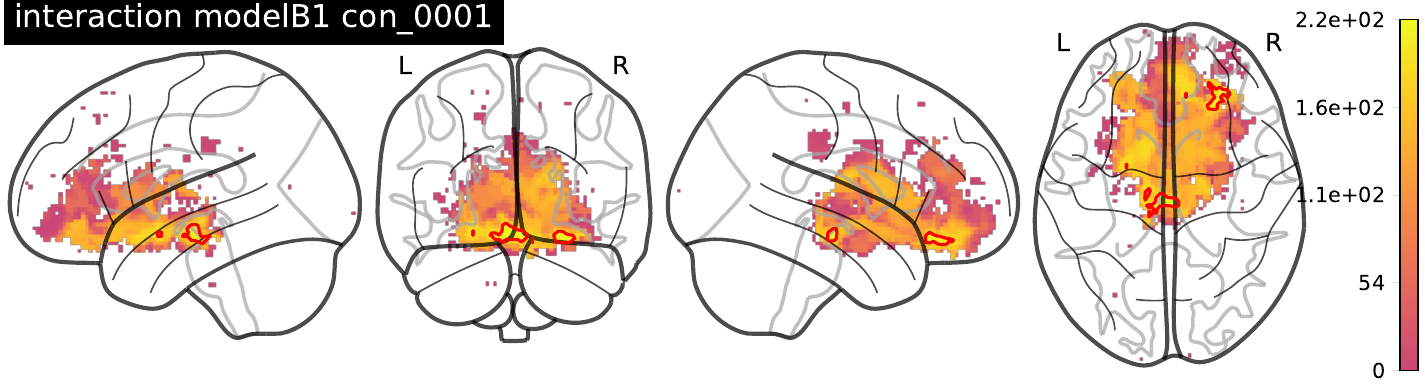 | ***Additionally corrected for SES index*** 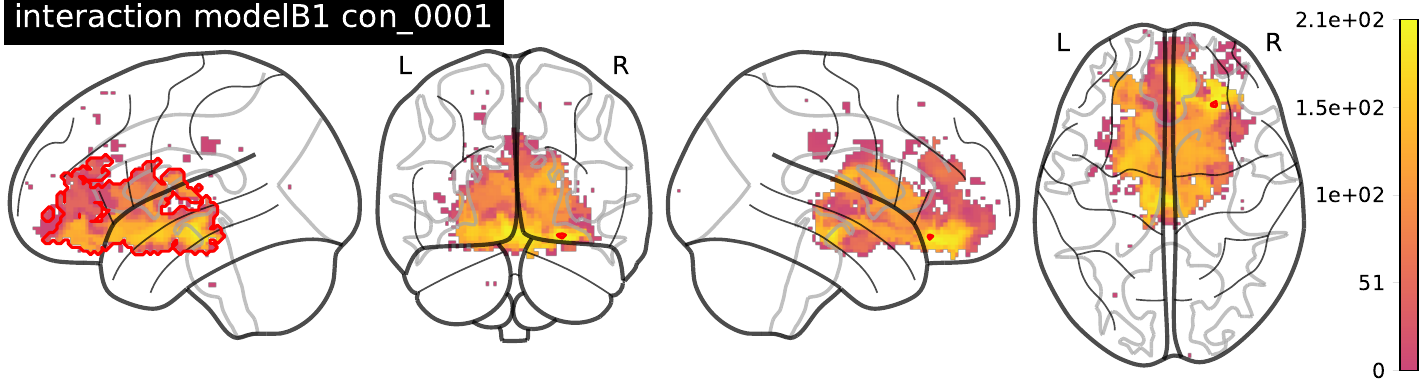 |
| ***Additionally corrected for pre-hunger*** 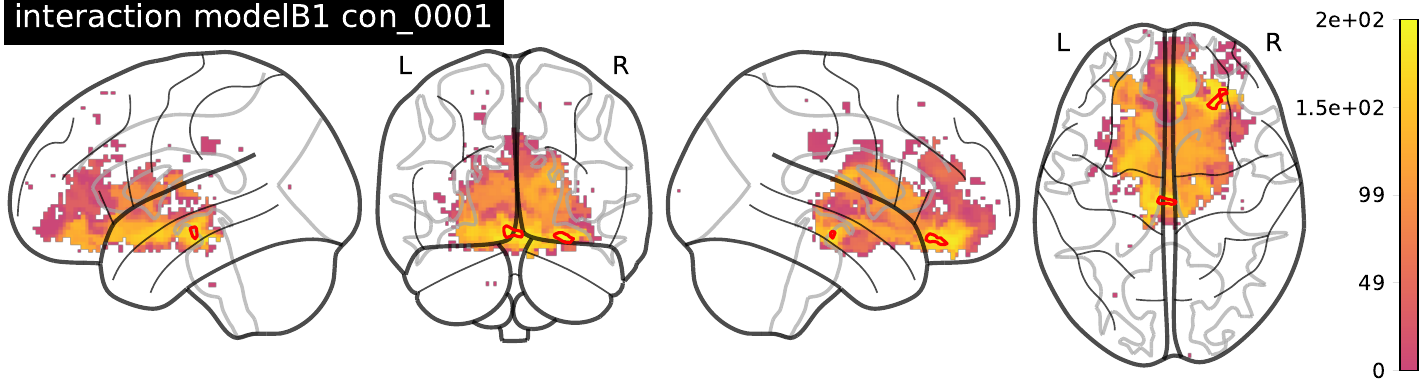 | ***Additionally corrected for VAIAK score*** 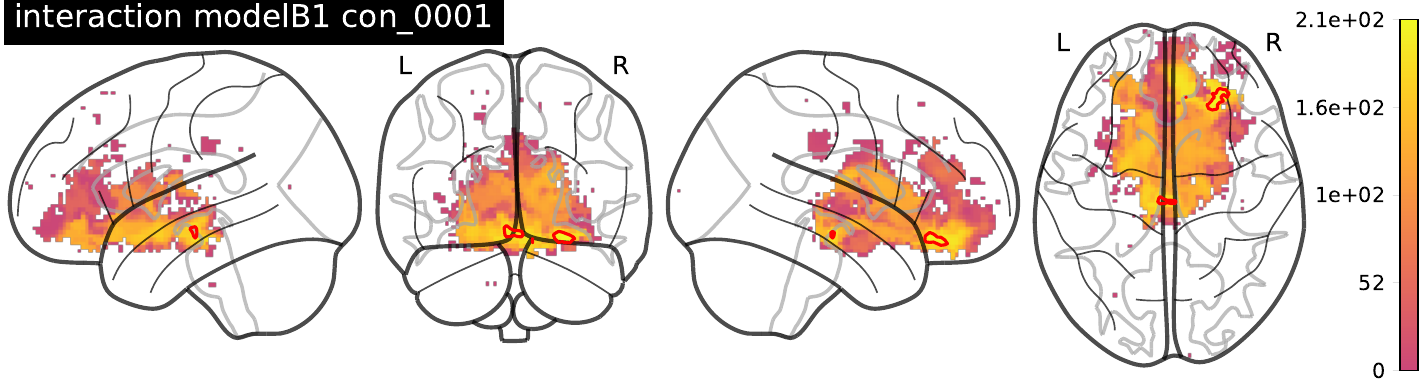 |
| ***Additionally corrected for well-being during scanning*** 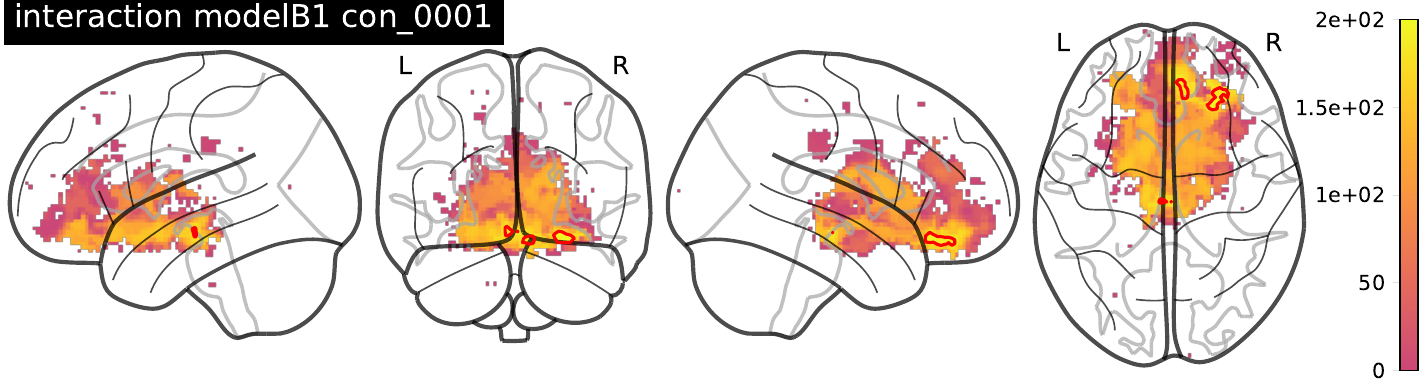 |  |

***SI Figure 7: Main results corrected for confounding factors age, sex and SES index.*** *Top: Main effect of wanting modulation (model A), Bottom: Interaction effect of timepoint*intervention showing prebiotic diet induced changes in food wanting-related activity contrasted to post-placebo intervention (model B1).*

### ***Model B1 – kcal only (con_0002) main effects***

### ***Additionally corrected for body fat***

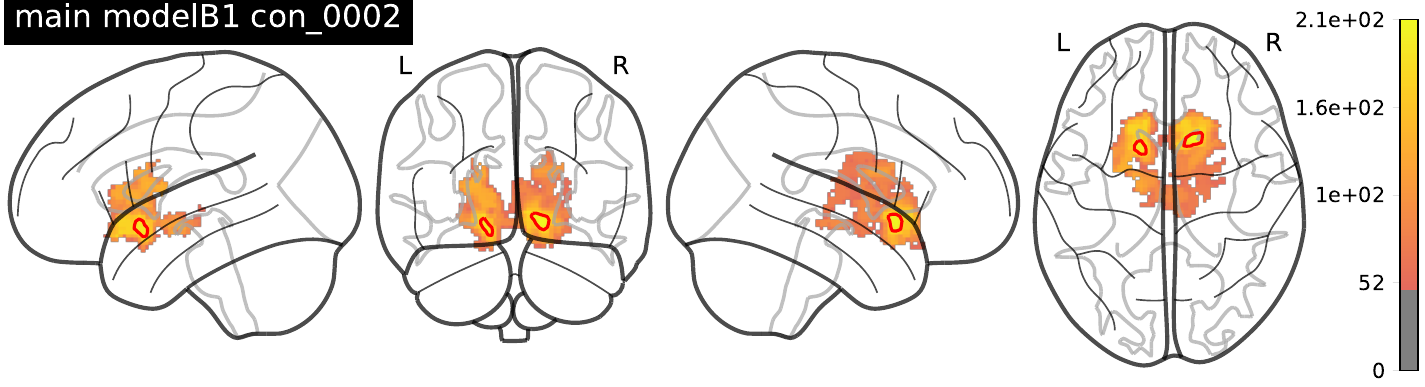

***SI Figure 8: Main results (model B1, kcal only) corrected for confounding factor body fat mass.***

### ***Model A – wanting modulation (food & art) effects***

### ***Additionally corrected for fiber intake***

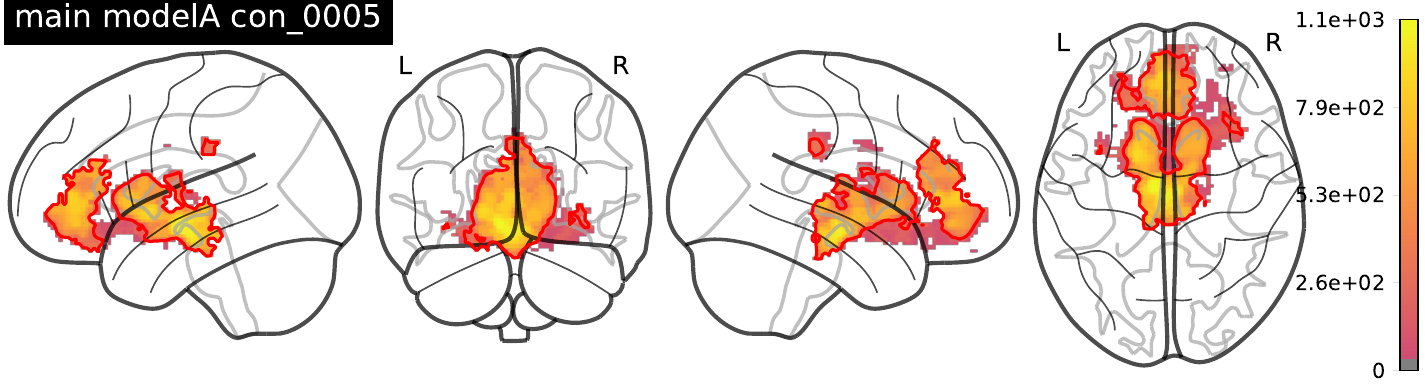

***Additionally corrected for body fat mass***

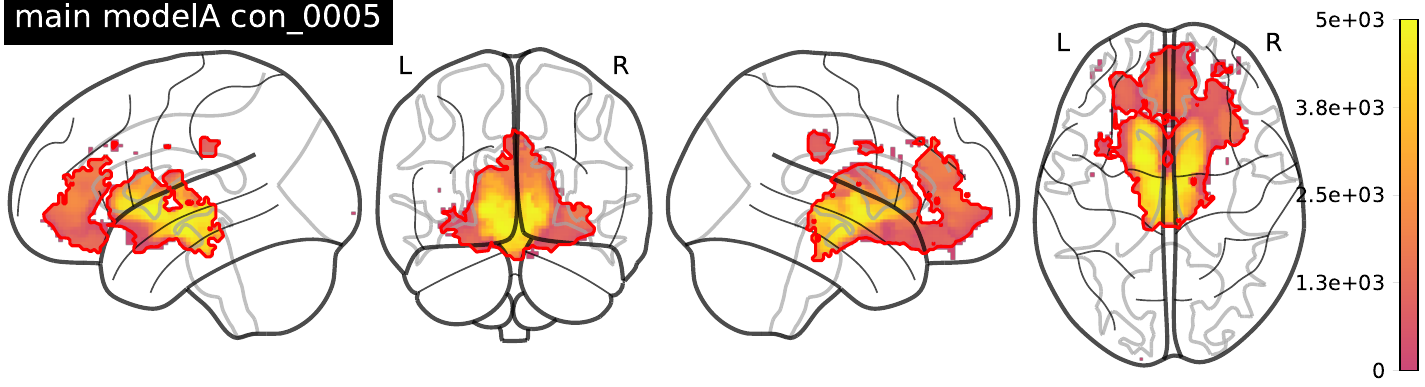

***Additionally corrected for pre-hunger***

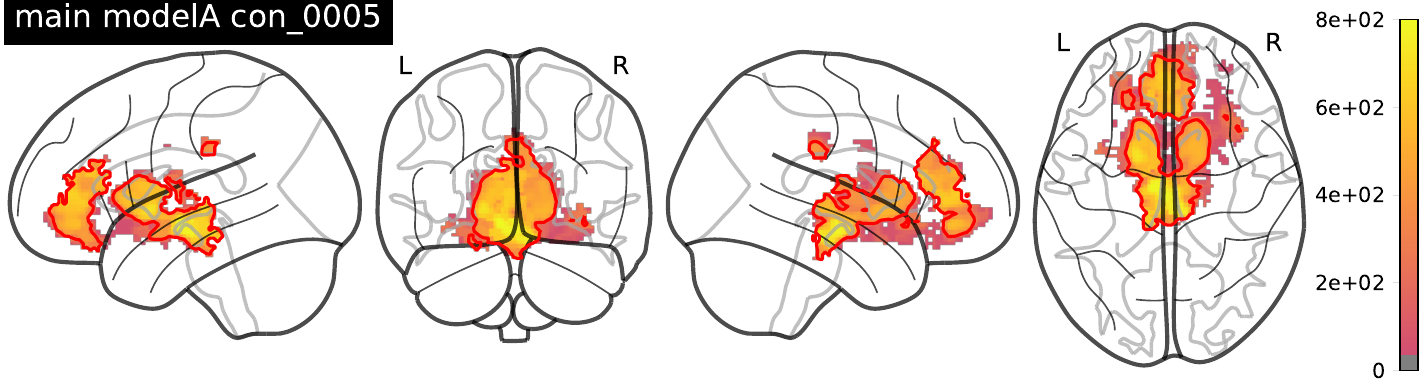

***Additionally corrected for post-hunger***

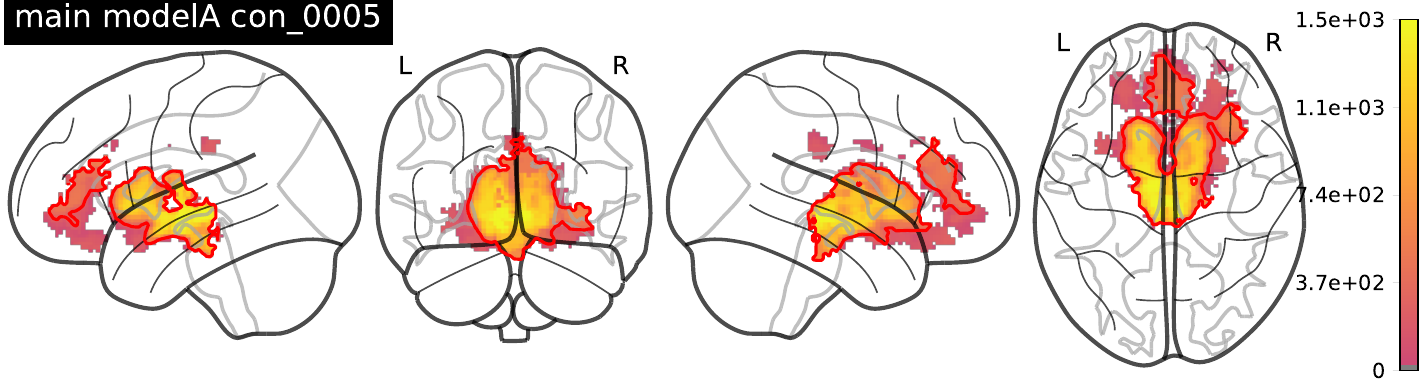

***Additionally corrected for VAIAK score***

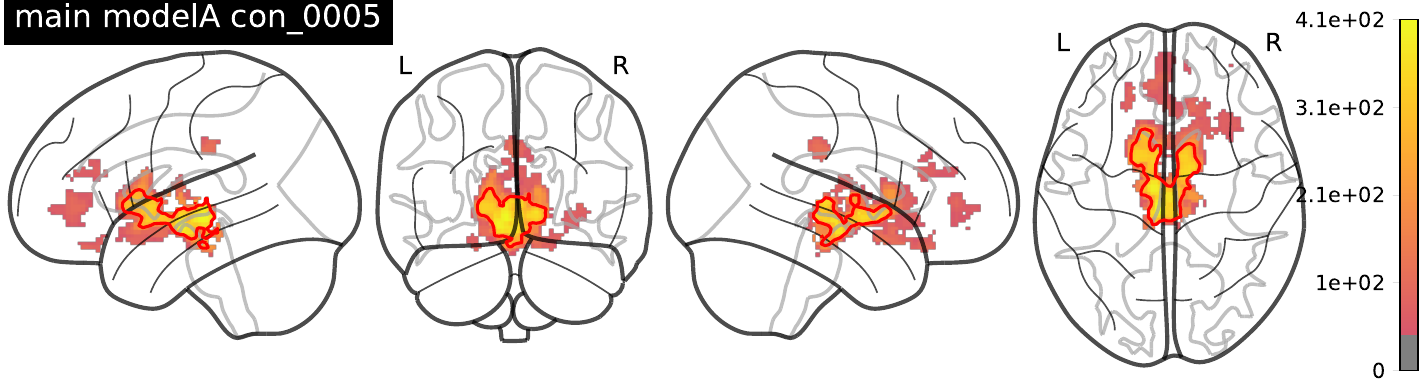

***Additionally corrected for well-being during scanning***

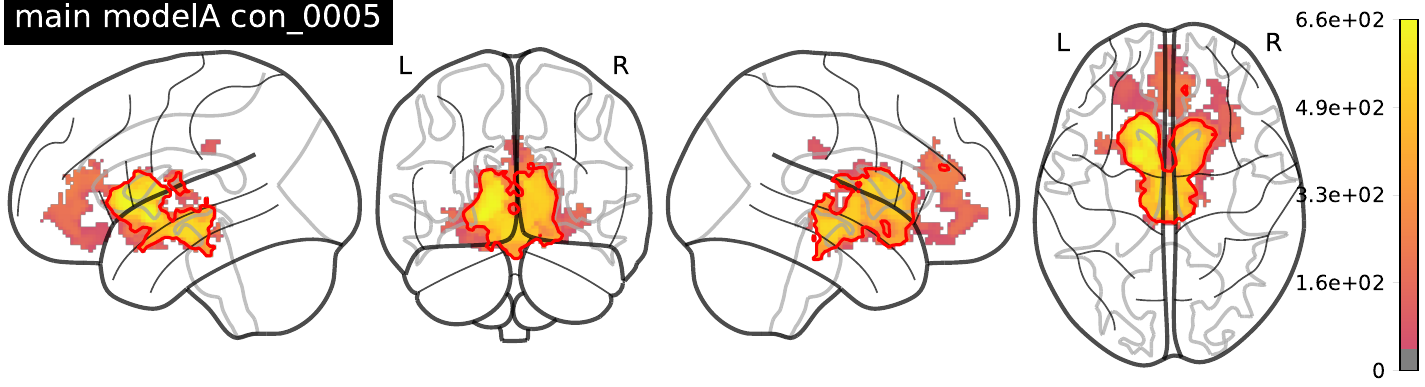

***Additionally corrected for SES index***

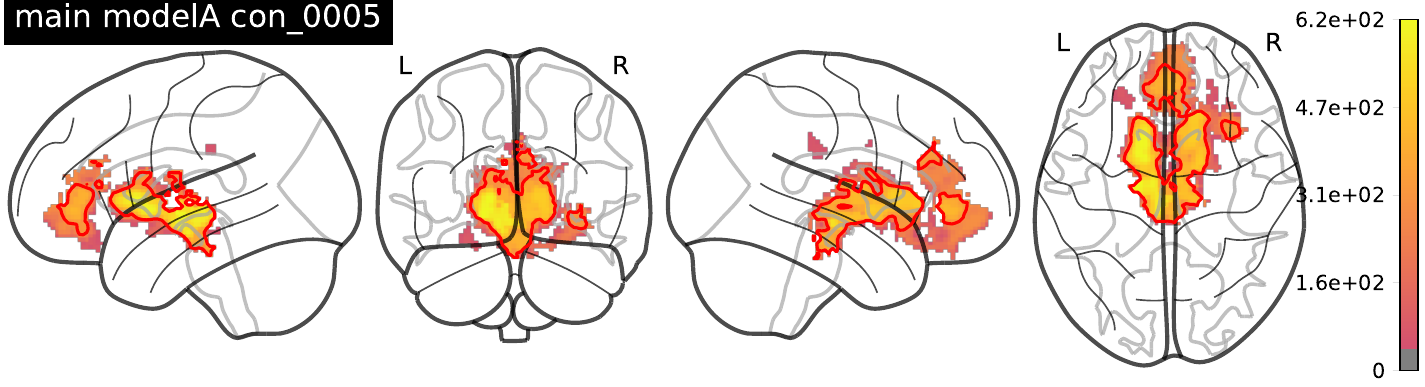

***SI Figure 9: Main results (model A (food and art wanting) corrected for various confounding factors.***

***Sensitivity and exploratory fMRI analyses***

To explore differences in fMRI results with regard to gender, we conducted gender-stratified analyses (19 women and 38 men, respectively) as pre-registered. Overall, results were similar to the whole-sample analysis, yet with one more cluster emerging in the male sample, and results in the female sample only did not survive pFWE-correction.

Further, we repeated analyses adding age, gender and SES index as confounding factors of no interest to the models. Here, the interaction effect of timepoint and intervention did not survive pFWE-correction, yet activation patterns were comparable to the uncorrected analysis (**SI_fMRI Figures 6-7**). See **SI_fMRI** and **SI_behav** for further constrained ROI-based and whole-brain as well as behavioral analyses according to the preregistration. All unthresholded TFCE maps are available on neurovault.org (<https://identifiers.org/neurovault.collection:14111>).

Note that deviant to the preregistration, we restricted further sensitivity analyses to the main contrast of interest that yielded significant results (i.e., food wanting by calorie content). As preregistered, we corrected interaction effects for *a priori* known confounders of interest (dietary fiber intake, gender-standardized % body fat, subjective hunger ratings, SES index, VAIAK score, well-being inside of scanner). Correcting for habitual fiber intake and well-being yielded the same three significant clusters, whereas when correcting for body fat mass, hunger or VAIAK only the VTA and the rOFC clusters survived p-FWE-correction. When correcting for SES index, only the clusters in the rOFC survived thresholding (**SI_fMRI Fig. 8**).

Also, when excluding one participant who reported depressive symptoms based on questionnaires or one participant who reported to not have taken the supplement for 48h (4 sachets), analyses showed similar results (VTA cluster, rOFC partly) (**see SI_fMRI Fig. 10 and SI_fMRI Table 1**).

excluded sub-30 (depressive symptoms):

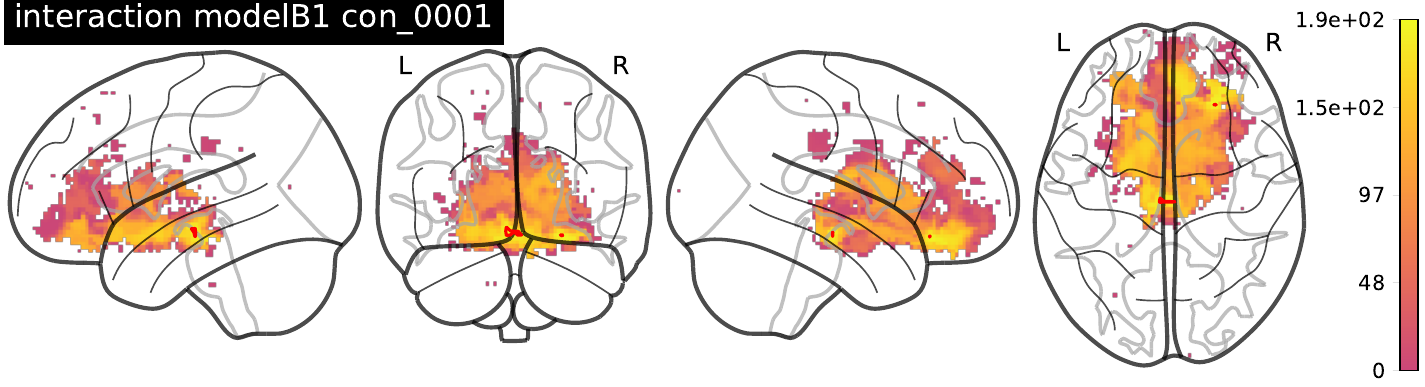

excluded sub-47 (>48h no supplement):

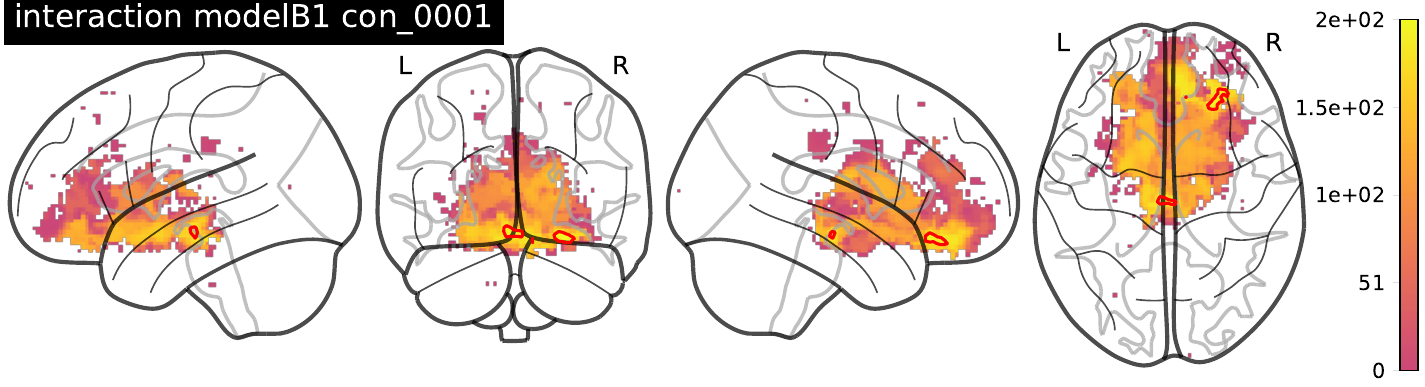

both excluded (n.s.):

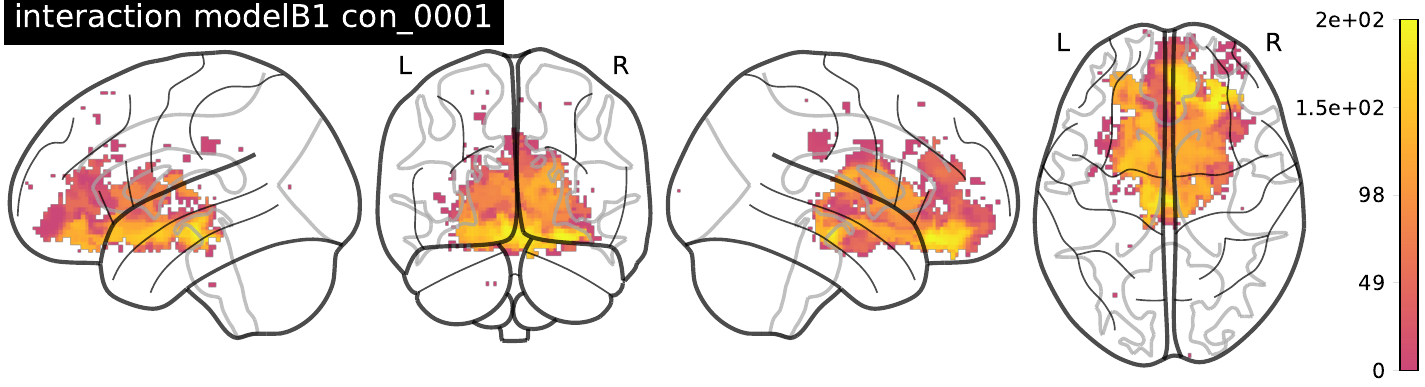

***SI Figure 10: Interaction results (model B1, kcal*wanting) regarding excluded participants.***

***SI Table 1: Interaction results (model B1, kcal*wanting) regarding excluded participants.***

| **Model** | **TFCE_**  **p(FWE-corr)** | **TFCE_**  **clustersize** | **peak_**  **p(FWE-corr)** | **peak_**  **p(FDR-corr)** | **peak_Z** | **peak_**  **p(unc)** | **x**  **{mm}** | **y**  **{mm}** | **z**  **{mm}** | **assigned region** |
| --- | --- | --- | --- | --- | --- | --- | --- | --- | --- | --- |
| **Model B1 (wanting*kcal)**  **- act**  **Excluded sub-30 ses-01** | 0.049 | 1 | 0.18 | 0.171 | 3.509 | 0.001 | 26 | 32 | -16 | right OFC |
|  | 0.047 | 10 | 0.239 | 0.171 | 3.406 | 0.001 | -4 | -20 | -14 | VTA |
| **Model B1 (wanting*kcal)**  **- act**  **Excluded sub-47 ses-04** | 0.038 | 21 | 0.132 | 0.128 | 3.595 | 0.001 | 26 | 32 | -16 | right OFC |
|  | 0.04 | 15 | 0.238 | 0.128 | 3.388 | 0.001 | -4 | -20 | -14 | VTA |
|  | 0.05 | 2 | 0.44 | 0.128 | 3.075 | 0.001 | 10 | 36 | -20 | right OFC |
| **Model B1 (wanting*kcal)**  **- act**  **Excluded sub-30 ses-01 and sub-47 ses-04** | - | - | - | - | - | - | - | - | - | - |

**Extracted brain activation clusters used for network analysis**

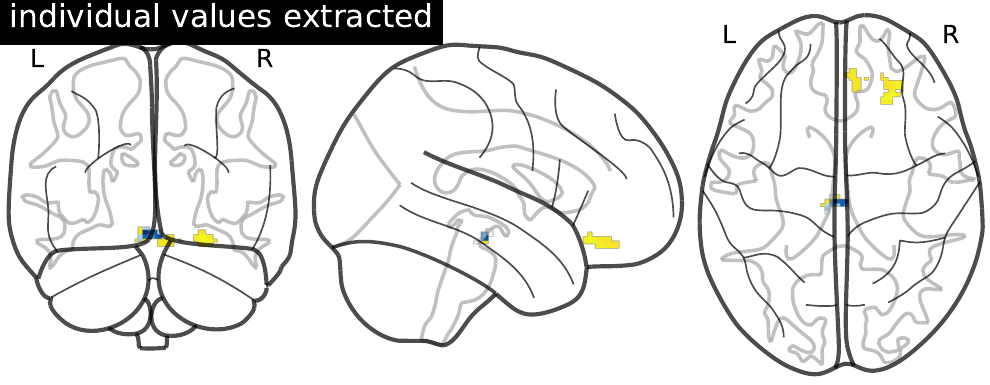

Using a secondary mask based on peak voxel activity of meta-analytic neurosynth maps, no significant clusters emerged for all models, except for model C2 (art liking slope > food liking slope), which resulted in one significant cluster in mPFC/ACC.

| TFCE_p(FWE-corr) | TFCE_ | TFCE_equivk | TFCE_ | peak_p(FWE-corr) | peak_p(FDR-corr) | peak_Z | peak_p(unc) | _x | y | z {mm} |
| --- | --- | --- | --- | --- | --- | --- | --- | --- | --- | --- |
| 0.011 | 12 |  |  | 0.017 | 0.041 | 3.338 | 0.001 | 0 | 34 | 16 |

For whole-brain analysis, no clusters survived in all models, except for model B1 (wanting*kcal), which resulted in four significant clusters in cerebellar regions.

| TFCE_p(FWE-corr) | TFCE_ | TFCE_equivk | TFCE_ | peak_p(FWE-corr) | peak_p(FDR-corr) | peak_Z | peak_p(unc) | _x | y | z {mm} |
| --- | --- | --- | --- | --- | --- | --- | --- | --- | --- | --- |
| 0.036 | 187 |  |  | 0.071 | 0.128 | 4.226 | 0.001 | -4 | -52 | -12 |
|  |  |  |  | 0.445 | 0.128 | 3.635 | 0.001 | 6 | -44 | -16 |
|  |  |  |  | 0.483 | 0.142 | 3.582 | 0.002 | 10 | -38 | -20 |
| 0.029 | 742 |  |  | 0.13 | 0.128 | 4.049 | 0.001 | -26 | -80 | -44 |
|  |  |  |  | 0.154 | 0.128 | 4.002 | 0.001 | -24 | -60 | -36 |
|  |  |  |  | 0.254 | 0.128 | 3.85 | 0.001 | -20 | -72 | -30 |
| 0.037 | 124 |  |  | 0.343 | 0.128 | 3.74 | 0.001 | -38 | -54 | -44 |
|  |  |  |  | 0.566 | 0.128 | 3.481 | 0.001 | -36 | -50 | -36 |
| 0.05 | 5 |  |  | 0.956 | 0.15 | 2.868 | 0.003 | -16 | -70 | -44 |
