## Supplementary material for "A prebiotic diet changes neural correlates of food decision-making in overweight adults: a randomized controlled within-subject cross-over trial": SI_microbiome

**Methods**

**Feces parameters**

Fecal samples were collected independently at home, where they were frozen and stored at -15 to -20°C. Samples were delivered to the institute in isolated boxes until they were stored at -80°C. Samples were aliquoted in frozen state. For microbial measurements, DNA was extracted and further analysis was done by GENEWIZ Germany GmbH, Leipzig, Germany. For metabolomics (SCFA measurements), samples were analyzed by the Center for Environmental Research (UfZ), Leipzig, Germany.

**SCFA measurements in blood and feces.**

*Metabolite extraction:*

Chemicals: Acetonitrile, formic acid and methanol were purchased from Sigma Aldrich (St Louis, MO, USA). D7-butyric acid was purchased from Cambridge Isotope Laboratories (Tewksbury, MA, USA). All short chain fatty acids standards (SCFAs) used for linear regression and quantitation were purchased from Sigma Aldrich (St Louis, MO, USA). All solvents for MS were of analytical grade purity. Experimental water (resistivity of 18.2 MΩ cm) was purified using a Milli‐Q system (Millipore, Milford, MA, USA).

For SCFAs the method of Han et al. (2015) was modified. First, 100 mg feces were mixed with 500 µl ACN:Water (1:1, v/v) and homogenized using a TissueLyser II (30 Hz, 10 min; Retsch Qiagen). After short centrifugation (2 min, 14000 rpm) 100 µl of the supernatant were added to 500 µl ACN:Water:methanol (3:1:2, v/v/v) and the sample was vortexed for 5 min. After sonication (5 min) and centrifugation (14.000 rpm, 4°C, 5 min) 550 µl of the supernatant were transferred into a new tube and evaporated to dryness. Pellet was reconstituted in 100 µl 50% and 38 µl used for further derivatization. Next, 20 µl serum and 2 µl of standards were diluted with 18 µl and 38 µl 100% ACN, respectively. For derivatization, both specimen, serum and feces supernatant, were combined with 2 µl D7-butyric acid (2 mM) used as internal standard, 20 µl 3-nitrophenylhydrazine in 50% ACN (200 mM) and 20 µl N-(3-dimethylaminopropyl)-N‘-ethylcarbodiimide hydrochloride in 50% ACN with 6% pyridine (120 mM). Incubation of the mixture was done for 30 min at 40 °C in a thermomixer (Eppendorf, Hamburg, Germany).

Prior to measurement, the resulting derivative was diluted 1:50 in 10% ACN. Of each sample 10 µl were injected into the UltiMate 3000 HPLC system (ThermoFisher Scientific™, Waltham, MA, USA) coupled online to a QTRAP® 5500 mass spectrometer (Sciex, Framingham, USA). Chromatographic separation of SCFAs was performed on an Acquity UPLC BEH C18 column (1.7µm, 2.1 x 100 mm) with H2O + 0.01 % formic acid and ACN + 0.01% formic acid as mobile phases. Constant flow rate was set to 0.35 ml and linear LC gradient was as follows: 0-2 min at 15% B, 2-17 min 15-50% B, 17-18 min 100 % B, 18-18.1 min 100-15% B, 18.1 -21 min 15 % B. Mass spectrometric measurement was performed in negative ionization mode. For identification and quantitation, a scheduled multiple reaction monitoring (MRM) method was used, with specific transitions for every SCFA. Peak areas of all samples and standards for linear regression were determined in Analyst® Software (v. 1.6.2, AB Sciex) and areas for single SCFAs were exported. Normalization and statistics were performed with in-house written R scripts.

Note that fecal concentrations were higher compared to serum concentrations, and that the number of available samples dropped considerably due to methodological challenges in SCFA measurements, including lack of remaining blood and feces material after the preceding analyses.

**16S rRNA sequencing.**

Sample preparation and sequencing was performed by GENEWIZ (GENEWIZ Germany GmbH, Leipzig) for sequencing. Briefly, following GENEWIZ standardized workflow. For each sample, paired-end reads were joined, low-quality reads were removed, reads were corrected, chimeras removed and Amplicon Sequence Variants (ASVs) were obtained. Taxonomy was annotated to the ASVs using the RDP database 95. The read counts per ASV with taxonomic annotation were normalized and relative abundances of each ASV and taxa were calculated using the R scripts Rhea. Visualization of all library-indexed genera was done as in 96 by inhouse written R-tools using ggplot2. Group statistics (4 groups: intervention, timepoint) consists of paired ANOVA Benjamini-Hochberg adjusted with pairwise post hoc Games-Howell.

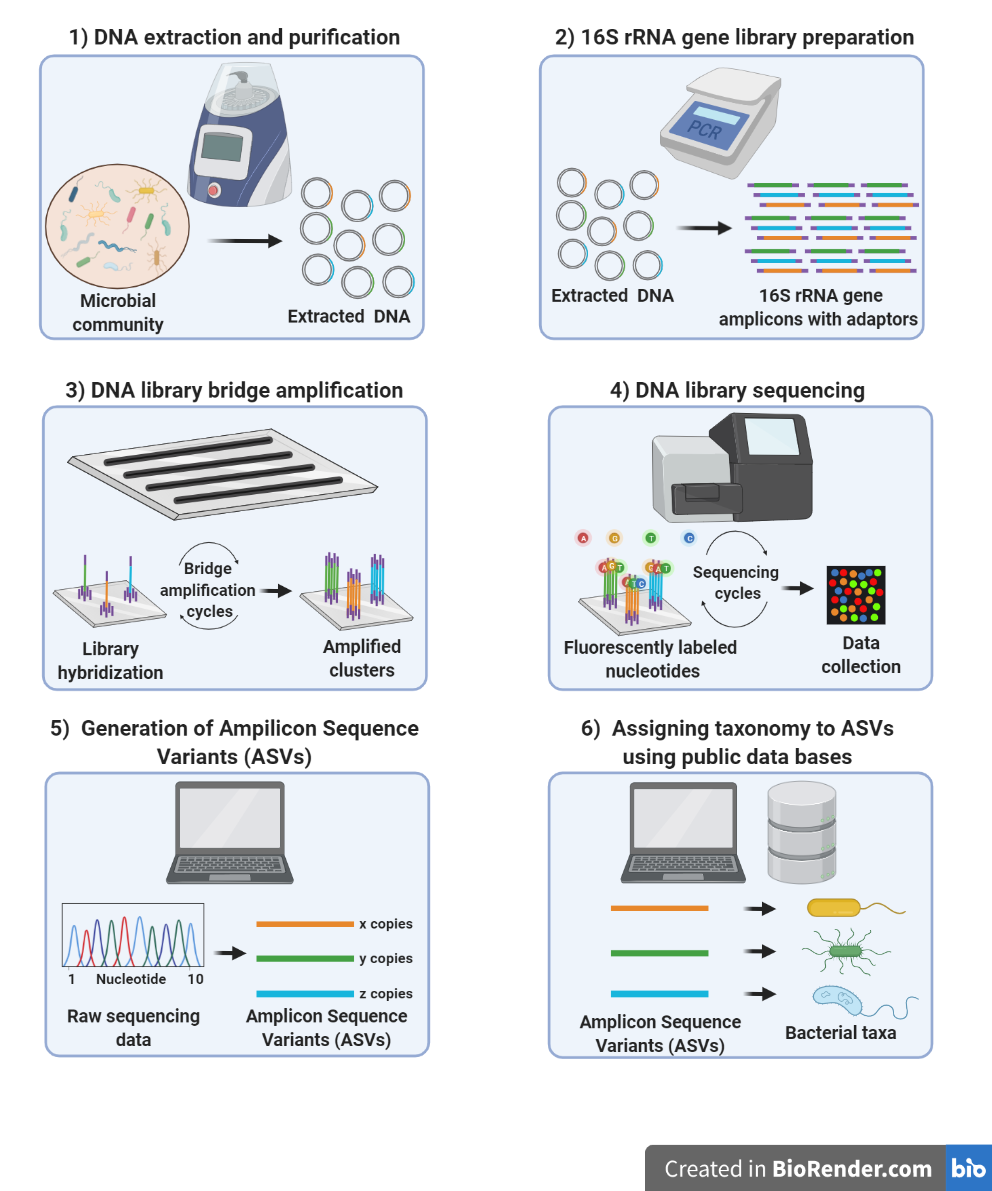

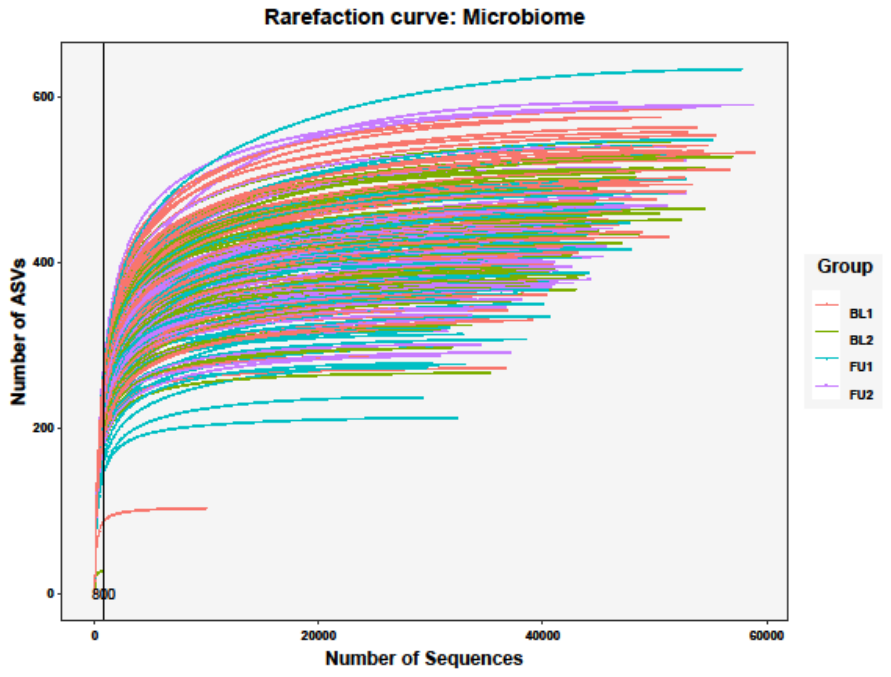

**SI Figure 1:** 16S-rRNA sequencing to ASV extraction.

**Results – Microbiotal outcome measures after intervention.**

**SI Table 1:** Significant shifts in gut-related variables of interest after prebiotic intervention, according to linear effect modelling.

|  | **intervention*timepoint effects** | | | | **ANOVA null model comparison** |
| --- | --- | --- | --- | --- | --- |
|  | **n_subj_** | **n_obs_** | **b** | **t** | **p** |
| Stool frequency | 59 | 201 | **1.26*** | **2.0*** | **0.045** |
| Bristol Stool Scale | 57 | 196 | -0.27 | -0.87 | 0.38 |
| Richness | 58 | 200 | **-53.05*** | **-3.13** | **0.0017** |
| Evenness | 58 | 200 | **-0.0084*** | **-4.93** | **<0.001** |
| Linear mixed modelling outcome compared to null model and model of interest as follows (ANOVA model comparison with p < 0.05): with the Formula: variable_of_interest ~ timepoint * intervention + timepoint + intervention + (1 \| subject) + age + gender. | | | | | |
| Shannon Effective | 58 | 200 | **-35.27*** | **-4.75** | **<0.001** |
| Simpson Effective | 58 | 200 | **-29.03*** | **-5.28** | **<0.001** |
| Linear mixed modelling outcome compared to null model and model of interest as follows (ANOVA model comparison with p < 0.05): with the Formula: variable_of_interest ~ timepoint * intervention + timepoint + intervention + (1 \| subject) + age + gender + time_of_day. | | | | | |

**Exploratory weighted network analysis (WGNCA)**

Using weighted network analysis we clustered microbiota genera to modules. In detail, data from participants with complete measures from all four timepoints (n_subj_ = 35) entered these network analyses and 4 out of 13 taxa modules were significantly correlated to prebiotic intervention (M05 r = 0.51, p < 0.001; M06 r = -0.23, p = 0.006; M08 r = -0.22, p = 0.007; M09 r = -0.20, p = 0.018). However, none of those 4 clusters correlated with prebiotic-induced changes in brain activation during decision-making. Similarly, neither hubs nor clusters of microbiota abundance differences before compared to after prebiotic intervention, nor hubs nor clusters of the microbiota pattern after prebiotic per se, correlated significantly with brain activation.

**Results – Network analysis.**

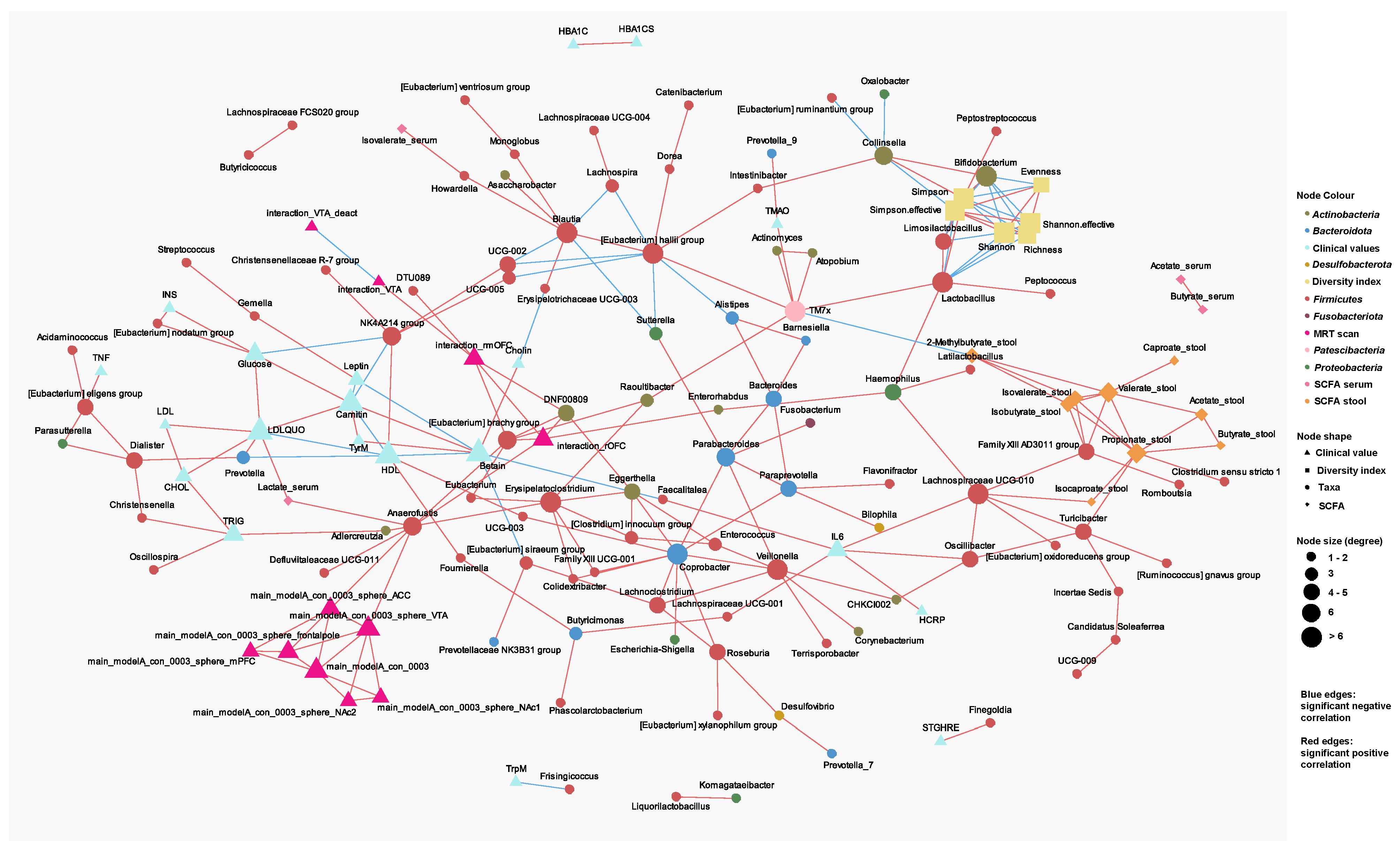

**SI Figure 2:** Network analysis.

**KEGG analysis**

We conducted functional capacity prediction of 16S-rRNA gene profiling data using the Tax4fun R-package and the Kyoto Encyclopedia of Genes and Genomes (KEGG) [2].This resulted in 8800 KEGG functional orthologues.

**SI Table 2:** Significant shifts in functional pathway capacity after prebiotic intervention, according to KEGG analysis and linear effect modelling.

| **KEGG pathway** | **intervention*timepoint effects** | | **ANOVA null model comparison** |
| --- | --- | --- | --- |
| **increased post-prebiotic intervention:** | **b** | **t** | **p** |
| ABC transporters (ko02010) | 0.55 | 2.63 | 0.009 |
| Acarbose and validamycin biosynthesis (ko00525) | 0.01 | 3.04 | 0.002 |
| Alanine aspartate and glutamate metabolism (ko00250) | 0.12 | 5.29 | < 0.001 |
| Aminoacyl tRNA biosynthesis (ko00970) | 0.11 | 2.62 | 0.009 |
| Arginine biosynthesis (ko00220) | 0.10 | 4.16 | < 0.001 |
| Carbapenem biosynthesis (ko00332) | 0.02 | 4.01 | < 0.001 |
| Cyanoamino acid metabolism (ko00460) | 0.03 | 2.38 | 0.017 |
| Cysteine and methionine metabolism (ko00270) | 0.25 | 5.16 | < 0.001 |
| D Glutamine and D glutamate metabolism (ko00471) | 0.01 | 2.62 | 0.009 |
| DNA replication (ko03030) | 0.06 | 2.58 | 0.010 |
| Ferroptosis (ko04216) | 0.03 | 3.74 | < 0.001 |
| Galactose metabolism (ko00052) | 0.09 | 2.42 | 0.015 |
| Glucosinolate biosynthesis (ko00966) | 0.00 | 2.34 | 0.019 |
| Glycine serine and threonine metabolism (ko00260) | 0.09 | 5.16 | < 0.001 |
| Homologous recombination (ko03440) | 0.11 | 3.98 | < 0.001 |
| Isoquinoline alkaloid biosynthesis (ko00950) | 0.05 | 5.25 | < 0.001 |
| Lysine biosynthesis (ko00300) | 0.06 | 2.20 | 0.027 |
| Mismatch repair (ko03430) | 0.06 | 2.95 | 0.003 |
| Nicotinate and nicotinamide metabolism (ko00760) | 0.08 | 6.35 | < 0.001 |
| Nucleotide excision repair (ko03420) | 0.08 | 3.89 | < 0.001 |
| Phenylalanine tyrosine and tryptophan biosynthesis (ko00400) | 0.10 | 2.87 | 0.004 |
| Phenylpropanoid biosynthesis (ko00940) | 0.03 | 2.60 | 0.009 |
| Polyketide sugar unit biosynthesis (ko00523) | 0.02 | 2.09 | 0.036 |
| Primary bile acid biosynthesis (ko00120) | 0.01 | 3.35 | 0.001 |
| Proteasome (ko03050) | 0.01 | 3.81 | < 0.001 |
| Purine metabolism (ko00230) | 0.19 | 4.08 | < 0.001 |
| Pyrimidine metabolism (ko00240) | 0.12 | 3.18 | 0.002 |
| Quorum sensing (ko02024) | 0.19 | 2.58 | 0.010 |
| Ribosome (ko03010) | 0.28 | 3.04 | 0.002 |
| RNA degradation (ko03018) | 0.04 | 2.80 | 0.005 |
| RNA polymerase (ko03020) | 0.03 | 3.23 | 0.001 |
| Secondary bile acid biosynthesis (ko00121) | 0.01 | 3.05 | 0.002 |
| Selenocompound metabolism (ko00450) | 0.08 | 5.98 | < 0.001 |
| Starch and sucrose metabolism (ko00500) | 0.18 | 2.74 | 0.006 |
| Taurine and hypotaurine metabolism (ko00430) | 0.03 | 6.53 | < 0.001 |
| Terpenoid backbone biosynthesis (ko00900) | 0.04 | 2.62 | 0.009 |
| Thiamine metabolism (ko00730) | 0.05 | 3.21 | 0.001 |
| Tropane piperidine and pyridine alkaloid biosynthesis (ko00960) | 0.04 | 4.70 | < 0.001 |
| Valine leucine and isoleucine biosynthesis (ko00290) | 0.04 | 2.64 | 0.008 |
| Vitamin B6 metabolism (ko00750) | 0.02 | 4.91 | < 0.001 |
| Zeatin biosynthesis (ko00908) | 0.00 | 2.25 | 0.024 |
| **decreased post-prebiotic intervention:** | **b** | **t** | **p** |
| Amino sugar and nucleotide sugar metabolism (ko00520) | -0.07 | -2.49 | 0.013 |
| Arachidonic acid metabolism (ko00590) | -0.01 | -2.90 | 0.004 |
| Atrazine degradation (ko00791) | -0.01 | -3.34 | 0.001 |
| Basal transcription factors (ko03022) | 0.00 | -3.36 | 0.001 |
| beta Alanine metabolism (ko00410) | -0.04 | -2.05 | 0.039 |
| Biofilm formation - Pseudomonas aeruginosa (ko02025) | -0.09 | -2.73 | 0.007 |
| Biosynthesis of siderophore group nonribosomal peptides (ko01053) | -0.04 | -2.27 | 0.023 |
| Biosynthesis of terpenoids and steroids (ko01062) | 0.00 | -3.56 | < 0.001 |
| Biosynthesis of type II polyketide products (ko01057) | 0.00 | -2.03 | 0.043 |
| Biosynthesis of unsaturated fatty acids (ko01040) | -0.03 | -2.62 | 0.009 |
| Biotin metabolism (ko00780) | -0.08 | -3.45 | 0.001 |
| Caprolactam degradation (ko00930) | -0.01 | -2.22 | 0.026 |
| Carotenoid biosynthesis (ko00906) | 0.00 | -3.05 | 0.002 |
| Cell cycle - Caulobacter (ko04112) | -0.06 | -2.10 | 0.035 |
| Citrate cycle - TCA cycle (ko00020) | -0.05 | -4.39 | < 0.001 |
| Fatty acid biosynthesis (ko00061) | -0.05 | -3.25 | 0.001 |
| Flavonoid biosynthesis (ko00941) | 0.00 | -2.29 | 0.022 |
| Fluorobenzoate degradation (ko00364) | -0.01 | -2.11 | 0.035 |
| Fructose and mannose metabolism (ko00051) | -0.16 | -2.48 | 0.013 |
| Glycerophospholipid metabolism (ko00564) | -0.02 | -2.00 | 0.045 |
| Glycolysis - Gluconeogenesis (ko00010) | -0.08 | -2.42 | 0.015 |
| Indole alkaloid biosynthesis (ko00901) | 0.00 | -2.47 | 0.014 |
| Inositol phosphate metabolism (ko00562) | -0.02 | -3.59 | < 0.001 |
| Limonene and pinene degradation (ko00903) | -0.02 | -1.99 | 0.046 |
| Lysine degradation (ko00310) | -0.06 | -2.06 | 0.039 |
| Methane metabolism (ko00680) | -0.06 | -4.09 | < 0.001 |
| Nitrotoluene degradation (ko00633) | -0.02 | -2.90 | 0.004 |
| Non homologous end joining (ko03450) | 0.00 | -3.47 | 0.001 |
| Nonribosomal peptide structures (ko01054) | -0.01 | -2.11 | 0.034 |
| Oxidative phosphorylation (ko00190) | -0.10 | -4.68 | < 0.001 |
| Phosphonate and phosphinate metabolism (ko00440) | -0.01 | -2.19 | 0.028 |
| Polycyclic aromatic hydrocarbon degradation (ko00624) | 0.00 | -2.51 | 0.012 |
| Porphyrin and chlorophyll metabolism (ko00860) | -0.16 | -6.77 | < 0.001 |
| Pyruvate metabolism (ko00620) | -0.10 | -4.53 | < 0.001 |
| RNA transport (ko03013) | -0.02 | -5.80 | < 0.001 |
| Steroid degradation (ko00984) | -0.01 | -3.48 | 0.001 |
| Stilbenoid. diarylheptanoid and gingerol biosynthesis (ko00945) | 0.00 | -2.29 | 0.022 |
| Styrene degradation (ko00643) | -0.01 | -2.94 | 0.003 |
| Sulfur metabolism (ko00920) | -0.09 | -2.32 | 0.020 |
| Sulfur relay system (ko04122) | -0.04 | -3.85 | < 0.001 |
| Tryptophan metabolism (ko00380) | -0.04 | -2.12 | 0.034 |
| Two component system (ko02020) | -1.01 | -5.43 | < 0.001 |
| Xylene degradation (ko00622) | -0.02 | -2.99 | 0.003 |

Linear mixed modelling outcome compared to null model and model of interest as follows (ANOVA model comparison with p < 0.05): with the Formula: pathyway_of_interest ~ timepoint * intervention + timepoint + intervention + (1 | subject). All models on n = 205 observations in n = 58 individuals and listed in alphabetical order of genera of interest.

**References.**

1	Han J, Lin K, Sequeira C, *et al.* An isotope-labeled chemical derivatization method for the quantitation of short-chain fatty acids in human feces by liquid chromatography–tandem mass spectrometry. *Anal Chim Acta* 2015;**854**:86–94.

2	Ogata H, Goto S, Fujibuchi W, *et al.* Computation with the KEGG pathway database. *BioSystems* 1998;**47**:119–28. doi:10.1016/S0303-2647(98)00017-3
