## Supplementary material for "A prebiotic diet changes neural correlates of food decision-making in overweight adults: a randomized controlled within-subject cross-over trial": SI_microbiome_Table3

### Correlations

| | | | interaction_VTA_change | interaction_rOFC_change | interaction_rmOFC_change | sumscore_GIQLI_change | Stool_frequency_change | STGHRE_change |
|---|---|---|---|---|---|---|---|---|
| Spearman's rho | interaction_VTA_change | Correlation Coefficient | 1,000 | ,344 | ,414 | -,162 | -,159 | ,043 |
| | | Sig. (2-tailed) | . | ,028 | ,005 | ,299 | ,316 | ,793 |
| | | N | 44 | 41 | 44 | 43 | 42 | 40 |
| | interaction_rOFC_change | Correlation Coefficient | ,344 | 1,000 | ,410 | -,295 | ,014 | ,070 |
| | | Sig. (2-tailed) | ,028 | . | ,008 | ,065 | ,934 | ,681 |
| | | N | 41 | 41 | 41 | 40 | 39 | 37 |
| | interaction_rmOFC_change | Correlation Coefficient | ,414 | ,410 | 1,000 | ,010 | ,027 | ,253 |
| | | Sig. (2-tailed) | ,005 | ,008 | . | ,947 | ,867 | ,116 |
| | | N | 44 | 41 | 44 | 43 | 42 | 40 |
| | sumscore_GIQLI_change | Correlation Coefficient | -,162 | -,295 | ,010 | 1,000 | ,063 | ,054 |
| | | Sig. (2-tailed) | ,299 | ,065 | ,947 | . | ,683 | ,738 |
| | | N | 43 | 40 | 43 | 45 | 44 | 41 |
| | Stool_frequency_change | Correlation Coefficient | -,159 | ,014 | ,027 | ,063 | 1,000 | -,082 |
| | | Sig. (2-tailed) | ,316 | ,934 | ,867 | ,683 | . | ,615 |
| | | N | 42 | 39 | 42 | 44 | 44 | 40 |
| | STGHRE_change | Correlation Coefficient | ,043 | ,070 | ,253 | ,054 | -,082 | 1,000 |
| | | Sig. (2-tailed) | ,793 | ,681 | ,116 | ,738 | ,615 | . |
| | | N | 40 | 37 | 40 | 41 | 40 | 42 |
| | STPYY_change | Correlation Coefficient | ,439 | ,348 | ,318 | -,159 | -,047 | ,011 |
| | | Sig. (2-tailed) | ,005 | ,035 | ,046 | ,321 | ,774 | ,947 |
| | | N | 40 | 37 | 40 | 41 | 40 | 42 |
| | STGLPT_change | Correlation Coefficient | ,057 | ,192 | -,012 | ,117 | ,011 | -,193 |
| | | Sig. (2-tailed) | ,727 | ,254 | ,943 | ,468 | ,946 | ,221 |
| | | N | 40 | 37 | 40 | 41 | 40 | 42 |

### Correlations

| | | | STPYY_change | STGLPT_change | age | BMI | FM_stand | Richness | shannon. effective |
|---|---|---|---|---|---|---|---|---|---|
| Spearman's rho | interaction_VTA_change | Correlation Coefficient | ,439 | ,057 | -,028 | -,251 | ,104 | ,182 | ,160 |
| | | Sig. (2-tailed) | ,005 | ,727 | ,854 | ,100 | ,500 | ,237 | ,299 |
| | | N | 40 | 40 | 44 | 44 | 44 | 44 | 44 |
| | interaction_rOFC_change | Correlation Coefficient | ,348 | ,192 | ,385 | -,273 | -,088 | ,041 | ,020 |
| | | Sig. (2-tailed) | ,035 | ,254 | ,013 | ,084 | ,585 | ,797 | ,901 |
| | | N | 37 | 37 | 41 | 41 | 41 | 41 | 41 |
| | interaction_rmOFC_change | Correlation Coefficient | ,318 | -,012 | ,167 | -,174 | -,031 | -,116 | -,125 |
| | | Sig. (2-tailed) | ,046 | ,943 | ,279 | ,259 | ,839 | ,453 | ,418 |
| | | N | 40 | 40 | 44 | 44 | 44 | 44 | 44 |
| | sumscore_GIQLI_change | Correlation Coefficient | -,159 | ,117 | ,175 | ,155 | ,154 | ,181 | ,181 |
| | | Sig. (2-tailed) | ,321 | ,468 | ,251 | ,309 | ,312 | ,235 | ,234 |
| | | N | 41 | 41 | 45 | 45 | 45 | 45 | 45 |
| | Stool_frequency_change | Correlation Coefficient | -,047 | ,011 | -,127 | -,034 | -,242 | -,278 | -,259 |
| | | Sig. (2-tailed) | ,774 | ,946 | ,410 | ,829 | ,114 | ,067 | ,090 |
| | | N | 40 | 40 | 44 | 44 | 44 | 44 | 44 |
| | STGHRE_change | Correlation Coefficient | ,011 | -,193 | -,051 | -,174 | -,351 | -,072 | -,072 |
| | | Sig. (2-tailed) | ,947 | ,221 | ,748 | ,270 | ,023 | ,652 | ,652 |
| | | N | 42 | 42 | 42 | 42 | 42 | 42 | 42 |
| | STPYY_change | Correlation Coefficient | 1,000 | ,113 | -,020 | -,078 | ,124 | ,137 | ,141 |
| | | Sig. (2-tailed) | . | ,478 | ,902 | ,625 | ,433 | ,388 | ,374 |
| | | N | 42 | 42 | 42 | 42 | 42 | 42 | 42 |
| | STGLPT_change | Correlation Coefficient | ,113 | 1,000 | -,094 | ,131 | ,150 | ,127 | ,044 |
| | | Sig. (2-tailed) | ,478 | . | ,556 | ,408 | ,344 | ,423 | ,780 |
| | | N | 42 | 42 | 42 | 42 | 42 | 42 | 42 |

### Correlations

| | | | simpson. effective | Evenness | CHOL_change | LDL_change | shannon. effective_change | Richness_change |
|---|---|---|---|---|---|---|---|---|
| Spearman's rho | interaction_VTA_change | Correlation Coefficient | ,081 | -,185 | -,066 | -,080 | ,241 | ,154 |
| | | Sig. (2-tailed) | ,603 | ,229 | ,669 | ,607 | ,120 | ,323 |
| | | N | 44 | 44 | 44 | 44 | 43 | 43 |
| | interaction_rOFC_change | Correlation Coefficient | -,039 | -,136 | ,041 | ,057 | -,253 | -,211 |
| | | Sig. (2-tailed) | ,811 | ,398 | ,800 | ,721 | ,115 | ,191 |
| | | N | 41 | 41 | 41 | 41 | 40 | 40 |
| | interaction_rmOFC_change | Correlation Coefficient | -,130 | -,124 | -,086 | -,199 | -,154 | -,177 |
| | | Sig. (2-tailed) | ,400 | ,422 | ,578 | ,196 | ,324 | ,256 |
| | | N | 44 | 44 | 44 | 44 | 43 | 43 |
| | sumscore_GIQLI_change | Correlation Coefficient | ,253 | ,071 | -,157 | -,079 | ,154 | ,122 |
| | | Sig. (2-tailed) | ,094 | ,644 | ,304 | ,608 | ,319 | ,431 |
| | | N | 45 | 45 | 45 | 45 | 44 | 44 |
| | Stool_frequency_change | Correlation Coefficient | -,216 | ,040 | -,200 | -,075 | -,270 | -,326 |
| | | Sig. (2-tailed) | ,159 | ,797 | ,194 | ,631 | ,080 | ,033 |
| | | N | 44 | 44 | 44 | 44 | 43 | 43 |
| | STGHRE_change | Correlation Coefficient | -,017 | -,022 | -,506 | -,532 | ,007 | -,070 |
| | | Sig. (2-tailed) | ,914 | ,889 | ,001 | ,000 | ,964 | ,659 |
| | | N | 42 | 42 | 42 | 42 | 42 | 42 |
| | STPYY_change | Correlation Coefficient | ,090 | -,060 | ,083 | ,080 | -,089 | -,204 |
| | | Sig. (2-tailed) | ,570 | ,706 | ,600 | ,615 | ,575 | ,196 |
| | | N | 42 | 42 | 42 | 42 | 42 | 42 |
| | STGLPT_change | Correlation Coefficient | -,017 | -,131 | ,230 | ,233 | ,014 | -,014 |
| | | Sig. (2-tailed) | ,917 | ,408 | ,143 | ,137 | ,928 | ,931 |
| | | N | 42 | 42 | 42 | 42 | 42 | 42 |

### Correlations

| | | | Evenness_change | Actinomyces_change | Aerostipes_change | Bifidobacterium_change | Blautia_change | Collinsella_change |
|---|---|---|---|---|---|---|---|---|
| Spearman's rho | interaction_VTA_change | Correlation Coefficient | ,112 | -,042 | -,264 | -,152 | -,238 | ,024 |
| | | Sig. (2-tailed) | ,473 | ,791 | ,088 | ,332 | ,124 | ,878 |
| | | N | 43 | 43 | 43 | 43 | 43 | 43 |
| | interaction_rOFC_change | Correlation Coefficient | -,227 | ,268 | -,206 | -,136 | ,179 | ,178 |
| | | Sig. (2-tailed) | ,159 | ,094 | ,201 | ,401 | ,270 | ,271 |
| | | N | 40 | 40 | 40 | 40 | 40 | 40 |
| | interaction_rmOFC_change | Correlation Coefficient | -,045 | ,173 | ,039 | ,098 | ,084 | ,150 |
| | | Sig. (2-tailed) | ,772 | ,268 | ,806 | ,533 | ,594 | ,339 |
| | | N | 43 | 43 | 43 | 43 | 43 | 43 |
| | sumscore_GIQLI_change | Correlation Coefficient | ,171 | -,130 | ,021 | -,109 | ,166 | -,202 |
| | | Sig. (2-tailed) | ,266 | ,401 | ,892 | ,482 | ,283 | ,189 |
| | | N | 44 | 44 | 44 | 44 | 44 | 44 |
| | Stool_frequency_change | Correlation Coefficient | ,106 | -,006 | ,325 | -,125 | ,115 | ,151 |
| | | Sig. (2-tailed) | ,498 | ,972 | ,034 | ,425 | ,465 | ,333 |
| | | N | 43 | 43 | 43 | 43 | 43 | 43 |
| | STGHRE_change | Correlation Coefficient | ,124 | ,008 | -,040 | -,225 | ,163 | ,014 |
| | | Sig. (2-tailed) | ,433 | ,958 | ,803 | ,151 | ,301 | ,930 |
| | | N | 42 | 42 | 42 | 42 | 42 | 42 |
| | STPYY_change | Correlation Coefficient | ,122 | ,084 | ,001 | ,012 | -,021 | ,176 |
| | | Sig. (2-tailed) | ,442 | ,599 | ,995 | ,940 | ,894 | ,266 |
| | | N | 42 | 42 | 42 | 42 | 42 | 42 |
| | STGLPT_change | Correlation Coefficient | ,073 | -,030 | ,067 | ,014 | ,209 | ,004 |
| | | Sig. (2-tailed) | ,644 | ,852 | ,673 | ,929 | ,184 | ,978 |
| | | N | 42 | 42 | 42 | 42 | 42 | 42 |

### Correlations

| | | | Desulfovibrio_change | Eggerthella_change | Erysipelatoclostridium_change | Faecalitalea_change | Family.XIII. AD3011. group_change | Family.XIII. UCG. 001_change |
|---|---|---|---|---|---|---|---|---|
| Spearman's rho | interaction_VTA_change | Correlation Coefficient | ,034 | ,020 | ,159 | -,099 | ,121 | -,080 |
| | | Sig. (2-tailed) | ,830 | ,899 | ,309 | ,526 | ,440 | ,611 |
| | | N | 43 | 43 | 43 | 43 | 43 | 43 |
| | interaction_rOFC_change | Correlation Coefficient | -,009 | ,022 | -,021 | -,018 | ,191 | ,064 |
| | | Sig. (2-tailed) | ,955 | ,892 | ,900 | ,911 | ,238 | ,696 |
| | | N | 40 | 40 | 40 | 40 | 40 | 40 |
| | interaction_rmOFC_change | Correlation Coefficient | -,084 | -,027 | -,063 | ,035 | ,070 | -,206 |
| | | Sig. (2-tailed) | ,591 | ,862 | ,688 | ,825 | ,655 | ,185 |
| | | N | 43 | 43 | 43 | 43 | 43 | 43 |
| | sumscore_GIQLI_change | Correlation Coefficient | ,059 | ,104 | ,190 | ,038 | ,180 | ,009 |
| | | Sig. (2-tailed) | ,704 | ,503 | ,216 | ,809 | ,241 | ,953 |
| | | N | 44 | 44 | 44 | 44 | 44 | 44 |
| | Stool_frequency_change | Correlation Coefficient | ,066 | -,131 | -,291 | -,040 | ,250 | -,054 |
| | | Sig. (2-tailed) | ,674 | ,403 | ,058 | ,798 | ,105 | ,731 |
| | | N | 43 | 43 | 43 | 43 | 43 | 43 |
| | STGHRE_change | Correlation Coefficient | -,095 | -,126 | -,075 | ,166 | ,125 | ,215 |
| | | Sig. (2-tailed) | ,551 | ,425 | ,639 | ,294 | ,430 | ,171 |
| | | N | 42 | 42 | 42 | 42 | 42 | 42 |
| | STPYY_change | Correlation Coefficient | -,370 | ,032 | -,017 | ,011 | ,171 | ,017 |
| | | Sig. (2-tailed) | ,016 | ,839 | ,913 | ,945 | ,279 | ,913 |
| | | N | 42 | 42 | 42 | 42 | 42 | 42 |
| | STGLPT_change | Correlation Coefficient | -,069 | ,103 | ,296 | ,090 | ,058 | ,205 |
| | | Sig. (2-tailed) | ,666 | ,518 | ,057 | ,570 | ,717 | ,192 |
| | | N | 42 | 42 | 42 | 42 | 42 | 42 |

### Correlations

| | | | Gemella_change | Gordonibacter_change | Holdemanella_change | Holdemania_change | Lachnospiraceae.FCS020.group_change | Lachnospiraceae.NK4A136.group_change |
|---|---|---|---|---|---|---|---|---|
| Spearman's rho | interaction_VTA_change | Correlation Coefficient | -,174 | ,025 | -,083 | -,116 | -,041 | ,286 |
| | | Sig. (2-tailed) | ,264 | ,876 | ,597 | ,459 | ,792 | ,063 |
| | | N | 43 | 43 | 43 | 43 | 43 | 43 |
| | interaction_rOFC_change | Correlation Coefficient | ,295 | ,044 | ,049 | -,259 | -,079 | -,093 |
| | | Sig. (2-tailed) | ,064 | ,785 | ,764 | ,107 | ,627 | ,566 |
| | | N | 40 | 40 | 40 | 40 | 40 | 40 |
| | interaction_rmOFC_change | Correlation Coefficient | ,196 | ,079 | ,005 | -,195 | ,028 | -,001 |
| | | Sig. (2-tailed) | ,209 | ,614 | ,972 | ,211 | ,860 | ,996 |
| | | N | 43 | 43 | 43 | 43 | 43 | 43 |
| | sumscore_GIQLI_change | Correlation Coefficient | ,086 | ,079 | ,160 | ,095 | ,133 | ,000 |
| | | Sig. (2-tailed) | ,577 | ,610 | ,301 | ,539 | ,390 | ,998 |
| | | N | 44 | 44 | 44 | 44 | 44 | 44 |
| | Stool_frequency_change | Correlation Coefficient | -,119 | -,099 | -,104 | -,016 | -,071 | -,013 |
| | | Sig. (2-tailed) | ,445 | ,526 | ,505 | ,919 | ,649 | ,933 |
| | | N | 43 | 43 | 43 | 43 | 43 | 43 |
| | STGHRE_change | Correlation Coefficient | -,105 | -,040 | -,104 | ,015 | -,078 | ,094 |
| | | Sig. (2-tailed) | ,510 | ,803 | ,514 | ,927 | ,624 | ,554 |
| | | N | 42 | 42 | 42 | 42 | 42 | 42 |
| | STPYY_change | Correlation Coefficient | -,139 | -,341 | -,043 | -,157 | ,076 | ,155 |
| | | Sig. (2-tailed) | ,381 | ,027 | ,785 | ,321 | ,633 | ,326 |
| | | N | 42 | 42 | 42 | 42 | 42 | 42 |
| | STGLPT_change | Correlation Coefficient | ,150 | ,132 | ,166 | -,266 | ,162 | -,079 |
| | | Sig. (2-tailed) | ,342 | ,404 | ,294 | ,089 | ,307 | ,621 |
| | | N | 42 | 42 | 42 | 42 | 42 | 42 |

### Correlations

| | | | Lactiplantibacillus_change | Lactobacillus_change | Libanicoccus_change | Ligilactobacillus_change | Limosilactobacillus_change | Roseburia_change |
|---|---|---|---|---|---|---|---|---|
| Spearman's rho | interaction_VTA_change | Correlation Coefficient | -,006 | -,023 | ,294 | -,175 | -,010 | ,137 |
| | | Sig. (2-tailed) | ,972 | ,882 | ,056 | ,262 | ,949 | ,380 |
| | | N | 43 | 43 | 43 | 43 | 43 | 43 |
| | interaction_rOFC_change | Correlation Coefficient | ,120 | ,046 | ,211 | -,105 | -,005 | ,146 |
| | | Sig. (2-tailed) | ,459 | ,778 | ,192 | ,521 | ,976 | ,369 |
| | | N | 40 | 40 | 40 | 40 | 40 | 40 |
| | interaction_rmOFC_change | Correlation Coefficient | ,397 | -,054 | ,223 | -,082 | -,070 | -,011 |
| | | Sig. (2-tailed) | ,008 | ,731 | ,151 | ,603 | ,654 | ,944 |
| | | N | 43 | 43 | 43 | 43 | 43 | 43 |
| | sumscore_GIQLI_change | Correlation Coefficient | ,168 | -,033 | -,057 | ,054 | ,058 | ,025 |
| | | Sig. (2-tailed) | ,276 | ,833 | ,712 | ,727 | ,706 | ,870 |
| | | N | 44 | 44 | 44 | 44 | 44 | 44 |
| | Stool_frequency_change | Correlation Coefficient | -,007 | ,105 | -,187 | ,060 | ,169 | -,316 |
| | | Sig. (2-tailed) | ,966 | ,504 | ,230 | ,703 | ,278 | ,039 |
| | | N | 43 | 43 | 43 | 43 | 43 | 43 |
| | STGHRE_change | Correlation Coefficient | ,131 | -,070 | -,107 | -,017 | -,196 | -,209 |
| | | Sig. (2-tailed) | ,407 | ,659 | ,499 | ,914 | ,212 | ,184 |
| | | N | 42 | 42 | 42 | 42 | 42 | 42 |
| | STPYY_change | Correlation Coefficient | -,079 | ,046 | ,039 | ,015 | ,027 | -,048 |
| | | Sig. (2-tailed) | ,618 | ,771 | ,807 | ,924 | ,864 | ,763 |
| | | N | 42 | 42 | 42 | 42 | 42 | 42 |
| | STGLPT_change | Correlation Coefficient | -,186 | -,181 | ,115 | ,038 | -,182 | -,035 |
| | | Sig. (2-tailed) | ,238 | ,251 | ,468 | ,813 | ,248 | ,824 |
| | | N | 42 | 42 | 42 | 42 | 42 | 42 |

### Correlations

| | | | Shuttleworthia_change | Subdoligranulum_change | UCG.003_change | X.Eubacterium..brachy.group_change | X.Ruminococcus..gauvreauii.group_change | X.Ruminococcus..torques.group_change |
|---|---|---|---|---|---|---|---|---|
| Spearman's rho | interaction_VTA_change | Correlation Coefficient | ,191 | -,384 | ,185 | ,115 | -,194 | -,229 |
| | | Sig. (2-tailed) | ,220 | ,011 | ,236 | ,464 | ,211 | ,140 |
| | | N | 43 | 43 | 43 | 43 | 43 | 43 |
| | interaction_rOFC_change | Correlation Coefficient | -,155 | -,159 | ,122 | -,177 | -,143 | -,194 |
| | | Sig. (2-tailed) | ,338 | ,329 | ,453 | ,275 | ,377 | ,230 |
| | | N | 40 | 40 | 40 | 40 | 40 | 40 |
| | interaction_rmOFC_change | Correlation Coefficient | ,032 | -,015 | -,094 | ,038 | -,024 | -,133 |
| | | Sig. (2-tailed) | ,838 | ,922 | ,551 | ,809 | ,879 | ,397 |
| | | N | 43 | 43 | 43 | 43 | 43 | 43 |
| | sumscore_GIQLI_change | Correlation Coefficient | ,210 | ,261 | -,255 | ,090 | ,113 | ,305 |
| | | Sig. (2-tailed) | ,172 | ,087 | ,095 | ,561 | ,465 | ,044 |
| | | N | 44 | 44 | 44 | 44 | 44 | 44 |
| | Stool_frequency_change | Correlation Coefficient | ,115 | ,083 | -,067 | -,196 | -,091 | ,271 |
| | | Sig. (2-tailed) | ,462 | ,598 | ,669 | ,209 | ,560 | ,079 |
| | | N | 43 | 43 | 43 | 43 | 43 | 43 |
| | STGHRE_change | Correlation Coefficient | -,164 | ,279 | -,059 | -,097 | -,100 | ,230 |
| | | Sig. (2-tailed) | ,300 | ,073 | ,711 | ,540 | ,528 | ,144 |
| | | N | 42 | 42 | 42 | 42 | 42 | 42 |
| | STPYY_change | Correlation Coefficient | -,068 | -,210 | -,176 | -,061 | -,063 | ,183 |
| | | Sig. (2-tailed) | ,669 | ,182 | ,264 | ,702 | ,693 | ,247 |
| | | N | 42 | 42 | 42 | 42 | 42 | 42 |
| | STGLPT_change | Correlation Coefficient | -,074 | ,009 | -,165 | ,179 | -,159 | ,092 |
| | | Sig. (2-tailed) | ,644 | ,957 | ,296 | ,256 | ,314 | ,561 |
| | | N | 42 | 42 | 42 | 42 | 42 | 42 |

### Correlations

| | | interaction_VTA_change | interaction_rOFC_change | interaction_rmOFC_change | sumscore_GIQLI_change | Stool_frequency_change | STGHRE_change |
|---|---|---|---|---|---|---|---|
| age | Correlation Coefficient | -,028 | ,385 | ,167 | ,175 | -,127 | -,051 |
| | Sig. (2-tailed) | ,854 | ,013 | ,279 | ,251 | ,410 | ,748 |
| | N | 44 | 41 | 44 | 45 | 44 | 42 |
| BMI | Correlation Coefficient | -,251 | -,273 | -,174 | ,155 | -,034 | -,174 |
| | Sig. (2-tailed) | ,100 | ,084 | ,259 | ,309 | ,829 | ,270 |
| | N | 44 | 41 | 44 | 45 | 44 | 42 |
| FM_stand | Correlation Coefficient | ,104 | -,088 | -,031 | ,154 | -,242 | -,351 |
| | Sig. (2-tailed) | ,500 | ,585 | ,839 | ,312 | ,114 | ,023 |
| | N | 44 | 41 | 44 | 45 | 44 | 42 |
| Richness | Correlation Coefficient | ,182 | ,041 | -,116 | ,181 | -,278 | -,072 |
| | Sig. (2-tailed) | ,237 | ,797 | ,453 | ,235 | ,067 | ,652 |
| | N | 44 | 41 | 44 | 45 | 44 | 42 |
| shannon.effective | Correlation Coefficient | ,160 | ,020 | -,125 | ,181 | -,259 | -,072 |
| | Sig. (2-tailed) | ,299 | ,901 | ,418 | ,234 | ,090 | ,652 |
| | N | 44 | 41 | 44 | 45 | 44 | 42 |
| simpson.effective | Correlation Coefficient | ,081 | -,039 | -,130 | ,253 | -,216 | -,017 |
| | Sig. (2-tailed) | ,603 | ,811 | ,400 | ,094 | ,159 | ,914 |
| | N | 44 | 41 | 44 | 45 | 44 | 42 |
| Evenness | Correlation Coefficient | -,185 | -,136 | -,124 | ,071 | ,040 | -,022 |
| | Sig. (2-tailed) | ,229 | ,398 | ,422 | ,644 | ,797 | ,889 |
| | N | 44 | 41 | 44 | 45 | 44 | 42 |
| CHOL_change | Correlation Coefficient | -,066 | ,041 | -,086 | -,157 | -,200 | -,506 |
| | Sig. (2-tailed) | ,669 | ,800 | ,578 | ,304 | ,194 | ,001 |
| | N | 44 | 41 | 44 | 45 | 44 | 42 |

### Correlations

| | | STPYY_change | STGLPT_change | age | BMI | FM_stand | Richness | shannon.effective |
|---|---|---|---|---|---|---|---|---|
| age | Correlation Coefficient | -,020 | -,094 | 1,000 | -,041 | -,055 | ,126 | ,065 |
| | Sig. (2-tailed) | ,902 | ,556 | . | ,789 | ,718 | ,402 | ,670 |
| | N | 42 | 42 | 46 | 46 | 46 | 46 | 46 |
| BMI | Correlation Coefficient | -,078 | ,131 | -,041 | 1,000 | ,463 | ,163 | ,111 |
| | Sig. (2-tailed) | ,625 | ,408 | ,789 | . | ,001 | ,278 | ,463 |
| | N | 42 | 42 | 46 | 46 | 46 | 46 | 46 |
| FM_stand | Correlation Coefficient | ,124 | ,150 | -,055 | ,463 | 1,000 | ,030 | -,027 |
| | Sig. (2-tailed) | ,433 | ,344 | ,718 | ,001 | . | ,842 | ,856 |
| | N | 42 | 42 | 46 | 46 | 46 | 46 | 46 |
| Richness | Correlation Coefficient | ,137 | ,127 | ,126 | ,163 | ,030 | 1,000 | ,949 |
| | Sig. (2-tailed) | ,388 | ,423 | ,402 | ,278 | ,842 | . | ,000 |
| | N | 42 | 42 | 46 | 46 | 46 | 46 | 46 |
| shannon.effective | Correlation Coefficient | ,141 | ,044 | ,065 | ,111 | -,027 | ,949 | 1,000 |
| | Sig. (2-tailed) | ,374 | ,780 | ,670 | ,463 | ,856 | ,000 | . |
| | N | 42 | 42 | 46 | 46 | 46 | 46 | 46 |
| simpson.effective | Correlation Coefficient | ,090 | -,017 | ,058 | ,068 | -,071 | ,886 | ,965 |
| | Sig. (2-tailed) | ,570 | ,917 | ,702 | ,654 | ,639 | ,000 | ,000 |
| | N | 42 | 42 | 46 | 46 | 46 | 46 | 46 |
| Evenness | Correlation Coefficient | -,060 | -,131 | -,096 | -,029 | -,094 | ,153 | ,405 |
| | Sig. (2-tailed) | ,706 | ,408 | ,524 | ,850 | ,536 | ,310 | ,005 |
| | N | 42 | 42 | 46 | 46 | 46 | 46 | 46 |
| CHOL_change | Correlation Coefficient | ,083 | ,230 | ,077 | ,238 | ,513 | ,010 | -,067 |
| | Sig. (2-tailed) | ,600 | ,143 | ,611 | ,111 | ,000 | ,948 | ,658 |
| | N | 42 | 42 | 46 | 46 | 46 | 46 | 46 |

### Correlations

| | | simpson. effective | Evenness | CHOL_change | LDL_change | shannon. effective_change | Richness_change |
|---|---|---|---|---|---|---|---|
| age | Correlation Coefficient | ,058 | -,096 | ,077 | ,052 | -,019 | ,089 |
| | Sig. (2-tailed) | ,702 | ,524 | ,611 | ,733 | ,903 | ,559 |
| | N | 46 | 46 | 46 | 46 | 45 | 45 |
| BMI | Correlation Coefficient | ,068 | -,029 | ,238 | ,202 | -,130 | -,105 |
| | Sig. (2-tailed) | ,654 | ,850 | ,111 | ,179 | ,395 | ,492 |
| | N | 46 | 46 | 46 | 46 | 45 | 45 |
| FM_stand | Correlation Coefficient | -,071 | -,094 | ,513 | ,478 | ,014 | ,167 |
| | Sig. (2-tailed) | ,639 | ,536 | ,000 | ,001 | ,927 | ,274 |
| | N | 46 | 46 | 46 | 46 | 45 | 45 |
| Richness | Correlation Coefficient | ,886 | ,153 | ,010 | -,042 | ,490 | ,500 |
| | Sig. (2-tailed) | ,000 | ,310 | ,948 | ,781 | ,001 | ,000 |
| | N | 46 | 46 | 46 | 46 | 45 | 45 |
| shannon.effective | Correlation Coefficient | ,965 | ,405 | -,067 | -,119 | ,570 | ,512 |
| | Sig. (2-tailed) | ,000 | ,005 | ,658 | ,430 | ,000 | ,000 |
| | N | 46 | 46 | 46 | 46 | 45 | 45 |
| simpson.effective | Correlation Coefficient | 1,000 | ,491 | -,114 | -,149 | ,586 | ,500 |
| | Sig. (2-tailed) | . | ,001 | ,449 | ,322 | ,000 | ,000 |
| | N | 46 | 46 | 46 | 46 | 45 | 45 |
| Evenness | Correlation Coefficient | ,491 | 1,000 | -,053 | -,082 | ,347 | ,163 |
| | Sig. (2-tailed) | ,001 | . | ,726 | ,588 | ,020 | ,285 |
| | N | 46 | 46 | 46 | 46 | 45 | 45 |
| CHOL_change | Correlation Coefficient | -,114 | -,053 | 1,000 | ,926 | -,144 | ,038 |
| | Sig. (2-tailed) | ,449 | ,726 | . | ,000 | ,344 | ,806 |
| | N | 46 | 46 | 46 | 46 | 45 | 45 |

### Correlations

| | | Evenness_change | Actinomyces_change | Aerostipes_change | Bifidobacterium_change | Blautia_change | Collinsella_change |
|---|---|---|---|---|---|---|---|
| age | Correlation Coefficient | -,205 | -,004 | -,078 | -,044 | ,083 | -,228 |
| | Sig. (2-tailed) | ,177 | ,979 | ,611 | ,774 | ,588 | ,133 |
| | N | 45 | 45 | 45 | 45 | 45 | 45 |
| BMI | Correlation Coefficient | -,086 | -,121 | ,105 | ,013 | -,132 | -,035 |
| | Sig. (2-tailed) | ,575 | ,429 | ,491 | ,933 | ,387 | ,820 |
| | N | 45 | 45 | 45 | 45 | 45 | 45 |
| FM_stand | Correlation Coefficient | -,303 | ,085 | -,157 | ,323 | -,159 | ,013 |
| | Sig. (2-tailed) | ,043 | ,577 | ,305 | ,031 | ,296 | ,932 |
| | N | 45 | 45 | 45 | 45 | 45 | 45 |
| Richness | Correlation Coefficient | ,137 | -,028 | ,011 | -,514 | -,145 | -,354 |
| | Sig. (2-tailed) | ,370 | ,856 | ,943 | ,000 | ,341 | ,017 |
| | N | 45 | 45 | 45 | 45 | 45 | 45 |
| shannon.effective | Correlation Coefficient | ,306 | -,017 | ,036 | -,582 | -,055 | -,322 |
| | Sig. (2-tailed) | ,041 | ,912 | ,816 | ,000 | ,722 | ,031 |
| | N | 45 | 45 | 45 | 45 | 45 | 45 |
| simpson.effective | Correlation Coefficient | ,373 | -,085 | ,058 | -,626 | -,032 | -,369 |
| | Sig. (2-tailed) | ,012 | ,577 | ,703 | ,000 | ,833 | ,012 |
| | N | 45 | 45 | 45 | 45 | 45 | 45 |
| Evenness | Correlation Coefficient | ,600 | ,010 | ,155 | -,401 | ,355 | -,060 |
| | Sig. (2-tailed) | ,000 | ,949 | ,309 | ,006 | ,017 | ,697 |
| | N | 45 | 45 | 45 | 45 | 45 | 45 |
| CHOL_change | Correlation Coefficient | -,308 | ,118 | -,260 | ,153 | -,016 | ,110 |
| | Sig. (2-tailed) | ,039 | ,441 | ,084 | ,316 | ,914 | ,474 |
| | N | 45 | 45 | 45 | 45 | 45 | 45 |

### Correlations

| | | Desulfovibrio_change | Eggerthella_change | Erysipelatoclostridium_change | Faecalitalea_change | Family.XIII. AD3011. group_change | Family.XIII. UCG. 001_change |
|---|---|---|---|---|---|---|---|
| age | Correlation Coefficient | ,135 | ,151 | ,005 | -,041 | -,057 | ,067 |
| | Sig. (2-tailed) | ,378 | ,322 | ,973 | ,788 | ,711 | ,663 |
| | N | 45 | 45 | 45 | 45 | 45 | 45 |
| BMI | Correlation Coefficient | -,268 | ,107 | ,250 | ,041 | ,033 | -,199 |
| | Sig. (2-tailed) | ,075 | ,485 | ,097 | ,791 | ,831 | ,190 |
| | N | 45 | 45 | 45 | 45 | 45 | 45 |
| FM_stand | Correlation Coefficient | -,284 | ,228 | ,207 | ,055 | -,024 | -,306 |
| | Sig. (2-tailed) | ,059 | ,131 | ,172 | ,721 | ,876 | ,041 |
| | N | 45 | 45 | 45 | 45 | 45 | 45 |
| Richness | Correlation Coefficient | -,048 | ,118 | ,120 | ,011 | ,350 | ,256 |
| | Sig. (2-tailed) | ,753 | ,438 | ,433 | ,942 | ,018 | ,090 |
| | N | 45 | 45 | 45 | 45 | 45 | 45 |
| shannon.effective | Correlation Coefficient | -,048 | ,104 | ,112 | ,026 | ,325 | ,236 |
| | Sig. (2-tailed) | ,755 | ,498 | ,464 | ,864 | ,029 | ,119 |
| | N | 45 | 45 | 45 | 45 | 45 | 45 |
| simpson.effective | Correlation Coefficient | -,059 | ,084 | ,126 | ,041 | ,293 | ,240 |
| | Sig. (2-tailed) | ,699 | ,583 | ,408 | ,788 | ,051 | ,113 |
| | N | 45 | 45 | 45 | 45 | 45 | 45 |
| Evenness | Correlation Coefficient | -,091 | ,081 | -,016 | ,117 | ,022 | -,001 |
| | Sig. (2-tailed) | ,553 | ,595 | ,919 | ,443 | ,885 | ,996 |
| | N | 45 | 45 | 45 | 45 | 45 | 45 |
| CHOL_change | Correlation Coefficient | -,270 | ,203 | ,152 | -,149 | -,174 | -,319 |
| | Sig. (2-tailed) | ,073 | ,181 | ,318 | ,328 | ,253 | ,033 |
| | N | 45 | 45 | 45 | 45 | 45 | 45 |

### Correlations

| | | Gemella_change | Gordonibacter_change | Holdemanella_change | Holdemania_change | Lachnospiraceae.FCS020.group_change | Lachnospiraceae.NK4A136.group_change |
|---|---|---|---|---|---|---|---|
| age | Correlation Coefficient | ,200 | ,239 | -,098 | ,177 | -,187 | ,152 |
| | Sig. (2-tailed) | ,187 | ,114 | ,523 | ,245 | ,220 | ,319 |
| | N | 45 | 45 | 45 | 45 | 45 | 45 |
| BMI | Correlation Coefficient | -,204 | ,173 | ,079 | ,284 | ,056 | ,109 |
| | Sig. (2-tailed) | ,180 | ,257 | ,606 | ,058 | ,713 | ,476 |
| | N | 45 | 45 | 45 | 45 | 45 | 45 |
| FM_stand | Correlation Coefficient | -,199 | ,090 | ,056 | ,017 | -,044 | ,114 |
| | Sig. (2-tailed) | ,189 | ,558 | ,713 | ,911 | ,774 | ,454 |
| | N | 45 | 45 | 45 | 45 | 45 | 45 |
| Richness | Correlation Coefficient | -,070 | ,244 | ,303 | ,395 | ,238 | ,177 |
| | Sig. (2-tailed) | ,648 | ,107 | ,043 | ,007 | ,115 | ,246 |
| | N | 45 | 45 | 45 | 45 | 45 | 45 |
| shannon.effective | Correlation Coefficient | -,050 | ,257 | ,318 | ,414 | ,193 | ,117 |
| | Sig. (2-tailed) | ,745 | ,089 | ,033 | ,005 | ,204 | ,443 |
| | N | 45 | 45 | 45 | 45 | 45 | 45 |
| simpson.effective | Correlation Coefficient | -,066 | ,266 | ,357 | ,438 | ,205 | ,108 |
| | Sig. (2-tailed) | ,667 | ,077 | ,016 | ,003 | ,178 | ,480 |
| | N | 45 | 45 | 45 | 45 | 45 | 45 |
| Evenness | Correlation Coefficient | -,034 | ,248 | ,092 | ,206 | -,019 | -,153 |
| | Sig. (2-tailed) | ,824 | ,101 | ,546 | ,175 | ,903 | ,316 |
| | N | 45 | 45 | 45 | 45 | 45 | 45 |
| CHOL_change | Correlation Coefficient | ,155 | ,192 | ,137 | -,201 | ,182 | ,013 |
| | Sig. (2-tailed) | ,311 | ,205 | ,369 | ,186 | ,232 | ,933 |
| | N | 45 | 45 | 45 | 45 | 45 | 45 |

### Correlations

| | | Lactiplantibacillus_change | Lactobacillus_change | Libanicoccus_change | Ligilactobacillus_change | Limosilactobacillus_change | Roseburia_change |
|---|---|---|---|---|---|---|---|
| age | Correlation Coefficient | -,003 | ,190 | ,250 | -,220 | ,280 | ,323 |
| | Sig. (2-tailed) | ,986 | ,212 | ,098 | ,146 | ,062 | ,030 |
| | N | 45 | 45 | 45 | 45 | 45 | 45 |
| BMI | Correlation Coefficient | -,024 | -,043 | ,042 | ,072 | -,257 | -,120 |
| | Sig. (2-tailed) | ,874 | ,777 | ,786 | ,637 | ,088 | ,434 |
| | N | 45 | 45 | 45 | 45 | 45 | 45 |
| FM_stand | Correlation Coefficient | ,182 | ,118 | -,048 | -,144 | -,212 | ,182 |
| | Sig. (2-tailed) | ,231 | ,440 | ,753 | ,345 | ,162 | ,232 |
| | N | 45 | 45 | 45 | 45 | 45 | 45 |
| Richness | Correlation Coefficient | -,214 | -,278 | ,110 | -,045 | -,141 | ,123 |
| | Sig. (2-tailed) | ,159 | ,065 | ,470 | ,768 | ,354 | ,419 |
| | N | 45 | 45 | 45 | 45 | 45 | 45 |
| shannon.effective | Correlation Coefficient | -,256 | -,322 | ,108 | ,004 | -,098 | ,096 |
| | Sig. (2-tailed) | ,090 | ,031 | ,481 | ,979 | ,523 | ,532 |
| | N | 45 | 45 | 45 | 45 | 45 | 45 |
| simpson.effective | Correlation Coefficient | -,284 | -,340 | ,007 | ,017 | -,108 | ,054 |
| | Sig. (2-tailed) | ,059 | ,022 | ,963 | ,914 | ,480 | ,726 |
| | N | 45 | 45 | 45 | 45 | 45 | 45 |
| Evenness | Correlation Coefficient | -,172 | -,226 | -,031 | ,145 | ,031 | -,126 |
| | Sig. (2-tailed) | ,258 | ,135 | ,837 | ,342 | ,839 | ,409 |
| | N | 45 | 45 | 45 | 45 | 45 | 45 |
| CHOL_change | Correlation Coefficient | -,052 | ,184 | -,032 | -,061 | -,076 | ,028 |
| | Sig. (2-tailed) | ,735 | ,225 | ,834 | ,689 | ,622 | ,857 |
| | N | 45 | 45 | 45 | 45 | 45 | 45 |

### Correlations

| | | Shuttleworthia_change | Subdoligranulum_change | UCG.003_change | X.Eubacterium..brachy.group_change | X.Ruminococcus..gauvreauii.group_change | X.Ruminococcus..torques.group_change |
|---|---|---|---|---|---|---|---|
| age | Correlation Coefficient | -,034 | -,033 | ,072 | -,090 | ,071 | -,302 |
| | Sig. (2-tailed) | ,825 | ,827 | ,638 | ,554 | ,642 | ,044 |
| | N | 45 | 45 | 45 | 45 | 45 | 45 |
| BMI | Correlation Coefficient | ,016 | ,175 | ,239 | -,048 | -,050 | ,053 |
| | Sig. (2-tailed) | ,915 | ,252 | ,113 | ,753 | ,744 | ,728 |
| | N | 45 | 45 | 45 | 45 | 45 | 45 |
| FM_stand | Correlation Coefficient | ,312 | -,188 | ,208 | ,150 | ,196 | ,019 |
| | Sig. (2-tailed) | ,037 | ,216 | ,170 | ,324 | ,196 | ,904 |
| | N | 45 | 45 | 45 | 45 | 45 | 45 |
| Richness | Correlation Coefficient | -,034 | -,033 | ,052 | ,244 | -,346 | ,079 |
| | Sig. (2-tailed) | ,826 | ,831 | ,736 | ,106 | ,020 | ,607 |
| | N | 45 | 45 | 45 | 45 | 45 | 45 |
| shannon.effective | Correlation Coefficient | -,090 | ,008 | ,083 | ,227 | -,288 | ,122 |
| | Sig. (2-tailed) | ,556 | ,956 | ,587 | ,133 | ,055 | ,425 |
| | N | 45 | 45 | 45 | 45 | 45 | 45 |
| simpson.effective | Correlation Coefficient | -,080 | ,101 | ,035 | ,229 | -,249 | ,163 |
| | Sig. (2-tailed) | ,603 | ,511 | ,818 | ,130 | ,099 | ,284 |
| | N | 45 | 45 | 45 | 45 | 45 | 45 |
| Evenness | Correlation Coefficient | -,186 | ,322 | ,030 | ,073 | ,176 | ,204 |
| | Sig. (2-tailed) | ,222 | ,031 | ,844 | ,632 | ,247 | ,179 |
| | N | 45 | 45 | 45 | 45 | 45 | 45 |
| CHOL_change | Correlation Coefficient | ,155 | -,110 | ,006 | ,000 | ,241 | -,067 |
| | Sig. (2-tailed) | ,308 | ,473 | ,971 | 1,000 | ,110 | ,663 |
| | N | 45 | 45 | 45 | 45 | 45 | 45 |

### Correlations

| | | | interaction_VTA_change | interaction_rOFC_change | interaction_rmOFC_change | sumscore_GIQLI_change | Stool_frequency_change | STGHRE_change |
|---|---|---|---|---|---|---|---|---|
| | LDL_change | Correlation Coefficient | -,080 | ,057 | -,199 | -,079 | -,075 | -,532 |
| | | Sig. (2-tailed) | ,607 | ,721 | ,196 | ,608 | ,631 | ,000 |
| | | N | 44 | 41 | 44 | 45 | 44 | 42 |
| | shannon.effective_change | Correlation Coefficient | ,241 | -,253 | -,154 | ,154 | -,270 | ,007 |
| | | Sig. (2-tailed) | ,120 | ,115 | ,324 | ,319 | ,080 | ,964 |
| | | N | 43 | 40 | 43 | 44 | 43 | 42 |
| | Richness_change | Correlation Coefficient | ,154 | -,211 | -,177 | ,122 | -,326 | -,070 |
| | | Sig. (2-tailed) | ,323 | ,191 | ,256 | ,431 | ,033 | ,659 |
| | | N | 43 | 40 | 43 | 44 | 43 | 42 |
| | Evenness_change | Correlation Coefficient | ,112 | -,227 | -,045 | ,171 | ,106 | ,124 |
| | | Sig. (2-tailed) | ,473 | ,159 | ,772 | ,266 | ,498 | ,433 |
| | | N | 43 | 40 | 43 | 44 | 43 | 42 |
| | Actinomyces_change | Correlation Coefficient | -,042 | ,268 | ,173 | -,130 | -,006 | ,008 |
| | | Sig. (2-tailed) | ,791 | ,094 | ,268 | ,401 | ,972 | ,958 |
| | | N | 43 | 40 | 43 | 44 | 43 | 42 |
| | Aerostipes_change | Correlation Coefficient | -,264 | -,206 | ,039 | ,021 | ,325 | -,040 |
| | | Sig. (2-tailed) | ,088 | ,201 | ,806 | ,892 | ,034 | ,803 |
| | | N | 43 | 40 | 43 | 44 | 43 | 42 |
| | Bifidobacterium_change | Correlation Coefficient | -,152 | -,136 | ,098 | -,109 | -,125 | -,225 |
| | | Sig. (2-tailed) | ,332 | ,401 | ,533 | ,482 | ,425 | ,151 |
| | | N | 43 | 40 | 43 | 44 | 43 | 42 |
| | Blautia_change | Correlation Coefficient | -,238 | ,179 | ,084 | ,166 | ,115 | ,163 |
| | | Sig. (2-tailed) | ,124 | ,270 | ,594 | ,283 | ,465 | ,301 |
| | | N | 43 | 40 | 43 | 44 | 43 | 42 |

### Correlations

| | | STPYY_change | STGLPT_change | age | BMI | FM_stand | Richness | shannon.effective |
|---|---|---|---|---|---|---|---|---|
| LDL_change | Correlation Coefficient | ,080 | ,233 | ,052 | ,202 | ,478 | -,042 | -,119 |
| | Sig. (2-tailed) | ,615 | ,137 | ,733 | ,179 | ,001 | ,781 | ,430 |
| | N | 42 | 42 | 46 | 46 | 46 | 46 | 46 |
| shannon.effective_change | Correlation Coefficient | -,089 | ,014 | -,019 | -,130 | ,014 | ,490 | ,570 |
| | Sig. (2-tailed) | ,575 | ,928 | ,903 | ,395 | ,927 | ,001 | ,000 |
| | N | 42 | 42 | 45 | 45 | 45 | 45 | 45 |
| Richness_change | Correlation Coefficient | -,204 | -,014 | ,089 | -,105 | ,167 | ,500 | ,512 |
| | Sig. (2-tailed) | ,196 | ,931 | ,559 | ,492 | ,274 | ,000 | ,000 |
| | N | 42 | 42 | 45 | 45 | 45 | 45 | 45 |
| Evenness_change | Correlation Coefficient | ,122 | ,073 | -,205 | -,086 | -,303 | ,137 | ,306 |
| | Sig. (2-tailed) | ,442 | ,644 | ,177 | ,575 | ,043 | ,370 | ,041 |
| | N | 42 | 42 | 45 | 45 | 45 | 45 | 45 |
| Actinomyces_change | Correlation Coefficient | ,084 | -,030 | -,004 | -,121 | ,085 | -,028 | -,017 |
| | Sig. (2-tailed) | ,599 | ,852 | ,979 | ,429 | ,577 | ,856 | ,912 |
| | N | 42 | 42 | 45 | 45 | 45 | 45 | 45 |
| Aerostipes_change | Correlation Coefficient | ,001 | ,067 | -,078 | ,105 | -,157 | ,011 | ,036 |
| | Sig. (2-tailed) | ,995 | ,673 | ,611 | ,491 | ,305 | ,943 | ,816 |
| | N | 42 | 42 | 45 | 45 | 45 | 45 | 45 |
| Bifidobacterium_change | Correlation Coefficient | ,012 | ,014 | -,044 | ,013 | ,323 | -,514 | -,582 |
| | Sig. (2-tailed) | ,940 | ,929 | ,774 | ,933 | ,031 | ,000 | ,000 |
| | N | 42 | 42 | 45 | 45 | 45 | 45 | 45 |
| Blautia_change | Correlation Coefficient | -,021 | ,209 | ,083 | -,132 | -,159 | -,145 | -,055 |
| | Sig. (2-tailed) | ,894 | ,184 | ,588 | ,387 | ,296 | ,341 | ,722 |
| | N | 42 | 42 | 45 | 45 | 45 | 45 | 45 |

### Correlations

| | | | simpson. effective | Evenness | CHOL_change | LDL_change | shannon. effective_change | Richness_change |
|---|---|---|---|---|---|---|---|---|
| | LDL_change | Correlation Coefficient | -,149 | -,082 | ,926 | 1,000 | -,187 | -,043 |
| | | Sig. (2-tailed) | ,322 | ,588 | ,000 | . | ,218 | ,779 |
| | | N | 46 | 46 | 46 | 46 | 45 | 45 |
| | shannon.effective_change | Correlation Coefficient | ,586 | ,347 | -,144 | -,187 | 1,000 | ,901 |
| | | Sig. (2-tailed) | ,000 | ,020 | ,344 | ,218 | . | ,000 |
| | | N | 45 | 45 | 45 | 45 | 45 | 45 |
| | Richness_change | Correlation Coefficient | ,500 | ,163 | ,038 | -,043 | ,901 | 1,000 |
| | | Sig. (2-tailed) | ,000 | ,285 | ,806 | ,779 | ,000 | . |
| | | N | 45 | 45 | 45 | 45 | 45 | 45 |
| | Evenness_change | Correlation Coefficient | ,373 | ,600 | -,308 | -,206 | ,451 | ,062 |
| | | Sig. (2-tailed) | ,012 | ,000 | ,039 | ,175 | ,002 | ,687 |
| | | N | 45 | 45 | 45 | 45 | 45 | 45 |
| | Actinomyces_change | Correlation Coefficient | -,085 | ,010 | ,118 | ,092 | ,176 | ,275 |
| | | Sig. (2-tailed) | ,577 | ,949 | ,441 | ,547 | ,247 | ,067 |
| | | N | 45 | 45 | 45 | 45 | 45 | 45 |
| | Aerostipes_change | Correlation Coefficient | ,058 | ,155 | -,260 | -,246 | -,101 | -,185 |
| | | Sig. (2-tailed) | ,703 | ,309 | ,084 | ,103 | ,508 | ,223 |
| | | N | 45 | 45 | 45 | 45 | 45 | 45 |
| | Bifidobacterium_change | Correlation Coefficient | -,626 | -,401 | ,153 | ,145 | -,414 | -,300 |
| | | Sig. (2-tailed) | ,000 | ,006 | ,316 | ,343 | ,005 | ,045 |
| | | N | 45 | 45 | 45 | 45 | 45 | 45 |
| | Blautia_change | Correlation Coefficient | -,032 | ,355 | -,016 | -,062 | ,010 | -,036 |
| | | Sig. (2-tailed) | ,833 | ,017 | ,914 | ,684 | ,947 | ,817 |
| | | N | 45 | 45 | 45 | 45 | 45 | 45 |

### Correlations

| | | | Evenness_change | Actinomyces_change | Aerostipes_change | Bifidobacterium_change | Blautia_change | Collinsella_change |
|---|---|---|---|---|---|---|---|---|
| | LDL_change | Correlation Coefficient | -,206 | ,092 | -,246 | ,145 | -,062 | ,085 |
| | | Sig. (2-tailed) | ,175 | ,547 | ,103 | ,343 | ,684 | ,580 |
| | | N | 45 | 45 | 45 | 45 | 45 | 45 |
| | shannon.effective_change | Correlation Coefficient | ,451 | ,176 | -,101 | -,414 | ,010 | -,269 |
| | | Sig. (2-tailed) | ,002 | ,247 | ,508 | ,005 | ,947 | ,074 |
| | | N | 45 | 45 | 45 | 45 | 45 | 45 |
| | Richness_change | Correlation Coefficient | ,062 | ,275 | -,185 | -,300 | -,036 | -,256 |
| | | Sig. (2-tailed) | ,687 | ,067 | ,223 | ,045 | ,817 | ,090 |
| | | N | 45 | 45 | 45 | 45 | 45 | 45 |
| | Evenness_change | Correlation Coefficient | 1,000 | -,099 | ,142 | -,427 | ,167 | -,115 |
| | | Sig. (2-tailed) | . | ,516 | ,352 | ,003 | ,273 | ,452 |
| | | N | 45 | 45 | 45 | 45 | 45 | 45 |
| | Actinomyces_change | Correlation Coefficient | -,099 | 1,000 | -,172 | ,075 | ,364 | ,391 |
| | | Sig. (2-tailed) | ,516 | . | ,259 | ,626 | ,014 | ,008 |
| | | N | 45 | 45 | 45 | 45 | 45 | 45 |
| | Aerostipes_change | Correlation Coefficient | ,142 | -,172 | 1,000 | -,064 | -,023 | -,083 |
| | | Sig. (2-tailed) | ,352 | ,259 | . | ,675 | ,881 | ,587 |
| | | N | 45 | 45 | 45 | 45 | 45 | 45 |
| | Bifidobacterium_change | Correlation Coefficient | -,427 | ,075 | -,064 | 1,000 | -,024 | ,312 |
| | | Sig. (2-tailed) | ,003 | ,626 | ,675 | . | ,876 | ,037 |
| | | N | 45 | 45 | 45 | 45 | 45 | 45 |
| | Blautia_change | Correlation Coefficient | ,167 | ,364 | -,023 | -,024 | 1,000 | ,173 |
| | | Sig. (2-tailed) | ,273 | ,014 | ,881 | ,876 | . | ,256 |
| | | N | 45 | 45 | 45 | 45 | 45 | 45 |

### Correlations

| | | | Desulfovibrio_change | Eggerthella_change | Erysipelatoclostridium_change | Faecalitalea_change | Family.XIII.AD3011.group_change | Family.XIII.UCG.001_change |
|---|---|---|---|---|---|---|---|---|
| | LDL_change | Correlation Coefficient | -,147 | ,137 | ,160 | -,170 | -,126 | -,286 |
| | | Sig. (2-tailed) | ,334 | ,371 | ,295 | ,263 | ,409 | ,056 |
| | | N | 45 | 45 | 45 | 45 | 45 | 45 |
| | shannon.effective_change | Correlation Coefficient | ,041 | -,006 | ,287 | -,093 | ,334 | ,194 |
| | | Sig. (2-tailed) | ,791 | ,966 | ,056 | ,543 | ,025 | ,201 |
| | | N | 45 | 45 | 45 | 45 | 45 | 45 |
| | Richness_change | Correlation Coefficient | ,028 | ,091 | ,267 | -,076 | ,297 | ,180 |
| | | Sig. (2-tailed) | ,857 | ,552 | ,077 | ,620 | ,048 | ,237 |
| | | N | 45 | 45 | 45 | 45 | 45 | 45 |
| | Evenness_change | Correlation Coefficient | ,122 | -,169 | ,145 | -,015 | ,144 | ,090 |
| | | Sig. (2-tailed) | ,423 | ,268 | ,341 | ,921 | ,345 | ,557 |
| | | N | 45 | 45 | 45 | 45 | 45 | 45 |
| | Actinomyces_change | Correlation Coefficient | -,057 | ,254 | -,007 | ,178 | ,214 | -,189 |
| | | Sig. (2-tailed) | ,711 | ,092 | ,966 | ,243 | ,159 | ,214 |
| | | N | 45 | 45 | 45 | 45 | 45 | 45 |
| | Aerostipes_change | Correlation Coefficient | ,062 | -,320 | -,375 | ,193 | ,098 | ,233 |
| | | Sig. (2-tailed) | ,684 | ,032 | ,011 | ,205 | ,524 | ,124 |
| | | N | 45 | 45 | 45 | 45 | 45 | 45 |
| | Bifidobacterium_change | Correlation Coefficient | -,004 | ,083 | -,023 | ,070 | -,494 | -,305 |
| | | Sig. (2-tailed) | ,978 | ,587 | ,879 | ,650 | ,001 | ,042 |
| | | N | 45 | 45 | 45 | 45 | 45 | 45 |
| | Blautia_change | Correlation Coefficient | ,021 | ,208 | ,012 | ,088 | ,014 | -,103 |
| | | Sig. (2-tailed) | ,891 | ,171 | ,937 | ,566 | ,927 | ,499 |
| | | N | 45 | 45 | 45 | 45 | 45 | 45 |

### Correlations

| | | Gemella_change | Gordonibacter_change | Holdemanella_change | Holdemania_change | Lachnospiraceae.FCS020.group_change | Lachnospiraceae.NK4A136.group_change |
|---|---|---|---|---|---|---|---|
| LDL_change | Correlation Coefficient | ,118 | ,030 | ,119 | -,187 | ,198 | ,030 |
| | Sig. (2-tailed) | ,439 | ,847 | ,437 | ,219 | ,193 | ,846 |
| | N | 45 | 45 | 45 | 45 | 45 | 45 |
| shannon.effective_change | Correlation Coefficient | ,046 | ,213 | ,130 | ,129 | ,090 | ,325 |
| | Sig. (2-tailed) | ,766 | ,160 | ,395 | ,400 | ,555 | ,029 |
| | N | 45 | 45 | 45 | 45 | 45 | 45 |
| Richness_change | Correlation Coefficient | ,103 | ,319 | ,156 | ,157 | ,105 | ,289 |
| | Sig. (2-tailed) | ,501 | ,033 | ,305 | ,304 | ,493 | ,055 |
| | N | 45 | 45 | 45 | 45 | 45 | 45 |
| Evenness_change | Correlation Coefficient | -,096 | -,097 | -,017 | ,045 | ,062 | ,138 |
| | Sig. (2-tailed) | ,532 | ,527 | ,911 | ,772 | ,686 | ,365 |
| | N | 45 | 45 | 45 | 45 | 45 | 45 |
| Actinomyces_change | Correlation Coefficient | ,497 | ,141 | ,092 | -,328 | ,082 | -,158 |
| | Sig. (2-tailed) | ,001 | ,357 | ,548 | ,028 | ,592 | ,300 |
| | N | 45 | 45 | 45 | 45 | 45 | 45 |
| Aerostipes_change | Correlation Coefficient | -,093 | -,081 | ,068 | ,138 | -,184 | -,157 |
| | Sig. (2-tailed) | ,542 | ,595 | ,657 | ,366 | ,227 | ,302 |
| | N | 45 | 45 | 45 | 45 | 45 | 45 |
| Bifidobacterium_change | Correlation Coefficient | ,039 | -,351 | -,172 | -,160 | -,103 | -,370 |
| | Sig. (2-tailed) | ,799 | ,018 | ,258 | ,294 | ,499 | ,012 |
| | N | 45 | 45 | 45 | 45 | 45 | 45 |
| Blautia_change | Correlation Coefficient | ,547 | ,121 | ,251 | -,244 | ,216 | -,377 |
| | Sig. (2-tailed) | ,000 | ,427 | ,096 | ,106 | ,154 | ,011 |
| | N | 45 | 45 | 45 | 45 | 45 | 45 |

### Correlations

| | | | Lactiplantibacillus_change | Lactobacillus_change | Libanicoccus_change | Ligilactobacillus_change | Limosilactobacillus_change | Roseburia_change |
|---|---|---|---|---|---|---|---|---|
| | LDL_change | Correlation Coefficient | -,114 | ,255 | -,003 | ,011 | -,056 | -,021 |
| | | Sig. (2-tailed) | ,455 | ,091 | ,986 | ,941 | ,713 | ,893 |
| | | N | 45 | 45 | 45 | 45 | 45 | 45 |
| | shannon.effective_change | Correlation Coefficient | -,116 | -,398 | -,164 | ,006 | ,038 | ,107 |
| | | Sig. (2-tailed) | ,446 | ,007 | ,281 | ,969 | ,803 | ,485 |
| | | N | 45 | 45 | 45 | 45 | 45 | 45 |
| | Richness_change | Correlation Coefficient | -,049 | -,220 | -,170 | -,101 | ,025 | ,211 |
| | | Sig. (2-tailed) | ,751 | ,147 | ,264 | ,508 | ,872 | ,165 |
| | | N | 45 | 45 | 45 | 45 | 45 | 45 |
| | Evenness_change | Correlation Coefficient | -,240 | -,370 | ,053 | ,320 | -,008 | -,188 |
| | | Sig. (2-tailed) | ,112 | ,012 | ,732 | ,032 | ,958 | ,216 |
| | | N | 45 | 45 | 45 | 45 | 45 | 45 |
| | Actinomyces_change | Correlation Coefficient | ,225 | -,099 | -,038 | ,003 | -,038 | -,040 |
| | | Sig. (2-tailed) | ,137 | ,519 | ,803 | ,982 | ,806 | ,792 |
| | | N | 45 | 45 | 45 | 45 | 45 | 45 |
| | Aerostipes_change | Correlation Coefficient | -,043 | -,033 | ,071 | -,024 | ,228 | -,111 |
| | | Sig. (2-tailed) | ,779 | ,828 | ,643 | ,874 | ,132 | ,468 |
| | | N | 45 | 45 | 45 | 45 | 45 | 45 |
| | Bifidobacterium_change | Correlation Coefficient | ,360 | ,159 | ,153 | ,072 | ,077 | ,142 |
| | | Sig. (2-tailed) | ,015 | ,297 | ,315 | ,640 | ,616 | ,353 |
| | | N | 45 | 45 | 45 | 45 | 45 | 45 |
| | Blautia_change | Correlation Coefficient | ,258 | -,086 | ,051 | ,133 | ,093 | -,313 |
| | | Sig. (2-tailed) | ,087 | ,575 | ,741 | ,384 | ,542 | ,037 |
| | | N | 45 | 45 | 45 | 45 | 45 | 45 |

### Correlations

| | | | Shuttleworthia_ change | Subdoligranulu m_change | UCG. 003_change | X.Eubacterium.. brachy. group_change | X. Ruminococcus. .gauvreauii. group_change | X. Ruminococcus. .torques. group_change |
|---|---|---|---|---|---|---|---|---|
| | LDL_change | Correlation Coefficient | ,231 | -,104 | -,018 | -,050 | ,241 | -,037 |
| | | Sig. (2-tailed) | ,126 | ,496 | ,907 | ,745 | ,111 | ,812 |
| | | N | 45 | 45 | 45 | 45 | 45 | 45 |
| | shannon.effective_change | Correlation Coefficient | -,047 | ,162 | ,051 | ,281 | ,010 | ,083 |
| | | Sig. (2-tailed) | ,758 | ,288 | ,738 | ,061 | ,950 | ,586 |
| | | N | 45 | 45 | 45 | 45 | 45 | 45 |
| | Richness_change | Correlation Coefficient | ,056 | ,054 | ,088 | ,364 | -,028 | ,019 |
| | | Sig. (2-tailed) | ,717 | ,722 | ,567 | ,014 | ,854 | ,904 |
| | | N | 45 | 45 | 45 | 45 | 45 | 45 |
| | Evenness_change | Correlation Coefficient | -,228 | ,348 | -,141 | -,061 | ,121 | ,242 |
| | | Sig. (2-tailed) | ,132 | ,019 | ,357 | ,689 | ,428 | ,109 |
| | | N | 45 | 45 | 45 | 45 | 45 | 45 |
| | Actinomyces_change | Correlation Coefficient | -,089 | ,121 | -,185 | ,205 | ,018 | ,286 |
| | | Sig. (2-tailed) | ,561 | ,428 | ,225 | ,178 | ,908 | ,056 |
| | | N | 45 | 45 | 45 | 45 | 45 | 45 |
| | Aerostipes_change | Correlation Coefficient | -,052 | ,163 | -,084 | ,134 | -,190 | ,134 |
| | | Sig. (2-tailed) | ,735 | ,285 | ,582 | ,381 | ,211 | ,380 |
| | | N | 45 | 45 | 45 | 45 | 45 | 45 |
| | Bifidobacterium_change | Correlation Coefficient | ,255 | -,264 | -,189 | -,085 | ,233 | -,128 |
| | | Sig. (2-tailed) | ,091 | ,079 | ,215 | ,580 | ,124 | ,402 |
| | | N | 45 | 45 | 45 | 45 | 45 | 45 |
| | Blautia_change | Correlation Coefficient | -,246 | ,239 | -,396 | ,195 | ,209 | ,288 |
| | | Sig. (2-tailed) | ,104 | ,114 | ,007 | ,199 | ,169 | ,055 |
| | | N | 45 | 45 | 45 | 45 | 45 | 45 |

### Correlations

| | | | interaction_VTA_change | interaction_rOFC_change | interaction_rmOFC_change | sumscore_GIQLI_change | Stool_frequency_change | STGHRE_change |
|---|---|---|---|---|---|---|---|---|
| | Collinsella_change | Correlation Coefficient | ,024 | ,178 | ,150 | -,202 | ,151 | ,014 |
| | | Sig. (2-tailed) | ,878 | ,271 | ,339 | ,189 | ,333 | ,930 |
| | | N | 43 | 40 | 43 | 44 | 43 | 42 |
| | Desulfovibrio_change | Correlation Coefficient | ,034 | -,009 | -,084 | ,059 | ,066 | -,095 |
| | | Sig. (2-tailed) | ,830 | ,955 | ,591 | ,704 | ,674 | ,551 |
| | | N | 43 | 40 | 43 | 44 | 43 | 42 |
| | Eggerthella_change | Correlation Coefficient | ,020 | ,022 | -,027 | ,104 | -,131 | -,126 |
| | | Sig. (2-tailed) | ,899 | ,892 | ,862 | ,503 | ,403 | ,425 |
| | | N | 43 | 40 | 43 | 44 | 43 | 42 |
| | Erysipelatoclostridium_change | Correlation Coefficient | ,159 | -,021 | -,063 | ,190 | -,291 | -,075 |
| | | Sig. (2-tailed) | ,309 | ,900 | ,688 | ,216 | ,058 | ,639 |
| | | N | 43 | 40 | 43 | 44 | 43 | 42 |
| | Faecalitalea_change | Correlation Coefficient | -,099 | -,018 | ,035 | ,038 | -,040 | ,166 |
| | | Sig. (2-tailed) | ,526 | ,911 | ,825 | ,809 | ,798 | ,294 |
| | | N | 43 | 40 | 43 | 44 | 43 | 42 |
| | Family.XIII.AD3011.group_change | Correlation Coefficient | ,121 | ,191 | ,070 | ,180 | ,250 | ,125 |
| | | Sig. (2-tailed) | ,440 | ,238 | ,655 | ,241 | ,105 | ,430 |
| | | N | 43 | 40 | 43 | 44 | 43 | 42 |
| | Family.XIII.UCG.001_change | Correlation Coefficient | -,080 | ,064 | -,206 | ,009 | -,054 | ,215 |
| | | Sig. (2-tailed) | ,611 | ,696 | ,185 | ,953 | ,731 | ,171 |
| | | N | 43 | 40 | 43 | 44 | 43 | 42 |
| | Gemella_change | Correlation Coefficient | -,174 | ,295 | ,196 | ,086 | -,119 | -,105 |
| | | Sig. (2-tailed) | ,264 | ,064 | ,209 | ,577 | ,445 | ,510 |
| | | N | 43 | 40 | 43 | 44 | 43 | 42 |

### Correlations

| | | | STPYY_change | STGLPT_change | age | BMI | FM_stand | Richness | shannon. effective |
|---|---|---|---|---|---|---|---|---|---|
| | Collinsella_change | Correlation Coefficient | ,176 | ,004 | -,228 | -,035 | ,013 | -,354 | -,322 |
| | | Sig. (2-tailed) | ,266 | ,978 | ,133 | ,820 | ,932 | ,017 | ,031 |
| | | N | 42 | 42 | 45 | 45 | 45 | 45 | 45 |
| | Desulfovibrio_change | Correlation Coefficient | -,370 | -,069 | ,135 | -,268 | -,284 | -,048 | -,048 |
| | | Sig. (2-tailed) | ,016 | ,666 | ,378 | ,075 | ,059 | ,753 | ,755 |
| | | N | 42 | 42 | 45 | 45 | 45 | 45 | 45 |
| | Eggerthella_change | Correlation Coefficient | ,032 | ,103 | ,151 | ,107 | ,228 | ,118 | ,104 |
| | | Sig. (2-tailed) | ,839 | ,518 | ,322 | ,485 | ,131 | ,438 | ,498 |
| | | N | 42 | 42 | 45 | 45 | 45 | 45 | 45 |
| | Erysipelatoclostridium_change | Correlation Coefficient | -,017 | ,296 | ,005 | ,250 | ,207 | ,120 | ,112 |
| | | Sig. (2-tailed) | ,913 | ,057 | ,973 | ,097 | ,172 | ,433 | ,464 |
| | | N | 42 | 42 | 45 | 45 | 45 | 45 | 45 |
| | Faecalitalea_change | Correlation Coefficient | ,011 | ,090 | -,041 | ,041 | ,055 | ,011 | ,026 |
| | | Sig. (2-tailed) | ,945 | ,570 | ,788 | ,791 | ,721 | ,942 | ,864 |
| | | N | 42 | 42 | 45 | 45 | 45 | 45 | 45 |
| | Family.XIII.AD3011.group_change | Correlation Coefficient | ,171 | ,058 | -,057 | ,033 | -,024 | ,350 | ,325 |
| | | Sig. (2-tailed) | ,279 | ,717 | ,711 | ,831 | ,876 | ,018 | ,029 |
| | | N | 42 | 42 | 45 | 45 | 45 | 45 | 45 |
| | Family.XIII.UCG.001_change | Correlation Coefficient | ,017 | ,205 | ,067 | -,199 | -,306 | ,256 | ,236 |
| | | Sig. (2-tailed) | ,913 | ,192 | ,663 | ,190 | ,041 | ,090 | ,119 |
| | | N | 42 | 42 | 45 | 45 | 45 | 45 | 45 |
| | Gemella_change | Correlation Coefficient | -,139 | ,150 | ,200 | -,204 | -,199 | -,070 | -,050 |
| | | Sig. (2-tailed) | ,381 | ,342 | ,187 | ,180 | ,189 | ,648 | ,745 |
| | | N | 42 | 42 | 45 | 45 | 45 | 45 | 45 |

### Correlations

| | | | simpson. effective | Evenness | CHOL_change | LDL_change | shannon. effective_change | Richness_change |
|---|---|---|---|---|---|---|---|---|
| | Collinsella_change | Correlation Coefficient | -,369 | -,060 | ,110 | ,085 | -,269 | -,256 |
| | | Sig. (2-tailed) | ,012 | ,697 | ,474 | ,580 | ,074 | ,090 |
| | | N | 45 | 45 | 45 | 45 | 45 | 45 |
| | Desulfovibrio_change | Correlation Coefficient | -,059 | -,091 | -,270 | -,147 | ,041 | ,028 |
| | | Sig. (2-tailed) | ,699 | ,553 | ,073 | ,334 | ,791 | ,857 |
| | | N | 45 | 45 | 45 | 45 | 45 | 45 |
| | Eggerthella_change | Correlation Coefficient | ,084 | ,081 | ,203 | ,137 | -,006 | ,091 |
| | | Sig. (2-tailed) | ,583 | ,595 | ,181 | ,371 | ,966 | ,552 |
| | | N | 45 | 45 | 45 | 45 | 45 | 45 |
| | Erysipelatoclostridium_change | Correlation Coefficient | ,126 | -,016 | ,152 | ,160 | ,287 | ,267 |
| | | Sig. (2-tailed) | ,408 | ,919 | ,318 | ,295 | ,056 | ,077 |
| | | N | 45 | 45 | 45 | 45 | 45 | 45 |
| | Faecalitalea_change | Correlation Coefficient | ,041 | ,117 | -,149 | -,170 | -,093 | -,076 |
| | | Sig. (2-tailed) | ,788 | ,443 | ,328 | ,263 | ,543 | ,620 |
| | | N | 45 | 45 | 45 | 45 | 45 | 45 |
| | Family.XIII.AD3011. group_change | Correlation Coefficient | ,293 | ,022 | -,174 | -,126 | ,334 | ,297 |
| | | Sig. (2-tailed) | ,051 | ,885 | ,253 | ,409 | ,025 | ,048 |
| | | N | 45 | 45 | 45 | 45 | 45 | 45 |
| | Family.XIII.UCG. 001_change | Correlation Coefficient | ,240 | -,001 | -,319 | -,286 | ,194 | ,180 |
| | | Sig. (2-tailed) | ,113 | ,996 | ,033 | ,056 | ,201 | ,237 |
| | | N | 45 | 45 | 45 | 45 | 45 | 45 |
| | Gemella_change | Correlation Coefficient | -,066 | -,034 | ,155 | ,118 | ,046 | ,103 |
| | | Sig. (2-tailed) | ,667 | ,824 | ,311 | ,439 | ,766 | ,501 |
| | | N | 45 | 45 | 45 | 45 | 45 | 45 |

### Correlations

| | | | Evenness_change | Actinomyces_change | Aerostipes_change | Bifidobacterium_change | Blautia_change | Collinsella_change |
|---|---|---|---|---|---|---|---|---|
| | Collinsella_change | Correlation Coefficient | -,115 | ,391 | -,083 | ,312 | ,173 | 1,000 |
| | | Sig. (2-tailed) | ,452 | ,008 | ,587 | ,037 | ,256 | . |
| | | N | 45 | 45 | 45 | 45 | 45 | 45 |
| | Desulfovibrio_change | Correlation Coefficient | ,122 | -,057 | ,062 | -,004 | ,021 | -,270 |
| | | Sig. (2-tailed) | ,423 | ,711 | ,684 | ,978 | ,891 | ,073 |
| | | N | 45 | 45 | 45 | 45 | 45 | 45 |
| | Eggerthella_change | Correlation Coefficient | -,169 | ,254 | -,320 | ,083 | ,208 | -,016 |
| | | Sig. (2-tailed) | ,268 | ,092 | ,032 | ,587 | ,171 | ,919 |
| | | N | 45 | 45 | 45 | 45 | 45 | 45 |
| | Erysipelatoclostridium_change | Correlation Coefficient | ,145 | -,007 | -,375 | -,023 | ,012 | ,054 |
| | | Sig. (2-tailed) | ,341 | ,966 | ,011 | ,879 | ,937 | ,724 |
| | | N | 45 | 45 | 45 | 45 | 45 | 45 |
| | Faecalitalea_change | Correlation Coefficient | -,015 | ,178 | ,193 | ,070 | ,088 | -,073 |
| | | Sig. (2-tailed) | ,921 | ,243 | ,205 | ,650 | ,566 | ,632 |
| | | N | 45 | 45 | 45 | 45 | 45 | 45 |
| | Family.XIII.AD3011.group_change | Correlation Coefficient | ,144 | ,214 | ,098 | -,494 | ,014 | -,084 |
| | | Sig. (2-tailed) | ,345 | ,159 | ,524 | ,001 | ,927 | ,583 |
| | | N | 45 | 45 | 45 | 45 | 45 | 45 |
| | Family.XIII.UCG.001_change | Correlation Coefficient | ,090 | -,189 | ,233 | -,305 | -,103 | -,363 |
| | | Sig. (2-tailed) | ,557 | ,214 | ,124 | ,042 | ,499 | ,014 |
| | | N | 45 | 45 | 45 | 45 | 45 | 45 |
| | Gemella_change | Correlation Coefficient | -,096 | ,497 | -,093 | ,039 | ,547 | ,322 |
| | | Sig. (2-tailed) | ,532 | ,001 | ,542 | ,799 | ,000 | ,031 |
| | | N | 45 | 45 | 45 | 45 | 45 | 45 |

### Correlations

| | | | Desulfovibrio_change | Eggerthella_change | Erysipelatoclostridium_change | Faecalitalea_change | Family.XIII.AD3011.group_change | Family.XIII.UCG.001_change |
|---|---|---|---|---|---|---|---|---|
| | Collinsella_change | Correlation Coefficient | -,270 | -,016 | ,054 | -,073 | -,084 | -,363 |
| | | Sig. (2-tailed) | ,073 | ,919 | ,724 | ,632 | ,583 | ,014 |
| | | N | 45 | 45 | 45 | 45 | 45 | 45 |
| | Desulfovibrio_change | Correlation Coefficient | 1,000 | -,155 | ,038 | ,067 | -,074 | ,158 |
| | | Sig. (2-tailed) | . | ,311 | ,805 | ,663 | ,629 | ,300 |
| | | N | 45 | 45 | 45 | 45 | 45 | 45 |
| | Eggerthella_change | Correlation Coefficient | -,155 | 1,000 | ,224 | ,365 | -,051 | -,217 |
| | | Sig. (2-tailed) | ,311 | . | ,139 | ,014 | ,737 | ,151 |
| | | N | 45 | 45 | 45 | 45 | 45 | 45 |
| | Erysipelatoclostridium_change | Correlation Coefficient | ,038 | ,224 | 1,000 | ,148 | ,065 | ,064 |
| | | Sig. (2-tailed) | ,805 | ,139 | . | ,331 | ,673 | ,676 |
| | | N | 45 | 45 | 45 | 45 | 45 | 45 |
| | Faecalitalea_change | Correlation Coefficient | ,067 | ,365 | ,148 | 1,000 | -,136 | ,064 |
| | | Sig. (2-tailed) | ,663 | ,014 | ,331 | . | ,374 | ,678 |
| | | N | 45 | 45 | 45 | 45 | 45 | 45 |
| | Family.XIII.AD3011.group_change | Correlation Coefficient | -,074 | -,051 | ,065 | -,136 | 1,000 | ,249 |
| | | Sig. (2-tailed) | ,629 | ,737 | ,673 | ,374 | . | ,100 |
| | | N | 45 | 45 | 45 | 45 | 45 | 45 |
| | Family.XIII.UCG.001_change | Correlation Coefficient | ,158 | -,217 | ,064 | ,064 | ,249 | 1,000 |
| | | Sig. (2-tailed) | ,300 | ,151 | ,676 | ,678 | ,100 | . |
| | | N | 45 | 45 | 45 | 45 | 45 | 45 |
| | Gemella_change | Correlation Coefficient | ,194 | ,023 | ,101 | ,008 | -,021 | -,127 |
| | | Sig. (2-tailed) | ,202 | ,879 | ,510 | ,957 | ,893 | ,405 |
| | | N | 45 | 45 | 45 | 45 | 45 | 45 |

### Correlations

| | | Gemella_change | Gordonibacter_change | Holdemanella_change | Holdemania_change | Lachnospiraceae.FCS020.group_change | Lachnospiraceae.NK4A136.group_change |
|---|---|---|---|---|---|---|---|
| Collinsella_change | Correlation Coefficient | ,322 | -,052 | ,071 | -,286 | ,106 | -,529 |
| | Sig. (2-tailed) | ,031 | ,737 | ,644 | ,057 | ,489 | ,000 |
| | N | 45 | 45 | 45 | 45 | 45 | 45 |
| Desulfovibrio_change | Correlation Coefficient | ,194 | -,078 | -,030 | ,003 | -,042 | -,024 |
| | Sig. (2-tailed) | ,202 | ,610 | ,845 | ,986 | ,782 | ,873 |
| | N | 45 | 45 | 45 | 45 | 45 | 45 |
| Eggerthella_change | Correlation Coefficient | ,023 | ,384 | ,098 | ,139 | ,020 | -,054 |
| | Sig. (2-tailed) | ,879 | ,009 | ,521 | ,362 | ,897 | ,727 |
| | N | 45 | 45 | 45 | 45 | 45 | 45 |
| Erysipelatoclostridium_change | Correlation Coefficient | ,101 | ,224 | ,088 | -,060 | ,162 | ,052 |
| | Sig. (2-tailed) | ,510 | ,140 | ,566 | ,694 | ,288 | ,736 |
| | N | 45 | 45 | 45 | 45 | 45 | 45 |
| Faecalitalea_change | Correlation Coefficient | ,008 | ,183 | ,085 | ,040 | -,221 | -,278 |
| | Sig. (2-tailed) | ,957 | ,230 | ,581 | ,794 | ,145 | ,064 |
| | N | 45 | 45 | 45 | 45 | 45 | 45 |
| Family.XIII.AD3011.group_change | Correlation Coefficient | -,021 | -,040 | ,227 | -,016 | ,038 | ,335 |
| | Sig. (2-tailed) | ,893 | ,792 | ,134 | ,916 | ,802 | ,024 |
| | N | 45 | 45 | 45 | 45 | 45 | 45 |
| Family.XIII.UCG.001_change | Correlation Coefficient | -,127 | -,023 | -,114 | ,170 | ,064 | ,207 |
| | Sig. (2-tailed) | ,405 | ,879 | ,458 | ,263 | ,676 | ,172 |
| | N | 45 | 45 | 45 | 45 | 45 | 45 |
| Gemella_change | Correlation Coefficient | 1,000 | ,128 | ,330 | -,465 | ,300 | -,431 |
| | Sig. (2-tailed) | . | ,402 | ,027 | ,001 | ,045 | ,003 |
| | N | 45 | 45 | 45 | 45 | 45 | 45 |

### Correlations

| | | | Lactiplantibacillus_change | Lactobacillus_change | Libanicoccus_change | Ligilactobacillus_change | Limosilactobacillus_change | Roseburia_change |
|---|---|---|---|---|---|---|---|---|
| | Collinsella_change | Correlation Coefficient | ,286 | -,006 | ,077 | ,218 | ,132 | -,219 |
| | | Sig. (2-tailed) | ,057 | ,969 | ,614 | ,150 | ,388 | ,149 |
| | | N | 45 | 45 | 45 | 45 | 45 | 45 |
| | Desulfovibrio_change | Correlation Coefficient | -,018 | ,201 | ,338 | -,054 | ,258 | ,130 |
| | | Sig. (2-tailed) | ,905 | ,186 | ,023 | ,725 | ,087 | ,393 |
| | | N | 45 | 45 | 45 | 45 | 45 | 45 |
| | Eggerthella_change | Correlation Coefficient | -,150 | ,141 | ,069 | -,030 | -,057 | -,037 |
| | | Sig. (2-tailed) | ,326 | ,356 | ,652 | ,845 | ,712 | ,807 |
| | | N | 45 | 45 | 45 | 45 | 45 | 45 |
| | Erysipelatoclostridium_change | Correlation Coefficient | -,149 | -,121 | ,019 | ,245 | -,069 | ,107 |
| | | Sig. (2-tailed) | ,329 | ,430 | ,903 | ,104 | ,652 | ,484 |
| | | N | 45 | 45 | 45 | 45 | 45 | 45 |
| | Faecalitalea_change | Correlation Coefficient | -,141 | ,157 | ,072 | -,043 | ,036 | ,126 |
| | | Sig. (2-tailed) | ,355 | ,303 | ,638 | ,778 | ,812 | ,411 |
| | | N | 45 | 45 | 45 | 45 | 45 | 45 |
| | Family.XIII.AD3011.group_change | Correlation Coefficient | ,185 | -,149 | -,323 | ,035 | -,104 | -,224 |
| | | Sig. (2-tailed) | ,224 | ,330 | ,031 | ,821 | ,496 | ,139 |
| | | N | 45 | 45 | 45 | 45 | 45 | 45 |
| | Family.XIII.UCG.001_change | Correlation Coefficient | -,151 | ,058 | -,146 | ,222 | ,128 | ,120 |
| | | Sig. (2-tailed) | ,322 | ,705 | ,337 | ,142 | ,402 | ,432 |
| | | N | 45 | 45 | 45 | 45 | 45 | 45 |
| | Gemella_change | Correlation Coefficient | ,304 | ,020 | ,122 | ,093 | ,253 | -,094 |
| | | Sig. (2-tailed) | ,043 | ,899 | ,424 | ,545 | ,094 | ,541 |
| | | N | 45 | 45 | 45 | 45 | 45 | 45 |

### Correlations

| | | | Shuttleworthia_change | Subdoligranulum_change | UCG.003_change | X.Eubacterium..brachy.group_change | X.Ruminococcus..gauvreauii.group_change | X.Ruminococcus..torques.group_change |
|---|---|---|---|---|---|---|---|---|
| | Collinsella_change | Correlation Coefficient | ,092 | ,025 | -,109 | -,063 | ,005 | ,158 |
| | | Sig. (2-tailed) | ,546 | ,869 | ,474 | ,682 | ,976 | ,300 |
| | | N | 45 | 45 | 45 | 45 | 45 | 45 |
| | Desulfovibrio_change | Correlation Coefficient | ,020 | -,113 | -,052 | ,100 | -,043 | -,151 |
| | | Sig. (2-tailed) | ,894 | ,462 | ,735 | ,513 | ,778 | ,323 |
| | | N | 45 | 45 | 45 | 45 | 45 | 45 |
| | Eggerthella_change | Correlation Coefficient | ,146 | ,022 | -,180 | ,125 | ,102 | -,038 |
| | | Sig. (2-tailed) | ,338 | ,886 | ,237 | ,412 | ,505 | ,804 |
| | | N | 45 | 45 | 45 | 45 | 45 | 45 |
| | Erysipelatoclostridium_change | Correlation Coefficient | ,078 | ,064 | ,000 | ,156 | ,038 | ,060 |
| | | Sig. (2-tailed) | ,613 | ,676 | ,999 | ,307 | ,803 | ,698 |
| | | N | 45 | 45 | 45 | 45 | 45 | 45 |
| | Faecalitalea_change | Correlation Coefficient | -,141 | ,039 | -,038 | ,302 | -,074 | ,190 |
| | | Sig. (2-tailed) | ,356 | ,801 | ,804 | ,044 | ,629 | ,211 |
| | | N | 45 | 45 | 45 | 45 | 45 | 45 |
| | Family.XIII.AD3011.group_change | Correlation Coefficient | -,048 | ,090 | -,018 | -,084 | -,026 | ,281 |
| | | Sig. (2-tailed) | ,752 | ,555 | ,905 | ,585 | ,867 | ,062 |
| | | N | 45 | 45 | 45 | 45 | 45 | 45 |
| | Family.XIII.UCG.001_change | Correlation Coefficient | -,280 | -,025 | ,070 | ,068 | -,256 | ,041 |
| | | Sig. (2-tailed) | ,062 | ,872 | ,645 | ,655 | ,089 | ,791 |
| | | N | 45 | 45 | 45 | 45 | 45 | 45 |
| | Gemella_change | Correlation Coefficient | -,146 | ,072 | -,274 | ,233 | ,000 | ,082 |
| | | Sig. (2-tailed) | ,339 | ,636 | ,069 | ,123 | ,998 | ,594 |
| | | N | 45 | 45 | 45 | 45 | 45 | 45 |

### Correlations

| | | interaction_VTA_change | interaction_rOFC_change | interaction_rmOFC_change | sumscore_GIQLI_change | Stool_frequency_change | STGHRE_change |
|---|---|---|---|---|---|---|---|
| Gordonibacter_change | Correlation Coefficient | ,025 | ,044 | ,079 | ,079 | -,099 | -,040 |
| | Sig. (2-tailed) | ,876 | ,785 | ,614 | ,610 | ,526 | ,803 |
| | N | 43 | 40 | 43 | 44 | 43 | 42 |
| Holdemanella_change | Correlation Coefficient | -,083 | ,049 | ,005 | ,160 | -,104 | -,104 |
| | Sig. (2-tailed) | ,597 | ,764 | ,972 | ,301 | ,505 | ,514 |
| | N | 43 | 40 | 43 | 44 | 43 | 42 |
| Holdemania_change | Correlation Coefficient | -,116 | -,259 | -,195 | ,095 | -,016 | ,015 |
| | Sig. (2-tailed) | ,459 | ,107 | ,211 | ,539 | ,919 | ,927 |
| | N | 43 | 40 | 43 | 44 | 43 | 42 |
| Lachnospiraceae.FCS020.group_change | Correlation Coefficient | -,041 | -,079 | ,028 | ,133 | -,071 | -,078 |
| | Sig. (2-tailed) | ,792 | ,627 | ,860 | ,390 | ,649 | ,624 |
| | N | 43 | 40 | 43 | 44 | 43 | 42 |
| Lachnospiraceae.NK4A136.group_change | Correlation Coefficient | ,286 | -,093 | -,001 | ,000 | -,013 | ,094 |
| | Sig. (2-tailed) | ,063 | ,566 | ,996 | ,998 | ,933 | ,554 |
| | N | 43 | 40 | 43 | 44 | 43 | 42 |
| Lactiplantibacillus_change | Correlation Coefficient | -,006 | ,120 | ,397 | ,168 | -,007 | ,131 |
| | Sig. (2-tailed) | ,972 | ,459 | ,008 | ,276 | ,966 | ,407 |
| | N | 43 | 40 | 43 | 44 | 43 | 42 |
| Lactobacillus_change | Correlation Coefficient | -,023 | ,046 | -,054 | -,033 | ,105 | -,070 |
| | Sig. (2-tailed) | ,882 | ,778 | ,731 | ,833 | ,504 | ,659 |
| | N | 43 | 40 | 43 | 44 | 43 | 42 |
| Libanicoccus_change | Correlation Coefficient | ,294 | ,211 | ,223 | -,057 | -,187 | -,107 |
| | Sig. (2-tailed) | ,056 | ,192 | ,151 | ,712 | ,230 | ,499 |
| | N | 43 | 40 | 43 | 44 | 43 | 42 |

### Correlations

| | | | STPYY_change | STGLPT_change | age | BMI | FM_stand | Richness | shannon.effective |
|---|---|---|---|---|---|---|---|---|---|
| | Gordonibacter_change | Correlation Coefficient | -,341 | ,132 | ,239 | ,173 | ,090 | ,244 | ,257 |
| | | Sig. (2-tailed) | ,027 | ,404 | ,114 | ,257 | ,558 | ,107 | ,089 |
| | | N | 42 | 42 | 45 | 45 | 45 | 45 | 45 |
| | Holdemanella_change | Correlation Coefficient | -,043 | ,166 | -,098 | ,079 | ,056 | ,303 | ,318 |
| | | Sig. (2-tailed) | ,785 | ,294 | ,523 | ,606 | ,713 | ,043 | ,033 |
| | | N | 42 | 42 | 45 | 45 | 45 | 45 | 45 |
| | Holdemania_change | Correlation Coefficient | -,157 | -,266 | ,177 | ,284 | ,017 | ,395 | ,414 |
| | | Sig. (2-tailed) | ,321 | ,089 | ,245 | ,058 | ,911 | ,007 | ,005 |
| | | N | 42 | 42 | 45 | 45 | 45 | 45 | 45 |
| | Lachnospiraceae.FCS020.group_change | Correlation Coefficient | ,076 | ,162 | -,187 | ,056 | -,044 | ,238 | ,193 |
| | | Sig. (2-tailed) | ,633 | ,307 | ,220 | ,713 | ,774 | ,115 | ,204 |
| | | N | 42 | 42 | 45 | 45 | 45 | 45 | 45 |
| | Lachnospiraceae.NK4A136.group_change | Correlation Coefficient | ,155 | -,079 | ,152 | ,109 | ,114 | ,177 | ,117 |
| | | Sig. (2-tailed) | ,326 | ,621 | ,319 | ,476 | ,454 | ,246 | ,443 |
| | | N | 42 | 42 | 45 | 45 | 45 | 45 | 45 |
| | Lactiplantibacillus_change | Correlation Coefficient | -,079 | -,186 | -,003 | -,024 | ,182 | -,214 | -,256 |
| | | Sig. (2-tailed) | ,618 | ,238 | ,986 | ,874 | ,231 | ,159 | ,090 |
| | | N | 42 | 42 | 45 | 45 | 45 | 45 | 45 |
| | Lactobacillus_change | Correlation Coefficient | ,046 | -,181 | ,190 | -,043 | ,118 | -,278 | -,322 |
| | | Sig. (2-tailed) | ,771 | ,251 | ,212 | ,777 | ,440 | ,065 | ,031 |
| | | N | 42 | 42 | 45 | 45 | 45 | 45 | 45 |
| | Libanicoccus_change | Correlation Coefficient | ,039 | ,115 | ,250 | ,042 | -,048 | ,110 | ,108 |
| | | Sig. (2-tailed) | ,807 | ,468 | ,098 | ,786 | ,753 | ,470 | ,481 |
| | | N | 42 | 42 | 45 | 45 | 45 | 45 | 45 |

### Correlations

| | | | simpson. effective | Evenness | CHOL_change | LDL_change | shannon. effective_change | Richness_change |
|---|---|---|---|---|---|---|---|---|
| | Gordonibacter_change | Correlation Coefficient | ,266 | ,248 | ,192 | ,030 | ,213 | ,319 |
| | | Sig. (2-tailed) | ,077 | ,101 | ,205 | ,847 | ,160 | ,033 |
| | | N | 45 | 45 | 45 | 45 | 45 | 45 |
| | Holdemanella_change | Correlation Coefficient | ,357 | ,092 | ,137 | ,119 | ,130 | ,156 |
| | | Sig. (2-tailed) | ,016 | ,546 | ,369 | ,437 | ,395 | ,305 |
| | | N | 45 | 45 | 45 | 45 | 45 | 45 |
| | Holdemania_change | Correlation Coefficient | ,438 | ,206 | -,201 | -,187 | ,129 | ,157 |
| | | Sig. (2-tailed) | ,003 | ,175 | ,186 | ,219 | ,400 | ,304 |
| | | N | 45 | 45 | 45 | 45 | 45 | 45 |
| | Lachnospiraceae.FCS020.group_change | Correlation Coefficient | ,205 | -,019 | ,182 | ,198 | ,090 | ,105 |
| | | Sig. (2-tailed) | ,178 | ,903 | ,232 | ,193 | ,555 | ,493 |
| | | N | 45 | 45 | 45 | 45 | 45 | 45 |
| | Lachnospiraceae. NK4A136.group_change | Correlation Coefficient | ,108 | -,153 | ,013 | ,030 | ,325 | ,289 |
| | | Sig. (2-tailed) | ,480 | ,316 | ,933 | ,846 | ,029 | ,055 |
| | | N | 45 | 45 | 45 | 45 | 45 | 45 |
| | Lactiplantibacillus_change | Correlation Coefficient | -,284 | -,172 | -,052 | -,114 | -,116 | -,049 |
| | | Sig. (2-tailed) | ,059 | ,258 | ,735 | ,455 | ,446 | ,751 |
| | | N | 45 | 45 | 45 | 45 | 45 | 45 |
| | Lactobacillus_change | Correlation Coefficient | -,340 | -,226 | ,184 | ,255 | -,398 | -,220 |
| | | Sig. (2-tailed) | ,022 | ,135 | ,225 | ,091 | ,007 | ,147 |
| | | N | 45 | 45 | 45 | 45 | 45 | 45 |
| | Libanicoccus_change | Correlation Coefficient | ,007 | -,031 | -,032 | -,003 | -,164 | -,170 |
| | | Sig. (2-tailed) | ,963 | ,837 | ,834 | ,986 | ,281 | ,264 |
| | | N | 45 | 45 | 45 | 45 | 45 | 45 |

### Correlations

| | | | Evenness_change | Actinomyces_change | Aerostipes_change | Bifidobacterium_change | Blautia_change | Collinsella_change |
|---|---|---|---|---|---|---|---|---|
| | Gordonibacter_change | Correlation Coefficient | -,097 | ,141 | -,081 | -,351 | ,121 | -,052 |
| | | Sig. (2-tailed) | ,527 | ,357 | ,595 | ,018 | ,427 | ,737 |
| | | N | 45 | 45 | 45 | 45 | 45 | 45 |
| | Holdemanella_change | Correlation Coefficient | -,017 | ,092 | ,068 | -,172 | ,251 | ,071 |
| | | Sig. (2-tailed) | ,911 | ,548 | ,657 | ,258 | ,096 | ,644 |
| | | N | 45 | 45 | 45 | 45 | 45 | 45 |
| | Holdemania_change | Correlation Coefficient | ,045 | -,328 | ,138 | -,160 | -,244 | -,286 |
| | | Sig. (2-tailed) | ,772 | ,028 | ,366 | ,294 | ,106 | ,057 |
| | | N | 45 | 45 | 45 | 45 | 45 | 45 |
| | Lachnospiraceae.FCS020.group_change | Correlation Coefficient | ,062 | ,082 | -,184 | -,103 | ,216 | ,106 |
| | | Sig. (2-tailed) | ,686 | ,592 | ,227 | ,499 | ,154 | ,489 |
| | | N | 45 | 45 | 45 | 45 | 45 | 45 |
| | Lachnospiraceae.NK4A136.group_change | Correlation Coefficient | ,138 | -,158 | -,157 | -,370 | -,377 | -,529 |
| | | Sig. (2-tailed) | ,365 | ,300 | ,302 | ,012 | ,011 | ,000 |
| | | N | 45 | 45 | 45 | 45 | 45 | 45 |
| | Lactiplantibacillus_change | Correlation Coefficient | -,240 | ,225 | -,043 | ,360 | ,258 | ,286 |
| | | Sig. (2-tailed) | ,112 | ,137 | ,779 | ,015 | ,087 | ,057 |
| | | N | 45 | 45 | 45 | 45 | 45 | 45 |
| | Lactobacillus_change | Correlation Coefficient | -,370 | -,099 | -,033 | ,159 | -,086 | -,006 |
| | | Sig. (2-tailed) | ,012 | ,519 | ,828 | ,297 | ,575 | ,969 |
| | | N | 45 | 45 | 45 | 45 | 45 | 45 |
| | Libanicoccus_change | Correlation Coefficient | ,053 | -,038 | ,071 | ,153 | ,051 | ,077 |
| | | Sig. (2-tailed) | ,732 | ,803 | ,643 | ,315 | ,741 | ,614 |
| | | N | 45 | 45 | 45 | 45 | 45 | 45 |

### Correlations

| | | | Desulfovibrio_change | Eggerthella_change | Erysipelatoclostridium_change | Faecalitalea_change | Family.XIII.AD3011.group_change | Family.XIII.UCG.001_change |
|---|---|---|---|---|---|---|---|---|
| | Gordonibacter_change | Correlation Coefficient | -,078 | ,384 | ,224 | ,183 | -,040 | -,023 |
| | | Sig. (2-tailed) | ,610 | ,009 | ,140 | ,230 | ,792 | ,879 |
| | | N | 45 | 45 | 45 | 45 | 45 | 45 |
| | Holdemanella_change | Correlation Coefficient | -,030 | ,098 | ,088 | ,085 | ,227 | -,114 |
| | | Sig. (2-tailed) | ,845 | ,521 | ,566 | ,581 | ,134 | ,458 |
| | | N | 45 | 45 | 45 | 45 | 45 | 45 |
| | Holdemania_change | Correlation Coefficient | ,003 | ,139 | -,060 | ,040 | -,016 | ,170 |
| | | Sig. (2-tailed) | ,986 | ,362 | ,694 | ,794 | ,916 | ,263 |
| | | N | 45 | 45 | 45 | 45 | 45 | 45 |
| | Lachnospiraceae.FCS020.group_change | Correlation Coefficient | -,042 | ,020 | ,162 | -,221 | ,038 | ,064 |
| | | Sig. (2-tailed) | ,782 | ,897 | ,288 | ,145 | ,802 | ,676 |
| | | N | 45 | 45 | 45 | 45 | 45 | 45 |
| | Lachnospiraceae.NK4A136.group_change | Correlation Coefficient | -,024 | -,054 | ,052 | -,278 | ,335 | ,207 |
| | | Sig. (2-tailed) | ,873 | ,727 | ,736 | ,064 | ,024 | ,172 |
| | | N | 45 | 45 | 45 | 45 | 45 | 45 |
| | Lactiplantibacillus_change | Correlation Coefficient | -,018 | -,150 | -,149 | -,141 | ,185 | -,151 |
| | | Sig. (2-tailed) | ,905 | ,326 | ,329 | ,355 | ,224 | ,322 |
| | | N | 45 | 45 | 45 | 45 | 45 | 45 |
| | Lactobacillus_change | Correlation Coefficient | ,201 | ,141 | -,121 | ,157 | -,149 | ,058 |
| | | Sig. (2-tailed) | ,186 | ,356 | ,430 | ,303 | ,330 | ,705 |
| | | N | 45 | 45 | 45 | 45 | 45 | 45 |
| | Libanicoccus_change | Correlation Coefficient | ,338 | ,069 | ,019 | ,072 | -,323 | -,146 |
| | | Sig. (2-tailed) | ,023 | ,652 | ,903 | ,638 | ,031 | ,337 |
| | | N | 45 | 45 | 45 | 45 | 45 | 45 |

### Correlations

| | | | Gemella_change | Gordonibacter_change | Holdemanella_change | Holdemania_change | Lachnospiraceae.FCS020.group_change | Lachnospiraceae.NK4A136.group_change |
|---|---|---|---|---|---|---|---|---|
| | Gordonibacter_change | Correlation Coefficient | ,128 | 1,000 | ,166 | ,164 | -,040 | ,003 |
| | | Sig. (2-tailed) | ,402 | . | ,275 | ,283 | ,792 | ,985 |
| | | N | 45 | 45 | 45 | 45 | 45 | 45 |
| | Holdemanella_change | Correlation Coefficient | ,330 | ,166 | 1,000 | ,013 | ,367 | -,336 |
| | | Sig. (2-tailed) | ,027 | ,275 | . | ,934 | ,013 | ,024 |
| | | N | 45 | 45 | 45 | 45 | 45 | 45 |
| | Holdemania_change | Correlation Coefficient | -,465 | ,164 | ,013 | 1,000 | -,172 | ,161 |
| | | Sig. (2-tailed) | ,001 | ,283 | ,934 | . | ,257 | ,290 |
| | | N | 45 | 45 | 45 | 45 | 45 | 45 |
| | Lachnospiraceae.FCS020.group_change | Correlation Coefficient | ,300 | -,040 | ,367 | -,172 | 1,000 | -,205 |
| | | Sig. (2-tailed) | ,045 | ,792 | ,013 | ,257 | . | ,177 |
| | | N | 45 | 45 | 45 | 45 | 45 | 45 |
| | Lachnospiraceae.NK4A136.group_change | Correlation Coefficient | -,431 | ,003 | -,336 | ,161 | -,205 | 1,000 |
| | | Sig. (2-tailed) | ,003 | ,985 | ,024 | ,290 | ,177 | . |
| | | N | 45 | 45 | 45 | 45 | 45 | 45 |
| | Lactiplantibacillus_change | Correlation Coefficient | ,304 | -,077 | ,033 | -,103 | ,056 | -,185 |
| | | Sig. (2-tailed) | ,043 | ,617 | ,831 | ,500 | ,713 | ,225 |
| | | N | 45 | 45 | 45 | 45 | 45 | 45 |
| | Lactobacillus_change | Correlation Coefficient | ,020 | -,064 | -,100 | -,023 | -,047 | -,079 |
| | | Sig. (2-tailed) | ,899 | ,677 | ,515 | ,879 | ,757 | ,604 |
| | | N | 45 | 45 | 45 | 45 | 45 | 45 |
| | Libanicoccus_change | Correlation Coefficient | ,122 | ,034 | -,152 | ,160 | -,142 | -,229 |
| | | Sig. (2-tailed) | ,424 | ,822 | ,319 | ,292 | ,353 | ,130 |
| | | N | 45 | 45 | 45 | 45 | 45 | 45 |

### Correlations

| | | | Lactiplantibacillus_change | Lactobacillus_change | Libanicoccus_change | Ligilactobacillus_change | Limosilactobacillus_change | Roseburia_change |
|---|---|---|---|---|---|---|---|---|
| | Gordonibacter_change | Correlation Coefficient | -,077 | -,064 | ,034 | -,110 | ,153 | -,032 |
| | | Sig. (2-tailed) | ,617 | ,677 | ,822 | ,471 | ,316 | ,834 |
| | | N | 45 | 45 | 45 | 45 | 45 | 45 |
| | Holdemanella_change | Correlation Coefficient | ,033 | -,100 | -,152 | -,017 | ,005 | -,121 |
| | | Sig. (2-tailed) | ,831 | ,515 | ,319 | ,914 | ,972 | ,429 |
| | | N | 45 | 45 | 45 | 45 | 45 | 45 |
| | Holdemania_change | Correlation Coefficient | -,103 | -,023 | ,160 | ,046 | ,031 | ,134 |
| | | Sig. (2-tailed) | ,500 | ,879 | ,292 | ,766 | ,841 | ,379 |
| | | N | 45 | 45 | 45 | 45 | 45 | 45 |
| | Lachnospiraceae.FCS020.group_change | Correlation Coefficient | ,056 | -,047 | -,142 | ,232 | -,109 | -,294 |
| | | Sig. (2-tailed) | ,713 | ,757 | ,353 | ,125 | ,478 | ,050 |
| | | N | 45 | 45 | 45 | 45 | 45 | 45 |
| | Lachnospiraceae.NK4A136.group_change | Correlation Coefficient | -,185 | -,079 | -,229 | -,276 | -,141 | ,128 |
| | | Sig. (2-tailed) | ,225 | ,604 | ,130 | ,066 | ,355 | ,401 |
| | | N | 45 | 45 | 45 | 45 | 45 | 45 |
| | Lactiplantibacillus_change | Correlation Coefficient | 1,000 | -,039 | -,036 | ,077 | ,124 | -,116 |
| | | Sig. (2-tailed) | . | ,801 | ,814 | ,615 | ,416 | ,447 |
| | | N | 45 | 45 | 45 | 45 | 45 | 45 |
| | Lactobacillus_change | Correlation Coefficient | -,039 | 1,000 | ,186 | ,068 | ,364 | -,085 |
| | | Sig. (2-tailed) | ,801 | . | ,220 | ,659 | ,014 | ,580 |
| | | N | 45 | 45 | 45 | 45 | 45 | 45 |
| | Libanicoccus_change | Correlation Coefficient | -,036 | ,186 | 1,000 | -,038 | ,060 | -,058 |
| | | Sig. (2-tailed) | ,814 | ,220 | . | ,806 | ,696 | ,705 |
| | | N | 45 | 45 | 45 | 45 | 45 | 45 |

### Correlations

| | | | Shuttleworthia_change | Subdoligranulum_change | UCG.003_change | X.Eubacterium..brachy.group_change | X.Ruminococcus..gauvreauii.group_change | X.Ruminococcus..torques.group_change |
|---|---|---|---|---|---|---|---|---|
| | Gordonibacter_change | Correlation Coefficient | ,055 | ,172 | ,223 | ,415 | -,214 | -,156 |
| | | Sig. (2-tailed) | ,720 | ,258 | ,141 | ,005 | ,158 | ,305 |
| | | N | 45 | 45 | 45 | 45 | 45 | 45 |
| | Holdemanella_change | Correlation Coefficient | -,001 | -,056 | -,306 | ,278 | -,057 | ,204 |
| | | Sig. (2-tailed) | ,996 | ,714 | ,041 | ,064 | ,709 | ,179 |
| | | N | 45 | 45 | 45 | 45 | 45 | 45 |
| | Holdemania_change | Correlation Coefficient | ,144 | -,059 | ,323 | ,009 | -,214 | -,192 |
| | | Sig. (2-tailed) | ,346 | ,702 | ,030 | ,953 | ,158 | ,206 |
| | | N | 45 | 45 | 45 | 45 | 45 | 45 |
| | Lachnospiraceae.FCS020.group_change | Correlation Coefficient | -,029 | ,214 | -,251 | ,108 | -,127 | ,105 |
| | | Sig. (2-tailed) | ,848 | ,158 | ,096 | ,478 | ,405 | ,493 |
| | | N | 45 | 45 | 45 | 45 | 45 | 45 |
| | Lachnospiraceae.NK4A136.group_change | Correlation Coefficient | ,009 | -,050 | ,227 | -,157 | ,083 | -,207 |
| | | Sig. (2-tailed) | ,954 | ,745 | ,133 | ,303 | ,587 | ,173 |
| | | N | 45 | 45 | 45 | 45 | 45 | 45 |
| | Lactiplantibacillus_change | Correlation Coefficient | ,071 | -,040 | -,063 | -,026 | ,178 | -,047 |
| | | Sig. (2-tailed) | ,645 | ,795 | ,680 | ,865 | ,241 | ,760 |
| | | N | 45 | 45 | 45 | 45 | 45 | 45 |
| | Lactobacillus_change | Correlation Coefficient | ,103 | -,337 | -,049 | -,032 | ,044 | -,111 |
| | | Sig. (2-tailed) | ,502 | ,024 | ,748 | ,837 | ,773 | ,466 |
| | | N | 45 | 45 | 45 | 45 | 45 | 45 |
| | Libanicoccus_change | Correlation Coefficient | -,018 | -,245 | ,026 | ,087 | -,159 | -,241 |
| | | Sig. (2-tailed) | ,909 | ,105 | ,863 | ,568 | ,297 | ,110 |
| | | N | 45 | 45 | 45 | 45 | 45 | 45 |

### Correlations

| | | | interaction_VTA_change | interaction_rOFC_change | interaction_rmOFC_change | sumscore_GIQLI_change | Stool_frequency_change | STGHRE_change |
|---|---|---|---|---|---|---|---|---|
| | Ligilactobacillus_change | Correlation Coefficient | -,175 | -,105 | -,082 | ,054 | ,060 | -,017 |
| | | Sig. (2-tailed) | ,262 | ,521 | ,603 | ,727 | ,703 | ,914 |
| | | N | 43 | 40 | 43 | 44 | 43 | 42 |
| | Limosilactobacillus_change | Correlation Coefficient | -,010 | -,005 | -,070 | ,058 | ,169 | -,196 |
| | | Sig. (2-tailed) | ,949 | ,976 | ,654 | ,706 | ,278 | ,212 |
| | | N | 43 | 40 | 43 | 44 | 43 | 42 |
| | Roseburia_change | Correlation Coefficient | ,137 | ,146 | -,011 | ,025 | -,316 | -,209 |
| | | Sig. (2-tailed) | ,380 | ,369 | ,944 | ,870 | ,039 | ,184 |
| | | N | 43 | 40 | 43 | 44 | 43 | 42 |
| | Shuttleworthia_change | Correlation Coefficient | ,191 | -,155 | ,032 | ,210 | ,115 | -,164 |
| | | Sig. (2-tailed) | ,220 | ,338 | ,838 | ,172 | ,462 | ,300 |
| | | N | 43 | 40 | 43 | 44 | 43 | 42 |
| | Subdoligranulum_change | Correlation Coefficient | -,384 | -,159 | -,015 | ,261 | ,083 | ,279 |
| | | Sig. (2-tailed) | ,011 | ,329 | ,922 | ,087 | ,598 | ,073 |
| | | N | 43 | 40 | 43 | 44 | 43 | 42 |
| | UCG.003_change | Correlation Coefficient | ,185 | ,122 | -,094 | -,255 | -,067 | -,059 |
| | | Sig. (2-tailed) | ,236 | ,453 | ,551 | ,095 | ,669 | ,711 |
| | | N | 43 | 40 | 43 | 44 | 43 | 42 |
| | X.Eubacterium..brachy.group_change | Correlation Coefficient | ,115 | -,177 | ,038 | ,090 | -,196 | -,097 |
| | | Sig. (2-tailed) | ,464 | ,275 | ,809 | ,561 | ,209 | ,540 |
| | | N | 43 | 40 | 43 | 44 | 43 | 42 |
| | X.Ruminococcus..gauvreauii.group_change | Correlation Coefficient | -,194 | -,143 | -,024 | ,113 | -,091 | -,100 |
| | | Sig. (2-tailed) | ,211 | ,377 | ,879 | ,465 | ,560 | ,528 |
| | | N | 43 | 40 | 43 | 44 | 43 | 42 |

### Correlations

| | | | STPYY_change | STGLPT_change | age | BMI | FM_stand | Richness | shannon. effective |
|---|---|---|---|---|---|---|---|---|---|
| | Ligilactobacillus_change | Correlation Coefficient | ,015 | ,038 | -,220 | ,072 | -,144 | -,045 | ,004 |
| | | Sig. (2-tailed) | ,924 | ,813 | ,146 | ,637 | ,345 | ,768 | ,979 |
| | | N | 42 | 42 | 45 | 45 | 45 | 45 | 45 |
| | Limosilactobacillus_change | Correlation Coefficient | ,027 | -,182 | ,280 | -,257 | -,212 | -,141 | -,098 |
| | | Sig. (2-tailed) | ,864 | ,248 | ,062 | ,088 | ,162 | ,354 | ,523 |
| | | N | 42 | 42 | 45 | 45 | 45 | 45 | 45 |
| | Roseburia_change | Correlation Coefficient | -,048 | -,035 | ,323 | -,120 | ,182 | ,123 | ,096 |
| | | Sig. (2-tailed) | ,763 | ,824 | ,030 | ,434 | ,232 | ,419 | ,532 |
| | | N | 42 | 42 | 45 | 45 | 45 | 45 | 45 |
| | Shuttleworthia_change | Correlation Coefficient | -,068 | -,074 | -,034 | ,016 | ,312 | -,034 | -,090 |
| | | Sig. (2-tailed) | ,669 | ,644 | ,825 | ,915 | ,037 | ,826 | ,556 |
| | | N | 42 | 42 | 45 | 45 | 45 | 45 | 45 |
| | Subdoligranulum_change | Correlation Coefficient | -,210 | ,009 | -,033 | ,175 | -,188 | -,033 | ,008 |
| | | Sig. (2-tailed) | ,182 | ,957 | ,827 | ,252 | ,216 | ,831 | ,956 |
| | | N | 42 | 42 | 45 | 45 | 45 | 45 | 45 |
| | UCG.003_change | Correlation Coefficient | -,176 | -,165 | ,072 | ,239 | ,208 | ,052 | ,083 |
| | | Sig. (2-tailed) | ,264 | ,296 | ,638 | ,113 | ,170 | ,736 | ,587 |
| | | N | 42 | 42 | 45 | 45 | 45 | 45 | 45 |
| | X.Eubacterium..brachy. group_change | Correlation Coefficient | -,061 | ,179 | -,090 | -,048 | ,150 | ,244 | ,227 |
| | | Sig. (2-tailed) | ,702 | ,256 | ,554 | ,753 | ,324 | ,106 | ,133 |
| | | N | 42 | 42 | 45 | 45 | 45 | 45 | 45 |
| | X.Ruminococcus.. gauvreauii.group_change | Correlation Coefficient | -,063 | -,159 | ,071 | -,050 | ,196 | -,346 | -,288 |
| | | Sig. (2-tailed) | ,693 | ,314 | ,642 | ,744 | ,196 | ,020 | ,055 |
| | | N | 42 | 42 | 45 | 45 | 45 | 45 | 45 |

### Correlations

| | | | simpson. effective | Evenness | CHOL_change | LDL_change | shannon. effective_change | Richness_change |
|---|---|---|---|---|---|---|---|---|
| | Ligilactobacillus_change | Correlation Coefficient | ,017 | ,145 | -,061 | ,011 | ,006 | -,101 |
| | | Sig. (2-tailed) | ,914 | ,342 | ,689 | ,941 | ,969 | ,508 |
| | | N | 45 | 45 | 45 | 45 | 45 | 45 |
| | Limosilactobacillus_change | Correlation Coefficient | -,108 | ,031 | -,076 | -,056 | ,038 | ,025 |
| | | Sig. (2-tailed) | ,480 | ,839 | ,622 | ,713 | ,803 | ,872 |
| | | N | 45 | 45 | 45 | 45 | 45 | 45 |
| | Roseburia_change | Correlation Coefficient | ,054 | -,126 | ,028 | -,021 | ,107 | ,211 |
| | | Sig. (2-tailed) | ,726 | ,409 | ,857 | ,893 | ,485 | ,165 |
| | | N | 45 | 45 | 45 | 45 | 45 | 45 |
| | Shuttleworthia_change | Correlation Coefficient | -,080 | -,186 | ,155 | ,231 | -,047 | ,056 |
| | | Sig. (2-tailed) | ,603 | ,222 | ,308 | ,126 | ,758 | ,717 |
| | | N | 45 | 45 | 45 | 45 | 45 | 45 |
| | Subdoligranulum_change | Correlation Coefficient | ,101 | ,322 | -,110 | -,104 | ,162 | ,054 |
| | | Sig. (2-tailed) | ,511 | ,031 | ,473 | ,496 | ,288 | ,722 |
| | | N | 45 | 45 | 45 | 45 | 45 | 45 |
| | UCG.003_change | Correlation Coefficient | ,035 | ,030 | ,006 | -,018 | ,051 | ,088 |
| | | Sig. (2-tailed) | ,818 | ,844 | ,971 | ,907 | ,738 | ,567 |
| | | N | 45 | 45 | 45 | 45 | 45 | 45 |
| | X.Eubacterium..brachy. group_change | Correlation Coefficient | ,229 | ,073 | ,000 | -,050 | ,281 | ,364 |
| | | Sig. (2-tailed) | ,130 | ,632 | 1,000 | ,745 | ,061 | ,014 |
| | | N | 45 | 45 | 45 | 45 | 45 | 45 |
| | X.Ruminococcus.. gauvreauii.group_change | Correlation Coefficient | -,249 | ,176 | ,241 | ,241 | ,010 | -,028 |
| | | Sig. (2-tailed) | ,099 | ,247 | ,110 | ,111 | ,950 | ,854 |
| | | N | 45 | 45 | 45 | 45 | 45 | 45 |

### Correlations

| | | | Evenness_change | Actinomyces_change | Aerostipes_change | Bifidobacterium_change | Blautia_change | Collinsella_change |
|---|---|---|---|---|---|---|---|---|
| | Ligilactobacillus_change | Correlation Coefficient | ,320 | ,003 | -,024 | ,072 | ,133 | ,218 |
| | | Sig. (2-tailed) | ,032 | ,982 | ,874 | ,640 | ,384 | ,150 |
| | | N | 45 | 45 | 45 | 45 | 45 | 45 |
| | Limosilactobacillus_change | Correlation Coefficient | -,008 | -,038 | ,228 | ,077 | ,093 | ,132 |
| | | Sig. (2-tailed) | ,958 | ,806 | ,132 | ,616 | ,542 | ,388 |
| | | N | 45 | 45 | 45 | 45 | 45 | 45 |
| | Roseburia_change | Correlation Coefficient | -,188 | -,040 | -,111 | ,142 | -,313 | -,219 |
| | | Sig. (2-tailed) | ,216 | ,792 | ,468 | ,353 | ,037 | ,149 |
| | | N | 45 | 45 | 45 | 45 | 45 | 45 |
| | Shuttleworthia_change | Correlation Coefficient | -,228 | -,089 | -,052 | ,255 | -,246 | ,092 |
| | | Sig. (2-tailed) | ,132 | ,561 | ,735 | ,091 | ,104 | ,546 |
| | | N | 45 | 45 | 45 | 45 | 45 | 45 |
| | Subdoligranulum_change | Correlation Coefficient | ,348 | ,121 | ,163 | -,264 | ,239 | ,025 |
| | | Sig. (2-tailed) | ,019 | ,428 | ,285 | ,079 | ,114 | ,869 |
| | | N | 45 | 45 | 45 | 45 | 45 | 45 |
| | UCG.003_change | Correlation Coefficient | -,141 | -,185 | -,084 | -,189 | -,396 | -,109 |
| | | Sig. (2-tailed) | ,357 | ,225 | ,582 | ,215 | ,007 | ,474 |
| | | N | 45 | 45 | 45 | 45 | 45 | 45 |
| | X.Eubacterium..brachy.group_change | Correlation Coefficient | -,061 | ,205 | ,134 | -,085 | ,195 | -,063 |
| | | Sig. (2-tailed) | ,689 | ,178 | ,381 | ,580 | ,199 | ,682 |
| | | N | 45 | 45 | 45 | 45 | 45 | 45 |
| | X.Ruminococcus..gauvreauii.group_change | Correlation Coefficient | ,121 | ,018 | -,190 | ,233 | ,209 | ,005 |
| | | Sig. (2-tailed) | ,428 | ,908 | ,211 | ,124 | ,169 | ,976 |
| | | N | 45 | 45 | 45 | 45 | 45 | 45 |

### Correlations

| | | | Desulfovibrio_change | Eggerthella_change | Erysipelatoclostridium_change | Faecalitalea_change | Family.XIII.AD3011.group_change | Family.XIII.UCG.001_change |
|---|---|---|---|---|---|---|---|---|
| | Ligilactobacillus_change | Correlation Coefficient | -,054 | -,030 | ,245 | -,043 | ,035 | ,222 |
| | | Sig. (2-tailed) | ,725 | ,845 | ,104 | ,778 | ,821 | ,142 |
| | | N | 45 | 45 | 45 | 45 | 45 | 45 |
| | Limosilactobacillus_change | Correlation Coefficient | ,258 | -,057 | -,069 | ,036 | -,104 | ,128 |
| | | Sig. (2-tailed) | ,087 | ,712 | ,652 | ,812 | ,496 | ,402 |
| | | N | 45 | 45 | 45 | 45 | 45 | 45 |
| | Roseburia_change | Correlation Coefficient | ,130 | -,037 | ,107 | ,126 | -,224 | ,120 |
| | | Sig. (2-tailed) | ,393 | ,807 | ,484 | ,411 | ,139 | ,432 |
| | | N | 45 | 45 | 45 | 45 | 45 | 45 |
| | Shuttleworthia_change | Correlation Coefficient | ,020 | ,146 | ,078 | -,141 | -,048 | -,280 |
| | | Sig. (2-tailed) | ,894 | ,338 | ,613 | ,356 | ,752 | ,062 |
| | | N | 45 | 45 | 45 | 45 | 45 | 45 |
| | Subdoligranulum_change | Correlation Coefficient | -,113 | ,022 | ,064 | ,039 | ,090 | -,025 |
| | | Sig. (2-tailed) | ,462 | ,886 | ,676 | ,801 | ,555 | ,872 |
| | | N | 45 | 45 | 45 | 45 | 45 | 45 |
| | UCG.003_change | Correlation Coefficient | -,052 | -,180 | ,000 | -,038 | -,018 | ,070 |
| | | Sig. (2-tailed) | ,735 | ,237 | ,999 | ,804 | ,905 | ,645 |
| | | N | 45 | 45 | 45 | 45 | 45 | 45 |
| | X.Eubacterium..brachy.group_change | Correlation Coefficient | ,100 | ,125 | ,156 | ,302 | -,084 | ,068 |
| | | Sig. (2-tailed) | ,513 | ,412 | ,307 | ,044 | ,585 | ,655 |
| | | N | 45 | 45 | 45 | 45 | 45 | 45 |
| | X.Ruminococcus..gauvreauii.group_change | Correlation Coefficient | -,043 | ,102 | ,038 | -,074 | -,026 | -,256 |
| | | Sig. (2-tailed) | ,778 | ,505 | ,803 | ,629 | ,867 | ,089 |
| | | N | 45 | 45 | 45 | 45 | 45 | 45 |

### Correlations

| | | Gemella_change | Gordonibacter_change | Holdemanella_change | Holdemania_change | Lachnospiraceae.FCS020.group_change | Lachnospiraceae.NK4A136.group_change |
|---|---|---|---|---|---|---|---|
| Ligilactobacillus_change | Correlation Coefficient | ,093 | -,110 | -,017 | ,046 | ,232 | -,276 |
| | Sig. (2-tailed) | ,545 | ,471 | ,914 | ,766 | ,125 | ,066 |
| | N | 45 | 45 | 45 | 45 | 45 | 45 |
| Limosilactobacillus_change | Correlation Coefficient | ,253 | ,153 | ,005 | ,031 | -,109 | -,141 |
| | Sig. (2-tailed) | ,094 | ,316 | ,972 | ,841 | ,478 | ,355 |
| | N | 45 | 45 | 45 | 45 | 45 | 45 |
| Roseburia_change | Correlation Coefficient | -,094 | -,032 | -,121 | ,134 | -,294 | ,128 |
| | Sig. (2-tailed) | ,541 | ,834 | ,429 | ,379 | ,050 | ,401 |
| | N | 45 | 45 | 45 | 45 | 45 | 45 |
| Shuttleworthia_change | Correlation Coefficient | -,146 | ,055 | -,001 | ,144 | -,029 | ,009 |
| | Sig. (2-tailed) | ,339 | ,720 | ,996 | ,346 | ,848 | ,954 |
| | N | 45 | 45 | 45 | 45 | 45 | 45 |
| Subdoligranulum_change | Correlation Coefficient | ,072 | ,172 | -,056 | -,059 | ,214 | -,050 |
| | Sig. (2-tailed) | ,636 | ,258 | ,714 | ,702 | ,158 | ,745 |
| | N | 45 | 45 | 45 | 45 | 45 | 45 |
| UCG.003_change | Correlation Coefficient | -,274 | ,223 | -,306 | ,323 | -,251 | ,227 |
| | Sig. (2-tailed) | ,069 | ,141 | ,041 | ,030 | ,096 | ,133 |
| | N | 45 | 45 | 45 | 45 | 45 | 45 |
| X.Eubacterium..brachy.group_change | Correlation Coefficient | ,233 | ,415 | ,278 | ,009 | ,108 | -,157 |
| | Sig. (2-tailed) | ,123 | ,005 | ,064 | ,953 | ,478 | ,303 |
| | N | 45 | 45 | 45 | 45 | 45 | 45 |
| X.Ruminococcus..gauvreauii.group_change | Correlation Coefficient | ,000 | -,214 | -,057 | -,214 | -,127 | ,083 |
| | Sig. (2-tailed) | ,998 | ,158 | ,709 | ,158 | ,405 | ,587 |
| | N | 45 | 45 | 45 | 45 | 45 | 45 |

### Correlations

| | | Lactiplantibacillus_change | Lactobacillus_change | Libanicoccus_change | Ligilactobacillus_change | Limosilactobacillus_change | Roseburia_change |
|---|---|---|---|---|---|---|---|
| Ligilactobacillus_change | Correlation Coefficient | ,077 | ,068 | -,038 | 1,000 | ,039 | -,232 |
| | Sig. (2-tailed) | ,615 | ,659 | ,806 | . | ,799 | ,125 |
| | N | 45 | 45 | 45 | 45 | 45 | 45 |
| Limosilactobacillus_change | Correlation Coefficient | ,124 | ,364 | ,060 | ,039 | 1,000 | ,063 |
| | Sig. (2-tailed) | ,416 | ,014 | ,696 | ,799 | . | ,681 |
| | N | 45 | 45 | 45 | 45 | 45 | 45 |
| Roseburia_change | Correlation Coefficient | -,116 | -,085 | -,058 | -,232 | ,063 | 1,000 |
| | Sig. (2-tailed) | ,447 | ,580 | ,705 | ,125 | ,681 | . |
| | N | 45 | 45 | 45 | 45 | 45 | 45 |
| Shuttleworthia_change | Correlation Coefficient | ,071 | ,103 | -,018 | -,075 | ,067 | ,080 |
| | Sig. (2-tailed) | ,645 | ,502 | ,909 | ,623 | ,661 | ,603 |
| | N | 45 | 45 | 45 | 45 | 45 | 45 |
| Subdoligranulum_change | Correlation Coefficient | -,040 | -,337 | -,245 | ,258 | -,188 | -,247 |
| | Sig. (2-tailed) | ,795 | ,024 | ,105 | ,088 | ,217 | ,102 |
| | N | 45 | 45 | 45 | 45 | 45 | 45 |
| UCG.003_change | Correlation Coefficient | -,063 | -,049 | ,026 | -,174 | -,070 | ,239 |
| | Sig. (2-tailed) | ,680 | ,748 | ,863 | ,253 | ,648 | ,114 |
| | N | 45 | 45 | 45 | 45 | 45 | 45 |
| X.Eubacterium..brachy. group_change | Correlation Coefficient | -,026 | -,032 | ,087 | -,023 | ,038 | -,006 |
| | Sig. (2-tailed) | ,865 | ,837 | ,568 | ,880 | ,803 | ,967 |
| | N | 45 | 45 | 45 | 45 | 45 | 45 |
| X.Ruminococcus.. gauvreauii.group_change | Correlation Coefficient | ,178 | ,044 | -,159 | ,026 | -,040 | ,033 |
| | Sig. (2-tailed) | ,241 | ,773 | ,297 | ,864 | ,794 | ,831 |
| | N | 45 | 45 | 45 | 45 | 45 | 45 |

### Correlations

| | | Shuttleworthia_ change | Subdoligranulu m_change | UCG. 003_change | X.Eubacterium.. brachy. group_change | X. Ruminococcus. .gauvreauii. group_change | X. Ruminococcus. .torques. group_change |
|---|---|---|---|---|---|---|---|
| Ligilactobacillus_change | Correlation Coefficient | -,075 | ,258 | -,174 | -,023 | ,026 | ,101 |
| | Sig. (2-tailed) | ,623 | ,088 | ,253 | ,880 | ,864 | ,511 |
| | N | 45 | 45 | 45 | 45 | 45 | 45 |
| Limosilactobacillus_change | Correlation Coefficient | ,067 | -,188 | -,070 | ,038 | -,040 | -,198 |
| | Sig. (2-tailed) | ,661 | ,217 | ,648 | ,803 | ,794 | ,192 |
| | N | 45 | 45 | 45 | 45 | 45 | 45 |
| Roseburia_change | Correlation Coefficient | ,080 | -,247 | ,239 | -,006 | ,033 | -,192 |
| | Sig. (2-tailed) | ,603 | ,102 | ,114 | ,967 | ,831 | ,205 |
| | N | 45 | 45 | 45 | 45 | 45 | 45 |
| Shuttleworthia_change | Correlation Coefficient | 1,000 | -,188 | -,004 | ,083 | -,090 | -,069 |
| | Sig. (2-tailed) | . | ,215 | ,980 | ,589 | ,554 | ,652 |
| | N | 45 | 45 | 45 | 45 | 45 | 45 |
| Subdoligranulum_change | Correlation Coefficient | -,188 | 1,000 | -,157 | -,039 | ,286 | ,201 |
| | Sig. (2-tailed) | ,215 | . | ,302 | ,797 | ,057 | ,185 |
| | N | 45 | 45 | 45 | 45 | 45 | 45 |
| UCG.003_change | Correlation Coefficient | -,004 | -,157 | 1,000 | -,044 | -,265 | -,349 |
| | Sig. (2-tailed) | ,980 | ,302 | . | ,774 | ,078 | ,019 |
| | N | 45 | 45 | 45 | 45 | 45 | 45 |
| X.Eubacterium..brachy. group_change | Correlation Coefficient | ,083 | -,039 | -,044 | 1,000 | -,241 | ,086 |
| | Sig. (2-tailed) | ,589 | ,797 | ,774 | . | ,111 | ,573 |
| | N | 45 | 45 | 45 | 45 | 45 | 45 |
| X.Ruminococcus.. gauvreauii.group_change | Correlation Coefficient | -,090 | ,286 | -,265 | -,241 | 1,000 | -,016 |
| | Sig. (2-tailed) | ,554 | ,057 | ,078 | ,111 | . | ,915 |
| | N | 45 | 45 | 45 | 45 | 45 | 45 |

### Correlations

| | | interaction_VTA_change | interaction_rOFC_change | interaction_rmOFC_change | sumscore_GIQLI_change | Stool_frequency_change | STGHRE_change |
|---|---|---|---|---|---|---|---|
| X.Ruminococcus..torques.group_change | Correlation Coefficient | -,229 | -,194 | -,133 | ,305 | ,271 | ,230 |
| | Sig. (2-tailed) | ,140 | ,230 | ,397 | ,044 | ,079 | ,144 |
| | N | 43 | 40 | 43 | 44 | 43 | 42 |

### Correlations

| | | STPYY_change | STGLPT_change | age | BMI | FM_stand | Richness | shannon.effective |
|---|---|---|---|---|---|---|---|---|
| X.Ruminococcus..torques.group_change | Correlation Coefficient | ,183 | ,092 | -,302 | ,053 | ,019 | ,079 | ,122 |
| | Sig. (2-tailed) | ,247 | ,561 | ,044 | ,728 | ,904 | ,607 | ,425 |
| | N | 42 | 42 | 45 | 45 | 45 | 45 | 45 |

### Correlations

| | | simpson.effective | Evenness | CHOL_change | LDL_change | shannon.effective_change | Richness_change |
|---|---|---|---|---|---|---|---|
| X.Ruminococcus..torques.group_change | Correlation Coefficient | ,163 | ,204 | -,067 | -,037 | ,083 | ,019 |
| | Sig. (2-tailed) | ,284 | ,179 | ,663 | ,812 | ,586 | ,904 |
| | N | 45 | 45 | 45 | 45 | 45 | 45 |

### Correlations

| | | Evenness_change | Actinomyces_change | Aerostipes_change | Bifidobacterium_change | Blautia_change | Collinsella_change |
|---|---|---|---|---|---|---|---|
| X.Ruminococcus..torques. group_change | Correlation Coefficient | ,242 | ,286 | ,134 | -,128 | ,288 | ,158 |
| | Sig. (2-tailed) | ,109 | ,056 | ,380 | ,402 | ,055 | ,300 |
| | N | 45 | 45 | 45 | 45 | 45 | 45 |

### Correlations

| | | Desulfovibrio_change | Eggerthella_change | Erysipelatoclostridium_change | Faecalitalea_change | Family.XIII. AD3011. group_change | Family.XIII. UCG. 001_change |
|---|---|---|---|---|---|---|---|
| X.Ruminococcus..torques. group_change | Correlation Coefficient | -,151 | -,038 | ,060 | ,190 | ,281 | ,041 |
| | Sig. (2-tailed) | ,323 | ,804 | ,698 | ,211 | ,062 | ,791 |
| | N | 45 | 45 | 45 | 45 | 45 | 45 |

### Correlations

| | | Gemella_change | Gordonibacter_change | Holdemanella_change | Holdemania_change | Lachnospiraceae.FCS020. group_change | Lachnospiraceae.NK4A136. group_change |
|---|---|---|---|---|---|---|---|
| X.Ruminococcus..torques. group_change | Correlation Coefficient | ,082 | -,156 | ,204 | -,192 | ,105 | -,207 |
| | Sig. (2-tailed) | ,594 | ,305 | ,179 | ,206 | ,493 | ,173 |
| | N | 45 | 45 | 45 | 45 | 45 | 45 |

### Correlations

| | | Lactiplantibacillus_change | Lactobacillus_change | Libanicoccus_change | Ligilactobacillus_change | Limosilactobacillus_change | Roseburia_change |
|---|---|---|---|---|---|---|---|
| X.Ruminococcus..torques.group_change | Correlation Coefficient | -,047 | -,111 | -,241 | ,101 | -,198 | -,192 |
| | Sig. (2-tailed) | ,760 | ,466 | ,110 | ,511 | ,192 | ,205 |
| | N | 45 | 45 | 45 | 45 | 45 | 45 |

### Correlations

| | | Shuttleworthia_change | Subdoligranulum_change | UCG.003_change | X.Eubacterium..brachy.group_change | X.Ruminococcus..gauvreauii.group_change | X.Ruminococcus..torques.group_change |
|---|---|---|---|---|---|---|---|
| X.Ruminococcus..torques.group_change | Correlation Coefficient | -,069 | ,201 | -,349 | ,086 | -,016 | 1,000 |
| | Sig. (2-tailed) | ,652 | ,185 | ,019 | ,573 | ,915 | . |
| | N | 45 | 45 | 45 | 45 | 45 | 45 |

### Correlations

| | | | interaction_VTA_change | interaction_rOFC_change | interaction_rmOFC_change | sumscore_GIQLI_change | Stool_frequency_change | STGHRE_change |
|---|---|---|---|---|---|---|---|---|
| Spearman's rho | interaction_VTA_change | Correlation Coefficient | 1,000 | ,344* | ,414** | -,162 | -,159 | ,043 |
| | | Sig. (2-tailed) | . | ,028 | ,005 | ,299 | ,316 | ,793 |
| | | N | 44 | 41 | 44 | 43 | 42 | 40 |
| | interaction_rOFC_change | Correlation Coefficient | ,344* | 1,000 | ,410** | -,295 | ,014 | ,070 |
| | | Sig. (2-tailed) | ,028 | . | ,008 | ,065 | ,934 | ,681 |
| | | N | 41 | 41 | 41 | 40 | 39 | 37 |
| | interaction_rmOFC_change | Correlation Coefficient | ,414** | ,410** | 1,000 | ,010 | ,027 | ,253 |
| | | Sig. (2-tailed) | ,005 | ,008 | . | ,947 | ,867 | ,116 |
| | | N | 44 | 41 | 44 | 43 | 42 | 40 |
| | sumscore_GIQLI_change | Correlation Coefficient | -,162 | -,295 | ,010 | 1,000 | ,063 | ,054 |
| | | Sig. (2-tailed) | ,299 | ,065 | ,947 | . | ,683 | ,738 |
| | | N | 43 | 40 | 43 | 45 | 44 | 41 |
| | Stool_frequency_change | Correlation Coefficient | -,159 | ,014 | ,027 | ,063 | 1,000 | -,082 |
| | | Sig. (2-tailed) | ,316 | ,934 | ,867 | ,683 | . | ,615 |
| | | N | 42 | 39 | 42 | 44 | 44 | 40 |
| | STGHRE_change | Correlation Coefficient | ,043 | ,070 | ,253 | ,054 | -,082 | 1,000 |
| | | Sig. (2-tailed) | ,793 | ,681 | ,116 | ,738 | ,615 | . |
| | | N | 40 | 37 | 40 | 41 | 40 | 42 |
| | STPYY_change | Correlation Coefficient | ,439** | ,348* | ,318* | -,159 | -,047 | ,011 |
| | | Sig. (2-tailed) | ,005 | ,035 | ,046 | ,321 | ,774 | ,947 |
| | | N | 40 | 37 | 40 | 41 | 40 | 42 |
| | STGLPT_change | Correlation Coefficient | ,057 | ,192 | -,012 | ,117 | ,011 | -,193 |
| | | Sig. (2-tailed) | ,727 | ,254 | ,943 | ,468 | ,946 | ,221 |
| | | N | 40 | 37 | 40 | 41 | 40 | 42 |

### Correlations

| | | | STPYY_change | STGLPT_change | age | BMI | FM_stand | Richness | shannon. effective |
|---|---|---|---|---|---|---|---|---|---|
| Spearman's rho | interaction_VTA_change | Correlation Coefficient | ,439** | ,057 | -,028 | -,251 | ,104 | ,182 | ,160 |
| | | Sig. (2-tailed) | ,005 | ,727 | ,854 | ,100 | ,500 | ,237 | ,299 |
| | | N | 40 | 40 | 44 | 44 | 44 | 44 | 44 |
| | interaction_rOFC_change | Correlation Coefficient | ,348* | ,192 | ,385* | -,273 | -,088 | ,041 | ,020 |
| | | Sig. (2-tailed) | ,035 | ,254 | ,013 | ,084 | ,585 | ,797 | ,901 |
| | | N | 37 | 37 | 41 | 41 | 41 | 41 | 41 |
| | interaction_rmOFC_change | Correlation Coefficient | ,318* | -,012 | ,167 | -,174 | -,031 | -,116 | -,125 |
| | | Sig. (2-tailed) | ,046 | ,943 | ,279 | ,259 | ,839 | ,453 | ,418 |
| | | N | 40 | 40 | 44 | 44 | 44 | 44 | 44 |
| | sumscore_GIQLI_change | Correlation Coefficient | -,159 | ,117 | ,175 | ,155 | ,154 | ,181 | ,181 |
| | | Sig. (2-tailed) | ,321 | ,468 | ,251 | ,309 | ,312 | ,235 | ,234 |
| | | N | 41 | 41 | 45 | 45 | 45 | 45 | 45 |
| | Stool_frequency_change | Correlation Coefficient | -,047 | ,011 | -,127 | -,034 | -,242 | -,278 | -,259 |
| | | Sig. (2-tailed) | ,774 | ,946 | ,410 | ,829 | ,114 | ,067 | ,090 |
| | | N | 40 | 40 | 44 | 44 | 44 | 44 | 44 |
| | STGHRE_change | Correlation Coefficient | ,011 | -,193 | -,051 | -,174 | -,351* | -,072 | -,072 |
| | | Sig. (2-tailed) | ,947 | ,221 | ,748 | ,270 | ,023 | ,652 | ,652 |
| | | N | 42 | 42 | 42 | 42 | 42 | 42 | 42 |
| | STPYY_change | Correlation Coefficient | 1,000 | ,113 | -,020 | -,078 | ,124 | ,137 | ,141 |
| | | Sig. (2-tailed) | . | ,478 | ,902 | ,625 | ,433 | ,388 | ,374 |
| | | N | 42 | 42 | 42 | 42 | 42 | 42 | 42 |
| | STGLPT_change | Correlation Coefficient | ,113 | 1,000 | -,094 | ,131 | ,150 | ,127 | ,044 |
| | | Sig. (2-tailed) | ,478 | . | ,556 | ,408 | ,344 | ,423 | ,780 |
| | | N | 42 | 42 | 42 | 42 | 42 | 42 | 42 |

### Correlations

| | | | simpson. effective | Evenness | CHOL_change | LDL_change | shannon. effective_change | Richness_change |
|---|---|---|---|---|---|---|---|---|
| Spearman's rho | interaction_VTA_change | Correlation Coefficient | ,081 | -,185 | -,066 | -,080 | ,241 | ,154 |
| | | Sig. (2-tailed) | ,603 | ,229 | ,669 | ,607 | ,120 | ,323 |
| | | N | 44 | 44 | 44 | 44 | 43 | 43 |
| | interaction_rOFC_change | Correlation Coefficient | -,039 | -,136 | ,041 | ,057 | -,253 | -,211 |
| | | Sig. (2-tailed) | ,811 | ,398 | ,800 | ,721 | ,115 | ,191 |
| | | N | 41 | 41 | 41 | 41 | 40 | 40 |
| | interaction_rmOFC_change | Correlation Coefficient | -,130 | -,124 | -,086 | -,199 | -,154 | -,177 |
| | | Sig. (2-tailed) | ,400 | ,422 | ,578 | ,196 | ,324 | ,256 |
| | | N | 44 | 44 | 44 | 44 | 43 | 43 |
| | sumscore_GIQLI_change | Correlation Coefficient | ,253 | ,071 | -,157 | -,079 | ,154 | ,122 |
| | | Sig. (2-tailed) | ,094 | ,644 | ,304 | ,608 | ,319 | ,431 |
| | | N | 45 | 45 | 45 | 45 | 44 | 44 |
| | Stool_frequency_change | Correlation Coefficient | -,216 | ,040 | -,200 | -,075 | -,270 | -,326* |
| | | Sig. (2-tailed) | ,159 | ,797 | ,194 | ,631 | ,080 | ,033 |
| | | N | 44 | 44 | 44 | 44 | 43 | 43 |
| | STGHRE_change | Correlation Coefficient | -,017 | -,022 | -,506** | -,532** | ,007 | -,070 |
| | | Sig. (2-tailed) | ,914 | ,889 | ,001 | ,000 | ,964 | ,659 |
| | | N | 42 | 42 | 42 | 42 | 42 | 42 |
| | STPYY_change | Correlation Coefficient | ,090 | -,060 | ,083 | ,080 | -,089 | -,204 |
| | | Sig. (2-tailed) | ,570 | ,706 | ,600 | ,615 | ,575 | ,196 |
| | | N | 42 | 42 | 42 | 42 | 42 | 42 |
| | STGLPT_change | Correlation Coefficient | -,017 | -,131 | ,230 | ,233 | ,014 | -,014 |
| | | Sig. (2-tailed) | ,917 | ,408 | ,143 | ,137 | ,928 | ,931 |
| | | N | 42 | 42 | 42 | 42 | 42 | 42 |

### Correlations

| | | | Evenness_change | Actinomyces_change | Aerostipes_change | Bifidobacterium_change | Blautia_change | Collinsella_change |
|---|---|---|---|---|---|---|---|---|
| Spearman's rho | interaction_VTA_change | Correlation Coefficient | ,112 | -,042 | -,264 | -,152 | -,238 | ,024 |
| | | Sig. (2-tailed) | ,473 | ,791 | ,088 | ,332 | ,124 | ,878 |
| | | N | 43 | 43 | 43 | 43 | 43 | 43 |
| | interaction_rOFC_change | Correlation Coefficient | -,227 | ,268 | -,206 | -,136 | ,179 | ,178 |
| | | Sig. (2-tailed) | ,159 | ,094 | ,201 | ,401 | ,270 | ,271 |
| | | N | 40 | 40 | 40 | 40 | 40 | 40 |
| | interaction_rmOFC_change | Correlation Coefficient | -,045 | ,173 | ,039 | ,098 | ,084 | ,150 |
| | | Sig. (2-tailed) | ,772 | ,268 | ,806 | ,533 | ,594 | ,339 |
| | | N | 43 | 43 | 43 | 43 | 43 | 43 |
| | sumscore_GIQLI_change | Correlation Coefficient | ,171 | -,130 | ,021 | -,109 | ,166 | -,202 |
| | | Sig. (2-tailed) | ,266 | ,401 | ,892 | ,482 | ,283 | ,189 |
| | | N | 44 | 44 | 44 | 44 | 44 | 44 |
| | Stool_frequency_change | Correlation Coefficient | ,106 | -,006 | ,325* | -,125 | ,115 | ,151 |
| | | Sig. (2-tailed) | ,498 | ,972 | ,034 | ,425 | ,465 | ,333 |
| | | N | 43 | 43 | 43 | 43 | 43 | 43 |
| | STGHRE_change | Correlation Coefficient | ,124 | ,008 | -,040 | -,225 | ,163 | ,014 |
| | | Sig. (2-tailed) | ,433 | ,958 | ,803 | ,151 | ,301 | ,930 |
| | | N | 42 | 42 | 42 | 42 | 42 | 42 |
| | STPYY_change | Correlation Coefficient | ,122 | ,084 | ,001 | ,012 | -,021 | ,176 |
| | | Sig. (2-tailed) | ,442 | ,599 | ,995 | ,940 | ,894 | ,266 |
| | | N | 42 | 42 | 42 | 42 | 42 | 42 |
| | STGLPT_change | Correlation Coefficient | ,073 | -,030 | ,067 | ,014 | ,209 | ,004 |
| | | Sig. (2-tailed) | ,644 | ,852 | ,673 | ,929 | ,184 | ,978 |
| | | N | 42 | 42 | 42 | 42 | 42 | 42 |

### Correlations

| | | | Desulfovibrio_change | Eggerthella_change | Erysipelatoclostridium_change | Faecalitalea_change | Family.XIII.AD3011.group_change | Family.XIII.UCG.001_change |
|---|---|---|---|---|---|---|---|---|
| Spearman's rho | interaction_VTA_change | Correlation Coefficient | ,034 | ,020 | ,159 | -,099 | ,121 | -,080 |
| | | Sig. (2-tailed) | ,830 | ,899 | ,309 | ,526 | ,440 | ,611 |
| | | N | 43 | 43 | 43 | 43 | 43 | 43 |
| | interaction_rOFC_change | Correlation Coefficient | -,009 | ,022 | -,021 | -,018 | ,191 | ,064 |
| | | Sig. (2-tailed) | ,955 | ,892 | ,900 | ,911 | ,238 | ,696 |
| | | N | 40 | 40 | 40 | 40 | 40 | 40 |
| | interaction_rmOFC_change | Correlation Coefficient | -,084 | -,027 | -,063 | ,035 | ,070 | -,206 |
| | | Sig. (2-tailed) | ,591 | ,862 | ,688 | ,825 | ,655 | ,185 |
| | | N | 43 | 43 | 43 | 43 | 43 | 43 |
| | sumscore_GIQLI_change | Correlation Coefficient | ,059 | ,104 | ,190 | ,038 | ,180 | ,009 |
| | | Sig. (2-tailed) | ,704 | ,503 | ,216 | ,809 | ,241 | ,953 |
| | | N | 44 | 44 | 44 | 44 | 44 | 44 |
| | Stool_frequency_change | Correlation Coefficient | ,066 | -,131 | -,291 | -,040 | ,250 | -,054 |
| | | Sig. (2-tailed) | ,674 | ,403 | ,058 | ,798 | ,105 | ,731 |
| | | N | 43 | 43 | 43 | 43 | 43 | 43 |
| | STGHRE_change | Correlation Coefficient | -,095 | -,126 | -,075 | ,166 | ,125 | ,215 |
| | | Sig. (2-tailed) | ,551 | ,425 | ,639 | ,294 | ,430 | ,171 |
| | | N | 42 | 42 | 42 | 42 | 42 | 42 |
| | STPYY_change | Correlation Coefficient | -,370* | ,032 | -,017 | ,011 | ,171 | ,017 |
| | | Sig. (2-tailed) | ,016 | ,839 | ,913 | ,945 | ,279 | ,913 |
| | | N | 42 | 42 | 42 | 42 | 42 | 42 |
| | STGLPT_change | Correlation Coefficient | -,069 | ,103 | ,296 | ,090 | ,058 | ,205 |
| | | Sig. (2-tailed) | ,666 | ,518 | ,057 | ,570 | ,717 | ,192 |
| | | N | 42 | 42 | 42 | 42 | 42 | 42 |

### Correlations

| | | | Gemella_change | Gordonibacter_change | Holdemanella_change | Holdemania_change | Lachnospiraceae.FCS020.group_change | Lachnospiraceae.NK4A136.group_change |
|---|---|---|---|---|---|---|---|---|
| Spearman's rho | interaction_VTA_change | Correlation Coefficient | -,174 | ,025 | -,083 | -,116 | -,041 | ,286 |
| | | Sig. (2-tailed) | ,264 | ,876 | ,597 | ,459 | ,792 | ,063 |
| | | N | 43 | 43 | 43 | 43 | 43 | 43 |
| | interaction_rOFC_change | Correlation Coefficient | ,295 | ,044 | ,049 | -,259 | -,079 | -,093 |
| | | Sig. (2-tailed) | ,064 | ,785 | ,764 | ,107 | ,627 | ,566 |
| | | N | 40 | 40 | 40 | 40 | 40 | 40 |
| | interaction_rmOFC_change | Correlation Coefficient | ,196 | ,079 | ,005 | -,195 | ,028 | -,001 |
| | | Sig. (2-tailed) | ,209 | ,614 | ,972 | ,211 | ,860 | ,996 |
| | | N | 43 | 43 | 43 | 43 | 43 | 43 |
| | sumscore_GIQLI_change | Correlation Coefficient | ,086 | ,079 | ,160 | ,095 | ,133 | ,000 |
| | | Sig. (2-tailed) | ,577 | ,610 | ,301 | ,539 | ,390 | ,998 |
| | | N | 44 | 44 | 44 | 44 | 44 | 44 |
| | Stool_frequency_change | Correlation Coefficient | -,119 | -,099 | -,104 | -,016 | -,071 | -,013 |
| | | Sig. (2-tailed) | ,445 | ,526 | ,505 | ,919 | ,649 | ,933 |
| | | N | 43 | 43 | 43 | 43 | 43 | 43 |
| | STGHRE_change | Correlation Coefficient | -,105 | -,040 | -,104 | ,015 | -,078 | ,094 |
| | | Sig. (2-tailed) | ,510 | ,803 | ,514 | ,927 | ,624 | ,554 |
| | | N | 42 | 42 | 42 | 42 | 42 | 42 |
| | STPYY_change | Correlation Coefficient | -,139 | -,341* | -,043 | -,157 | ,076 | ,155 |
| | | Sig. (2-tailed) | ,381 | ,027 | ,785 | ,321 | ,633 | ,326 |
| | | N | 42 | 42 | 42 | 42 | 42 | 42 |
| | STGLPT_change | Correlation Coefficient | ,150 | ,132 | ,166 | -,266 | ,162 | -,079 |
| | | Sig. (2-tailed) | ,342 | ,404 | ,294 | ,089 | ,307 | ,621 |
| | | N | 42 | 42 | 42 | 42 | 42 | 42 |

### Correlations

| | | | Lactiplantibacillus_change | Lactobacillus_change | Libanicoccus_change | Ligilactobacillus_change | Limosilactobacillus_change | Roseburia_change |
|---|---|---|---|---|---|---|---|---|
| Spearman's rho | interaction_VTA_change | Correlation Coefficient | -,006 | -,023 | ,294 | -,175 | -,010 | ,137 |
| | | Sig. (2-tailed) | ,972 | ,882 | ,056 | ,262 | ,949 | ,380 |
| | | N | 43 | 43 | 43 | 43 | 43 | 43 |
| | interaction_rOFC_change | Correlation Coefficient | ,120 | ,046 | ,211 | -,105 | -,005 | ,146 |
| | | Sig. (2-tailed) | ,459 | ,778 | ,192 | ,521 | ,976 | ,369 |
| | | N | 40 | 40 | 40 | 40 | 40 | 40 |
| | interaction_rmOFC_change | Correlation Coefficient | ,397** | -,054 | ,223 | -,082 | -,070 | -,011 |
| | | Sig. (2-tailed) | ,008 | ,731 | ,151 | ,603 | ,654 | ,944 |
| | | N | 43 | 43 | 43 | 43 | 43 | 43 |
| | sumscore_GIQLI_change | Correlation Coefficient | ,168 | -,033 | -,057 | ,054 | ,058 | ,025 |
| | | Sig. (2-tailed) | ,276 | ,833 | ,712 | ,727 | ,706 | ,870 |
| | | N | 44 | 44 | 44 | 44 | 44 | 44 |
| | Stool_frequency_change | Correlation Coefficient | -,007 | ,105 | -,187 | ,060 | ,169 | -,316* |
| | | Sig. (2-tailed) | ,966 | ,504 | ,230 | ,703 | ,278 | ,039 |
| | | N | 43 | 43 | 43 | 43 | 43 | 43 |
| | STGHRE_change | Correlation Coefficient | ,131 | -,070 | -,107 | -,017 | -,196 | -,209 |
| | | Sig. (2-tailed) | ,407 | ,659 | ,499 | ,914 | ,212 | ,184 |
| | | N | 42 | 42 | 42 | 42 | 42 | 42 |
| | STPYY_change | Correlation Coefficient | -,079 | ,046 | ,039 | ,015 | ,027 | -,048 |
| | | Sig. (2-tailed) | ,618 | ,771 | ,807 | ,924 | ,864 | ,763 |
| | | N | 42 | 42 | 42 | 42 | 42 | 42 |
| | STGLPT_change | Correlation Coefficient | -,186 | -,181 | ,115 | ,038 | -,182 | -,035 |
| | | Sig. (2-tailed) | ,238 | ,251 | ,468 | ,813 | ,248 | ,824 |
| | | N | 42 | 42 | 42 | 42 | 42 | 42 |

### Correlations

| | | | Shuttleworthia_change | Subdoligranulum_change | UCG.003_change | X.Eubacterium..brachy.group_change | X.Ruminococcus..gauvreauii.group_change | X.Ruminococcus..torques.group_change |
|---|---|---|---|---|---|---|---|---|
| Spearman's rho | interaction_VTA_change | Correlation Coefficient | ,191 | -,384* | ,185 | ,115 | -,194 | -,229 |
| | | Sig. (2-tailed) | ,220 | ,011 | ,236 | ,464 | ,211 | ,140 |
| | | N | 43 | 43 | 43 | 43 | 43 | 43 |
| | interaction_rOFC_change | Correlation Coefficient | -,155 | -,159 | ,122 | -,177 | -,143 | -,194 |
| | | Sig. (2-tailed) | ,338 | ,329 | ,453 | ,275 | ,377 | ,230 |
| | | N | 40 | 40 | 40 | 40 | 40 | 40 |
| | interaction_rmOFC_change | Correlation Coefficient | ,032 | -,015 | -,094 | ,038 | -,024 | -,133 |
| | | Sig. (2-tailed) | ,838 | ,922 | ,551 | ,809 | ,879 | ,397 |
| | | N | 43 | 43 | 43 | 43 | 43 | 43 |
| | sumscore_GIQLI_change | Correlation Coefficient | ,210 | ,261 | -,255 | ,090 | ,113 | ,305* |
| | | Sig. (2-tailed) | ,172 | ,087 | ,095 | ,561 | ,465 | ,044 |
| | | N | 44 | 44 | 44 | 44 | 44 | 44 |
| | Stool_frequency_change | Correlation Coefficient | ,115 | ,083 | -,067 | -,196 | -,091 | ,271 |
| | | Sig. (2-tailed) | ,462 | ,598 | ,669 | ,209 | ,560 | ,079 |
| | | N | 43 | 43 | 43 | 43 | 43 | 43 |
| | STGHRE_change | Correlation Coefficient | -,164 | ,279 | -,059 | -,097 | -,100 | ,230 |
| | | Sig. (2-tailed) | ,300 | ,073 | ,711 | ,540 | ,528 | ,144 |
| | | N | 42 | 42 | 42 | 42 | 42 | 42 |
| | STPYY_change | Correlation Coefficient | -,068 | -,210 | -,176 | -,061 | -,063 | ,183 |
| | | Sig. (2-tailed) | ,669 | ,182 | ,264 | ,702 | ,693 | ,247 |
| | | N | 42 | 42 | 42 | 42 | 42 | 42 |
| | STGLPT_change | Correlation Coefficient | -,074 | ,009 | -,165 | ,179 | -,159 | ,092 |
| | | Sig. (2-tailed) | ,644 | ,957 | ,296 | ,256 | ,314 | ,561 |
| | | N | 42 | 42 | 42 | 42 | 42 | 42 |

### Correlations

| | | interaction_VTA_change | interaction_rOFC_change | interaction_rmOFC_change | sumscore_GIQLI_change | Stool_frequency_change | STGHRE_change |
|---|---|---|---|---|---|---|---|
| age | Correlation Coefficient | -,028 | ,385* | ,167 | ,175 | -,127 | -,051 |
| | Sig. (2-tailed) | ,854 | ,013 | ,279 | ,251 | ,410 | ,748 |
| | N | 44 | 41 | 44 | 45 | 44 | 42 |
| BMI | Correlation Coefficient | -,251 | -,273 | -,174 | ,155 | -,034 | -,174 |
| | Sig. (2-tailed) | ,100 | ,084 | ,259 | ,309 | ,829 | ,270 |
| | N | 44 | 41 | 44 | 45 | 44 | 42 |
| FM_stand | Correlation Coefficient | ,104 | -,088 | -,031 | ,154 | -,242 | -,351* |
| | Sig. (2-tailed) | ,500 | ,585 | ,839 | ,312 | ,114 | ,023 |
| | N | 44 | 41 | 44 | 45 | 44 | 42 |
| Richness | Correlation Coefficient | ,182 | ,041 | -,116 | ,181 | -,278 | -,072 |
| | Sig. (2-tailed) | ,237 | ,797 | ,453 | ,235 | ,067 | ,652 |
| | N | 44 | 41 | 44 | 45 | 44 | 42 |
| shannon.effective | Correlation Coefficient | ,160 | ,020 | -,125 | ,181 | -,259 | -,072 |
| | Sig. (2-tailed) | ,299 | ,901 | ,418 | ,234 | ,090 | ,652 |
| | N | 44 | 41 | 44 | 45 | 44 | 42 |
| simpson.effective | Correlation Coefficient | ,081 | -,039 | -,130 | ,253 | -,216 | -,017 |
| | Sig. (2-tailed) | ,603 | ,811 | ,400 | ,094 | ,159 | ,914 |
| | N | 44 | 41 | 44 | 45 | 44 | 42 |
| Evenness | Correlation Coefficient | -,185 | -,136 | -,124 | ,071 | ,040 | -,022 |
| | Sig. (2-tailed) | ,229 | ,398 | ,422 | ,644 | ,797 | ,889 |
| | N | 44 | 41 | 44 | 45 | 44 | 42 |
| CHOL_change | Correlation Coefficient | -,066 | ,041 | -,086 | -,157 | -,200 | -,506** |
| | Sig. (2-tailed) | ,669 | ,800 | ,578 | ,304 | ,194 | ,001 |
| | N | 44 | 41 | 44 | 45 | 44 | 42 |

### Correlations

| | | STPYY_change | STGLPT_change | age | BMI | FM_stand | Richness | shannon.effective |
|---|---|---|---|---|---|---|---|---|
| age | Correlation Coefficient | -,020 | -,094 | 1,000 | -,041 | -,055 | ,126 | ,065 |
| | Sig. (2-tailed) | ,902 | ,556 | . | ,789 | ,718 | ,402 | ,670 |
| | N | 42 | 42 | 46 | 46 | 46 | 46 | 46 |
| BMI | Correlation Coefficient | -,078 | ,131 | -,041 | 1,000 | ,463** | ,163 | ,111 |
| | Sig. (2-tailed) | ,625 | ,408 | ,789 | . | ,001 | ,278 | ,463 |
| | N | 42 | 42 | 46 | 46 | 46 | 46 | 46 |
| FM_stand | Correlation Coefficient | ,124 | ,150 | -,055 | ,463** | 1,000 | ,030 | -,027 |
| | Sig. (2-tailed) | ,433 | ,344 | ,718 | ,001 | . | ,842 | ,856 |
| | N | 42 | 42 | 46 | 46 | 46 | 46 | 46 |
| Richness | Correlation Coefficient | ,137 | ,127 | ,126 | ,163 | ,030 | 1,000 | ,949** |
| | Sig. (2-tailed) | ,388 | ,423 | ,402 | ,278 | ,842 | . | ,000 |
| | N | 42 | 42 | 46 | 46 | 46 | 46 | 46 |
| shannon.effective | Correlation Coefficient | ,141 | ,044 | ,065 | ,111 | -,027 | ,949** | 1,000 |
| | Sig. (2-tailed) | ,374 | ,780 | ,670 | ,463 | ,856 | ,000 | . |
| | N | 42 | 42 | 46 | 46 | 46 | 46 | 46 |
| simpson.effective | Correlation Coefficient | ,090 | -,017 | ,058 | ,068 | -,071 | ,886** | ,965** |
| | Sig. (2-tailed) | ,570 | ,917 | ,702 | ,654 | ,639 | ,000 | ,000 |
| | N | 42 | 42 | 46 | 46 | 46 | 46 | 46 |
| Evenness | Correlation Coefficient | -,060 | -,131 | -,096 | -,029 | -,094 | ,153 | ,405** |
| | Sig. (2-tailed) | ,706 | ,408 | ,524 | ,850 | ,536 | ,310 | ,005 |
| | N | 42 | 42 | 46 | 46 | 46 | 46 | 46 |
| CHOL_change | Correlation Coefficient | ,083 | ,230 | ,077 | ,238 | ,513** | ,010 | -,067 |
| | Sig. (2-tailed) | ,600 | ,143 | ,611 | ,111 | ,000 | ,948 | ,658 |
| | N | 42 | 42 | 46 | 46 | 46 | 46 | 46 |

### Correlations

| | | simpson.effective | Evenness | CHOL_change | LDL_change | shannon.effective_change | Richness_change |
|---|---|---|---|---|---|---|---|
| age | Correlation Coefficient | ,058 | -,096 | ,077 | ,052 | -,019 | ,089 |
| | Sig. (2-tailed) | ,702 | ,524 | ,611 | ,733 | ,903 | ,559 |
| | N | 46 | 46 | 46 | 46 | 45 | 45 |
| BMI | Correlation Coefficient | ,068 | -,029 | ,238 | ,202 | -,130 | -,105 |
| | Sig. (2-tailed) | ,654 | ,850 | ,111 | ,179 | ,395 | ,492 |
| | N | 46 | 46 | 46 | 46 | 45 | 45 |
| FM_stand | Correlation Coefficient | -,071 | -,094 | ,513** | ,478** | ,014 | ,167 |
| | Sig. (2-tailed) | ,639 | ,536 | ,000 | ,001 | ,927 | ,274 |
| | N | 46 | 46 | 46 | 46 | 45 | 45 |
| Richness | Correlation Coefficient | ,886** | ,153 | ,010 | -,042 | ,490** | ,500** |
| | Sig. (2-tailed) | ,000 | ,310 | ,948 | ,781 | ,001 | ,000 |
| | N | 46 | 46 | 46 | 46 | 45 | 45 |
| shannon.effective | Correlation Coefficient | ,965** | ,405** | -,067 | -,119 | ,570** | ,512** |
| | Sig. (2-tailed) | ,000 | ,005 | ,658 | ,430 | ,000 | ,000 |
| | N | 46 | 46 | 46 | 46 | 45 | 45 |
| simpson.effective | Correlation Coefficient | 1,000 | ,491** | -,114 | -,149 | ,586** | ,500** |
| | Sig. (2-tailed) | . | ,001 | ,449 | ,322 | ,000 | ,000 |
| | N | 46 | 46 | 46 | 46 | 45 | 45 |
| Evenness | Correlation Coefficient | ,491** | 1,000 | -,053 | -,082 | ,347* | ,163 |
| | Sig. (2-tailed) | ,001 | . | ,726 | ,588 | ,020 | ,285 |
| | N | 46 | 46 | 46 | 46 | 45 | 45 |
| CHOL_change | Correlation Coefficient | -,114 | -,053 | 1,000 | ,926** | -,144 | ,038 |
| | Sig. (2-tailed) | ,449 | ,726 | . | ,000 | ,344 | ,806 |
| | N | 46 | 46 | 46 | 46 | 45 | 45 |

### Correlations

| | | Evenness_change | Actinomyces_change | Aerostipes_change | Bifidobacterium_change | Blautia_change | Collinsella_change |
|---|---|---|---|---|---|---|---|
| age | Correlation Coefficient | -,205 | -,004 | -,078 | -,044 | ,083 | -,228 |
| | Sig. (2-tailed) | ,177 | ,979 | ,611 | ,774 | ,588 | ,133 |
| | N | 45 | 45 | 45 | 45 | 45 | 45 |
| BMI | Correlation Coefficient | -,086 | -,121 | ,105 | ,013 | -,132 | -,035 |
| | Sig. (2-tailed) | ,575 | ,429 | ,491 | ,933 | ,387 | ,820 |
| | N | 45 | 45 | 45 | 45 | 45 | 45 |
| FM_stand | Correlation Coefficient | -,303* | ,085 | -,157 | ,323* | -,159 | ,013 |
| | Sig. (2-tailed) | ,043 | ,577 | ,305 | ,031 | ,296 | ,932 |
| | N | 45 | 45 | 45 | 45 | 45 | 45 |
| Richness | Correlation Coefficient | ,137 | -,028 | ,011 | -,514** | -,145 | -,354* |
| | Sig. (2-tailed) | ,370 | ,856 | ,943 | ,000 | ,341 | ,017 |
| | N | 45 | 45 | 45 | 45 | 45 | 45 |
| shannon.effective | Correlation Coefficient | ,306* | -,017 | ,036 | -,582** | -,055 | -,322* |
| | Sig. (2-tailed) | ,041 | ,912 | ,816 | ,000 | ,722 | ,031 |
| | N | 45 | 45 | 45 | 45 | 45 | 45 |
| simpson.effective | Correlation Coefficient | ,373* | -,085 | ,058 | -,626** | -,032 | -,369* |
| | Sig. (2-tailed) | ,012 | ,577 | ,703 | ,000 | ,833 | ,012 |
| | N | 45 | 45 | 45 | 45 | 45 | 45 |
| Evenness | Correlation Coefficient | ,600** | ,010 | ,155 | -,401** | ,355* | -,060 |
| | Sig. (2-tailed) | ,000 | ,949 | ,309 | ,006 | ,017 | ,697 |
| | N | 45 | 45 | 45 | 45 | 45 | 45 |
| CHOL_change | Correlation Coefficient | -,308* | ,118 | -,260 | ,153 | -,016 | ,110 |
| | Sig. (2-tailed) | ,039 | ,441 | ,084 | ,316 | ,914 | ,474 |
| | N | 45 | 45 | 45 | 45 | 45 | 45 |

### Correlations

| | | | Desulfovibrio_change | Eggerthella_change | Erysipelatoclostridium_change | Faecalitalea_change | Family.XIII. AD3011. group_change | Family.XIII. UCG. 001_change |
|---|---|---|---|---|---|---|---|---|
| | age | Correlation Coefficient | ,135 | ,151 | ,005 | -,041 | -,057 | ,067 |
| | | Sig. (2-tailed) | ,378 | ,322 | ,973 | ,788 | ,711 | ,663 |
| | | N | 45 | 45 | 45 | 45 | 45 | 45 |
| | BMI | Correlation Coefficient | -,268 | ,107 | ,250 | ,041 | ,033 | -,199 |
| | | Sig. (2-tailed) | ,075 | ,485 | ,097 | ,791 | ,831 | ,190 |
| | | N | 45 | 45 | 45 | 45 | 45 | 45 |
| | FM_stand | Correlation Coefficient | -,284 | ,228 | ,207 | ,055 | -,024 | -,306* |
| | | Sig. (2-tailed) | ,059 | ,131 | ,172 | ,721 | ,876 | ,041 |
| | | N | 45 | 45 | 45 | 45 | 45 | 45 |
| | Richness | Correlation Coefficient | -,048 | ,118 | ,120 | ,011 | ,350* | ,256 |
| | | Sig. (2-tailed) | ,753 | ,438 | ,433 | ,942 | ,018 | ,090 |
| | | N | 45 | 45 | 45 | 45 | 45 | 45 |
| | shannon.effective | Correlation Coefficient | -,048 | ,104 | ,112 | ,026 | ,325* | ,236 |
| | | Sig. (2-tailed) | ,755 | ,498 | ,464 | ,864 | ,029 | ,119 |
| | | N | 45 | 45 | 45 | 45 | 45 | 45 |
| | simpson.effective | Correlation Coefficient | -,059 | ,084 | ,126 | ,041 | ,293 | ,240 |
| | | Sig. (2-tailed) | ,699 | ,583 | ,408 | ,788 | ,051 | ,113 |
| | | N | 45 | 45 | 45 | 45 | 45 | 45 |
| | Evenness | Correlation Coefficient | -,091 | ,081 | -,016 | ,117 | ,022 | -,001 |
| | | Sig. (2-tailed) | ,553 | ,595 | ,919 | ,443 | ,885 | ,996 |
| | | N | 45 | 45 | 45 | 45 | 45 | 45 |
| | CHOL_change | Correlation Coefficient | -,270 | ,203 | ,152 | -,149 | -,174 | -,319* |
| | | Sig. (2-tailed) | ,073 | ,181 | ,318 | ,328 | ,253 | ,033 |
| | | N | 45 | 45 | 45 | 45 | 45 | 45 |

### Correlations

| | | Gemella_change | Gordonibacter_change | Holdemanella_change | Holdemania_change | Lachnospiraceae.FCS020.group_change | Lachnospiraceae.NK4A136.group_change |
|---|---|---|---|---|---|---|---|
| age | Correlation Coefficient | ,200 | ,239 | -,098 | ,177 | -,187 | ,152 |
| | Sig. (2-tailed) | ,187 | ,114 | ,523 | ,245 | ,220 | ,319 |
| | N | 45 | 45 | 45 | 45 | 45 | 45 |
| BMI | Correlation Coefficient | -,204 | ,173 | ,079 | ,284 | ,056 | ,109 |
| | Sig. (2-tailed) | ,180 | ,257 | ,606 | ,058 | ,713 | ,476 |
| | N | 45 | 45 | 45 | 45 | 45 | 45 |
| FM_stand | Correlation Coefficient | -,199 | ,090 | ,056 | ,017 | -,044 | ,114 |
| | Sig. (2-tailed) | ,189 | ,558 | ,713 | ,911 | ,774 | ,454 |
| | N | 45 | 45 | 45 | 45 | 45 | 45 |
| Richness | Correlation Coefficient | -,070 | ,244 | ,303* | ,395** | ,238 | ,177 |
| | Sig. (2-tailed) | ,648 | ,107 | ,043 | ,007 | ,115 | ,246 |
| | N | 45 | 45 | 45 | 45 | 45 | 45 |
| shannon.effective | Correlation Coefficient | -,050 | ,257 | ,318* | ,414** | ,193 | ,117 |
| | Sig. (2-tailed) | ,745 | ,089 | ,033 | ,005 | ,204 | ,443 |
| | N | 45 | 45 | 45 | 45 | 45 | 45 |
| simpson.effective | Correlation Coefficient | -,066 | ,266 | ,357* | ,438** | ,205 | ,108 |
| | Sig. (2-tailed) | ,667 | ,077 | ,016 | ,003 | ,178 | ,480 |
| | N | 45 | 45 | 45 | 45 | 45 | 45 |
| Evenness | Correlation Coefficient | -,034 | ,248 | ,092 | ,206 | -,019 | -,153 |
| | Sig. (2-tailed) | ,824 | ,101 | ,546 | ,175 | ,903 | ,316 |
| | N | 45 | 45 | 45 | 45 | 45 | 45 |
| CHOL_change | Correlation Coefficient | ,155 | ,192 | ,137 | -,201 | ,182 | ,013 |
| | Sig. (2-tailed) | ,311 | ,205 | ,369 | ,186 | ,232 | ,933 |
| | N | 45 | 45 | 45 | 45 | 45 | 45 |

### Correlations

| | | Lactiplantibacillus_change | Lactobacillus_change | Libanicoccus_change | Ligilactobacillus_change | Limosilactobacillus_change | Roseburia_change |
|---|---|---|---|---|---|---|---|
| age | Correlation Coefficient | -,003 | ,190 | ,250 | -,220 | ,280 | ,323* |
| | Sig. (2-tailed) | ,986 | ,212 | ,098 | ,146 | ,062 | ,030 |
| | N | 45 | 45 | 45 | 45 | 45 | 45 |
| BMI | Correlation Coefficient | -,024 | -,043 | ,042 | ,072 | -,257 | -,120 |
| | Sig. (2-tailed) | ,874 | ,777 | ,786 | ,637 | ,088 | ,434 |
| | N | 45 | 45 | 45 | 45 | 45 | 45 |
| FM_stand | Correlation Coefficient | ,182 | ,118 | -,048 | -,144 | -,212 | ,182 |
| | Sig. (2-tailed) | ,231 | ,440 | ,753 | ,345 | ,162 | ,232 |
| | N | 45 | 45 | 45 | 45 | 45 | 45 |
| Richness | Correlation Coefficient | -,214 | -,278 | ,110 | -,045 | -,141 | ,123 |
| | Sig. (2-tailed) | ,159 | ,065 | ,470 | ,768 | ,354 | ,419 |
| | N | 45 | 45 | 45 | 45 | 45 | 45 |
| shannon.effective | Correlation Coefficient | -,256 | -,322* | ,108 | ,004 | -,098 | ,096 |
| | Sig. (2-tailed) | ,090 | ,031 | ,481 | ,979 | ,523 | ,532 |
| | N | 45 | 45 | 45 | 45 | 45 | 45 |
| simpson.effective | Correlation Coefficient | -,284 | -,340* | ,007 | ,017 | -,108 | ,054 |
| | Sig. (2-tailed) | ,059 | ,022 | ,963 | ,914 | ,480 | ,726 |
| | N | 45 | 45 | 45 | 45 | 45 | 45 |
| Evenness | Correlation Coefficient | -,172 | -,226 | -,031 | ,145 | ,031 | -,126 |
| | Sig. (2-tailed) | ,258 | ,135 | ,837 | ,342 | ,839 | ,409 |
| | N | 45 | 45 | 45 | 45 | 45 | 45 |
| CHOL_change | Correlation Coefficient | -,052 | ,184 | -,032 | -,061 | -,076 | ,028 |
| | Sig. (2-tailed) | ,735 | ,225 | ,834 | ,689 | ,622 | ,857 |
| | N | 45 | 45 | 45 | 45 | 45 | 45 |

### Correlations

| | | Shuttleworthia_change | Subdoligranulum_change | UCG.003_change | X.Eubacterium..brachy.group_change | X.Ruminococcus..gauvreauii.group_change | X.Ruminococcus..torques.group_change |
|---|---|---|---|---|---|---|---|
| age | Correlation Coefficient | -,034 | -,033 | ,072 | -,090 | ,071 | -,302* |
| | Sig. (2-tailed) | ,825 | ,827 | ,638 | ,554 | ,642 | ,044 |
| | N | 45 | 45 | 45 | 45 | 45 | 45 |
| BMI | Correlation Coefficient | ,016 | ,175 | ,239 | -,048 | -,050 | ,053 |
| | Sig. (2-tailed) | ,915 | ,252 | ,113 | ,753 | ,744 | ,728 |
| | N | 45 | 45 | 45 | 45 | 45 | 45 |
| FM_stand | Correlation Coefficient | ,312* | -,188 | ,208 | ,150 | ,196 | ,019 |
| | Sig. (2-tailed) | ,037 | ,216 | ,170 | ,324 | ,196 | ,904 |
| | N | 45 | 45 | 45 | 45 | 45 | 45 |
| Richness | Correlation Coefficient | -,034 | -,033 | ,052 | ,244 | -,346* | ,079 |
| | Sig. (2-tailed) | ,826 | ,831 | ,736 | ,106 | ,020 | ,607 |
| | N | 45 | 45 | 45 | 45 | 45 | 45 |
| shannon.effective | Correlation Coefficient | -,090 | ,008 | ,083 | ,227 | -,288 | ,122 |
| | Sig. (2-tailed) | ,556 | ,956 | ,587 | ,133 | ,055 | ,425 |
| | N | 45 | 45 | 45 | 45 | 45 | 45 |
| simpson.effective | Correlation Coefficient | -,080 | ,101 | ,035 | ,229 | -,249 | ,163 |
| | Sig. (2-tailed) | ,603 | ,511 | ,818 | ,130 | ,099 | ,284 |
| | N | 45 | 45 | 45 | 45 | 45 | 45 |
| Evenness | Correlation Coefficient | -,186 | ,322* | ,030 | ,073 | ,176 | ,204 |
| | Sig. (2-tailed) | ,222 | ,031 | ,844 | ,632 | ,247 | ,179 |
| | N | 45 | 45 | 45 | 45 | 45 | 45 |
| CHOL_change | Correlation Coefficient | ,155 | -,110 | ,006 | ,000 | ,241 | -,067 |
| | Sig. (2-tailed) | ,308 | ,473 | ,971 | 1,000 | ,110 | ,663 |
| | N | 45 | 45 | 45 | 45 | 45 | 45 |

### Correlations

| | | interaction_VTA_change | interaction_rOFC_change | interaction_rmOFC_change | sumscore_GIQLI_change | Stool_frequency_change | STGHRE_change |
|---|---|---|---|---|---|---|---|
| LDL_change | Correlation Coefficient | -,080 | ,057 | -,199 | -,079 | -,075 | -,532** |
| | Sig. (2-tailed) | ,607 | ,721 | ,196 | ,608 | ,631 | ,000 |
| | N | 44 | 41 | 44 | 45 | 44 | 42 |
| shannon.effective_change | Correlation Coefficient | ,241 | -,253 | -,154 | ,154 | -,270 | ,007 |
| | Sig. (2-tailed) | ,120 | ,115 | ,324 | ,319 | ,080 | ,964 |
| | N | 43 | 40 | 43 | 44 | 43 | 42 |
| Richness_change | Correlation Coefficient | ,154 | -,211 | -,177 | ,122 | -,326* | -,070 |
| | Sig. (2-tailed) | ,323 | ,191 | ,256 | ,431 | ,033 | ,659 |
| | N | 43 | 40 | 43 | 44 | 43 | 42 |
| Evenness_change | Correlation Coefficient | ,112 | -,227 | -,045 | ,171 | ,106 | ,124 |
| | Sig. (2-tailed) | ,473 | ,159 | ,772 | ,266 | ,498 | ,433 |
| | N | 43 | 40 | 43 | 44 | 43 | 42 |
| Actinomyces_change | Correlation Coefficient | -,042 | ,268 | ,173 | -,130 | -,006 | ,008 |
| | Sig. (2-tailed) | ,791 | ,094 | ,268 | ,401 | ,972 | ,958 |
| | N | 43 | 40 | 43 | 44 | 43 | 42 |
| Aerostipes_change | Correlation Coefficient | -,264 | -,206 | ,039 | ,021 | ,325* | -,040 |
| | Sig. (2-tailed) | ,088 | ,201 | ,806 | ,892 | ,034 | ,803 |
| | N | 43 | 40 | 43 | 44 | 43 | 42 |
| Bifidobacterium_change | Correlation Coefficient | -,152 | -,136 | ,098 | -,109 | -,125 | -,225 |
| | Sig. (2-tailed) | ,332 | ,401 | ,533 | ,482 | ,425 | ,151 |
| | N | 43 | 40 | 43 | 44 | 43 | 42 |
| Blautia_change | Correlation Coefficient | -,238 | ,179 | ,084 | ,166 | ,115 | ,163 |
| | Sig. (2-tailed) | ,124 | ,270 | ,594 | ,283 | ,465 | ,301 |
| | N | 43 | 40 | 43 | 44 | 43 | 42 |

### Correlations

| | | STPYY_change | STGLPT_change | age | BMI | FM_stand | Richness | shannon.effective |
|---|---|---|---|---|---|---|---|---|
| LDL_change | Correlation Coefficient | ,080 | ,233 | ,052 | ,202 | ,478** | -,042 | -,119 |
| | Sig. (2-tailed) | ,615 | ,137 | ,733 | ,179 | ,001 | ,781 | ,430 |
| | N | 42 | 42 | 46 | 46 | 46 | 46 | 46 |
| shannon.effective_change | Correlation Coefficient | -,089 | ,014 | -,019 | -,130 | ,014 | ,490** | ,570** |
| | Sig. (2-tailed) | ,575 | ,928 | ,903 | ,395 | ,927 | ,001 | ,000 |
| | N | 42 | 42 | 45 | 45 | 45 | 45 | 45 |
| Richness_change | Correlation Coefficient | -,204 | -,014 | ,089 | -,105 | ,167 | ,500** | ,512** |
| | Sig. (2-tailed) | ,196 | ,931 | ,559 | ,492 | ,274 | ,000 | ,000 |
| | N | 42 | 42 | 45 | 45 | 45 | 45 | 45 |
| Evenness_change | Correlation Coefficient | ,122 | ,073 | -,205 | -,086 | -,303* | ,137 | ,306* |
| | Sig. (2-tailed) | ,442 | ,644 | ,177 | ,575 | ,043 | ,370 | ,041 |
| | N | 42 | 42 | 45 | 45 | 45 | 45 | 45 |
| Actinomyces_change | Correlation Coefficient | ,084 | -,030 | -,004 | -,121 | ,085 | -,028 | -,017 |
| | Sig. (2-tailed) | ,599 | ,852 | ,979 | ,429 | ,577 | ,856 | ,912 |
| | N | 42 | 42 | 45 | 45 | 45 | 45 | 45 |
| Aerostipes_change | Correlation Coefficient | ,001 | ,067 | -,078 | ,105 | -,157 | ,011 | ,036 |
| | Sig. (2-tailed) | ,995 | ,673 | ,611 | ,491 | ,305 | ,943 | ,816 |
| | N | 42 | 42 | 45 | 45 | 45 | 45 | 45 |
| Bifidobacterium_change | Correlation Coefficient | ,012 | ,014 | -,044 | ,013 | ,323* | -,514** | -,582** |
| | Sig. (2-tailed) | ,940 | ,929 | ,774 | ,933 | ,031 | ,000 | ,000 |
| | N | 42 | 42 | 45 | 45 | 45 | 45 | 45 |
| Blautia_change | Correlation Coefficient | -,021 | ,209 | ,083 | -,132 | -,159 | -,145 | -,055 |
| | Sig. (2-tailed) | ,894 | ,184 | ,588 | ,387 | ,296 | ,341 | ,722 |
| | N | 42 | 42 | 45 | 45 | 45 | 45 | 45 |

### Correlations

| | | simpson. effective | Evenness | CHOL_change | LDL_change | shannon. effective_change | Richness_change |
|---|---|---|---|---|---|---|---|
| LDL_change | Correlation Coefficient | -,149 | -,082 | ,926** | 1,000 | -,187 | -,043 |
| | Sig. (2-tailed) | ,322 | ,588 | ,000 | . | ,218 | ,779 |
| | N | 46 | 46 | 46 | 46 | 45 | 45 |
| shannon.effective_change | Correlation Coefficient | ,586** | ,347* | -,144 | -,187 | 1,000 | ,901** |
| | Sig. (2-tailed) | ,000 | ,020 | ,344 | ,218 | . | ,000 |
| | N | 45 | 45 | 45 | 45 | 45 | 45 |
| Richness_change | Correlation Coefficient | ,500** | ,163 | ,038 | -,043 | ,901** | 1,000 |
| | Sig. (2-tailed) | ,000 | ,285 | ,806 | ,779 | ,000 | . |
| | N | 45 | 45 | 45 | 45 | 45 | 45 |
| Evenness_change | Correlation Coefficient | ,373* | ,600** | -,308* | -,206 | ,451** | ,062 |
| | Sig. (2-tailed) | ,012 | ,000 | ,039 | ,175 | ,002 | ,687 |
| | N | 45 | 45 | 45 | 45 | 45 | 45 |
| Actinomyces_change | Correlation Coefficient | -,085 | ,010 | ,118 | ,092 | ,176 | ,275 |
| | Sig. (2-tailed) | ,577 | ,949 | ,441 | ,547 | ,247 | ,067 |
| | N | 45 | 45 | 45 | 45 | 45 | 45 |
| Aerostipes_change | Correlation Coefficient | ,058 | ,155 | -,260 | -,246 | -,101 | -,185 |
| | Sig. (2-tailed) | ,703 | ,309 | ,084 | ,103 | ,508 | ,223 |
| | N | 45 | 45 | 45 | 45 | 45 | 45 |
| Bifidobacterium_change | Correlation Coefficient | -,626** | -,401** | ,153 | ,145 | -,414** | -,300* |
| | Sig. (2-tailed) | ,000 | ,006 | ,316 | ,343 | ,005 | ,045 |
| | N | 45 | 45 | 45 | 45 | 45 | 45 |
| Blautia_change | Correlation Coefficient | -,032 | ,355* | -,016 | -,062 | ,010 | -,036 |
| | Sig. (2-tailed) | ,833 | ,017 | ,914 | ,684 | ,947 | ,817 |
| | N | 45 | 45 | 45 | 45 | 45 | 45 |

### Correlations

| | | | Evenness_change | Actinomyces_change | Aerostipes_change | Bifidobacterium_change | Blautia_change | Collinsella_change |
|---|---|---|---|---|---|---|---|---|
| | LDL_change | Correlation Coefficient | -,206 | ,092 | -,246 | ,145 | -,062 | ,085 |
| | | Sig. (2-tailed) | ,175 | ,547 | ,103 | ,343 | ,684 | ,580 |
| | | N | 45 | 45 | 45 | 45 | 45 | 45 |
| | shannon.effective_change | Correlation Coefficient | ,451** | ,176 | -,101 | -,414** | ,010 | -,269 |
| | | Sig. (2-tailed) | ,002 | ,247 | ,508 | ,005 | ,947 | ,074 |
| | | N | 45 | 45 | 45 | 45 | 45 | 45 |
| | Richness_change | Correlation Coefficient | ,062 | ,275 | -,185 | -,300* | -,036 | -,256 |
| | | Sig. (2-tailed) | ,687 | ,067 | ,223 | ,045 | ,817 | ,090 |
| | | N | 45 | 45 | 45 | 45 | 45 | 45 |
| | Evenness_change | Correlation Coefficient | 1,000 | -,099 | ,142 | -,427** | ,167 | -,115 |
| | | Sig. (2-tailed) | . | ,516 | ,352 | ,003 | ,273 | ,452 |
| | | N | 45 | 45 | 45 | 45 | 45 | 45 |
| | Actinomyces_change | Correlation Coefficient | -,099 | 1,000 | -,172 | ,075 | ,364* | ,391** |
| | | Sig. (2-tailed) | ,516 | . | ,259 | ,626 | ,014 | ,008 |
| | | N | 45 | 45 | 45 | 45 | 45 | 45 |
| | Aerostipes_change | Correlation Coefficient | ,142 | -,172 | 1,000 | -,064 | -,023 | -,083 |
| | | Sig. (2-tailed) | ,352 | ,259 | . | ,675 | ,881 | ,587 |
| | | N | 45 | 45 | 45 | 45 | 45 | 45 |
| | Bifidobacterium_change | Correlation Coefficient | -,427** | ,075 | -,064 | 1,000 | -,024 | ,312* |
| | | Sig. (2-tailed) | ,003 | ,626 | ,675 | . | ,876 | ,037 |
| | | N | 45 | 45 | 45 | 45 | 45 | 45 |
| | Blautia_change | Correlation Coefficient | ,167 | ,364* | -,023 | -,024 | 1,000 | ,173 |
| | | Sig. (2-tailed) | ,273 | ,014 | ,881 | ,876 | . | ,256 |
| | | N | 45 | 45 | 45 | 45 | 45 | 45 |

### Correlations

| | | Desulfovibrio_change | Eggerthella_change | Erysipelatoclostridium_change | Faecalitalea_change | Family.XIII.AD3011.group_change | Family.XIII.UCG.001_change |
|---|---|---|---|---|---|---|---|
| LDL_change | Correlation Coefficient | -,147 | ,137 | ,160 | -,170 | -,126 | -,286 |
| | Sig. (2-tailed) | ,334 | ,371 | ,295 | ,263 | ,409 | ,056 |
| | N | 45 | 45 | 45 | 45 | 45 | 45 |
| shannon.effective_change | Correlation Coefficient | ,041 | -,006 | ,287 | -,093 | ,334* | ,194 |
| | Sig. (2-tailed) | ,791 | ,966 | ,056 | ,543 | ,025 | ,201 |
| | N | 45 | 45 | 45 | 45 | 45 | 45 |
| Richness_change | Correlation Coefficient | ,028 | ,091 | ,267 | -,076 | ,297* | ,180 |
| | Sig. (2-tailed) | ,857 | ,552 | ,077 | ,620 | ,048 | ,237 |
| | N | 45 | 45 | 45 | 45 | 45 | 45 |
| Evenness_change | Correlation Coefficient | ,122 | -,169 | ,145 | -,015 | ,144 | ,090 |
| | Sig. (2-tailed) | ,423 | ,268 | ,341 | ,921 | ,345 | ,557 |
| | N | 45 | 45 | 45 | 45 | 45 | 45 |
| Actinomyces_change | Correlation Coefficient | -,057 | ,254 | -,007 | ,178 | ,214 | -,189 |
| | Sig. (2-tailed) | ,711 | ,092 | ,966 | ,243 | ,159 | ,214 |
| | N | 45 | 45 | 45 | 45 | 45 | 45 |
| Aerostipes_change | Correlation Coefficient | ,062 | -,320* | -,375* | ,193 | ,098 | ,233 |
| | Sig. (2-tailed) | ,684 | ,032 | ,011 | ,205 | ,524 | ,124 |
| | N | 45 | 45 | 45 | 45 | 45 | 45 |
| Bifidobacterium_change | Correlation Coefficient | -,004 | ,083 | -,023 | ,070 | -,494** | -,305* |
| | Sig. (2-tailed) | ,978 | ,587 | ,879 | ,650 | ,001 | ,042 |
| | N | 45 | 45 | 45 | 45 | 45 | 45 |
| Blautia_change | Correlation Coefficient | ,021 | ,208 | ,012 | ,088 | ,014 | -,103 |
| | Sig. (2-tailed) | ,891 | ,171 | ,937 | ,566 | ,927 | ,499 |
| | N | 45 | 45 | 45 | 45 | 45 | 45 |

### Correlations

| | | Gemella_change | Gordonibacter_change | Holdemanella_change | Holdemania_change | Lachnospiraceae.FCS020.group_change | Lachnospiraceae.NK4A136.group_change |
|---|---|---|---|---|---|---|---|
| LDL_change | Correlation Coefficient | ,118 | ,030 | ,119 | -,187 | ,198 | ,030 |
| | Sig. (2-tailed) | ,439 | ,847 | ,437 | ,219 | ,193 | ,846 |
| | N | 45 | 45 | 45 | 45 | 45 | 45 |
| shannon.effective_change | Correlation Coefficient | ,046 | ,213 | ,130 | ,129 | ,090 | ,325* |
| | Sig. (2-tailed) | ,766 | ,160 | ,395 | ,400 | ,555 | ,029 |
| | N | 45 | 45 | 45 | 45 | 45 | 45 |
| Richness_change | Correlation Coefficient | ,103 | ,319* | ,156 | ,157 | ,105 | ,289 |
| | Sig. (2-tailed) | ,501 | ,033 | ,305 | ,304 | ,493 | ,055 |
| | N | 45 | 45 | 45 | 45 | 45 | 45 |
| Evenness_change | Correlation Coefficient | -,096 | -,097 | -,017 | ,045 | ,062 | ,138 |
| | Sig. (2-tailed) | ,532 | ,527 | ,911 | ,772 | ,686 | ,365 |
| | N | 45 | 45 | 45 | 45 | 45 | 45 |
| Actinomyces_change | Correlation Coefficient | ,497** | ,141 | ,092 | -,328* | ,082 | -,158 |
| | Sig. (2-tailed) | ,001 | ,357 | ,548 | ,028 | ,592 | ,300 |
| | N | 45 | 45 | 45 | 45 | 45 | 45 |
| Aerostipes_change | Correlation Coefficient | -,093 | -,081 | ,068 | ,138 | -,184 | -,157 |
| | Sig. (2-tailed) | ,542 | ,595 | ,657 | ,366 | ,227 | ,302 |
| | N | 45 | 45 | 45 | 45 | 45 | 45 |
| Bifidobacterium_change | Correlation Coefficient | ,039 | -,351* | -,172 | -,160 | -,103 | -,370* |
| | Sig. (2-tailed) | ,799 | ,018 | ,258 | ,294 | ,499 | ,012 |
| | N | 45 | 45 | 45 | 45 | 45 | 45 |
| Blautia_change | Correlation Coefficient | ,547** | ,121 | ,251 | -,244 | ,216 | -,377* |
| | Sig. (2-tailed) | ,000 | ,427 | ,096 | ,106 | ,154 | ,011 |
| | N | 45 | 45 | 45 | 45 | 45 | 45 |

### Correlations

| | | Lactiplantibacillus_change | Lactobacillus_change | Libanicoccus_change | Ligilactobacillus_change | Limosilactobacillus_change | Roseburia_change |
|---|---|---|---|---|---|---|---|
| LDL_change | Correlation Coefficient | -,114 | ,255 | -,003 | ,011 | -,056 | -,021 |
| | Sig. (2-tailed) | ,455 | ,091 | ,986 | ,941 | ,713 | ,893 |
| | N | 45 | 45 | 45 | 45 | 45 | 45 |
| shannon.effective_change | Correlation Coefficient | -,116 | -,398** | -,164 | ,006 | ,038 | ,107 |
| | Sig. (2-tailed) | ,446 | ,007 | ,281 | ,969 | ,803 | ,485 |
| | N | 45 | 45 | 45 | 45 | 45 | 45 |
| Richness_change | Correlation Coefficient | -,049 | -,220 | -,170 | -,101 | ,025 | ,211 |
| | Sig. (2-tailed) | ,751 | ,147 | ,264 | ,508 | ,872 | ,165 |
| | N | 45 | 45 | 45 | 45 | 45 | 45 |
| Evenness_change | Correlation Coefficient | -,240 | -,370* | ,053 | ,320* | -,008 | -,188 |
| | Sig. (2-tailed) | ,112 | ,012 | ,732 | ,032 | ,958 | ,216 |
| | N | 45 | 45 | 45 | 45 | 45 | 45 |
| Actinomyces_change | Correlation Coefficient | ,225 | -,099 | -,038 | ,003 | -,038 | -,040 |
| | Sig. (2-tailed) | ,137 | ,519 | ,803 | ,982 | ,806 | ,792 |
| | N | 45 | 45 | 45 | 45 | 45 | 45 |
| Aerostipes_change | Correlation Coefficient | -,043 | -,033 | ,071 | -,024 | ,228 | -,111 |
| | Sig. (2-tailed) | ,779 | ,828 | ,643 | ,874 | ,132 | ,468 |
| | N | 45 | 45 | 45 | 45 | 45 | 45 |
| Bifidobacterium_change | Correlation Coefficient | ,360* | ,159 | ,153 | ,072 | ,077 | ,142 |
| | Sig. (2-tailed) | ,015 | ,297 | ,315 | ,640 | ,616 | ,353 |
| | N | 45 | 45 | 45 | 45 | 45 | 45 |
| Blautia_change | Correlation Coefficient | ,258 | -,086 | ,051 | ,133 | ,093 | -,313* |
| | Sig. (2-tailed) | ,087 | ,575 | ,741 | ,384 | ,542 | ,037 |
| | N | 45 | 45 | 45 | 45 | 45 | 45 |

### Correlations

| | | Shuttleworthia_change | Subdoligranulum_change | UCG.003_change | X.Eubacterium..brachy.group_change | X.Ruminococcus..gauvreauii.group_change | X.Ruminococcus..torques.group_change |
|---|---|---|---|---|---|---|---|
| LDL_change | Correlation Coefficient | ,231 | -,104 | -,018 | -,050 | ,241 | -,037 |
| | Sig. (2-tailed) | ,126 | ,496 | ,907 | ,745 | ,111 | ,812 |
| | N | 45 | 45 | 45 | 45 | 45 | 45 |
| shannon.effective_change | Correlation Coefficient | -,047 | ,162 | ,051 | ,281 | ,010 | ,083 |
| | Sig. (2-tailed) | ,758 | ,288 | ,738 | ,061 | ,950 | ,586 |
| | N | 45 | 45 | 45 | 45 | 45 | 45 |
| Richness_change | Correlation Coefficient | ,056 | ,054 | ,088 | ,364* | -,028 | ,019 |
| | Sig. (2-tailed) | ,717 | ,722 | ,567 | ,014 | ,854 | ,904 |
| | N | 45 | 45 | 45 | 45 | 45 | 45 |
| Evenness_change | Correlation Coefficient | -,228 | ,348* | -,141 | -,061 | ,121 | ,242 |
| | Sig. (2-tailed) | ,132 | ,019 | ,357 | ,689 | ,428 | ,109 |
| | N | 45 | 45 | 45 | 45 | 45 | 45 |
| Actinomyces_change | Correlation Coefficient | -,089 | ,121 | -,185 | ,205 | ,018 | ,286 |
| | Sig. (2-tailed) | ,561 | ,428 | ,225 | ,178 | ,908 | ,056 |
| | N | 45 | 45 | 45 | 45 | 45 | 45 |
| Aerostipes_change | Correlation Coefficient | -,052 | ,163 | -,084 | ,134 | -,190 | ,134 |
| | Sig. (2-tailed) | ,735 | ,285 | ,582 | ,381 | ,211 | ,380 |
| | N | 45 | 45 | 45 | 45 | 45 | 45 |
| Bifidobacterium_change | Correlation Coefficient | ,255 | -,264 | -,189 | -,085 | ,233 | -,128 |
| | Sig. (2-tailed) | ,091 | ,079 | ,215 | ,580 | ,124 | ,402 |
| | N | 45 | 45 | 45 | 45 | 45 | 45 |
| Blautia_change | Correlation Coefficient | -,246 | ,239 | -,396** | ,195 | ,209 | ,288 |
| | Sig. (2-tailed) | ,104 | ,114 | ,007 | ,199 | ,169 | ,055 |
| | N | 45 | 45 | 45 | 45 | 45 | 45 |

### Correlations

| | | | interaction_VTA_change | interaction_rOFC_change | interaction_rmOFC_change | sumscore_GIQLI_change | Stool_frequency_change | STGHRE_change |
|---|---|---|---|---|---|---|---|---|
| | Collinsella_change | Correlation Coefficient | ,024 | ,178 | ,150 | -,202 | ,151 | ,014 |
| | | Sig. (2-tailed) | ,878 | ,271 | ,339 | ,189 | ,333 | ,930 |
| | | N | 43 | 40 | 43 | 44 | 43 | 42 |
| | Desulfovibrio_change | Correlation Coefficient | ,034 | -,009 | -,084 | ,059 | ,066 | -,095 |
| | | Sig. (2-tailed) | ,830 | ,955 | ,591 | ,704 | ,674 | ,551 |
| | | N | 43 | 40 | 43 | 44 | 43 | 42 |
| | Eggerthella_change | Correlation Coefficient | ,020 | ,022 | -,027 | ,104 | -,131 | -,126 |
| | | Sig. (2-tailed) | ,899 | ,892 | ,862 | ,503 | ,403 | ,425 |
| | | N | 43 | 40 | 43 | 44 | 43 | 42 |
| | Erysipelatoclostridium_change | Correlation Coefficient | ,159 | -,021 | -,063 | ,190 | -,291 | -,075 |
| | | Sig. (2-tailed) | ,309 | ,900 | ,688 | ,216 | ,058 | ,639 |
| | | N | 43 | 40 | 43 | 44 | 43 | 42 |
| | Faecalitalea_change | Correlation Coefficient | -,099 | -,018 | ,035 | ,038 | -,040 | ,166 |
| | | Sig. (2-tailed) | ,526 | ,911 | ,825 | ,809 | ,798 | ,294 |
| | | N | 43 | 40 | 43 | 44 | 43 | 42 |
| | Family.XIII.AD3011.group_change | Correlation Coefficient | ,121 | ,191 | ,070 | ,180 | ,250 | ,125 |
| | | Sig. (2-tailed) | ,440 | ,238 | ,655 | ,241 | ,105 | ,430 |
| | | N | 43 | 40 | 43 | 44 | 43 | 42 |
| | Family.XIII.UCG.001_change | Correlation Coefficient | -,080 | ,064 | -,206 | ,009 | -,054 | ,215 |
| | | Sig. (2-tailed) | ,611 | ,696 | ,185 | ,953 | ,731 | ,171 |
| | | N | 43 | 40 | 43 | 44 | 43 | 42 |
| | Gemella_change | Correlation Coefficient | -,174 | ,295 | ,196 | ,086 | -,119 | -,105 |
| | | Sig. (2-tailed) | ,264 | ,064 | ,209 | ,577 | ,445 | ,510 |
| | | N | 43 | 40 | 43 | 44 | 43 | 42 |

### Correlations

| | | | STPYY_change | STGLPT_change | age | BMI | FM_stand | Richness | shannon.effective |
|---|---|---|---|---|---|---|---|---|---|
| | Collinsella_change | Correlation Coefficient | ,176 | ,004 | -,228 | -,035 | ,013 | -,354* | -,322* |
| | | Sig. (2-tailed) | ,266 | ,978 | ,133 | ,820 | ,932 | ,017 | ,031 |
| | | N | 42 | 42 | 45 | 45 | 45 | 45 | 45 |
| | Desulfovibrio_change | Correlation Coefficient | -,370* | -,069 | ,135 | -,268 | -,284 | -,048 | -,048 |
| | | Sig. (2-tailed) | ,016 | ,666 | ,378 | ,075 | ,059 | ,753 | ,755 |
| | | N | 42 | 42 | 45 | 45 | 45 | 45 | 45 |
| | Eggerthella_change | Correlation Coefficient | ,032 | ,103 | ,151 | ,107 | ,228 | ,118 | ,104 |
| | | Sig. (2-tailed) | ,839 | ,518 | ,322 | ,485 | ,131 | ,438 | ,498 |
| | | N | 42 | 42 | 45 | 45 | 45 | 45 | 45 |
| | Erysipelatoclostridium_change | Correlation Coefficient | -,017 | ,296 | ,005 | ,250 | ,207 | ,120 | ,112 |
| | | Sig. (2-tailed) | ,913 | ,057 | ,973 | ,097 | ,172 | ,433 | ,464 |
| | | N | 42 | 42 | 45 | 45 | 45 | 45 | 45 |
| | Faecalitalea_change | Correlation Coefficient | ,011 | ,090 | -,041 | ,041 | ,055 | ,011 | ,026 |
| | | Sig. (2-tailed) | ,945 | ,570 | ,788 | ,791 | ,721 | ,942 | ,864 |
| | | N | 42 | 42 | 45 | 45 | 45 | 45 | 45 |
| | Family.XIII.AD3011.group_change | Correlation Coefficient | ,171 | ,058 | -,057 | ,033 | -,024 | ,350* | ,325* |
| | | Sig. (2-tailed) | ,279 | ,717 | ,711 | ,831 | ,876 | ,018 | ,029 |
| | | N | 42 | 42 | 45 | 45 | 45 | 45 | 45 |
| | Family.XIII.UCG.001_change | Correlation Coefficient | ,017 | ,205 | ,067 | -,199 | -,306* | ,256 | ,236 |
| | | Sig. (2-tailed) | ,913 | ,192 | ,663 | ,190 | ,041 | ,090 | ,119 |
| | | N | 42 | 42 | 45 | 45 | 45 | 45 | 45 |
| | Gemella_change | Correlation Coefficient | -,139 | ,150 | ,200 | -,204 | -,199 | -,070 | -,050 |
| | | Sig. (2-tailed) | ,381 | ,342 | ,187 | ,180 | ,189 | ,648 | ,745 |
| | | N | 42 | 42 | 45 | 45 | 45 | 45 | 45 |

### Correlations

| | | | simpson. effective | Evenness | CHOL_change | LDL_change | shannon. effective_change | Richness_change |
|---|---|---|---|---|---|---|---|---|
| | Collinsella_change | Correlation Coefficient | -,369* | -,060 | ,110 | ,085 | -,269 | -,256 |
| | | Sig. (2-tailed) | ,012 | ,697 | ,474 | ,580 | ,074 | ,090 |
| | | N | 45 | 45 | 45 | 45 | 45 | 45 |
| | Desulfovibrio_change | Correlation Coefficient | -,059 | -,091 | -,270 | -,147 | ,041 | ,028 |
| | | Sig. (2-tailed) | ,699 | ,553 | ,073 | ,334 | ,791 | ,857 |
| | | N | 45 | 45 | 45 | 45 | 45 | 45 |
| | Eggerthella_change | Correlation Coefficient | ,084 | ,081 | ,203 | ,137 | -,006 | ,091 |
| | | Sig. (2-tailed) | ,583 | ,595 | ,181 | ,371 | ,966 | ,552 |
| | | N | 45 | 45 | 45 | 45 | 45 | 45 |
| | Erysipelatoclostridium_change | Correlation Coefficient | ,126 | -,016 | ,152 | ,160 | ,287 | ,267 |
| | | Sig. (2-tailed) | ,408 | ,919 | ,318 | ,295 | ,056 | ,077 |
| | | N | 45 | 45 | 45 | 45 | 45 | 45 |
| | Faecalitalea_change | Correlation Coefficient | ,041 | ,117 | -,149 | -,170 | -,093 | -,076 |
| | | Sig. (2-tailed) | ,788 | ,443 | ,328 | ,263 | ,543 | ,620 |
| | | N | 45 | 45 | 45 | 45 | 45 | 45 |
| | Family.XIII.AD3011. group_change | Correlation Coefficient | ,293 | ,022 | -,174 | -,126 | ,334* | ,297* |
| | | Sig. (2-tailed) | ,051 | ,885 | ,253 | ,409 | ,025 | ,048 |
| | | N | 45 | 45 | 45 | 45 | 45 | 45 |
| | Family.XIII.UCG. 001_change | Correlation Coefficient | ,240 | -,001 | -,319* | -,286 | ,194 | ,180 |
| | | Sig. (2-tailed) | ,113 | ,996 | ,033 | ,056 | ,201 | ,237 |
| | | N | 45 | 45 | 45 | 45 | 45 | 45 |
| | Gemella_change | Correlation Coefficient | -,066 | -,034 | ,155 | ,118 | ,046 | ,103 |
| | | Sig. (2-tailed) | ,667 | ,824 | ,311 | ,439 | ,766 | ,501 |
| | | N | 45 | 45 | 45 | 45 | 45 | 45 |

### Correlations

| | | | Evenness_change | Actinomyces_change | Aerostipes_change | Bifidobacterium_change | Blautia_change | Collinsella_change |
|---|---|---|---|---|---|---|---|---|
| | Collinsella_change | Correlation Coefficient | -,115 | ,391** | -,083 | ,312* | ,173 | 1,000 |
| | | Sig. (2-tailed) | ,452 | ,008 | ,587 | ,037 | ,256 | . |
| | | N | 45 | 45 | 45 | 45 | 45 | 45 |
| | Desulfovibrio_change | Correlation Coefficient | ,122 | -,057 | ,062 | -,004 | ,021 | -,270 |
| | | Sig. (2-tailed) | ,423 | ,711 | ,684 | ,978 | ,891 | ,073 |
| | | N | 45 | 45 | 45 | 45 | 45 | 45 |
| | Eggerthella_change | Correlation Coefficient | -,169 | ,254 | -,320* | ,083 | ,208 | -,016 |
| | | Sig. (2-tailed) | ,268 | ,092 | ,032 | ,587 | ,171 | ,919 |
| | | N | 45 | 45 | 45 | 45 | 45 | 45 |
| | Erysipelatoclostridium_change | Correlation Coefficient | ,145 | -,007 | -,375* | -,023 | ,012 | ,054 |
| | | Sig. (2-tailed) | ,341 | ,966 | ,011 | ,879 | ,937 | ,724 |
| | | N | 45 | 45 | 45 | 45 | 45 | 45 |
| | Faecalitalea_change | Correlation Coefficient | -,015 | ,178 | ,193 | ,070 | ,088 | -,073 |
| | | Sig. (2-tailed) | ,921 | ,243 | ,205 | ,650 | ,566 | ,632 |
| | | N | 45 | 45 | 45 | 45 | 45 | 45 |
| | Family.XIII.AD3011.group_change | Correlation Coefficient | ,144 | ,214 | ,098 | -,494** | ,014 | -,084 |
| | | Sig. (2-tailed) | ,345 | ,159 | ,524 | ,001 | ,927 | ,583 |
| | | N | 45 | 45 | 45 | 45 | 45 | 45 |
| | Family.XIII.UCG.001_change | Correlation Coefficient | ,090 | -,189 | ,233 | -,305* | -,103 | -,363* |
| | | Sig. (2-tailed) | ,557 | ,214 | ,124 | ,042 | ,499 | ,014 |
| | | N | 45 | 45 | 45 | 45 | 45 | 45 |
| | Gemella_change | Correlation Coefficient | -,096 | ,497** | -,093 | ,039 | ,547** | ,322* |
| | | Sig. (2-tailed) | ,532 | ,001 | ,542 | ,799 | ,000 | ,031 |
| | | N | 45 | 45 | 45 | 45 | 45 | 45 |

### Correlations

| | | | Desulfovibrio_change | Eggerthella_change | Erysipelatoclostridium_change | Faecalitalea_change | Family.XIII.AD3011.group_change | Family.XIII.UCG.001_change |
|---|---|---|---|---|---|---|---|---|
| | Collinsella_change | Correlation Coefficient | -,270 | -,016 | ,054 | -,073 | -,084 | -,363* |
| | | Sig. (2-tailed) | ,073 | ,919 | ,724 | ,632 | ,583 | ,014 |
| | | N | 45 | 45 | 45 | 45 | 45 | 45 |
| | Desulfovibrio_change | Correlation Coefficient | 1,000 | -,155 | ,038 | ,067 | -,074 | ,158 |
| | | Sig. (2-tailed) | . | ,311 | ,805 | ,663 | ,629 | ,300 |
| | | N | 45 | 45 | 45 | 45 | 45 | 45 |
| | Eggerthella_change | Correlation Coefficient | -,155 | 1,000 | ,224 | ,365* | -,051 | -,217 |
| | | Sig. (2-tailed) | ,311 | . | ,139 | ,014 | ,737 | ,151 |
| | | N | 45 | 45 | 45 | 45 | 45 | 45 |
| | Erysipelatoclostridium_change | Correlation Coefficient | ,038 | ,224 | 1,000 | ,148 | ,065 | ,064 |
| | | Sig. (2-tailed) | ,805 | ,139 | . | ,331 | ,673 | ,676 |
| | | N | 45 | 45 | 45 | 45 | 45 | 45 |
| | Faecalitalea_change | Correlation Coefficient | ,067 | ,365* | ,148 | 1,000 | -,136 | ,064 |
| | | Sig. (2-tailed) | ,663 | ,014 | ,331 | . | ,374 | ,678 |
| | | N | 45 | 45 | 45 | 45 | 45 | 45 |
| | Family.XIII.AD3011.group_change | Correlation Coefficient | -,074 | -,051 | ,065 | -,136 | 1,000 | ,249 |
| | | Sig. (2-tailed) | ,629 | ,737 | ,673 | ,374 | . | ,100 |
| | | N | 45 | 45 | 45 | 45 | 45 | 45 |
| | Family.XIII.UCG.001_change | Correlation Coefficient | ,158 | -,217 | ,064 | ,064 | ,249 | 1,000 |
| | | Sig. (2-tailed) | ,300 | ,151 | ,676 | ,678 | ,100 | . |
| | | N | 45 | 45 | 45 | 45 | 45 | 45 |
| | Gemella_change | Correlation Coefficient | ,194 | ,023 | ,101 | ,008 | -,021 | -,127 |
| | | Sig. (2-tailed) | ,202 | ,879 | ,510 | ,957 | ,893 | ,405 |
| | | N | 45 | 45 | 45 | 45 | 45 | 45 |

### Correlations

| | | | Gemella_change | Gordonibacter_change | Holdemanella_change | Holdemania_change | Lachnospiraceae.FCS020.group_change | Lachnospiraceae.NK4A136.group_change |
|---|---|---|---|---|---|---|---|---|
| | Collinsella_change | Correlation Coefficient | ,322* | -,052 | ,071 | -,286 | ,106 | -,529** |
| | | Sig. (2-tailed) | ,031 | ,737 | ,644 | ,057 | ,489 | ,000 |
| | | N | 45 | 45 | 45 | 45 | 45 | 45 |
| | Desulfovibrio_change | Correlation Coefficient | ,194 | -,078 | -,030 | ,003 | -,042 | -,024 |
| | | Sig. (2-tailed) | ,202 | ,610 | ,845 | ,986 | ,782 | ,873 |
| | | N | 45 | 45 | 45 | 45 | 45 | 45 |
| | Eggerthella_change | Correlation Coefficient | ,023 | ,384** | ,098 | ,139 | ,020 | -,054 |
| | | Sig. (2-tailed) | ,879 | ,009 | ,521 | ,362 | ,897 | ,727 |
| | | N | 45 | 45 | 45 | 45 | 45 | 45 |
| | Erysipelatoclostridium_change | Correlation Coefficient | ,101 | ,224 | ,088 | -,060 | ,162 | ,052 |
| | | Sig. (2-tailed) | ,510 | ,140 | ,566 | ,694 | ,288 | ,736 |
| | | N | 45 | 45 | 45 | 45 | 45 | 45 |
| | Faecalitalea_change | Correlation Coefficient | ,008 | ,183 | ,085 | ,040 | -,221 | -,278 |
| | | Sig. (2-tailed) | ,957 | ,230 | ,581 | ,794 | ,145 | ,064 |
| | | N | 45 | 45 | 45 | 45 | 45 | 45 |
| | Family.XIII.AD3011.group_change | Correlation Coefficient | -,021 | -,040 | ,227 | -,016 | ,038 | ,335* |
| | | Sig. (2-tailed) | ,893 | ,792 | ,134 | ,916 | ,802 | ,024 |
| | | N | 45 | 45 | 45 | 45 | 45 | 45 |
| | Family.XIII.UCG.001_change | Correlation Coefficient | -,127 | -,023 | -,114 | ,170 | ,064 | ,207 |
| | | Sig. (2-tailed) | ,405 | ,879 | ,458 | ,263 | ,676 | ,172 |
| | | N | 45 | 45 | 45 | 45 | 45 | 45 |
| | Gemella_change | Correlation Coefficient | 1,000 | ,128 | ,330* | -,465** | ,300* | -,431** |
| | | Sig. (2-tailed) | . | ,402 | ,027 | ,001 | ,045 | ,003 |
| | | N | 45 | 45 | 45 | 45 | 45 | 45 |

### Correlations

| | | | Lactiplantibacillus_change | Lactobacillus_change | Libanicoccus_change | Ligilactobacillus_change | Limosilactobacillus_change | Roseburia_change |
|---|---|---|---|---|---|---|---|---|
| | Collinsella_change | Correlation Coefficient | ,286 | -,006 | ,077 | ,218 | ,132 | -,219 |
| | | Sig. (2-tailed) | ,057 | ,969 | ,614 | ,150 | ,388 | ,149 |
| | | N | 45 | 45 | 45 | 45 | 45 | 45 |
| | Desulfovibrio_change | Correlation Coefficient | -,018 | ,201 | ,338* | -,054 | ,258 | ,130 |
| | | Sig. (2-tailed) | ,905 | ,186 | ,023 | ,725 | ,087 | ,393 |
| | | N | 45 | 45 | 45 | 45 | 45 | 45 |
| | Eggerthella_change | Correlation Coefficient | -,150 | ,141 | ,069 | -,030 | -,057 | -,037 |
| | | Sig. (2-tailed) | ,326 | ,356 | ,652 | ,845 | ,712 | ,807 |
| | | N | 45 | 45 | 45 | 45 | 45 | 45 |
| | Erysipelatoclostridium_change | Correlation Coefficient | -,149 | -,121 | ,019 | ,245 | -,069 | ,107 |
| | | Sig. (2-tailed) | ,329 | ,430 | ,903 | ,104 | ,652 | ,484 |
| | | N | 45 | 45 | 45 | 45 | 45 | 45 |
| | Faecalitalea_change | Correlation Coefficient | -,141 | ,157 | ,072 | -,043 | ,036 | ,126 |
| | | Sig. (2-tailed) | ,355 | ,303 | ,638 | ,778 | ,812 | ,411 |
| | | N | 45 | 45 | 45 | 45 | 45 | 45 |
| | Family.XIII.AD3011.group_change | Correlation Coefficient | ,185 | -,149 | -,323* | ,035 | -,104 | -,224 |
| | | Sig. (2-tailed) | ,224 | ,330 | ,031 | ,821 | ,496 | ,139 |
| | | N | 45 | 45 | 45 | 45 | 45 | 45 |
| | Family.XIII.UCG.001_change | Correlation Coefficient | -,151 | ,058 | -,146 | ,222 | ,128 | ,120 |
| | | Sig. (2-tailed) | ,322 | ,705 | ,337 | ,142 | ,402 | ,432 |
| | | N | 45 | 45 | 45 | 45 | 45 | 45 |
| | Gemella_change | Correlation Coefficient | ,304* | ,020 | ,122 | ,093 | ,253 | -,094 |
| | | Sig. (2-tailed) | ,043 | ,899 | ,424 | ,545 | ,094 | ,541 |
| | | N | 45 | 45 | 45 | 45 | 45 | 45 |

### Correlations

| | | | Shuttleworthia_change | Subdoligranulum_change | UCG.003_change | X.Eubacterium..brachy.group_change | X.Ruminococcus.gauvreauii.group_change | X.Ruminococcus.torques.group_change |
|---|---|---|---|---|---|---|---|---|
| | Collinsella_change | Correlation Coefficient | ,092 | ,025 | -,109 | -,063 | ,005 | ,158 |
| | | Sig. (2-tailed) | ,546 | ,869 | ,474 | ,682 | ,976 | ,300 |
| | | N | 45 | 45 | 45 | 45 | 45 | 45 |
| | Desulfovibrio_change | Correlation Coefficient | ,020 | -,113 | -,052 | ,100 | -,043 | -,151 |
| | | Sig. (2-tailed) | ,894 | ,462 | ,735 | ,513 | ,778 | ,323 |
| | | N | 45 | 45 | 45 | 45 | 45 | 45 |
| | Eggerthella_change | Correlation Coefficient | ,146 | ,022 | -,180 | ,125 | ,102 | -,038 |
| | | Sig. (2-tailed) | ,338 | ,886 | ,237 | ,412 | ,505 | ,804 |
| | | N | 45 | 45 | 45 | 45 | 45 | 45 |
| | Erysipelatoclostridium_change | Correlation Coefficient | ,078 | ,064 | ,000 | ,156 | ,038 | ,060 |
| | | Sig. (2-tailed) | ,613 | ,676 | ,999 | ,307 | ,803 | ,698 |
| | | N | 45 | 45 | 45 | 45 | 45 | 45 |
| | Faecalitalea_change | Correlation Coefficient | -,141 | ,039 | -,038 | ,302* | -,074 | ,190 |
| | | Sig. (2-tailed) | ,356 | ,801 | ,804 | ,044 | ,629 | ,211 |
| | | N | 45 | 45 | 45 | 45 | 45 | 45 |
| | Family.XIII.AD3011.group_change | Correlation Coefficient | -,048 | ,090 | -,018 | -,084 | -,026 | ,281 |
| | | Sig. (2-tailed) | ,752 | ,555 | ,905 | ,585 | ,867 | ,062 |
| | | N | 45 | 45 | 45 | 45 | 45 | 45 |
| | Family.XIII.UCG.001_change | Correlation Coefficient | -,280 | -,025 | ,070 | ,068 | -,256 | ,041 |
| | | Sig. (2-tailed) | ,062 | ,872 | ,645 | ,655 | ,089 | ,791 |
| | | N | 45 | 45 | 45 | 45 | 45 | 45 |
| | Gemella_change | Correlation Coefficient | -,146 | ,072 | -,274 | ,233 | ,000 | ,082 |
| | | Sig. (2-tailed) | ,339 | ,636 | ,069 | ,123 | ,998 | ,594 |
| | | N | 45 | 45 | 45 | 45 | 45 | 45 |

### Correlations

| | | | interaction_VTA_change | interaction_rOFC_change | interaction_rmOFC_change | sumscore_GIQLI_change | Stool_frequency_change | STGHRE_change |
|---|---|---|---|---|---|---|---|---|
| | Gordonibacter_change | Correlation Coefficient | ,025 | ,044 | ,079 | ,079 | -,099 | -,040 |
| | | Sig. (2-tailed) | ,876 | ,785 | ,614 | ,610 | ,526 | ,803 |
| | | N | 43 | 40 | 43 | 44 | 43 | 42 |
| | Holdemanella_change | Correlation Coefficient | -,083 | ,049 | ,005 | ,160 | -,104 | -,104 |
| | | Sig. (2-tailed) | ,597 | ,764 | ,972 | ,301 | ,505 | ,514 |
| | | N | 43 | 40 | 43 | 44 | 43 | 42 |
| | Holdemania_change | Correlation Coefficient | -,116 | -,259 | -,195 | ,095 | -,016 | ,015 |
| | | Sig. (2-tailed) | ,459 | ,107 | ,211 | ,539 | ,919 | ,927 |
| | | N | 43 | 40 | 43 | 44 | 43 | 42 |
| | Lachnospiraceae.FCS020.group_change | Correlation Coefficient | -,041 | -,079 | ,028 | ,133 | -,071 | -,078 |
| | | Sig. (2-tailed) | ,792 | ,627 | ,860 | ,390 | ,649 | ,624 |
| | | N | 43 | 40 | 43 | 44 | 43 | 42 |
| | Lachnospiraceae.NK4A136.group_change | Correlation Coefficient | ,286 | -,093 | -,001 | ,000 | -,013 | ,094 |
| | | Sig. (2-tailed) | ,063 | ,566 | ,996 | ,998 | ,933 | ,554 |
| | | N | 43 | 40 | 43 | 44 | 43 | 42 |
| | Lactiplantibacillus_change | Correlation Coefficient | -,006 | ,120 | ,397** | ,168 | -,007 | ,131 |
| | | Sig. (2-tailed) | ,972 | ,459 | ,008 | ,276 | ,966 | ,407 |
| | | N | 43 | 40 | 43 | 44 | 43 | 42 |
| | Lactobacillus_change | Correlation Coefficient | -,023 | ,046 | -,054 | -,033 | ,105 | -,070 |
| | | Sig. (2-tailed) | ,882 | ,778 | ,731 | ,833 | ,504 | ,659 |
| | | N | 43 | 40 | 43 | 44 | 43 | 42 |
| | Libanicoccus_change | Correlation Coefficient | ,294 | ,211 | ,223 | -,057 | -,187 | -,107 |
| | | Sig. (2-tailed) | ,056 | ,192 | ,151 | ,712 | ,230 | ,499 |
| | | N | 43 | 40 | 43 | 44 | 43 | 42 |

### Correlations

| | | STPYY_change | STGLPT_change | age | BMI | FM_stand | Richness | shannon. effective |
|---|---|---|---|---|---|---|---|---|
| Gordonibacter_change | Correlation Coefficient | -,341* | ,132 | ,239 | ,173 | ,090 | ,244 | ,257 |
| | Sig. (2-tailed) | ,027 | ,404 | ,114 | ,257 | ,558 | ,107 | ,089 |
| | N | 42 | 42 | 45 | 45 | 45 | 45 | 45 |
| Holdemanella_change | Correlation Coefficient | -,043 | ,166 | -,098 | ,079 | ,056 | ,303* | ,318* |
| | Sig. (2-tailed) | ,785 | ,294 | ,523 | ,606 | ,713 | ,043 | ,033 |
| | N | 42 | 42 | 45 | 45 | 45 | 45 | 45 |
| Holdemania_change | Correlation Coefficient | -,157 | -,266 | ,177 | ,284 | ,017 | ,395** | ,414** |
| | Sig. (2-tailed) | ,321 | ,089 | ,245 | ,058 | ,911 | ,007 | ,005 |
| | N | 42 | 42 | 45 | 45 | 45 | 45 | 45 |
| Lachnospiraceae.FCS020.group_change | Correlation Coefficient | ,076 | ,162 | -,187 | ,056 | -,044 | ,238 | ,193 |
| | Sig. (2-tailed) | ,633 | ,307 | ,220 | ,713 | ,774 | ,115 | ,204 |
| | N | 42 | 42 | 45 | 45 | 45 | 45 | 45 |
| Lachnospiraceae.NK4A136.group_change | Correlation Coefficient | ,155 | -,079 | ,152 | ,109 | ,114 | ,177 | ,117 |
| | Sig. (2-tailed) | ,326 | ,621 | ,319 | ,476 | ,454 | ,246 | ,443 |
| | N | 42 | 42 | 45 | 45 | 45 | 45 | 45 |
| Lactiplantibacillus_change | Correlation Coefficient | -,079 | -,186 | -,003 | -,024 | ,182 | -,214 | -,256 |
| | Sig. (2-tailed) | ,618 | ,238 | ,986 | ,874 | ,231 | ,159 | ,090 |
| | N | 42 | 42 | 45 | 45 | 45 | 45 | 45 |
| Lactobacillus_change | Correlation Coefficient | ,046 | -,181 | ,190 | -,043 | ,118 | -,278 | -,322* |
| | Sig. (2-tailed) | ,771 | ,251 | ,212 | ,777 | ,440 | ,065 | ,031 |
| | N | 42 | 42 | 45 | 45 | 45 | 45 | 45 |
| Libanicoccus_change | Correlation Coefficient | ,039 | ,115 | ,250 | ,042 | -,048 | ,110 | ,108 |
| | Sig. (2-tailed) | ,807 | ,468 | ,098 | ,786 | ,753 | ,470 | ,481 |
| | N | 42 | 42 | 45 | 45 | 45 | 45 | 45 |

### Correlations

| | | | simpson. effective | Evenness | CHOL_change | LDL_change | shannon. effective_change | Richness_change |
|---|---|---|---|---|---|---|---|---|
| | Gordonibacter_change | Correlation Coefficient | ,266 | ,248 | ,192 | ,030 | ,213 | ,319* |
| | | Sig. (2-tailed) | ,077 | ,101 | ,205 | ,847 | ,160 | ,033 |
| | | N | 45 | 45 | 45 | 45 | 45 | 45 |
| | Holdemanella_change | Correlation Coefficient | ,357* | ,092 | ,137 | ,119 | ,130 | ,156 |
| | | Sig. (2-tailed) | ,016 | ,546 | ,369 | ,437 | ,395 | ,305 |
| | | N | 45 | 45 | 45 | 45 | 45 | 45 |
| | Holdemania_change | Correlation Coefficient | ,438** | ,206 | -,201 | -,187 | ,129 | ,157 |
| | | Sig. (2-tailed) | ,003 | ,175 | ,186 | ,219 | ,400 | ,304 |
| | | N | 45 | 45 | 45 | 45 | 45 | 45 |
| | Lachnospiraceae.FCS020.group_change | Correlation Coefficient | ,205 | -,019 | ,182 | ,198 | ,090 | ,105 |
| | | Sig. (2-tailed) | ,178 | ,903 | ,232 | ,193 | ,555 | ,493 |
| | | N | 45 | 45 | 45 | 45 | 45 | 45 |
| | Lachnospiraceae.NK4A136.group_change | Correlation Coefficient | ,108 | -,153 | ,013 | ,030 | ,325* | ,289 |
| | | Sig. (2-tailed) | ,480 | ,316 | ,933 | ,846 | ,029 | ,055 |
| | | N | 45 | 45 | 45 | 45 | 45 | 45 |
| | Lactiplantibacillus_change | Correlation Coefficient | -,284 | -,172 | -,052 | -,114 | -,116 | -,049 |
| | | Sig. (2-tailed) | ,059 | ,258 | ,735 | ,455 | ,446 | ,751 |
| | | N | 45 | 45 | 45 | 45 | 45 | 45 |
| | Lactobacillus_change | Correlation Coefficient | -,340* | -,226 | ,184 | ,255 | -,398** | -,220 |
| | | Sig. (2-tailed) | ,022 | ,135 | ,225 | ,091 | ,007 | ,147 |
| | | N | 45 | 45 | 45 | 45 | 45 | 45 |
| | Libanicoccus_change | Correlation Coefficient | ,007 | -,031 | -,032 | -,003 | -,164 | -,170 |
| | | Sig. (2-tailed) | ,963 | ,837 | ,834 | ,986 | ,281 | ,264 |
| | | N | 45 | 45 | 45 | 45 | 45 | 45 |

### Correlations

| | | | Evenness_change | Actinomyces_change | Aerostipes_change | Bifidobacterium_change | Blautia_change | Collinsella_change |
|---|---|---|---|---|---|---|---|---|
| | Gordonibacter_change | Correlation Coefficient | -,097 | ,141 | -,081 | -,351* | ,121 | -,052 |
| | | Sig. (2-tailed) | ,527 | ,357 | ,595 | ,018 | ,427 | ,737 |
| | | N | 45 | 45 | 45 | 45 | 45 | 45 |
| | Holdemanella_change | Correlation Coefficient | -,017 | ,092 | ,068 | -,172 | ,251 | ,071 |
| | | Sig. (2-tailed) | ,911 | ,548 | ,657 | ,258 | ,096 | ,644 |
| | | N | 45 | 45 | 45 | 45 | 45 | 45 |
| | Holdemania_change | Correlation Coefficient | ,045 | -,328* | ,138 | -,160 | -,244 | -,286 |
| | | Sig. (2-tailed) | ,772 | ,028 | ,366 | ,294 | ,106 | ,057 |
| | | N | 45 | 45 | 45 | 45 | 45 | 45 |
| | Lachnospiraceae.FCS020.group_change | Correlation Coefficient | ,062 | ,082 | -,184 | -,103 | ,216 | ,106 |
| | | Sig. (2-tailed) | ,686 | ,592 | ,227 | ,499 | ,154 | ,489 |
| | | N | 45 | 45 | 45 | 45 | 45 | 45 |
| | Lachnospiraceae.NK4A136.group_change | Correlation Coefficient | ,138 | -,158 | -,157 | -,370* | -,377* | -,529** |
| | | Sig. (2-tailed) | ,365 | ,300 | ,302 | ,012 | ,011 | ,000 |
| | | N | 45 | 45 | 45 | 45 | 45 | 45 |
| | Lactiplantibacillus_change | Correlation Coefficient | -,240 | ,225 | -,043 | ,360* | ,258 | ,286 |
| | | Sig. (2-tailed) | ,112 | ,137 | ,779 | ,015 | ,087 | ,057 |
| | | N | 45 | 45 | 45 | 45 | 45 | 45 |
| | Lactobacillus_change | Correlation Coefficient | -,370* | -,099 | -,033 | ,159 | -,086 | -,006 |
| | | Sig. (2-tailed) | ,012 | ,519 | ,828 | ,297 | ,575 | ,969 |
| | | N | 45 | 45 | 45 | 45 | 45 | 45 |
| | Libanicoccus_change | Correlation Coefficient | ,053 | -,038 | ,071 | ,153 | ,051 | ,077 |
| | | Sig. (2-tailed) | ,732 | ,803 | ,643 | ,315 | ,741 | ,614 |
| | | N | 45 | 45 | 45 | 45 | 45 | 45 |

### Correlations

| | | Desulfovibrio_change | Eggerthella_change | Erysipelatoclostridium_change | Faecalitalea_change | Family.XIII.AD3011.group_change | Family.XIII.UCG.001_change |
|---|---|---|---|---|---|---|---|
| Gordonibacter_change | Correlation Coefficient | -,078 | ,384** | ,224 | ,183 | -,040 | -,023 |
| | Sig. (2-tailed) | ,610 | ,009 | ,140 | ,230 | ,792 | ,879 |
| | N | 45 | 45 | 45 | 45 | 45 | 45 |
| Holdemanella_change | Correlation Coefficient | -,030 | ,098 | ,088 | ,085 | ,227 | -,114 |
| | Sig. (2-tailed) | ,845 | ,521 | ,566 | ,581 | ,134 | ,458 |
| | N | 45 | 45 | 45 | 45 | 45 | 45 |
| Holdemania_change | Correlation Coefficient | ,003 | ,139 | -,060 | ,040 | -,016 | ,170 |
| | Sig. (2-tailed) | ,986 | ,362 | ,694 | ,794 | ,916 | ,263 |
| | N | 45 | 45 | 45 | 45 | 45 | 45 |
| Lachnospiraceae.FCS020.group_change | Correlation Coefficient | -,042 | ,020 | ,162 | -,221 | ,038 | ,064 |
| | Sig. (2-tailed) | ,782 | ,897 | ,288 | ,145 | ,802 | ,676 |
| | N | 45 | 45 | 45 | 45 | 45 | 45 |
| Lachnospiraceae.NK4A136.group_change | Correlation Coefficient | -,024 | -,054 | ,052 | -,278 | ,335* | ,207 |
| | Sig. (2-tailed) | ,873 | ,727 | ,736 | ,064 | ,024 | ,172 |
| | N | 45 | 45 | 45 | 45 | 45 | 45 |
| Lactiplantibacillus_change | Correlation Coefficient | -,018 | -,150 | -,149 | -,141 | ,185 | -,151 |
| | Sig. (2-tailed) | ,905 | ,326 | ,329 | ,355 | ,224 | ,322 |
| | N | 45 | 45 | 45 | 45 | 45 | 45 |
| Lactobacillus_change | Correlation Coefficient | ,201 | ,141 | -,121 | ,157 | -,149 | ,058 |
| | Sig. (2-tailed) | ,186 | ,356 | ,430 | ,303 | ,330 | ,705 |
| | N | 45 | 45 | 45 | 45 | 45 | 45 |
| Libanicoccus_change | Correlation Coefficient | ,338* | ,069 | ,019 | ,072 | -,323* | -,146 |
| | Sig. (2-tailed) | ,023 | ,652 | ,903 | ,638 | ,031 | ,337 |
| | N | 45 | 45 | 45 | 45 | 45 | 45 |

### Correlations

| | | | Gemella_change | Gordonibacter_change | Holdemanella_change | Holdemania_change | Lachnospiraceae.FCS020.group_change | Lachnospiraceae.NK4A136.group_change |
|---|---|---|---|---|---|---|---|---|
| | Gordonibacter_change | Correlation Coefficient | ,128 | 1,000 | ,166 | ,164 | -,040 | ,003 |
| | | Sig. (2-tailed) | ,402 | . | ,275 | ,283 | ,792 | ,985 |
| | | N | 45 | 45 | 45 | 45 | 45 | 45 |
| | Holdemanella_change | Correlation Coefficient | ,330* | ,166 | 1,000 | ,013 | ,367* | -,336* |
| | | Sig. (2-tailed) | ,027 | ,275 | . | ,934 | ,013 | ,024 |
| | | N | 45 | 45 | 45 | 45 | 45 | 45 |
| | Holdemania_change | Correlation Coefficient | -,465** | ,164 | ,013 | 1,000 | -,172 | ,161 |
| | | Sig. (2-tailed) | ,001 | ,283 | ,934 | . | ,257 | ,290 |
| | | N | 45 | 45 | 45 | 45 | 45 | 45 |
| | Lachnospiraceae.FCS020.group_change | Correlation Coefficient | ,300* | -,040 | ,367* | -,172 | 1,000 | -,205 |
| | | Sig. (2-tailed) | ,045 | ,792 | ,013 | ,257 | . | ,177 |
| | | N | 45 | 45 | 45 | 45 | 45 | 45 |
| | Lachnospiraceae.NK4A136.group_change | Correlation Coefficient | -,431** | ,003 | -,336* | ,161 | -,205 | 1,000 |
| | | Sig. (2-tailed) | ,003 | ,985 | ,024 | ,290 | ,177 | . |
| | | N | 45 | 45 | 45 | 45 | 45 | 45 |
| | Lactiplantibacillus_change | Correlation Coefficient | ,304* | -,077 | ,033 | -,103 | ,056 | -,185 |
| | | Sig. (2-tailed) | ,043 | ,617 | ,831 | ,500 | ,713 | ,225 |
| | | N | 45 | 45 | 45 | 45 | 45 | 45 |
| | Lactobacillus_change | Correlation Coefficient | ,020 | -,064 | -,100 | -,023 | -,047 | -,079 |
| | | Sig. (2-tailed) | ,899 | ,677 | ,515 | ,879 | ,757 | ,604 |
| | | N | 45 | 45 | 45 | 45 | 45 | 45 |
| | Libanicoccus_change | Correlation Coefficient | ,122 | ,034 | -,152 | ,160 | -,142 | -,229 |
| | | Sig. (2-tailed) | ,424 | ,822 | ,319 | ,292 | ,353 | ,130 |
| | | N | 45 | 45 | 45 | 45 | 45 | 45 |

### Correlations

| | | Lactiplantibacillus_change | Lactobacillus_change | Libanicoccus_change | Ligilactobacillus_change | Limosilactobacillus_change | Roseburia_change |
|---|---|---|---|---|---|---|---|
| Gordonibacter_change | Correlation Coefficient | -,077 | -,064 | ,034 | -,110 | ,153 | -,032 |
| | Sig. (2-tailed) | ,617 | ,677 | ,822 | ,471 | ,316 | ,834 |
| | N | 45 | 45 | 45 | 45 | 45 | 45 |
| Holdemanella_change | Correlation Coefficient | ,033 | -,100 | -,152 | -,017 | ,005 | -,121 |
| | Sig. (2-tailed) | ,831 | ,515 | ,319 | ,914 | ,972 | ,429 |
| | N | 45 | 45 | 45 | 45 | 45 | 45 |
| Holdemania_change | Correlation Coefficient | -,103 | -,023 | ,160 | ,046 | ,031 | ,134 |
| | Sig. (2-tailed) | ,500 | ,879 | ,292 | ,766 | ,841 | ,379 |
| | N | 45 | 45 | 45 | 45 | 45 | 45 |
| Lachnospiraceae.FCS020.group_change | Correlation Coefficient | ,056 | -,047 | -,142 | ,232 | -,109 | -,294 |
| | Sig. (2-tailed) | ,713 | ,757 | ,353 | ,125 | ,478 | ,050 |
| | N | 45 | 45 | 45 | 45 | 45 | 45 |
| Lachnospiraceae.NK4A136.group_change | Correlation Coefficient | -,185 | -,079 | -,229 | -,276 | -,141 | ,128 |
| | Sig. (2-tailed) | ,225 | ,604 | ,130 | ,066 | ,355 | ,401 |
| | N | 45 | 45 | 45 | 45 | 45 | 45 |
| Lactiplantibacillus_change | Correlation Coefficient | 1,000 | -,039 | -,036 | ,077 | ,124 | -,116 |
| | Sig. (2-tailed) | . | ,801 | ,814 | ,615 | ,416 | ,447 |
| | N | 45 | 45 | 45 | 45 | 45 | 45 |
| Lactobacillus_change | Correlation Coefficient | -,039 | 1,000 | ,186 | ,068 | ,364* | -,085 |
| | Sig. (2-tailed) | ,801 | . | ,220 | ,659 | ,014 | ,580 |
| | N | 45 | 45 | 45 | 45 | 45 | 45 |
| Libanicoccus_change | Correlation Coefficient | -,036 | ,186 | 1,000 | -,038 | ,060 | -,058 |
| | Sig. (2-tailed) | ,814 | ,220 | . | ,806 | ,696 | ,705 |
| | N | 45 | 45 | 45 | 45 | 45 | 45 |

### Correlations

| | | | Shuttleworthia_change | Subdoligranulum_change | UCG.003_change | X.Eubacterium..brachy.group_change | X.Ruminococcus..gauvreauii.group_change | X.Ruminococcus..torques.group_change |
|---|---|---|---|---|---|---|---|---|
| | Gordonibacter_change | Correlation Coefficient | ,055 | ,172 | ,223 | ,415** | -,214 | -,156 |
| | | Sig. (2-tailed) | ,720 | ,258 | ,141 | ,005 | ,158 | ,305 |
| | | N | 45 | 45 | 45 | 45 | 45 | 45 |
| | Holdemanella_change | Correlation Coefficient | -,001 | -,056 | -,306* | ,278 | -,057 | ,204 |
| | | Sig. (2-tailed) | ,996 | ,714 | ,041 | ,064 | ,709 | ,179 |
| | | N | 45 | 45 | 45 | 45 | 45 | 45 |
| | Holdemania_change | Correlation Coefficient | ,144 | -,059 | ,323* | ,009 | -,214 | -,192 |
| | | Sig. (2-tailed) | ,346 | ,702 | ,030 | ,953 | ,158 | ,206 |
| | | N | 45 | 45 | 45 | 45 | 45 | 45 |
| | Lachnospiraceae.FCS020.group_change | Correlation Coefficient | -,029 | ,214 | -,251 | ,108 | -,127 | ,105 |
| | | Sig. (2-tailed) | ,848 | ,158 | ,096 | ,478 | ,405 | ,493 |
| | | N | 45 | 45 | 45 | 45 | 45 | 45 |
| | Lachnospiraceae.NK4A136.group_change | Correlation Coefficient | ,009 | -,050 | ,227 | -,157 | ,083 | -,207 |
| | | Sig. (2-tailed) | ,954 | ,745 | ,133 | ,303 | ,587 | ,173 |
| | | N | 45 | 45 | 45 | 45 | 45 | 45 |
| | Lactiplantibacillus_change | Correlation Coefficient | ,071 | -,040 | -,063 | -,026 | ,178 | -,047 |
| | | Sig. (2-tailed) | ,645 | ,795 | ,680 | ,865 | ,241 | ,760 |
| | | N | 45 | 45 | 45 | 45 | 45 | 45 |
| | Lactobacillus_change | Correlation Coefficient | ,103 | -,337* | -,049 | -,032 | ,044 | -,111 |
| | | Sig. (2-tailed) | ,502 | ,024 | ,748 | ,837 | ,773 | ,466 |
| | | N | 45 | 45 | 45 | 45 | 45 | 45 |
| | Libanicoccus_change | Correlation Coefficient | -,018 | -,245 | ,026 | ,087 | -,159 | -,241 |
| | | Sig. (2-tailed) | ,909 | ,105 | ,863 | ,568 | ,297 | ,110 |
| | | N | 45 | 45 | 45 | 45 | 45 | 45 |

### Correlations

| | | interaction_VTA_change | interaction_rOFC_change | interaction_rmOFC_change | sumscore_GIQLI_change | Stool_frequency_change | STGHRE_change |
|---|---|---|---|---|---|---|---|
| Ligilactobacillus_change | Correlation Coefficient | -,175 | -,105 | -,082 | ,054 | ,060 | -,017 |
| | Sig. (2-tailed) | ,262 | ,521 | ,603 | ,727 | ,703 | ,914 |
| | N | 43 | 40 | 43 | 44 | 43 | 42 |
| Limosilactobacillus_change | Correlation Coefficient | -,010 | -,005 | -,070 | ,058 | ,169 | -,196 |
| | Sig. (2-tailed) | ,949 | ,976 | ,654 | ,706 | ,278 | ,212 |
| | N | 43 | 40 | 43 | 44 | 43 | 42 |
| Roseburia_change | Correlation Coefficient | ,137 | ,146 | -,011 | ,025 | -,316* | -,209 |
| | Sig. (2-tailed) | ,380 | ,369 | ,944 | ,870 | ,039 | ,184 |
| | N | 43 | 40 | 43 | 44 | 43 | 42 |
| Shuttleworthia_change | Correlation Coefficient | ,191 | -,155 | ,032 | ,210 | ,115 | -,164 |
| | Sig. (2-tailed) | ,220 | ,338 | ,838 | ,172 | ,462 | ,300 |
| | N | 43 | 40 | 43 | 44 | 43 | 42 |
| Subdoligranulum_change | Correlation Coefficient | -,384* | -,159 | -,015 | ,261 | ,083 | ,279 |
| | Sig. (2-tailed) | ,011 | ,329 | ,922 | ,087 | ,598 | ,073 |
| | N | 43 | 40 | 43 | 44 | 43 | 42 |
| UCG.003_change | Correlation Coefficient | ,185 | ,122 | -,094 | -,255 | -,067 | -,059 |
| | Sig. (2-tailed) | ,236 | ,453 | ,551 | ,095 | ,669 | ,711 |
| | N | 43 | 40 | 43 | 44 | 43 | 42 |
| X.Eubacterium..brachy. group_change | Correlation Coefficient | ,115 | -,177 | ,038 | ,090 | -,196 | -,097 |
| | Sig. (2-tailed) | ,464 | ,275 | ,809 | ,561 | ,209 | ,540 |
| | N | 43 | 40 | 43 | 44 | 43 | 42 |
| X.Ruminococcus.. gauvreauii.group_change | Correlation Coefficient | -,194 | -,143 | -,024 | ,113 | -,091 | -,100 |
| | Sig. (2-tailed) | ,211 | ,377 | ,879 | ,465 | ,560 | ,528 |
| | N | 43 | 40 | 43 | 44 | 43 | 42 |

### Correlations

| | | | STPYY_change | STGLPT_change | age | BMI | FM_stand | Richness | shannon.effective |
|---|---|---|---|---|---|---|---|---|---|
| | Ligilactobacillus_change | Correlation Coefficient | ,015 | ,038 | -,220 | ,072 | -,144 | -,045 | ,004 |
| | | Sig. (2-tailed) | ,924 | ,813 | ,146 | ,637 | ,345 | ,768 | ,979 |
| | | N | 42 | 42 | 45 | 45 | 45 | 45 | 45 |
| | Limosilactobacillus_change | Correlation Coefficient | ,027 | -,182 | ,280 | -,257 | -,212 | -,141 | -,098 |
| | | Sig. (2-tailed) | ,864 | ,248 | ,062 | ,088 | ,162 | ,354 | ,523 |
| | | N | 42 | 42 | 45 | 45 | 45 | 45 | 45 |
| | Roseburia_change | Correlation Coefficient | -,048 | -,035 | ,323* | -,120 | ,182 | ,123 | ,096 |
| | | Sig. (2-tailed) | ,763 | ,824 | ,030 | ,434 | ,232 | ,419 | ,532 |
| | | N | 42 | 42 | 45 | 45 | 45 | 45 | 45 |
| | Shuttleworthia_change | Correlation Coefficient | -,068 | -,074 | -,034 | ,016 | ,312* | -,034 | -,090 |
| | | Sig. (2-tailed) | ,669 | ,644 | ,825 | ,915 | ,037 | ,826 | ,556 |
| | | N | 42 | 42 | 45 | 45 | 45 | 45 | 45 |
| | Subdoligranulum_change | Correlation Coefficient | -,210 | ,009 | -,033 | ,175 | -,188 | -,033 | ,008 |
| | | Sig. (2-tailed) | ,182 | ,957 | ,827 | ,252 | ,216 | ,831 | ,956 |
| | | N | 42 | 42 | 45 | 45 | 45 | 45 | 45 |
| | UCG.003_change | Correlation Coefficient | -,176 | -,165 | ,072 | ,239 | ,208 | ,052 | ,083 |
| | | Sig. (2-tailed) | ,264 | ,296 | ,638 | ,113 | ,170 | ,736 | ,587 |
| | | N | 42 | 42 | 45 | 45 | 45 | 45 | 45 |
| | X.Eubacterium..brachy.group_change | Correlation Coefficient | -,061 | ,179 | -,090 | -,048 | ,150 | ,244 | ,227 |
| | | Sig. (2-tailed) | ,702 | ,256 | ,554 | ,753 | ,324 | ,106 | ,133 |
| | | N | 42 | 42 | 45 | 45 | 45 | 45 | 45 |
| | X.Ruminococcus..gauvreauii.group_change | Correlation Coefficient | -,063 | -,159 | ,071 | -,050 | ,196 | -,346* | -,288 |
| | | Sig. (2-tailed) | ,693 | ,314 | ,642 | ,744 | ,196 | ,020 | ,055 |
| | | N | 42 | 42 | 45 | 45 | 45 | 45 | 45 |

### Correlations

| | | simpson. effective | Evenness | CHOL_change | LDL_change | shannon. effective_change | Richness_change |
|---|---|---|---|---|---|---|---|
| Ligilactobacillus_change | Correlation Coefficient | ,017 | ,145 | -,061 | ,011 | ,006 | -,101 |
| | Sig. (2-tailed) | ,914 | ,342 | ,689 | ,941 | ,969 | ,508 |
| | N | 45 | 45 | 45 | 45 | 45 | 45 |
| Limosilactobacillus_change | Correlation Coefficient | -,108 | ,031 | -,076 | -,056 | ,038 | ,025 |
| | Sig. (2-tailed) | ,480 | ,839 | ,622 | ,713 | ,803 | ,872 |
| | N | 45 | 45 | 45 | 45 | 45 | 45 |
| Roseburia_change | Correlation Coefficient | ,054 | -,126 | ,028 | -,021 | ,107 | ,211 |
| | Sig. (2-tailed) | ,726 | ,409 | ,857 | ,893 | ,485 | ,165 |
| | N | 45 | 45 | 45 | 45 | 45 | 45 |
| Shuttleworthia_change | Correlation Coefficient | -,080 | -,186 | ,155 | ,231 | -,047 | ,056 |
| | Sig. (2-tailed) | ,603 | ,222 | ,308 | ,126 | ,758 | ,717 |
| | N | 45 | 45 | 45 | 45 | 45 | 45 |
| Subdoligranulum_change | Correlation Coefficient | ,101 | ,322* | -,110 | -,104 | ,162 | ,054 |
| | Sig. (2-tailed) | ,511 | ,031 | ,473 | ,496 | ,288 | ,722 |
| | N | 45 | 45 | 45 | 45 | 45 | 45 |
| UCG.003_change | Correlation Coefficient | ,035 | ,030 | ,006 | -,018 | ,051 | ,088 |
| | Sig. (2-tailed) | ,818 | ,844 | ,971 | ,907 | ,738 | ,567 |
| | N | 45 | 45 | 45 | 45 | 45 | 45 |
| X.Eubacterium..brachy. group_change | Correlation Coefficient | ,229 | ,073 | ,000 | -,050 | ,281 | ,364* |
| | Sig. (2-tailed) | ,130 | ,632 | 1,000 | ,745 | ,061 | ,014 |
| | N | 45 | 45 | 45 | 45 | 45 | 45 |
| X.Ruminococcus.. gauvreauii.group_change | Correlation Coefficient | -,249 | ,176 | ,241 | ,241 | ,010 | -,028 |
| | Sig. (2-tailed) | ,099 | ,247 | ,110 | ,111 | ,950 | ,854 |
| | N | 45 | 45 | 45 | 45 | 45 | 45 |

### Correlations

| | | | Evenness_change | Actinomyces_change | Aerostipes_change | Bifidobacterium_change | Blautia_change | Collinsella_change |
|---|---|---|---|---|---|---|---|---|
| | Ligilactobacillus_change | Correlation Coefficient | ,320* | ,003 | -,024 | ,072 | ,133 | ,218 |
| | | Sig. (2-tailed) | ,032 | ,982 | ,874 | ,640 | ,384 | ,150 |
| | | N | 45 | 45 | 45 | 45 | 45 | 45 |
| | Limosilactobacillus_change | Correlation Coefficient | -,008 | -,038 | ,228 | ,077 | ,093 | ,132 |
| | | Sig. (2-tailed) | ,958 | ,806 | ,132 | ,616 | ,542 | ,388 |
| | | N | 45 | 45 | 45 | 45 | 45 | 45 |
| | Roseburia_change | Correlation Coefficient | -,188 | -,040 | -,111 | ,142 | -,313* | -,219 |
| | | Sig. (2-tailed) | ,216 | ,792 | ,468 | ,353 | ,037 | ,149 |
| | | N | 45 | 45 | 45 | 45 | 45 | 45 |
| | Shuttleworthia_change | Correlation Coefficient | -,228 | -,089 | -,052 | ,255 | -,246 | ,092 |
| | | Sig. (2-tailed) | ,132 | ,561 | ,735 | ,091 | ,104 | ,546 |
| | | N | 45 | 45 | 45 | 45 | 45 | 45 |
| | Subdoligranulum_change | Correlation Coefficient | ,348* | ,121 | ,163 | -,264 | ,239 | ,025 |
| | | Sig. (2-tailed) | ,019 | ,428 | ,285 | ,079 | ,114 | ,869 |
| | | N | 45 | 45 | 45 | 45 | 45 | 45 |
| | UCG.003_change | Correlation Coefficient | -,141 | -,185 | -,084 | -,189 | -,396** | -,109 |
| | | Sig. (2-tailed) | ,357 | ,225 | ,582 | ,215 | ,007 | ,474 |
| | | N | 45 | 45 | 45 | 45 | 45 | 45 |
| | X.Eubacterium..brachy. group_change | Correlation Coefficient | -,061 | ,205 | ,134 | -,085 | ,195 | -,063 |
| | | Sig. (2-tailed) | ,689 | ,178 | ,381 | ,580 | ,199 | ,682 |
| | | N | 45 | 45 | 45 | 45 | 45 | 45 |
| | X.Ruminococcus.. gauvreauii.group_change | Correlation Coefficient | ,121 | ,018 | -,190 | ,233 | ,209 | ,005 |
| | | Sig. (2-tailed) | ,428 | ,908 | ,211 | ,124 | ,169 | ,976 |
| | | N | 45 | 45 | 45 | 45 | 45 | 45 |

### Correlations

| | | Desulfovibrio_change | Eggerthella_change | Erysipelatoclostridium_change | Faecalitalea_change | Family.XIII. AD3011. group_change | Family.XIII. UCG. 001_change |
|---|---|---|---|---|---|---|---|
| Ligilactobacillus_change | Correlation Coefficient | -,054 | -,030 | ,245 | -,043 | ,035 | ,222 |
| | Sig. (2-tailed) | ,725 | ,845 | ,104 | ,778 | ,821 | ,142 |
| | N | 45 | 45 | 45 | 45 | 45 | 45 |
| Limosilactobacillus_change | Correlation Coefficient | ,258 | -,057 | -,069 | ,036 | -,104 | ,128 |
| | Sig. (2-tailed) | ,087 | ,712 | ,652 | ,812 | ,496 | ,402 |
| | N | 45 | 45 | 45 | 45 | 45 | 45 |
| Roseburia_change | Correlation Coefficient | ,130 | -,037 | ,107 | ,126 | -,224 | ,120 |
| | Sig. (2-tailed) | ,393 | ,807 | ,484 | ,411 | ,139 | ,432 |
| | N | 45 | 45 | 45 | 45 | 45 | 45 |
| Shuttleworthia_change | Correlation Coefficient | ,020 | ,146 | ,078 | -,141 | -,048 | -,280 |
| | Sig. (2-tailed) | ,894 | ,338 | ,613 | ,356 | ,752 | ,062 |
| | N | 45 | 45 | 45 | 45 | 45 | 45 |
| Subdoligranulum_change | Correlation Coefficient | -,113 | ,022 | ,064 | ,039 | ,090 | -,025 |
| | Sig. (2-tailed) | ,462 | ,886 | ,676 | ,801 | ,555 | ,872 |
| | N | 45 | 45 | 45 | 45 | 45 | 45 |
| UCG.003_change | Correlation Coefficient | -,052 | -,180 | ,000 | -,038 | -,018 | ,070 |
| | Sig. (2-tailed) | ,735 | ,237 | ,999 | ,804 | ,905 | ,645 |
| | N | 45 | 45 | 45 | 45 | 45 | 45 |
| X.Eubacterium..brachy. group_change | Correlation Coefficient | ,100 | ,125 | ,156 | ,302* | -,084 | ,068 |
| | Sig. (2-tailed) | ,513 | ,412 | ,307 | ,044 | ,585 | ,655 |
| | N | 45 | 45 | 45 | 45 | 45 | 45 |
| X.Ruminococcus.. gauvreauii.group_change | Correlation Coefficient | -,043 | ,102 | ,038 | -,074 | -,026 | -,256 |
| | Sig. (2-tailed) | ,778 | ,505 | ,803 | ,629 | ,867 | ,089 |
| | N | 45 | 45 | 45 | 45 | 45 | 45 |

### Correlations

| | | Gemella_change | Gordonibacter_change | Holdemanella_change | Holdemania_change | Lachnospiraceae.FCS020.group_change | Lachnospiraceae.NK4A136.group_change |
|---|---|---|---|---|---|---|---|
| Ligilactobacillus_change | Correlation Coefficient | ,093 | -,110 | -,017 | ,046 | ,232 | -,276 |
| | Sig. (2-tailed) | ,545 | ,471 | ,914 | ,766 | ,125 | ,066 |
| | N | 45 | 45 | 45 | 45 | 45 | 45 |
| Limosilactobacillus_change | Correlation Coefficient | ,253 | ,153 | ,005 | ,031 | -,109 | -,141 |
| | Sig. (2-tailed) | ,094 | ,316 | ,972 | ,841 | ,478 | ,355 |
| | N | 45 | 45 | 45 | 45 | 45 | 45 |
| Roseburia_change | Correlation Coefficient | -,094 | -,032 | -,121 | ,134 | -,294 | ,128 |
| | Sig. (2-tailed) | ,541 | ,834 | ,429 | ,379 | ,050 | ,401 |
| | N | 45 | 45 | 45 | 45 | 45 | 45 |
| Shuttleworthia_change | Correlation Coefficient | -,146 | ,055 | -,001 | ,144 | -,029 | ,009 |
| | Sig. (2-tailed) | ,339 | ,720 | ,996 | ,346 | ,848 | ,954 |
| | N | 45 | 45 | 45 | 45 | 45 | 45 |
| Subdoligranulum_change | Correlation Coefficient | ,072 | ,172 | -,056 | -,059 | ,214 | -,050 |
| | Sig. (2-tailed) | ,636 | ,258 | ,714 | ,702 | ,158 | ,745 |
| | N | 45 | 45 | 45 | 45 | 45 | 45 |
| UCG.003_change | Correlation Coefficient | -,274 | ,223 | -,306* | ,323* | -,251 | ,227 |
| | Sig. (2-tailed) | ,069 | ,141 | ,041 | ,030 | ,096 | ,133 |
| | N | 45 | 45 | 45 | 45 | 45 | 45 |
| X.Eubacterium..brachy.group_change | Correlation Coefficient | ,233 | ,415** | ,278 | ,009 | ,108 | -,157 |
| | Sig. (2-tailed) | ,123 | ,005 | ,064 | ,953 | ,478 | ,303 |
| | N | 45 | 45 | 45 | 45 | 45 | 45 |
| X.Ruminococcus..gauvreauii.group_change | Correlation Coefficient | ,000 | -,214 | -,057 | -,214 | -,127 | ,083 |
| | Sig. (2-tailed) | ,998 | ,158 | ,709 | ,158 | ,405 | ,587 |
| | N | 45 | 45 | 45 | 45 | 45 | 45 |

### Correlations

| | | | Lactiplantibacillus_change | Lactobacillus_change | Libanicoccus_change | Ligilactobacillus_change | Limosilactobacillus_change | Roseburia_change |
|---|---|---|---|---|---|---|---|---|
| | Ligilactobacillus_change | Correlation Coefficient | ,077 | ,068 | -,038 | 1,000 | ,039 | -,232 |
| | | Sig. (2-tailed) | ,615 | ,659 | ,806 | . | ,799 | ,125 |
| | | N | 45 | 45 | 45 | 45 | 45 | 45 |
| | Limosilactobacillus_change | Correlation Coefficient | ,124 | ,364* | ,060 | ,039 | 1,000 | ,063 |
| | | Sig. (2-tailed) | ,416 | ,014 | ,696 | ,799 | . | ,681 |
| | | N | 45 | 45 | 45 | 45 | 45 | 45 |
| | Roseburia_change | Correlation Coefficient | -,116 | -,085 | -,058 | -,232 | ,063 | 1,000 |
| | | Sig. (2-tailed) | ,447 | ,580 | ,705 | ,125 | ,681 | . |
| | | N | 45 | 45 | 45 | 45 | 45 | 45 |
| | Shuttleworthia_change | Correlation Coefficient | ,071 | ,103 | -,018 | -,075 | ,067 | ,080 |
| | | Sig. (2-tailed) | ,645 | ,502 | ,909 | ,623 | ,661 | ,603 |
| | | N | 45 | 45 | 45 | 45 | 45 | 45 |
| | Subdoligranulum_change | Correlation Coefficient | -,040 | -,337* | -,245 | ,258 | -,188 | -,247 |
| | | Sig. (2-tailed) | ,795 | ,024 | ,105 | ,088 | ,217 | ,102 |
| | | N | 45 | 45 | 45 | 45 | 45 | 45 |
| | UCG.003_change | Correlation Coefficient | -,063 | -,049 | ,026 | -,174 | -,070 | ,239 |
| | | Sig. (2-tailed) | ,680 | ,748 | ,863 | ,253 | ,648 | ,114 |
| | | N | 45 | 45 | 45 | 45 | 45 | 45 |
| | X.Eubacterium..brachy. group_change | Correlation Coefficient | -,026 | -,032 | ,087 | -,023 | ,038 | -,006 |
| | | Sig. (2-tailed) | ,865 | ,837 | ,568 | ,880 | ,803 | ,967 |
| | | N | 45 | 45 | 45 | 45 | 45 | 45 |
| | X.Ruminococcus.. gauvreauii.group_change | Correlation Coefficient | ,178 | ,044 | -,159 | ,026 | -,040 | ,033 |
| | | Sig. (2-tailed) | ,241 | ,773 | ,297 | ,864 | ,794 | ,831 |
| | | N | 45 | 45 | 45 | 45 | 45 | 45 |

### Correlations

| | | | Shuttleworthia_change | Subdoligranulum_change | UCG.003_change | X.Eubacterium..brachy.group_change | X.Ruminococcus..gauvreauii.group_change | X.Ruminococcus..torques.group_change |
|---|---|---|---|---|---|---|---|---|
| | Ligilactobacillus_change | Correlation Coefficient | -,075 | ,258 | -,174 | -,023 | ,026 | ,101 |
| | | Sig. (2-tailed) | ,623 | ,088 | ,253 | ,880 | ,864 | ,511 |
| | | N | 45 | 45 | 45 | 45 | 45 | 45 |
| | Limosilactobacillus_change | Correlation Coefficient | ,067 | -,188 | -,070 | ,038 | -,040 | -,198 |
| | | Sig. (2-tailed) | ,661 | ,217 | ,648 | ,803 | ,794 | ,192 |
| | | N | 45 | 45 | 45 | 45 | 45 | 45 |
| | Roseburia_change | Correlation Coefficient | ,080 | -,247 | ,239 | -,006 | ,033 | -,192 |
| | | Sig. (2-tailed) | ,603 | ,102 | ,114 | ,967 | ,831 | ,205 |
| | | N | 45 | 45 | 45 | 45 | 45 | 45 |
| | Shuttleworthia_change | Correlation Coefficient | 1,000 | -,188 | -,004 | ,083 | -,090 | -,069 |
| | | Sig. (2-tailed) | . | ,215 | ,980 | ,589 | ,554 | ,652 |
| | | N | 45 | 45 | 45 | 45 | 45 | 45 |
| | Subdoligranulum_change | Correlation Coefficient | -,188 | 1,000 | -,157 | -,039 | ,286 | ,201 |
| | | Sig. (2-tailed) | ,215 | . | ,302 | ,797 | ,057 | ,185 |
| | | N | 45 | 45 | 45 | 45 | 45 | 45 |
| | UCG.003_change | Correlation Coefficient | -,004 | -,157 | 1,000 | -,044 | -,265 | -,349* |
| | | Sig. (2-tailed) | ,980 | ,302 | . | ,774 | ,078 | ,019 |
| | | N | 45 | 45 | 45 | 45 | 45 | 45 |
| | X.Eubacterium..brachy.group_change | Correlation Coefficient | ,083 | -,039 | -,044 | 1,000 | -,241 | ,086 |
| | | Sig. (2-tailed) | ,589 | ,797 | ,774 | . | ,111 | ,573 |
| | | N | 45 | 45 | 45 | 45 | 45 | 45 |
| | X.Ruminococcus..gauvreauii.group_change | Correlation Coefficient | -,090 | ,286 | -,265 | -,241 | 1,000 | -,016 |
| | | Sig. (2-tailed) | ,554 | ,057 | ,078 | ,111 | . | ,915 |
| | | N | 45 | 45 | 45 | 45 | 45 | 45 |

### Correlations

| | | interaction_VTA_change | interaction_rOFC_change | interaction_rmOFC_change | sumscore_GIQLI_change | Stool_frequency_change | STGHRE_change |
|---|---|---|---|---|---|---|---|
| X.Ruminococcus..torques.group_change | Correlation Coefficient | -,229 | -,194 | -,133 | ,305* | ,271 | ,230 |
| | Sig. (2-tailed) | ,140 | ,230 | ,397 | ,044 | ,079 | ,144 |
| | N | 43 | 40 | 43 | 44 | 43 | 42 |

### Correlations

| | | STPYY_change | STGLPT_change | age | BMI | FM_stand | Richness | shannon.effective |
|---|---|---|---|---|---|---|---|---|
| X.Ruminococcus..torques.group_change | Correlation Coefficient | ,183 | ,092 | -,302* | ,053 | ,019 | ,079 | ,122 |
| | Sig. (2-tailed) | ,247 | ,561 | ,044 | ,728 | ,904 | ,607 | ,425 |
| | N | 42 | 42 | 45 | 45 | 45 | 45 | 45 |

### Correlations

| | | simpson.effective | Evenness | CHOL_change | LDL_change | shannon.effective_change | Richness_change |
|---|---|---|---|---|---|---|---|
| X.Ruminococcus..torques.group_change | Correlation Coefficient | ,163 | ,204 | -,067 | -,037 | ,083 | ,019 |
| | Sig. (2-tailed) | ,284 | ,179 | ,663 | ,812 | ,586 | ,904 |
| | N | 45 | 45 | 45 | 45 | 45 | 45 |

### Correlations

| | | Evenness_change | Actinomyces_change | Aerostipes_change | Bifidobacterium_change | Blautia_change | Collinsella_change |
|---|---|---|---|---|---|---|---|
| X.Ruminococcus..torques.group_change | Correlation Coefficient | ,242 | ,286 | ,134 | -,128 | ,288 | ,158 |
| | Sig. (2-tailed) | ,109 | ,056 | ,380 | ,402 | ,055 | ,300 |
| | N | 45 | 45 | 45 | 45 | 45 | 45 |

### Correlations

| | | Desulfovibrio_change | Eggerthella_change | Erysipelatoclostridium_change | Faecalitalea_change | Family.XIII.AD3011.group_change | Family.XIII.UCG.001_change |
|---|---|---|---|---|---|---|---|
| X.Ruminococcus..torques.group_change | Correlation Coefficient | -,151 | -,038 | ,060 | ,190 | ,281 | ,041 |
| | Sig. (2-tailed) | ,323 | ,804 | ,698 | ,211 | ,062 | ,791 |
| | N | 45 | 45 | 45 | 45 | 45 | 45 |

### Correlations

| | | Gemella_change | Gordonibacter_change | Holdemanella_change | Holdemania_change | Lachnospiraceae.FCS020.group_change | Lachnospiraceae.NK4A136.group_change |
|---|---|---|---|---|---|---|---|
| X.Ruminococcus..torques.group_change | Correlation Coefficient | ,082 | -,156 | ,204 | -,192 | ,105 | -,207 |
| | Sig. (2-tailed) | ,594 | ,305 | ,179 | ,206 | ,493 | ,173 |
| | N | 45 | 45 | 45 | 45 | 45 | 45 |

### Correlations

| | | Lactiplantibacillus_change | Lactobacillus_change | Libanicoccus_change | Ligilactobacillus_change | Limosilactobacillus_change | Roseburia_change |
|---|---|---|---|---|---|---|---|
| X.Ruminococcus..torques. group_change | Correlation Coefficient | -,047 | -,111 | -,241 | ,101 | -,198 | -,192 |
| | Sig. (2-tailed) | ,760 | ,466 | ,110 | ,511 | ,192 | ,205 |
| | N | 45 | 45 | 45 | 45 | 45 | 45 |

### Correlations

| | | Shuttleworthia_change | Subdoligranulum_change | UCG. 003_change | X.Eubacterium.. brachy. group_change | X. Ruminococcus. .gauvreauii. group_change | X. Ruminococcus. .torques. group_change |
|---|---|---|---|---|---|---|---|
| X.Ruminococcus..torques. group_change | Correlation Coefficient | -,069 | ,201 | -,349* | ,086 | -,016 | 1,000 |
| | Sig. (2-tailed) | ,652 | ,185 | ,019 | ,573 | ,915 | . |
| | N | 45 | 45 | 45 | 45 | 45 | 45 |

*. Correlation is significant at the 0.05 level (2-tailed).

**. Correlation is significant at the 0.01 level (2-tailed).
