## Supplementary material for "A prebiotic diet changes neural correlates of food decision-making in overweight adults: a randomized controlled within-subject cross-over trial": SI_behav

Additional behavioral assessments

Liking ratings of food and art stimuli were collected similar to wanting ratings, yet outside the MR scanner and after all pre- and post-intervention visits. In one session (about 1h30min), 720 stimuli were presented (all from the wanting task across all sessions, plus additional ones) in the same technical manner as described above. Participants were asked “How much do you like this in general” (German original: “Wie sehr mögen Sie dies generell?”) and responded by moving a trial-by-trial randomly placed cursor on a 8-point Likert scale (1 = “not at all”, 8 = “absolutely”; German original: 1 = “überhaupt nicht”, 8 = “äußert gern”) with arrow buttons. Diverging from the MR setting, here each stimuli was presented up to 10s and participants could actively confirm their rating choice by clicking the space bar. Participants were explicitly instructed to report general liking, and that no post-experiment reward was provided. Time of day was not standardized and fasted state not acquired for this day.

*[Detailed instructions: “Sie können eine Wertung zwischen 1 und 8 auswählen. 1 bedeutet, dass Sie das Lebensmittel so abstoßend finden, dass Sie es unter keinen Umständen essen würden, und 8 bedeutet, dass Sie das Lebensmittel so lecker finden, dass Sie es jederzeit sehr gerne essen würden. Für die Kunstbilder bedeutet 1, dass Sie das Bild so hässlich finden, dass Sie es nicht ansehen möchten, und 8 bedeutet, dass Sie es so schön finden, dass Sie den Blick nicht abwenden möchten. Zum Fortfahren bitte LEERTASTE drücken.”]*

Food intake. The DEGS-1 German Food Frequency Questionnaire (FFQ) [1] was used to assess habitual dietary intake for the last 24h and the last 7 days at each timepoint. We developed a pipeline to compute daily nutrient intake based on self-reported dietary habits [2]. We did this by combining computed mean daily portion [g] based on DEGS-1 FFQ and corresponding nutrient information based on reference nutrient data (using the German Nutrient Reference Database “Bundeslebensmittelschlüssel” version 3.02) for each of the 53 items. This resulted in mean daily intake of macro- and micronutrients, e.g. daily fiber intake in grams.

Traits. The following questionnaires were administered once for each individual at the pre-baseline assessment: personality traits (NEOFFI-30) [3], Three-Factor Eating Questionnaire (TFEQ) [4], Eating Disorder Examination Questionnaire (EDEQ) [5], art knowledge (VAIAK) [6], physical activity (IPAQ), general well-being (8-item Eurohis QoL and 5-item WHO-5), trait anxiety (STADI-T) [7] and impulsivity (BIS-15) [8].

States. The following questionnaires were administered at each intervention visit: sleep quality of the last 24h and last 7 days (SF-A/R, SF-B/R) [9], gastrointestinal quality of life (GIQLI) [10], personality states (BFMM), changes to physical activity, depressive symptoms (Beck Depression Inventory, BDI) [11], well-being (WHO-5), state anxiety (STADI-S) [7], mood (POMS) [12], affect (PANAS) [13].

**Behavioral hypotheses** **and codes**

H_behav_0: Food is more wanted than art (between-subject)

Main effect of stimulus category (food vs. art)

R1 <- lmer(wanting ~ timepoint+stim_cat+(1|subj), data=food_and_art_wanting)

R0 <- lmer(wanting ~ timepoint+(1|subj), data=food_and_art_wanting)

Main effect of stimulus category (food vs. art) dependent on hunger ratings.

Note: The mean of pre- and post-task subjective hunger ratings (VAS) were used.

R2 <- lmer(wanting ~ stim_cat*hunger+timepoint+stim_cat+hunger+(1|subj), data=food_and_art_wanting)

R0 <- lmer(wanting ~ hunger+timepoint+stim_cat+(1|subj), data=food_and_art_wanting)

H_behav_A: Overall intervention effect on wanting (food vs. art)

R1 <- lmer(wanting ~ timepoint*intervention*stim_cat+timepoint*stim_cat+timepoint*intervention+stim_cat*intervention+timepoint+stim_cat+intervention+(1|subj), data=food_and_art_wanting)

R0 <- lmer(wanting ~ timepoint*stim_cat+timepoint*intervention+stim_cat*intervention+timepoint+stim_cat+intervention+(1|subj), data=food_and_art_wanting)

H_behav_B: Caloric content effect on wanting (food only)

R1 <- lmer(wanting ~ timepoint*intervention*kcal_100g+intervention*kcal_100g+timepoint*kcal_100g+timepoint*intervention+intervention+timepoint+kcal_100g+(1|subj), data=food_wanting)

R0 <- lmer(wanting ~ intervention*kcal_100g+timepoint*kcal_100g+timepoint*intervention+intervention+timepoint+kcal_100g+(1|subj), data=food_wanting)

Note: We computed the same models (R1 and R0) for fiber_100g instead of kcal_100g. Analysis part B includes food pictures only.

H_behav_C: Influence of liking on wanting (extended H_behav_A)

R1 <- lmer(wanting ~ timepoint*intervention*stim_cat+timepoint*stim_cat+timepoint*intervention+stim_cat*intervention+liking+timepoint+stim_cat+intervention+(1|subj), data=food_and_art_wanting)

R0 <- lmer(wanting ~ timepoint*stim_cat+timepoint*intervention+stim_cat*intervention+timepoint+stim_cat+intervention+(1|subj), data=food_and_art_wanting)

Note that all models residuals were normally distributed, if not stated otherwise.

Behavioral Results

Table 1: Wanting ratings by stimulus category and stimulus type by timepoint for each intervention arm. Based on means of individuals for each stimulus type. sd = standard deviation.

| **timepoint** | **intervention** | **stim_category** | **stim_type** | **variable** | **n** | **mean** | **sd** |
| --- | --- | --- | --- | --- | --- | --- | --- |
| **BL** | placebo | F | cal1 | wanting | 53 | 3.394 | 1.15 |
| **BL** | placebo | F | cal2 | wanting | 53 | 4.173 | 1.078 |
| **BL** | placebo | F | cal3 | wanting | 53 | 3.844 | 1.107 |
| **BL** | placebo | F | cal4 | wanting | 53 | 3.521 | 1.155 |
| **BL** | placebo | NF | animals | wanting | 53 | 3.662 | 1.164 |
| **BL** | placebo | NF | plants | wanting | 53 | 3.245 | 1.181 |
| **BL** | placebo | NF | objects | wanting | 53 | 2.401 | 0.87 |
| **BL** | fiber | F | cal1 | wanting | 55 | 3.532 | 1.235 |
| **BL** | fiber | F | cal2 | wanting | 55 | 4.204 | 1.221 |
| **BL** | fiber | F | cal3 | wanting | 55 | 3.797 | 1.21 |
| **BL** | fiber | F | cal4 | wanting | 55 | 3.724 | 1.168 |
| **BL** | fiber | NF | animals | wanting | 55 | 3.608 | 1.323 |
| **BL** | fiber | NF | plants | wanting | 55 | 3.562 | 1.378 |
| **BL** | fiber | NF | objects | wanting | 55 | 2.399 | 0.91 |
| **FU** | placebo | F | cal1 | wanting | 49 | 3.294 | 1.286 |
| **FU** | placebo | F | cal2 | wanting | 49 | 3.991 | 1.288 |
| **FU** | placebo | F | cal3 | wanting | 49 | 3.653 | 1.059 |
| **FU** | placebo | F | cal4 | wanting | 49 | 3.391 | 1.112 |
| **FU** | placebo | NF | animals | wanting | 49 | 3.422 | 1.217 |
| **FU** | placebo | NF | plants | wanting | 49 | 3.329 | 1.412 |
| **FU** | placebo | NF | objects | wanting | 49 | 2.369 | 947 |
| **FU** | fiber | F | cal1 | wanting | 47 | 3.212 | 1.218 |
| **FU** | fiber | F | cal2 | wanting | 47 | 4.074 | 1.34 |
| **FU** | fiber | F | cal3 | wanting | 47 | 3.755 | 1.186 |
| **FU** | fiber | F | cal4 | wanting | 47 | 3.398 | 1.116 |
| **FU** | fiber | NF | animals | wanting | 47 | 3.447 | 1.338 |
| **FU** | fiber | NF | plants | wanting | 47 | 3.165 | 1.328 |
| **FU** | fiber | NF | objects | wanting | 47 | 2.259 | 906 |

Preregistered linear models for model 1/A (food vs. art), model 2/A (intervention effect) for different stimulus classes (stimulus category, stimulus type) for either average across class or stimulus-by-stimulus values (note number of observations: n_obs_category_ > 1,470, n_obs_stimulus_ > 32,000) are reported here.

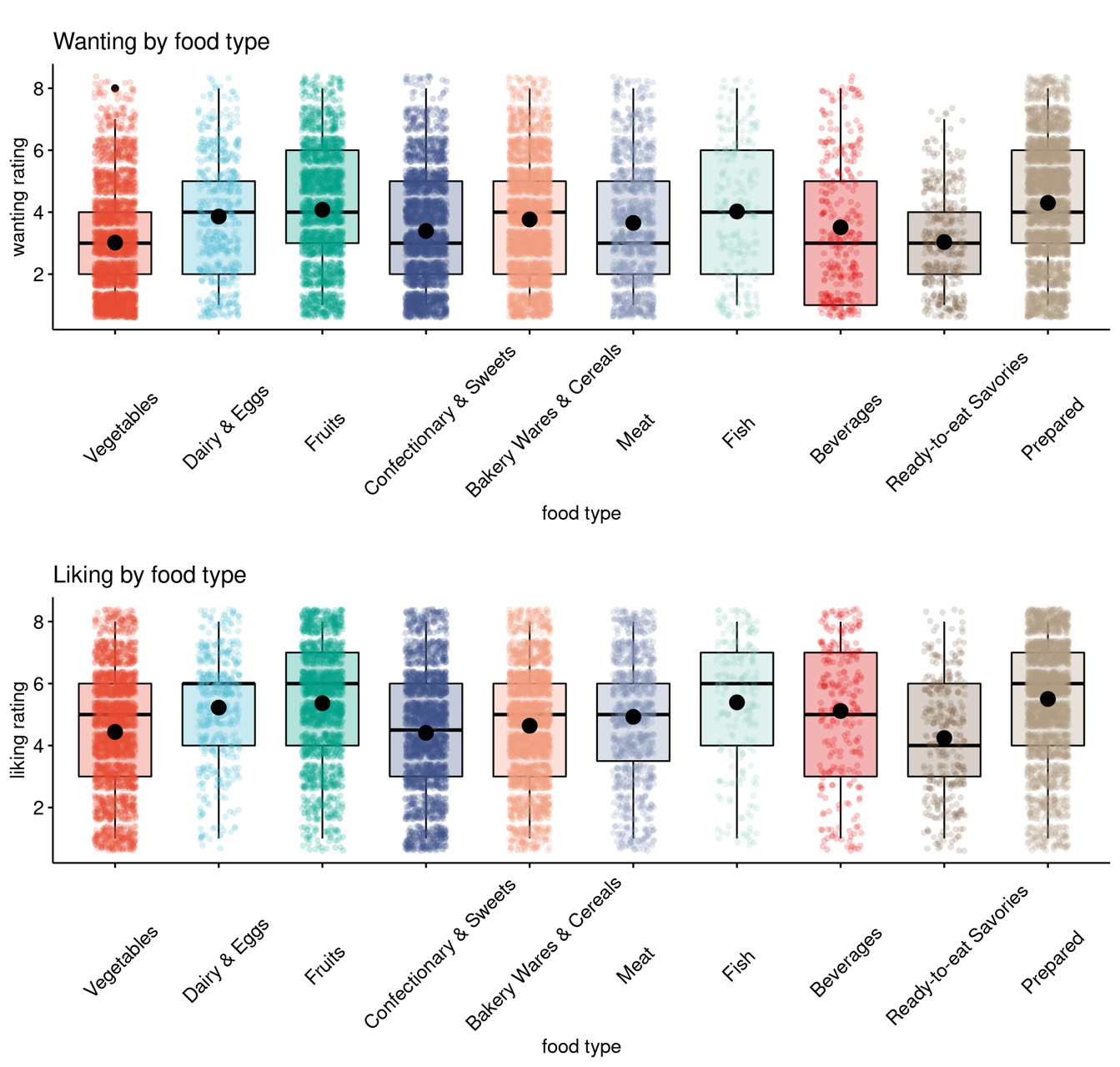

**SI-Fig. 2: Distribution of food wanting and liking ratings by food type.**

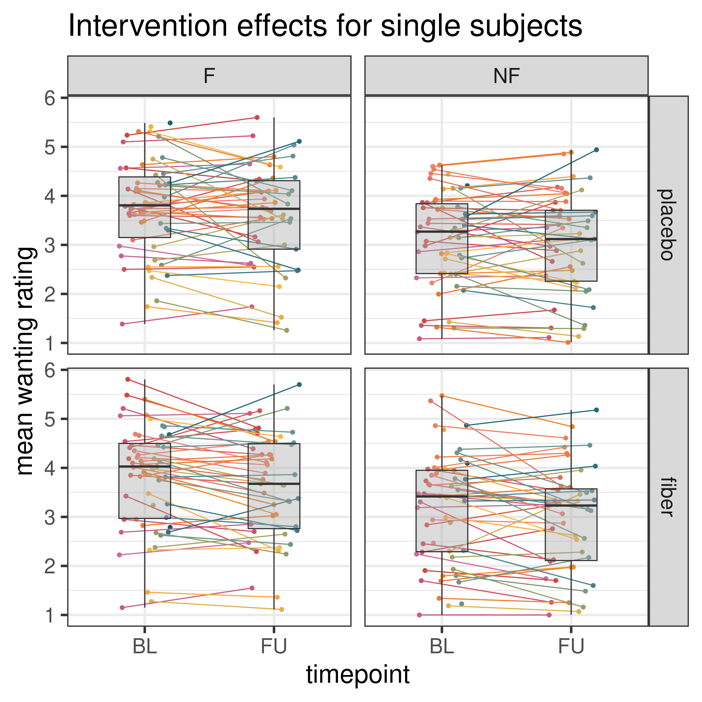

**SI-Fig. 3: Intervention effects on wanting ratings by stimulus category and timepoint.** Average and individual ratings by timepoint and by intervention depicting inter-individual variability in wanting ratings.

Food items higher in protein/100g (b = 0.02, t = 3.35), lower in fiber/100g (b = -0.06, t = -3.51), and to a lesser extent, lower in carbohydrates/100g (b = -0.004, t = -2.22) were more wanted (p_all_ < .03).

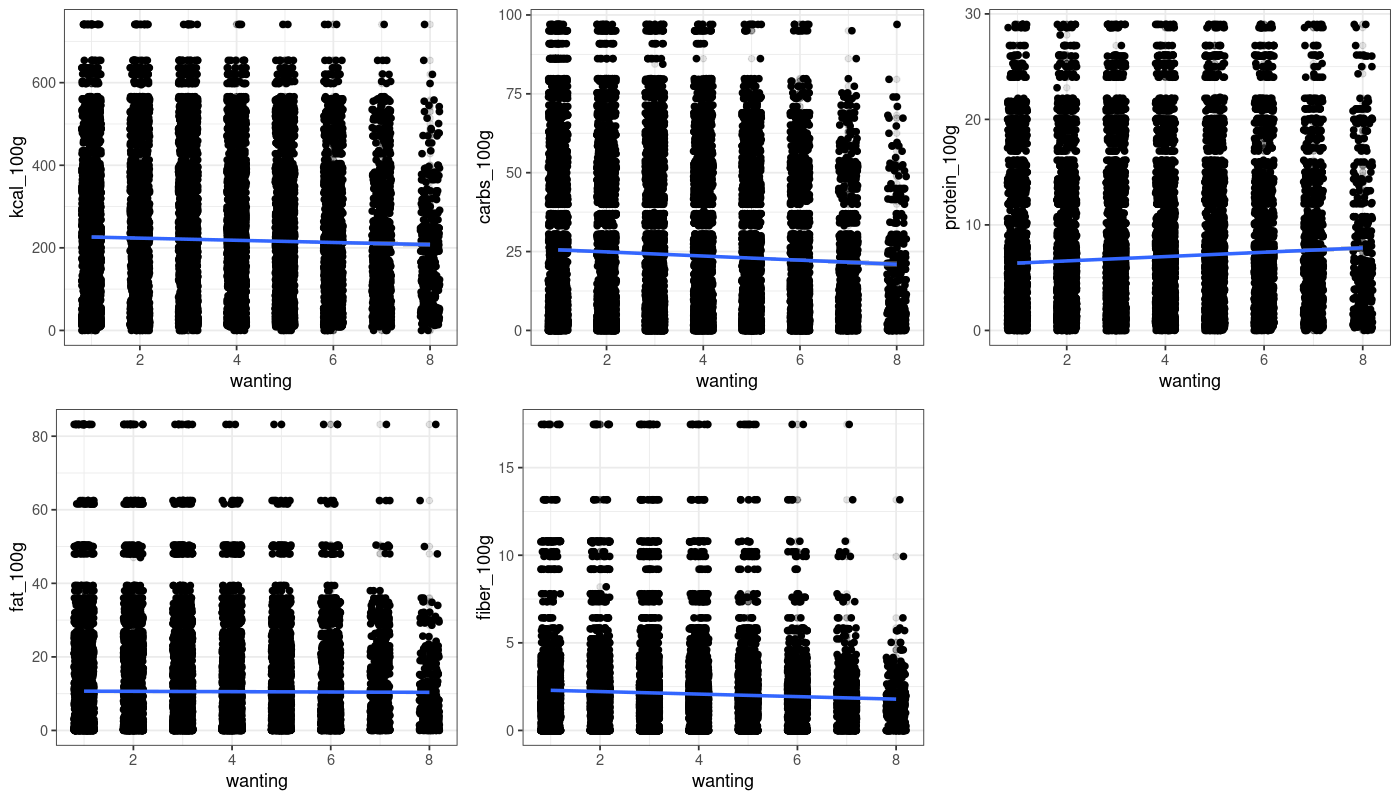

**SI-Fig. 4: Food wanting ratings correlate with nutrient content.**

**Model 1/A: Main effect of stimulus category (food vs. art)**

H_behav_0.1: Individual wanting is higher for food compared to art wanting for between-subject analysis (b = 1.01, t = 54.8, null model comparison p<.001).

**SI-Table 2: Mixed effects linear model results on the subjective wanting for food and art stimuli on the level of stimulus category.**

| **fixed effects** | **estimate** | **SE** | **t-value** |
| --- | --- | --- | --- |
| (intercept) | 2.77 | 0.10 | 28.03 |
| time (follow-up) | -0.12 | 0.02 | -6.20 |
| stim_category (food) | **1.01***** | **0.018** | **55.10** |

***Formula****: wanting ~ timepoint + stim_category + (1 | subject).* ***REML*** *criterion at convergence: 123415, n_obs_ = 32,111, groups: n_subj_ = 59.* ***Significance****,* ********* *p < 0.001*

**Additional analysis for stimulus type**

**SI-Table 3: Mixed effects linear model results on the subjective wanting for food and art stimuli on the level of stimulus type.**

| **fixed effects** | **estimate** | **SE** | **t-value** |
| --- | --- | --- | --- |
| (Intercept) | 3.64 | 0.10 | 35.18 |
| time (follow-up) | -0.12 | 0.02 | -6.35 |
| **stim_typecal1** | **-0.18***** | **0.04** | **-4.30** |
| **stim_typecal2** | **0.57***** | **0.04** | **13.86** |
| **stim_typecal3** | **0.22***** | **0.04** | **5.29** |
| **stim_typecal4** | **-0.03***** | **0.04** | **-0.83** |
| **stim_typeobjects** | **-1.19***** | **0.04** | **-33.09** |
| **stim_typeplants** | **-0.21***** | **0.05** | **-4.67** |

*Formula: wanting ~ timepoint + stim_type + (1 | subject). REML criterion at convergence: 121431, n_obs_ = 32,111, groups: n_subj_ = 59. Significance, *** p < 0.001*

Additional analysis for food type (10 types)

All types of food are more liked than Vegetables with fruits, fish and prepared most liked (between-subject).

**SI-Table 4: Mixed effects linear model results on the subjective wanting for food and art stimuli on the level of food-pics type (food only).**

| **fixed effects** | **estimate** | **SE** | **t-value** |
| --- | --- | --- | --- |
| (intercept) | 3.11 | 0.13 | 24.51 |
| time (follow-up) | -0.15 | 0.03 | -5.90 |
| dairy & eggs | **0.84***** | 0.07 | 11.29 |
| fruits | **1.06***** | 0.05 | 22.90 |
| confectionary & sweets | **0.38***** | 0.04 | 8.77 |
| bakery wares & cereals | **0.75***** | 0.04 | 17.72 |
| meat | **0.63***** | 0.06 | 11.08 |
| fish | **1.01***** | 0.11 | 9.48 |
| beverages | **0.50***** | 0.10 | 5.15 |
| ready-to-eat savories | **0.03***** | 0.09 | 0.37 |
| prepared | **1.28***** | 0.04 | 30.50 |

*Formula: wanting ~ timepoint + food_pics_type + (1 | subject), data = data_F_only. REML criterion at convergence: 61111, n_obs_ = 16,071, groups: n_subj_ = 59. Significance, *** p < 0.001*

**Additional analysis for nutrient content (macronutrients)**

**SI-Table 5: Mixed effects linear model results on the subjective wanting for food and art stimuli per nutrient content (food only).** Less fiber content and higher amounts of protein and carbohydrates related to higher wanting.

| **fixed effects** | **estimate** | **SE** | **t-value** |
| --- | --- | --- | --- |
| (Intercept) | 3.84 | 0.14 | 27.79 |
| kcal_100g | -0.00 | 0.00 | -0.87 |
| time (follow-up) | -0.15 | 0.02 | -6.11 |
| (Intercept) | 3.94 | 0.14 | 29.11 |
| **fiber_100g** | **-0.06***** | **0.02** | **-3.51** |
| time (follow-up) | -0.15 | 0.02 | -6.11 |
| (Intercept) | 3.67 | 0.14 | 27.01 |
| **protein_100g** | **0.02***** | **0.01** | **3.35** |
| time (follow-up) | -0.15 | 0.02 | -6.11 |
| (Intercept) | 3.80 | 0.13 | 28.46 |
| fat_100g | 0.00 | 0.00 | 0.10 |
| time (follow-up) | -0.15 | 0.02 | -6.11 |
| (Intercept) | 3.89 | 0.14 | 28.77 |
| **carbs_100g** | **-3.5*10-3*** | **0.00** | **-2.22** |
| time (follow-up) | -0.15 | 0.02 | -6.11 |

*Formula: wanting ~ timepoint + nutrient_of_interest_pics_type + (1 | subject) + (1 | image_number), data = data_F_only. For each model n_obs_ = 16,071, groups: n_subj_ = 59, n_images_ = 410. Significance, * / *** ANOVA null-full model comparison p < 0.05 / p < 0.001*

**Additional analysis for H_behav_0.1 with hunger rating as covariate**

**SI-Table 6: Mixed effects linear model results on the subjective wanting for subjective hunger rating on wanting by stimulus category.**

| **fixed effects** | **estimate** | **SE** | **t-value** |
| --- | --- | --- | --- |
| (intercept) | 2.84 | 0.11 | 25.79 |
| stim_category (food) | 0.34 | 0.05 | 6.28 |
| hunger (mean pre-/post-wanting task) | -0.02 | 0.01 | -1.34 |
| time (follow-up) | -0.11 | 0.02 | -5.81 |
| stim_category (food) * hunger_mean | 0.15 | 0.01 | 13.02 |

*Formula: wanting ~ stim_category * hunger_mean_wanting + timepoint + stim_category + hunger_mean_wanting + (1 | subject). REML criterion at convergence: 123231, n_obs_ = 32,111, groups: n_subj_ = 59. Significance, *** p < 0.001*

**Model 2/A: Intervention effect**

H_behav_A0: Individual food wanting compared to art wanting is not different after a two-week high-fiber intervention, when looking at stimulus category (R1 with timepoint*intervention*stim_category vs. R0 p = 0.31), but for stimulus type (R1 with timepoint*intervention*stim_type vs. R0, p = 0.002), and not for food_pics type (R1 with timepoint*intervention*food_pics_type vs. R0, p = 0.20).

**SI-Table 7: Mixed effects linear model results on the subjective wanting for post-intervention by stimulus category.**

| **fixed effects** | **estimate** | **SE** | **t-value** |
| --- | --- | --- | --- |
| (intercept) | 2.72 | 0.10 | 26.81 |
| time (follow-up) | -0.01 | 0.04 | -0.16 |
| intervention (prebiotic) | 0.08 | 0.04 | 2.15 |
| stim_category (food) | 1.01 | 0.04 | 28.09 |
| time (follow-up) * intervention (prebiotic) | -0.16 | 0.05 | -3.06 |
| time (follow-up) * stim_category (food) | -0.09 | 0.05 | -1.78 |
| intervention (prebiotic) * stim_category (food) | 0.05 | 0.05 | 0.92 |
| time (follow-up) * intervention (prebiotic) * stim_category (food) | 0.07 | 0.07 | 0.90 |

*Formula: wanting ~ timepoint*intervention*stim_category+timepoint*stim_category+timepoint*intervention+stim_category*intervention+timepoint+stim_category+intervention+(1|subject). REML criterion at convergence: 123415, n_obs_ = 32,111, groups: n_subj_ = 59.*

**SI-Table 8: Mixed effects linear model results on the subjective wanting for post-intervention by stimulus type.**

| **fixed effects** | **estimate** | **SE** | **t-value** |
| --- | --- | --- | --- |
| (intercept) | 3.39 | 0.11 | 30.86 |
| time (follow-up) | -0.04 | 0.07 | -0.57 |
| intervention (prebiotic) | 0.18 | 0.07 | 2.57 |
| stim_type (cal2) | 0.77 | 0.07 | 11.11 |
| stim_type (cal3) | 0.45 | 0.07 | 6.39 |
| stim_type (cal4) | 0.13 | 0.07 | 1.80 |
| stim_type (animals) | 0.28 | 0.08 | 3.47 |
| stim_type (plants) | -0.15 | 0.08 | -1.88 |
| stim_type (objects) | -0.99 | 0.06 | -17.21 |
| time (follow-up) * intervention (prebiotic) | -0.26 | 0.10 | -2.58 |
| time (follow-up) * stim_type (cal2) | -0.09 | 0.10 | -0.84 |
| time (follow-up) * stim_type (cal3) | -0.10 | 0.10 | -0.97 |
| time (follow-up) * stim_type (cal4) | -0.04 | 0.10 | -0.44 |
| time (follow-up) * stim_type (animals) | -0.16 | 0.12 | -1.38 |
| time (follow-up) * stim_type (plants) | 0.17 | 0.12 | 1.49 |
| time (follow-up) * stim_type (objects) | 0.05 | 0.08 | 0.57 |
| intervention (prebiotic) * stim_type (cal2) | -0.10 | 0.10 | -1.03 |
| intervention (prebiotic) * stim_type (cal3) | -0.18 | 0.10 | -1.85 |
| intervention (prebiotic) * stim_type (cal4) | 0.06 | 0.10 | 0.66 |
| intervention (prebiotic) * stim_type (animals) | -0.20 | 0.11 | -1.80 |
| intervention (prebiotic) * stim_type (plants) | 0.18 | 0.11 | 1.59 |
| intervention (prebiotic) * stim_type (objects) | -0.14 | 0.08 | -1.70 |
| **time (follow-up) * intervention (prebiotic) * stim_type (cal2)** | **0.27**** | **0.14** | **1.91** |
| **time (follow-up) * intervention (prebiotic) * stim_type (cal3)** | **0.37**** | **0.14** | **2.58** |
| **time (follow-up) * intervention (prebiotic) * stim_type (cal4)** | **0.03**** | **0.14** | **0.19** |
| **time (follow-up) * intervention (prebiotic) * stim_type (animals)** | **0.33**** | **0.16** | **1.97** |
| **time (follow-up) * intervention (prebiotic) * stim_type (plants)** | **-0.25**** | **0.16** | **-1.51** |
| **time (follow-up) * intervention (prebiotic) * stim_type (objects)** | **0.13**** | **0.12** | **1.07** |

*Formula: wanting ~ timepoint*intervention*stim_type+timepoint*stim_type+timepoint*intervention+stim_type*intervention+timepoint+stim_type+intervention+(1|subject). REML criterion at convergence: 121445, n_obs_ = 32,111, groups: n_subj_ = 59. Significance, ** p < 0.01*

**Model 2: Impact of hunger on intervention effect**

**SI-Table 9: Mixed effects linear model results on the subjective wanting for post-intervention by stimulus category dependent on hunger rating.**

| **fixed effects** | **estimate** | **SE** | **t-value** |
| --- | --- | --- | --- |
| (intercept) | 3.05 | 0.13 | 23.58 |
| time (follow-up) | -0.45 | 0.11 | -4.02 |
| intervention (prebiotic) | -0.24 | 0.11 | -2.18 |
| stim_category (food) | 0.48 | 0.11 | 4.58 |
| mean hunger rating | -0.08 | 0.02 | -3.94 |
| time (follow-up) * intervention (prebiotic) | 0.47 | 0.16 | 2.95 |
| time (follow-up) * stim_category (food) | -0.11 | 0.15 | -0.69 |
| intervention (prebiotic) * stim_category (food) | -0.19 | 0.15 | -1.29 |
| time (follow-up) * mean hunger rating | 0.10 | 0.02 | 4.20 |
| intervention (prebiotic) * mean hunger rating | 0.08 | 0.02 | 3.21 |
| stim_category (food) * mean hunger rating | 0.12 | 0.02 | 5.34 |
| time (follow-up) * intervention (prebiotic) * stim_category (food) | 0.04 | 0.22 | 0.18 |
| time (follow-up) * intervention (prebiotic) * mean hunger rating | -0.15 | 0.03 | -4.27 |
| time (follow-up) * stim_category (food) * mean hunger rating | 0.00 | 0.03 | 0.09 |
| intervention (prebiotic) * stim_category (food) * mean hunger rating | 0.04 | 0.03 | 1.27 |
| **time (follow-up) * intervention (prebiotic) * stim_category (food) * mean hunger rating** | **0.02***** | **0.05***** | **0.35***** |

*Formula: wanting ~ timepoint * intervention * stim_category * hunger_mean_wanting + timepoint * intervention * stim_category + timepoint * stim_category + timepoint * intervention + stim_category * intervention + timepoint + stim_category + intervention + (1 | subject). REML criterion at convergence: 123228, n_obs_ = 32,110 , groups: n_subj_ = 59. Significance, *** p < 0.001*

**H_behav_3/B: Nutrient content effect on wanting (food only)**

H_behav_B1: Individual food wanting is not different for kcal_100g content after a two-week high-fiber intervention across all food stimuli (null model comparison p = .86).

**SI-Table 10: Mixed effects linear model results on the subjective wanting for post-intervention dependent on caloric content (kcal / 100g).**

| **fixed effects** | **estimate** | **SE** | **t-value** |
| --- | --- | --- | --- |
| (intercept) | 3.92 | 0.13 | 29.92 |
| time (follow-up) | -0.01 | 0.06 | -0.20 |
| intervention (prebiotic) | 0.05 | 0.06 | 0.91 |
| kcal_100g | -3.45*10^-4^ | 1.47*10^-4^ | -2.38 |
| session (session 2, 3, 4) | -0.21 | 0.04 | -5.64 |
| time (follow-up) * intervention (prebiotic) | -0.07 | 0.08 | -0.87 |
| time (follow-up) * kcal_100g | -6.1*10^-5^ | 2.11*10^-4^ | -0.29 |
| intervention (prebiotic) * kcal_100g | 1.87*10^-4^ | 2.05*10^-4^ | 0.91 |
| time (follow-up) * intervention (prebiotic) * kcal_100g | -5.42*10^-5^ | 2.98*10^-4^ | -0.18 |

*Formula: wanting ~ timepoint * intervention * kcal_100g + intervention * kcal_100g + timepoint * kcal_100g + timepoint * intervention + intervention + timepoint + kcal_100g + (1 | subject) + session_1_2, data: data_F_only). REML criterion at convergence: 62283, n_obs_ = 16,071, groups: n_subj_ = 59.*

H_behav_B2*:* Individual food wanting is not different for fiber_100g content after a two-week high-fiber intervention across all food stimuli (null model comparison p = 0.34).

**SI-Table 11: Mixed effects linear model results on the subjective wanting for post-intervention dependent on fiber content (fiber / 100g).**

| **fixed effects** | **estimate** | **SE** | **t-value** |
| --- | --- | --- | --- |
| (intercept) | 3.98 | 0.13 | 30.72 |
| time (follow-up) | -0.01 | 0.06 | -0.17 |
| intervention (prebiotic) | 0.05 | 0.05 | 1.05 |
| fiber_100g | -0.07 | 0.01 | -5.40 |
| session (session 2, 3, 4) | -0.21 | 0.04 | -5.66 |
| time (follow-up) * intervention (prebiotic) | -0.04 | 0.08 | -0.48 |
| time (follow-up) * fiber_100g | 0.005 | 0.02 | 0.28 |
| intervention (prebiotic) * fiber_100g | 0.02 | 0.02 | 1.10 |
| time (follow-up) * intervention (prebiotic) * fiber_100g | -0.02 | 0.03 | -0.96 |

*Formula: wanting ~ timepoint * intervention * fiber_100g + intervention * fiber_100g + timepoint * fiber_100g + timepoint * intervention + intervention + timepoint + fiber_100g + (1 | subject) + session_1_2, data: data_F_only). REML criterion at convergence: 62170, n_obs_ = 16,071, groups: n_subj_ = 59. Significance, *** p < 0.001.*

**Model C: Liking as a potential confounding variable on wanting ratings**

Test if subjective liking is a confounding variable for subjective wanting. Note that less datapoints could be included due to incomplete liking ratings.

**SI-Table 12: Mixed effects linear model results on the subjective wanting for post-intervention dependent on subjective liking ratings.**

| **fixed effects** | **estimate** | **SE** | **t-value** |
| --- | --- | --- | --- |
| (intercept) | 1.10 | 0.09 | 11.84 |
| time (follow-up) | -0.05 | 0.03 | -1.34 |
| intervention (prebiotic) | 0.07 | 0.03 | 2.07 |
| stim_category (food) | 0.11 | 0.04 | 3.15 |
| **liking** | **0.51***** | **0.01** | **102.46** |
| time (follow-up) * intervention (prebiotic) | -0.12 | 0.05 | -2.48 |
| time (follow-up) * stim_category (food) | -0.09 | 0.05 | -1.79 |
| intervention (prebiotic) * stim_category (food) | 0.01 | 0.05 | 0.21 |
| time (follow-up) * intervention (prebiotic) * stim_category (food) | 0.03 | 0.07 | 0.49 |

*Formula: wanting ~ timepoint * intervention * stim_category + timepoint * stim_category + timepoint * intervention + stim_category * intervention + liking + timepoint + stim_category + intervention + (1 | subject), (data = data_liking_only). REML criterion at convergence: 96357, n_obs_ = 27,445, groups: n_subj_ = 45. Significance, *** p < 0.001.*

Interpretation: Yes, subjective liking has a significant positive impact on wanting ratings (p <.001).

**Additional analyses: Test order effects of visits on wanting ratings and intervention effects**

Check if session order (1 2 3 4) has an impact on wanting ratings.

**SI-Table 13: Mixed effects linear model results on the subjective wanting for post-intervention dependent on session (1 2 3 4).**

| **fixed effects** | **estimate** | **SE** | **t-value** |
| --- | --- | --- | --- |
| (intercept) | 3.39 | 0.10 | 33.94 |
| intervention (prebiotic) | -0.10 | 0.03 | -3.56 |
| time (follow-up) | -0.36 | 0.03 | -10.35 |
| **session (ses-02)** | **0.18***** | **0.03** | **6.31** |
| **session (ses-03)** | **-0.16***** | **0.03** | **-5.99** |
| intervention (prebiotic) * time (follow-up) | 0.13 | 0.04 | 3.42 |

*Note: Fixed-effect model matrix is rank deficient so dropping 1 column / coefficient. Formula: wanting ~ intervention * timepoint + session + (1 | subject). REML criterion at convergence: 126212, n_obs_ = 32,111 , groups: n_subj_ = 59. Significance, *** p < 0.001.*

Interpretation: Yes, the session order has a significant impact on wanting ratings (p <.001).

Check if first session compared to all others (1 2 2 2) has an impact on wanting ratings.

**SI-Table 14: Mixed effects linear model results on the subjective wanting for post-intervention dependent on first session (1 2 2 2).**

| **fixed effects** | **estimate** | **SE** | **t-value** |
| --- | --- | --- | --- |
| (intercept) | 3.30 | 0.10 | 32.88 |
| intervention (prebiotic) | 0.10 | 0.03 | 3.53 |
| time (follow-up) | 0.03 | 0.03 | 0.93 |
| **session (sessions 2, 3, 4)** | **-0.16***** | **0.03***** | **-5.96***** |
| intervention (prebiotic) * time (follow-up) | -0.12 | 0.04 | -3.13 |

*Formula: wanting ~ intervention * timepoint + session + (1 | subject). REML criterion at convergence: 126246, n_obs_ = 32,111 , groups: n_subj_ = 59. Significance, *** p < 0.001.*

Interpretation: Yes, the first testing session has a significant positive impact on wanting ratings (p <.001).

Check if first session compared to all others (1 2 2 2) has an impact on post-intervention change in wanting ratings by stimulus category (F vs. NF).

**SI-Table 15: Mixed effects linear model results on the subjective wanting for post-intervention by stimulus category accounting for first session (1 2 2 2).**

| **fixed effects** | **estimate** | **SE** | **t-value** |
| --- | --- | --- | --- |
| (intercept) | 2.79 | 0.10 | 27.45 |
| time (follow-up) | 0.08 | 0.04 | 1.93 |
| intervention (prebiotic) | 0.07 | 0.04 | 2.05 |
| stim_category (food) | 1.01 | 0.04 | 28.10 |
| session (sessions 2, 3, 4) | -0.16 | 0.03 | -6.26 |
| intervention (prebiotic) * time (follow-up) | -0.16 | 0.05 | -2.98 |
| time (follow-up) * stim_category (food) | -0.09 | 0.05 | -1.77 |
| intervention (prebiotic) * stim_category (food) | 0.05 | 0.05 | 0.93 |
| time (follow-up) * intervention (prebiotic) * stim_category (food) | 0.07 | 0.07 | 0.89 |

*Formula: wanting ~ timepoint * intervention * stim_category + timepoint * stim_category + timepoint * intervention + stim_category * intervention + timepoint + stim_category + intervention + (1 | subject) + session_1_2. REML criterion at convergence: 123381, n_obs_ = 32,110 , groups: n_subj_ = 59. Model comparison, p = 0.37.*

-> No, there is no significant difference to the model with or without session as covariate (p = 0.37).

Check if first session compared to all others (1 2 2 2) has an impact on post-intervention change in wanting ratings by stimulus type (cal1, cal2, cal3, cal4, animals, plants, objects).

**SI-Table 16: Mixed effects linear model results on the subjective wanting for post-intervention by stimulus type accounting for first session (1 2 2 2).**

| **fixed effects** | **estimate** | **SE** | **t-value** |
| --- | --- | --- | --- |
| (intercept) | 3.83 | 0.12 | 32.72 |
| time (follow-up) | -0.22 | 0.07 | -2.97 |
| intervention (prebiotic) | -0.16 | 0.07 | -2.34 |
| stim_type (cal2) | 0.67 | 0.07 | 9.91 |
| stim_type (cal3) | 0.27 | 0.07 | 3.91 |
| stim_type (cal4) | 0.19 | 0.07 | 2.78 |
| stim_type (animals) | 0.08 | 0.08 | 0.96 |
| stim_type (plants) | 0.03 | 0.08 | 0.35 |
| stim_type (objects) | -1.13 | 0.06 | -20.11 |
| session (sessions 2, 3, 4) | -0.17 | 0.03 | -6.52 |
| intervention (prebiotic) * time (follow-up) | 0.24 | 0.10 | 2.42 |
| time (follow-up) * stim_type (cal2) | 0.19 | 0.10 | 1.87 |
| time (follow-up) * stim_type (cal3) | 0.27 | 0.10 | 2.69 |
| time (follow-up) * stim_type (cal4) | -0.02 | 0.10 | -0.17 |
| time (follow-up) * stim_type (animals) | 0.16 | 0.12 | 1.42 |
| time (follow-up) * stim_type (plants) | -0.07 | 0.12 | -0.64 |
| time (follow-up) * stim_type (objects) | 0.17 | 0.08 | 2.10 |
| intervention (prebiotic) * stim_type (cal2) | 0.10 | 0.10 | 0.99 |
| intervention (prebiotic) * stim_type (cal3) | 0.17 | 0.10 | 1.78 |
| intervention (prebiotic) * stim_type (cal4) | -0.06 | 0.10 | -0.61 |
| intervention (prebiotic) * stim_type (animals) | 0.19 | 0.11 | 1.66 |
| intervention (prebiotic) * stim_type (plants) | -0.15 | 0.11 | -1.30 |
| intervention (prebiotic) * stim_type (objects) | 0.10 | 0.08 | 1.26 |
| **time (follow-up) * intervention (prebiotic) * stim_type (cal2)** | **-0.27**** | **0.14** | **-1.89** |
| **time (follow-up) * intervention (prebiotic) * stim_type (cal3)** | **-0.36**** | **0.14** | **-2.54** |
| **time (follow-up) * intervention (prebiotic) * stim_type (cal4)** | **-0.03**** | **0.14** | **-0.23** |
| **time (follow-up) * intervention (prebiotic) * stim_type (animals)** | **-0.31**** | **0.16** | **-1.88** |
| **time (follow-up) * intervention (prebiotic) * stim_type (plants)** | **0.22**** | **0.16** | **1.31** |
| **time (follow-up) * intervention (prebiotic) * stim_type (objects)** | **-0.09**** | **0.12** | **-0.77** |

*Formula: wanting ~ timepoint * intervention * stim_type + timepoint * stim_ type + timepoint * intervention + stim_ type * intervention + timepoint + stim_ type + intervention + (1 | subject) + session_1_2. REML criterion at convergence: 121409, n_obs_ = 32,111, groups: n_subj_ = 59. Significance, ** p = 0.0018*

-> Yes, the significant difference post-intervention for stimulus type for all items remains with session as covariate (p = 0.0018).

Check if first session compared to all others (1 2 2 2) has an impact on post-intervention change in wanting ratings by stimulus type (cal1, cal2, cal3, cal4) for food items only.

**SI-Table 17: Mixed effects linear model results on the subjective wanting for post-intervention by stimulus type accounting for first session (1 2 2 2) for food items only.**

| **fixed effects** | **estimate** | **SE** | **t-value** |
| --- | --- | --- | --- |
| (intercept) | 3.51 | 0.13 | 26.10 |
| time (follow-up) | 0.06 | 0.08 | 0.74 |
| intervention (prebiotic) | 0.15 | 0.07 | 2.05 |
| stim_type (cal2) | 0.78 | 0.07 | 10.81 |
| stim_type (cal3) | 0.45 | 0.07 | 6.22 |
| stim_type (cal4) | 0.13 | 0.07 | 1.74 |
| session (sessions 2, 3, 4) | -0.21 | 0.04 | -5.74 |
| intervention (prebiotic) * time (follow-up) | -0.25 | 0.10 | -2.40 |
| time (follow-up) * stim_type (cal2) | -0.08 | 0.10 | -0.81 |
| time (follow-up) * stim_type (cal3) | -0.10 | 0.10 | -0.94 |
| time (follow-up) * stim_type (cal4) | -0.04 | 0.10 | -0.42 |
| intervention (prebiotic) * stim_type (cal2) | -0.10 | 0.10 | -1.00 |
| intervention (prebiotic) * stim_type (cal3) | -0.18 | 0.10 | -1.78 |
| intervention (prebiotic) * stim_type (cal4) | 0.07 | 0.10 | 0.68 |
| **time (follow-up) * intervention (prebiotic) * stim_type (cal2)** | 0.27* | 0.15* | 1.83* |
| **time (follow-up) * intervention (prebiotic) * stim_type (cal3)** | 0.36* | 0.15* | 2.48* |
| **time (follow-up) * intervention (prebiotic) * stim_type (cal4)** | 0.03* | 0.15* | 0.19* |

*Formula: wanting ~ timepoint * intervention * stim_type + timepoint * stim_ type + timepoint * intervention + stim_ type * intervention + timepoint + stim_ type + intervention + (1 | subject) + session_1_2 (data= data_F_only). REML criterion at convergence: 62081, n_obs_ = 16,071, groups: n_subj_ = 59. Significance, * p < 0.03.*

—> Yes, the significant difference post-intervention for wanting by stimulus type for food only remains with session as covariate (p = 0.03).

**Effects of subjective hunger**

Individuals’ subjective hunger ratings during fMRI sessions were diverse and not significantly different after prebiotic intervention compared to placebo (interaction b = -0.33, t = -1.18; null model comparison p > 0.23, **SI_behav Table 6, 18-19**). However, the less hungry they were, the lower their overall wanting rating for food compared to art items (b = 0.15, t = 13.0, null model comparison p < .001, **SI_behav Table 9**). When adding mean hunger as an interaction term to the model, this effect appeared marginally more pronounced after prebiotics (time*group*category*hunger: b = 0.01, t = 0.3, model comparison p < .001).

SI-Table 18: Subjective hunger ratings by timepoint for each intervention arm. Ratings were measured using a Likert scale inside MR scanner at 10 and 40 min after 10% energy intake using a breakfast shake, with a scale from 1 (not at all) to 8 (extremely).

| Timepoint | Intervention | n | hunger rating | | | |
| --- | --- | --- | --- | --- | --- | --- |
|  |  |  | 10 min postprandial | 40 min postprandia | Mean 10-40 min | |
|  |  |  | mean ± SD | mean ± SD | mean ± SD | change± SD |
| BL | prebiotic | 55 | 4.25 ± 1.76 | 5.16 ± 1.71 | 4.71 ± 1.66 | - |
| FU | prebiotic | 48 | 4.13 ± 1.66 | 4.72 ± 1.73 | 4.43 ± 1.60 | -0.28 ± 1.32 |
| BL | placebo | 53 | 3.77 ± 1.69 | 4.79 ± 1.65 | 4.28 ± 1.58 | - |
| FU | placebo | 49 | 3.92 ± 1.68 | 4.67 ± 1.72 | 4.30 ± 1.54 | 0.05 ± 1.37 |

**SI-Table 19: Mixed effects linear model results on the effects of prebiotic intervention on subjective hunger ratings (average).** Ratings were measured using a Likert scale inside MR scanner at 10 and 40 min after 10% energy intake and averaged, with a scale from 1 (not at all) to 8 (extremely). Model comparison, p > 0.23.

| ***fixed effects*** | **estimate** | **SE** | **t-value** |
| --- | --- | --- | --- |
| (intercept) | 4.04 | 0.84 | 4.84 |
| time (follow-up) | 0.04 | 0.20 | 0.18 |
| group (prebiotics) | 0.43 | 0.20 | 2.20 |
| age | 0.02 | 0.03 | 0.57 |
| gender (male) | -0.32 | 0.41 | -0.78 |
| time*group | -0.33 | 0.28 | -1.18 |

***Formula****: hunger ~ timepoint * intervention + (1 | subject) + age + gender.* ***REML*** *criterion at convergence: 689, Number of observations: 204, groups: participants, 59.*

**Additional wanting models with “true wanting” models considering weighted ratings.**

We did not further explore interaction effects for wanting ratings modelled as dependent outcome variable in three different ways (1, individual wanting – individual liking; 2, individual wanting - individual liking - population mean of wanting; 3 individual wanting* population mean of wanting) per item to simplify results.
